# Efficacy and safety of pharmacological interventions for the treatment of cocaine use disorder: a systematic review and network meta-analysis

**DOI:** 10.64898/2025.12.10.25341965

**Authors:** Craig Paterson, Thomas Parkhouse, Chloe Burke, Monika Halicka, Jennifer C Palmer, Hend Gabr, Rebecca Wilson, Katie E Webster, Francesca Spiga, Sarah Dawson, Deborah M Caldwell, Jennifer Scott, Julian PT Higgins, Jelena Savović

## Abstract

**Background and Aims:** Cocaine use disorder (CUD) is an increasingly widespread concern worldwide. Various pharmacological treatments have been investigated for CUD, but their efficacy remains unclear, and none have been licensed for treatment. Therefore, we assessed the comparative effectiveness, safety, and acceptability of pharmacological interventions for the treatment of CUD and prevention of relapse.

**Design:** Systematic review and network meta-analyses of double-blind randomized controlled trials (RCTs). PROSPERO (CRD42024596434). We assessed risk of bias using the Cochrane Risk of Bias 2 tool and certainty of evidence was assessed using the Confidence in Network Meta-analysis framework.

**Setting:** Any in/outpatient setting, with no restriction on geographic location.

**Participants:** Adults with diagnosed CUD. Participants with co-occurring substance use disorders and/or common mental health conditions were eligible.

**Interventions:** Pharmacologicalinterventions specifically tested for the treatment of CUD with a minimum follow-up of 4 weeks.

**Measurements:** We assessed 12 effectiveness, three acceptability, and two safety outcomes. Our primary outcomes were continuous abstinence, point abstinence, dropout for any reason, and serious adverse events. We also examined longest duration of continuous abstinence, extent of cocaine use, craving, severity of dependence, dropout due to adverse events, adherence, and mortality.

**Findings:** We included 218 reports of 163 studies (14871 participants) assessing 89 unique medications, grouped into 8 categories. Most study results had some concerns or high risk of bias, and the results of syntheses were generally judged to be very low certainty evidence. As such, the findings ought to be interpreted with caution. No single treatment type showed consistent beneficial effects across all effectiveness outcomes and there was limited evidence that results varied by intervention type or presence of co-occurring comorbidity.

**Conclusions:** Despite continued work in this area, effective pharmacological treatments for CUD remain elusive.

## BACKGROUND

According to the UN World Drug Report 2025, an estimated 25 million individuals over the age of 15 years used cocaine in the past year, representing 0.5% of the global population (1). Only a fraction will go on to experience cocaine use disorder (CUD), but for those that do, it is characterized by compulsive use of cocaine despite the physical or psychological harm it may cause (2). Diagnosis is typically established according to the American Psychiatric Association Diagnostic and Statistical Manual of Mental Disorders (DSM-5), where an individual displays at least two of 11 physiological and behavioural symptoms (3). Estimates suggest that 20% of individuals who use cocaine meet criteria for, or may develop, the disorder within 2 years (4,5). Chronic use of cocaine is associated with increased all-cause mortality, acute and chronic cardiovascular pathology (6), as well as neurological deficits such as impaired attention and working memory, and response inhibition (7). At a societal level, cocaine use is associated with increased violence, crime and the transmission of HIV and hepatitis C (8,9).

Despite concerted efforts in this area, no standard care pathway for CUD exists, even as demand for specialist treatment continues to rise (10). A previous review of randomized controlled trials (RCTs) investigating the use of psychosocial and pharmacological interventions for CUD concluded that only contingency management (a psychosocial intervention) showed a significant reduction in cocaine use amongst adults (11). While promising, psychosocial interventions may be limited in their ability to address the physical symptoms of CUD such as cravings and withdrawal. Pharmacological interventions may have a role in treating CUD, as they have for other substance use disorders, such as alcohol (12,13) and opioids (14). However, despite promising results from RCTs investigating a wide variety of medications (e.g., psychostimulants, antidepressants, anticonvulsants), systematic reviews have failed to demonstrate clear or consistent benefits of pharmacological interventions for CUD (11,15,16). As a result, no established recommendation for pharmacotherapy exists and no drugs have specific approval for use in the treatment of CUD. This gap underscores the need for an up-to-date systematic review and network meta-analysis (NMA) to provide a coherent comparison of the relative effects of these numerous treatment options.

### Aims and objectives

This review aimed to determine the comparative effectiveness, safety, and acceptability of different pharmacological interventions for the treatment of CUD and prevention of relapse.

## METHODS

This review was pre-registered with PROSPERO (CRD42024596434) and NIHR (NIHR165378). Input from people with lived experience of cocaine use, informed protocol development. This review is reported following PRISMA guidelines (17).

### Eligibility criteria

We sought RCTs with at least four weeks of follow-up from baseline. To be eligible for inclusion, trial participants and personnel must have been blinded to intervention allocation, which may have been evidenced by use of a placebo. Individually-randomized or cluster-randomized trials were eligible, as were trials with factorial and cross-over designs. However, only data from the first period of a cross-over trial was eligible. Studies reported only in conference abstracts, ongoing trial protocols, and other “grey literature” were ineligible.

#### Population

Eligible studies were of individuals aged ≥ 18 years with cocaine use disorder or dependence (including crack cocaine), according to formal diagnostic criteria (e.g., DSM-5, DSM IV, or ICD classification). For convenience, we use the term CUD throughout, which should be interpreted as including cocaine dependence. Studies of individuals diagnosed with cocaine abuse or where a significant majority (>80%) of participants were not diagnosed with CUD were excluded.

Participants with co-occurring substance use disorders (SUD) and those with common mental health conditions (e.g., depression) were eligible. However, studies in participants with co-occurring bipolar, schizophrenic, dissociative, delirium, dementia or amnestic disorders were excluded. Studies of individuals who were in receipt of prescription stimulants within six months of trial participation, studies of opportunistic screening and treatment, and studies conducted solely in laboratory settings were also excluded.

#### Interventions

Any pharmacological intervention (including combinations) was eligible if it was used for the treatment of CUD. No restrictions were placed on intervention setting (inpatient vs outpatient), treatment supervision, or presence of co-occurring psychosocial interventions or support, providing a direct comparison for the evaluation of the pharmacological intervention effect was available.

#### Comparators

Eligible comparators were placebos or any other eligible pharmacological intervention. Psychosocial interventions in isolation were not eligible comparators. Studies only comparing the effectiveness of different doses, formulations, release or delivery schedules of the same pharmacological agent, but with no distinct comparator, were included in the systematic review but not the NMAs.

#### Outcomes and timepoints of interest

Eligible outcomes are detailed in Table 1 and include 12 effectiveness, three acceptability, and two safety outcomes. Where studies reported multiple measures for a given outcome, we followed a hierarchy of preference or conducted a within-trial synthesis to combine estimates taking account of correlations between outcomes (Supporting information S1.1). Where studies did not report outcomes and/or timepoints of interest, they were included in the systematic review, but not NMAs.

**Table 1.** Review outcomes and time points of interest with definitions.

| Outcome | Definition | Time point | Studies with outcome | Studies in NMA | Classes in NMA |
| --- | --- | --- | --- | --- | --- |
| Effectiveness |  |  |  |  |  |
| Primary |  |  |  |  |  |
| Continuous abstinence | Whether a participant was abstinent from cocaine use for a specified duration (confirmed using more than 2 consecutive tests) of 1 week or more at (i) any point during the treatment period or through to the end of treatment and (ii) the longest follow-up. Only confirmed or verified outcomes were eligible (e.g., urinalysis), solely self-report measures were excluded | During treatment | 50 | 46 | 9 |
|  |  | During follow-up | 5 | 5 | 4 |
| Point abstinence | Whether the participant was abstinent at the time of assessment, based on confirmed or verified outcomes, as above. | End of treatment | 27 | 22 | 8 |
|  |  | End of follow-up | 7 | 7 | 7 |
| Secondary |  |  |  |  |  |
| Longest duration of continuous abstinence | Defined as the longest duration of continuous abstinence during the treatment period, based on self-report and/or urinalysis | During treatment | 20 | 19 | 6 |
| Extent of use – frequency | How often participants self-reported using cocaine in a given period | During treatment | 54 | 40 | 9 |
|  |  | During follow-up | 8 | 7 | 5 |
| Extent of use – quantity | How much cocaine participants used in a given period | During treatment | 30 | 19 | 8 |
|  |  | During follow-up | 5 | 4 | 5 |
| Extent of use – urine BE levels | Level of BE detected in participant's urine as a measure of recent cocaine use | During treatment | 27 | 12 | 7 |
|  |  | During follow up | 4 | 0 | 0 |
| Extent of use – proportion of UDS | Proportion of positive/negative urinary drug screenings during a given period | During treatment | 62 | 45 | 8 |
|  |  | During follow-up | 2 | 0 | 0 |
| Craving | As measured by any validated scale, at the end of treatment | End of treatment | 83 | 39 | 8 |
| Severity of dependence | Defined as a change in the number of formal CUD diagnostic criteria met by participants | End of treatment | 1 | 0 | 0 |
| Acceptability |  |  |  |  |  |
| Primary |  |  |  |  |  |
| Dropout from treatment for any reason | Defined as non-completion of the full treatment period for any reason | End of treatment | 141 | 135 | 9 |
| Secondary |  |  |  |  |  |
| Dropout due to adverse events | Defined as non-completion of the full treatment period due to adverse events | End of treatment | 57 | 39 | 9 |
| Adherence | Defined as adherence to study medication protocol (rather than adherence to adjuvant therapy) | During treatment | 67 | 52 | 9 |
| Safety |  |  |  |  |  |
| Primary |  |  |  |  |  |
| Serious adverse events | As defined by the study authors | End of treatment | 88 | 41 | 9 |
| Secondary |  |  |  |  |  |
| Mortality | From any cause | End of treatment | 19 | 4 | 5 |

BE: benzoylecgonine; CUD: cocaine use disorder; NMA: network meta-analysis; UDS: urine drug screen.

### Searches and study selection

Eligible RCTs were identified in two stages. First, we searched MEDLINE-Ovid (2^nd^ February 2024) for relevant systematic reviews from the preceding 5 years and created a list of all RCTs included in eight identified reviews (11,15,16,18–22). Full text reports of all identified RCTs (n=237) were assessed for eligibility by two reviewers independently, using the LaserAI platform (www.laser.ai). In a second stage one year later, we searched for primary studies in MEDLINE-Ovid, PsycINFO-Ovid, and the Cochrane Central Register of Controlled Trials (CENTRAL) (2^nd^ February 2025). Full search strategies in Supporting information S1.2). No date restrictions were applied to this search, although results were restricted to reports published in English. Primary study search results were screened by two reviewers independently. Discrepancies at any stage were resolved by discussion with a third reviewer or the wider review team.

### Data extraction and processing

Standardized forms, created and piloted in LaserAI, were used to extract study information by a single reviewer, and then cross-checked in detail by a second reviewer. Discrepancies were resolved by discussion with a third reviewer or the wider review team. During extraction, AI model suggestions from LaserAI were used, with all suggestions adjudicated by a human reviewer.

For dichotomous outcomes, we extracted arm-level counts of participants with event, number of participants with available outcome data, number of participants retained at end of treatment, and number of participants randomized into each arm. The choice of denominator used for each analysis depended on the type of outcome. For safety and acceptability outcomes, the denominator was the number of participants randomized. For effectiveness outcomes, where it was reported, the participants included in analyses was the denominator, otherwise we used the number of participants who completed treatment.

An extraction hierarchy was established for continuous outcomes. Following NICE TSU guidance (23), continuous outcome data was extracted according to a hierarchy with preference for means and standard deviations of change from baseline (CfB). For multi-arm trials including two or more of the same medication (e.g., two different doses of the same drug), we pooled the outcome data into a single arm for subsequent analysis before including them in NMAs. This approach prevents double-counting participants and avoids including correlated comparisons in the analyses.

### Risk of bias assessment

We assessed risk of bias using the Cochrane Risk of Bias 2 tool (RoB2) (24) at the results level for all outcomes included in quantitative synthesis. Risk of bias was assessed by a single reviewer and checked in detail by a second reviewer. Discrepancies were resolved by discussion with a third reviewer or the wider review team.

### Synthesis of results

We conducted a NMA, allowing the relative effects of all pharmacological interventions to be compared simultaneously in a single analysis (25). We fitted all NMA models in a Bayesian framework using the multinma package (26) in R (v4.5.1) (27). Random-effects models were used as the primary approach (assuming a common between-study heterogeneity standard deviation (SD), which we denote as τ), with common-effect models fitted as sensitivity analyses. Before undertaking the synthesis, we assessed transitivity by examining the similarity of the distribution of potential effect modifiers across the comparisons in the network. Potential effect modifiers were of intervention duration, treatment setting, age, and co-occurring SUD. Global consistency was examined by comparing model fit between a model assuming consistency and a model allowing for inconsistency. If there was evidence of inconsistency between direct and indirect evidence, we planned further evaluation using a local loop-based approach (node-splitting) (23). Full analysis details, including specific priors, are detailed in Supporting information S1.3.

To aid interpretation, we re-expressed SMD results as MD based on the most commonly reported measure for a given outcome, which is articulated under each relevant subsection (full details of methods used for conversions and reinterpretations are presented in Supporting information S1.4). As such, results are expressed as MD for continuous outcomes and OR for dichotomous outcomes, with 95% credible intervals (CrI). Point estimates of relative effect represent the 50% quantile (median) of the posterior distribution, and 95% CrI represent its 2.5 and 97.5% quantiles.

#### Sensitivity and subgroup analyses

We planned to explore several potential effect modifiers, including: intervention type (i.e., studies designed to initiate abstinence vs those designed to prevent relapse), presence of co-occurring substance use disorder/comorbidity, predominant cocaine type used, presence of co-interventions (e.g., counselling or other psychosocial interventions), and setting and delivery of interventions (inpatient vs outpatient, supervised vs unsupervised). These were conducted on primary outcomes only, using a meta-regression approach (Supporting information S1.3).

#### Certainty of evidence

We assessed the certainty of the evidence using the CINeMA framework (28). Assessments were completed by one reviewer and checked thoroughly by a second. Any discrepancies were resolved by discussion with a third reviewer or the wider review team. In the absence of established thresholds for minimum clinically important differences (MCID), we used conservative estimates of standardized mean difference (SMD) = −0.2 or 0.2, and odds ratio (OR) <0.8 or >1.25, in line with our previous work. Our overall certainty of evidence judgements were reached by starting at high certainty and downgrading by one level for each domain with some concerns, and by two levels for each domain with major concerns.

## RESULTS

### Included studies

We included 218 reports of 163 studies, including 14871 participants (**Error! Reference source not found.**). 151 studies contributed data to at least one NMA. Eight (29–36) of the remaining 12 studies provided narrative outcomes only and four studies (37–40) contributed no outcomes to this review. Characteristics of included studies are presented in Table 2. The sample size of studies ranged between 14 and 464 participants (median: 81 participants). Studies predominantly recruited mixed gender samples, although one study recruited only females and 19 recruited only males. The mixed samples were predominantly male (median: 76%, range: 48-98%) and the average age ranged from 28 to 52 years (median: 39 years). Treatment duration ranged from 1 day to 34 weeks, with a median duration of 12 weeks. Most studies combined pharmacological treatments with adjunctive therapy (95%).

**Table 2.**
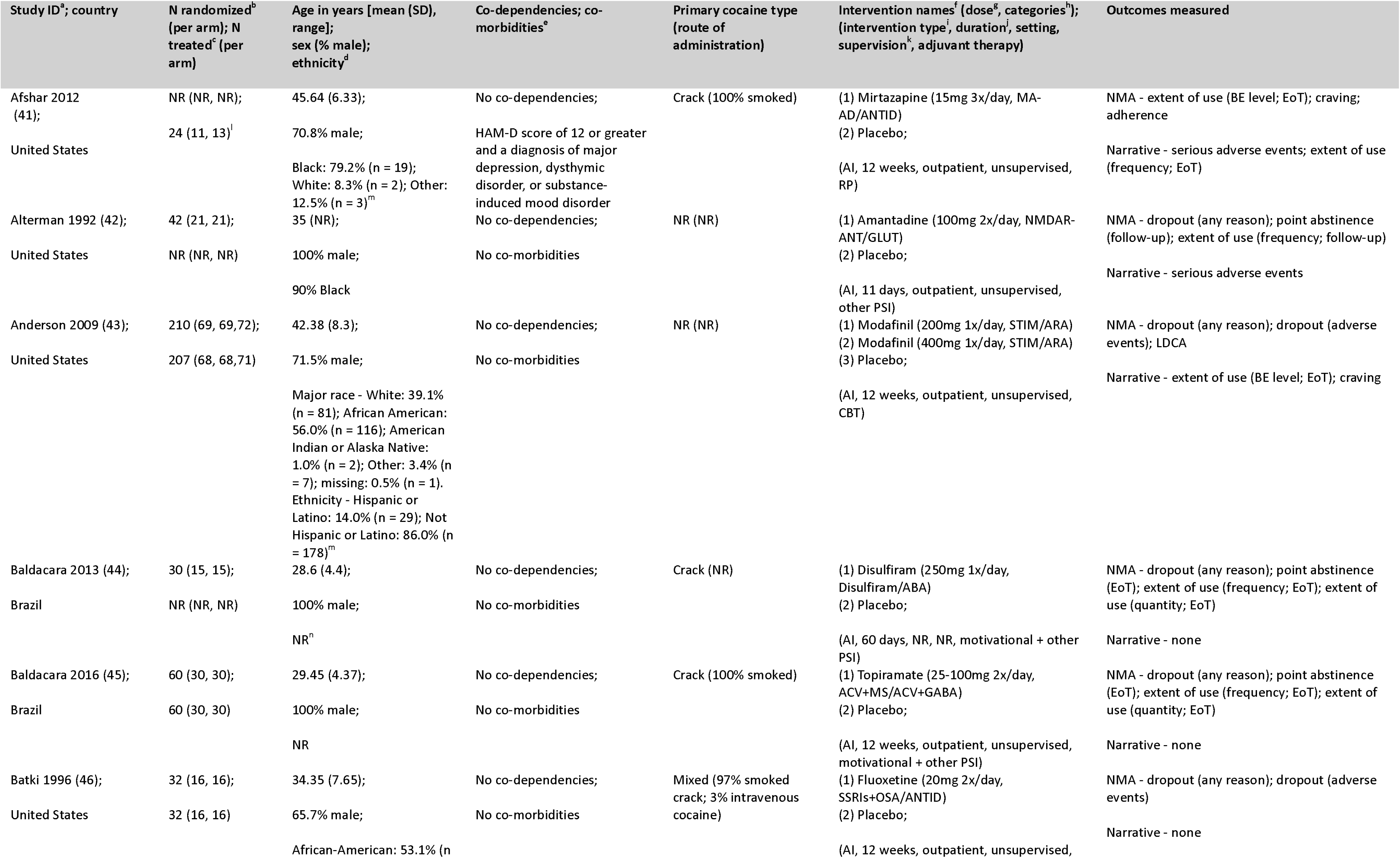
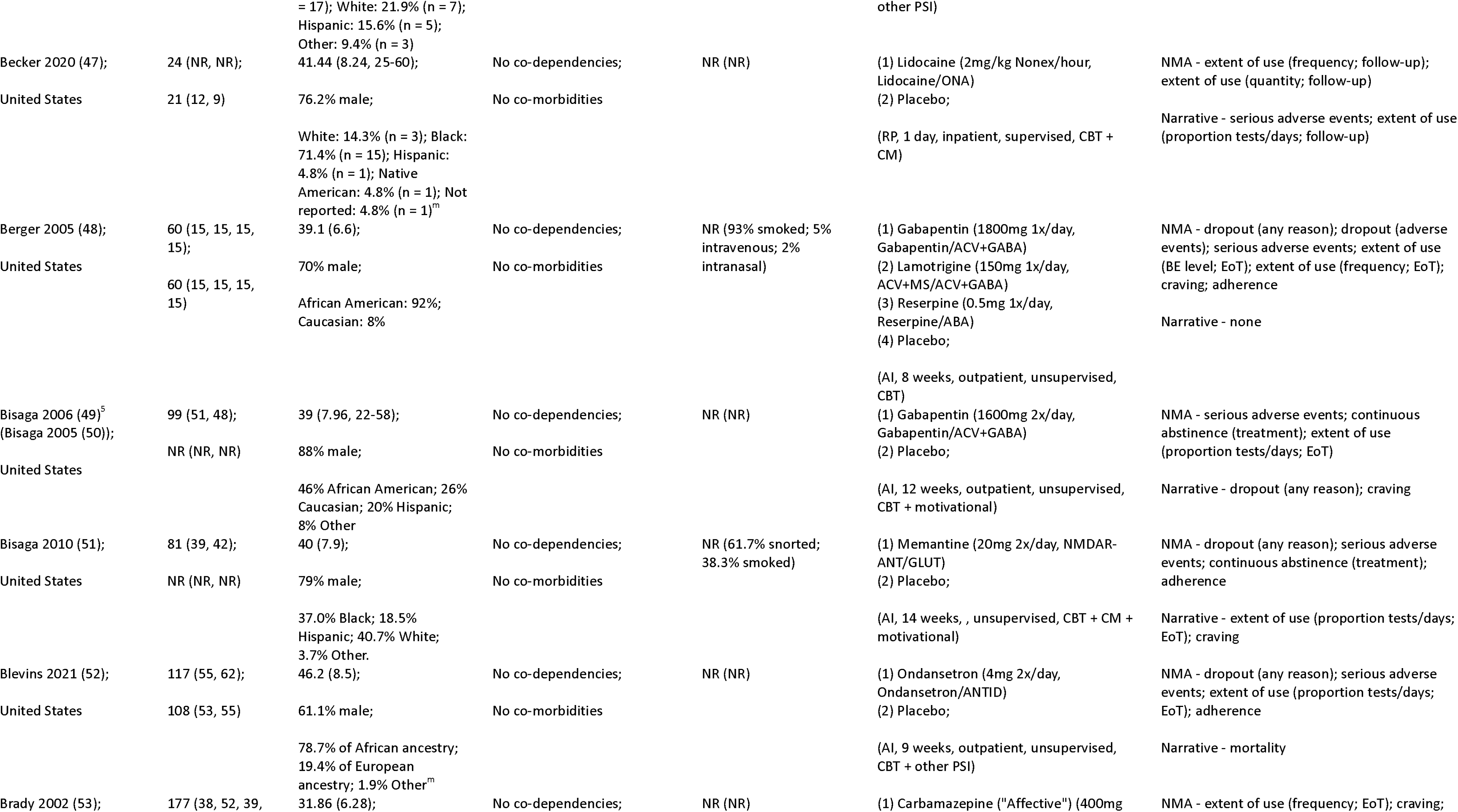
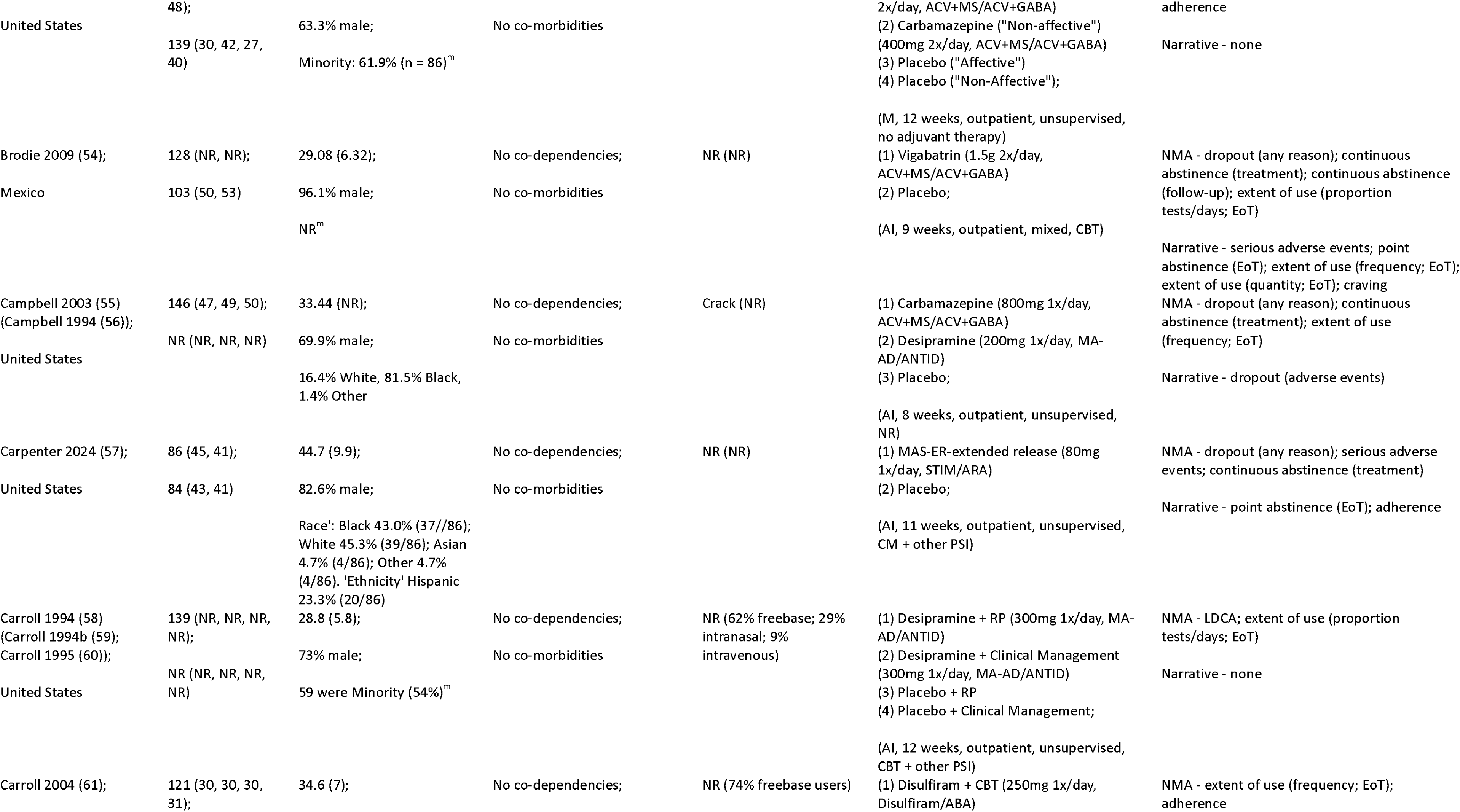
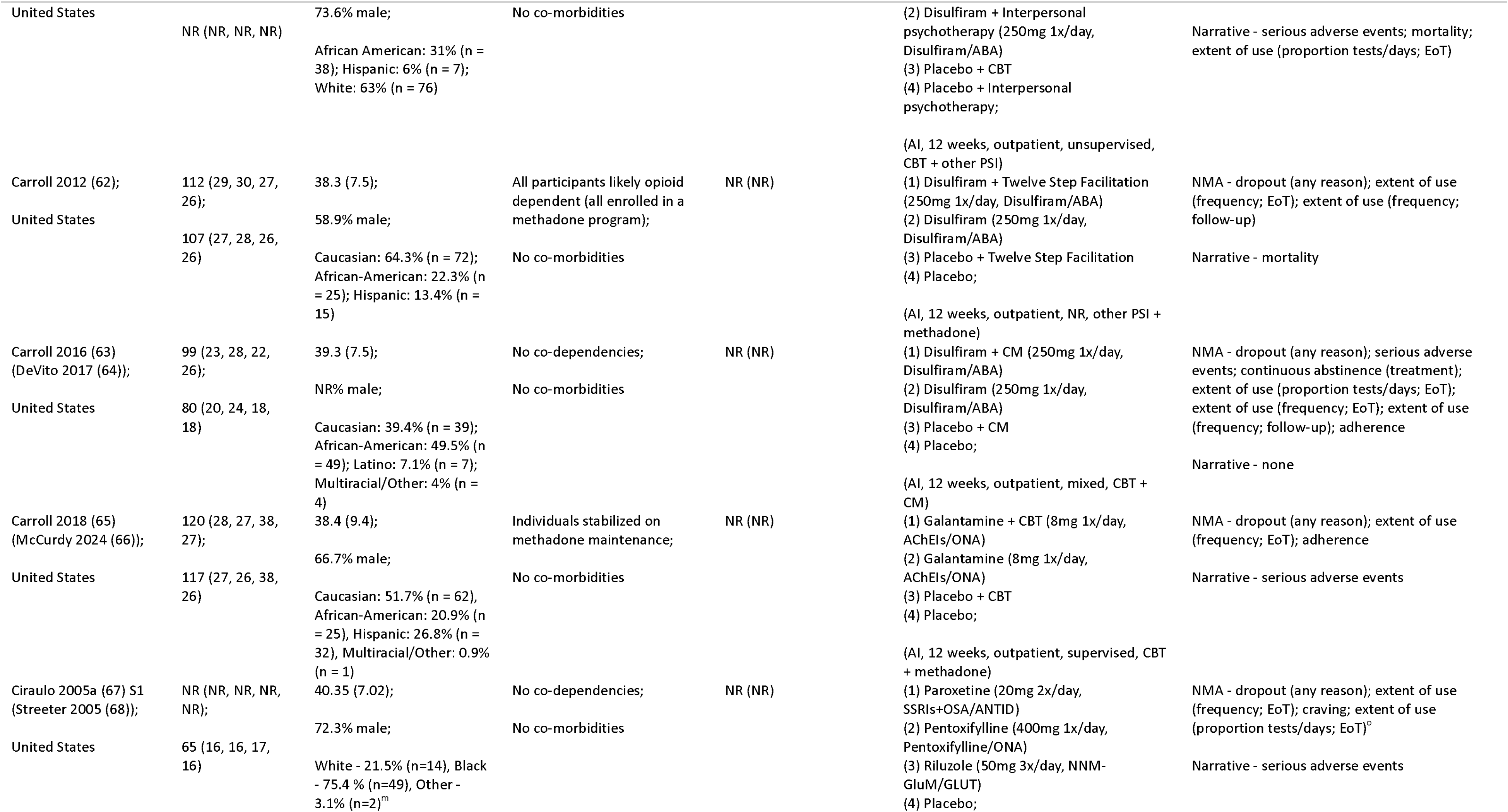
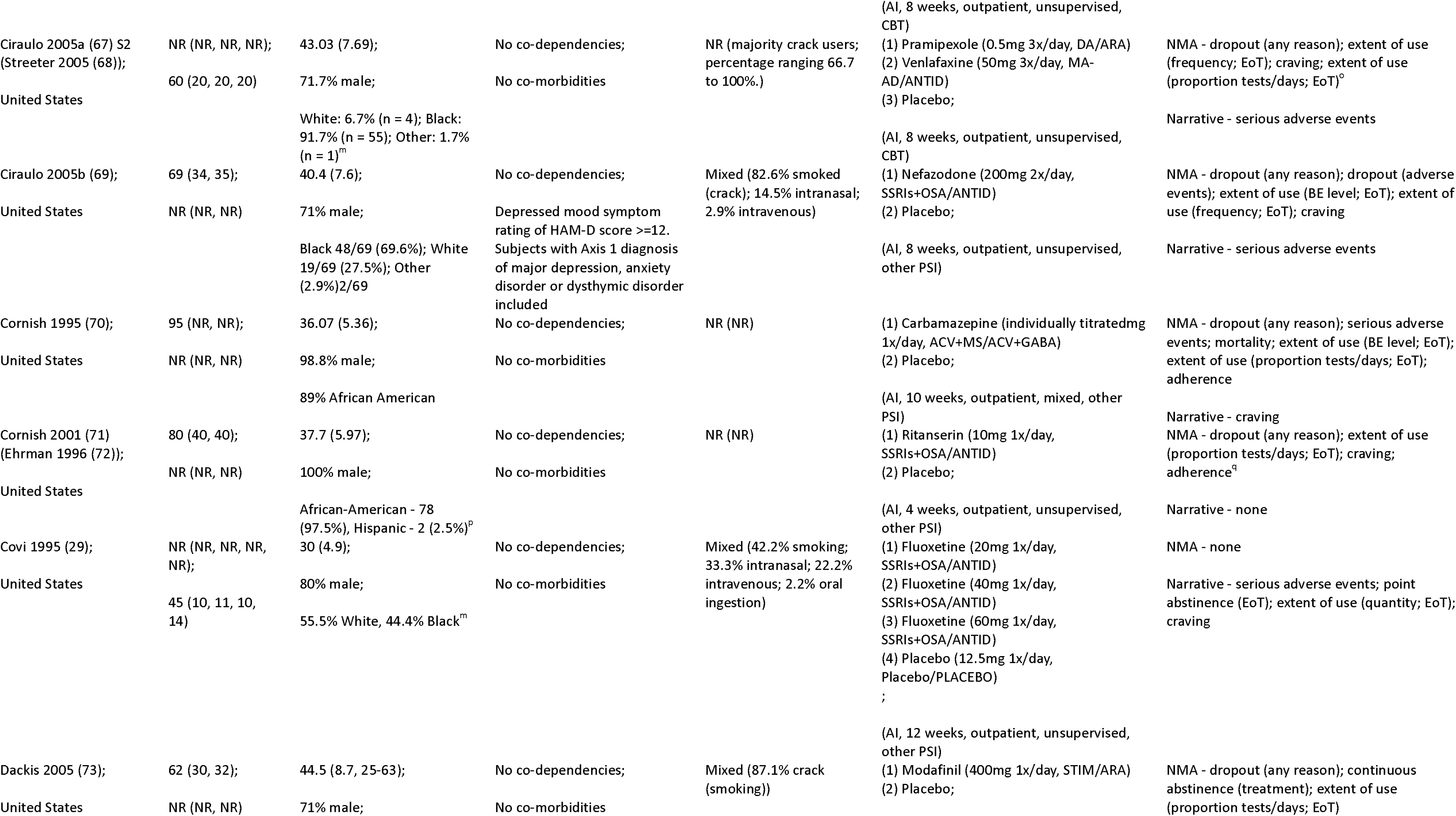
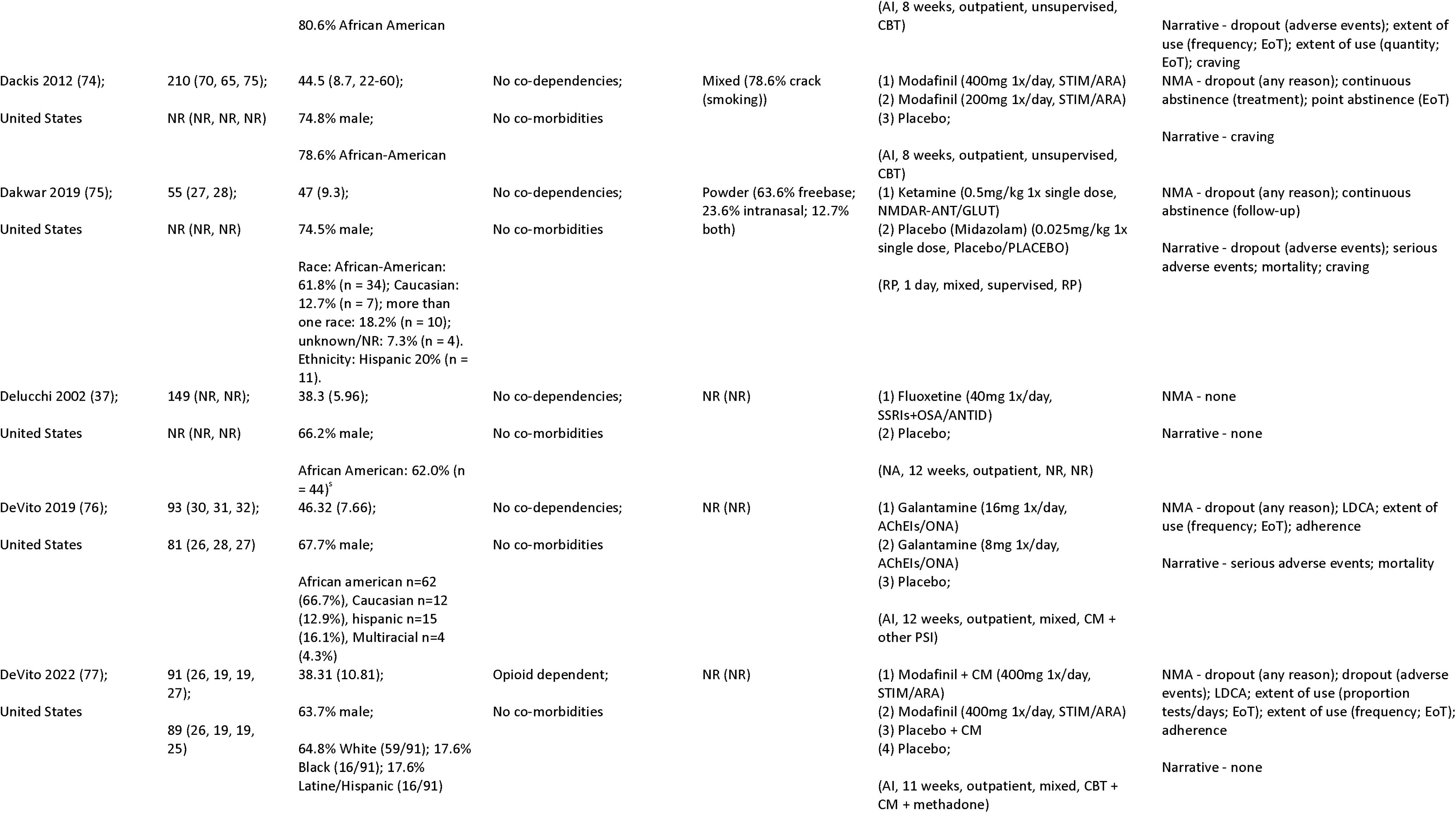
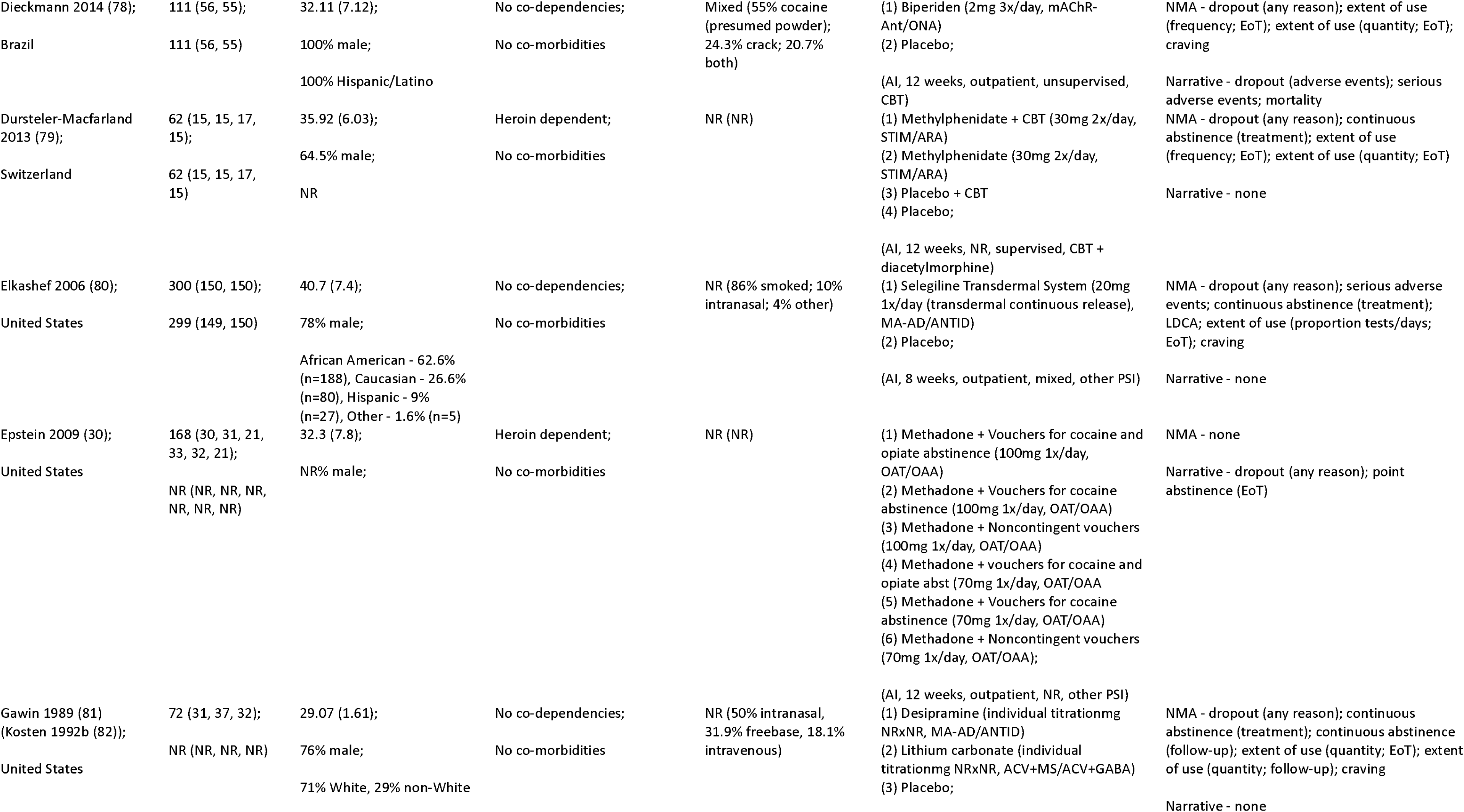
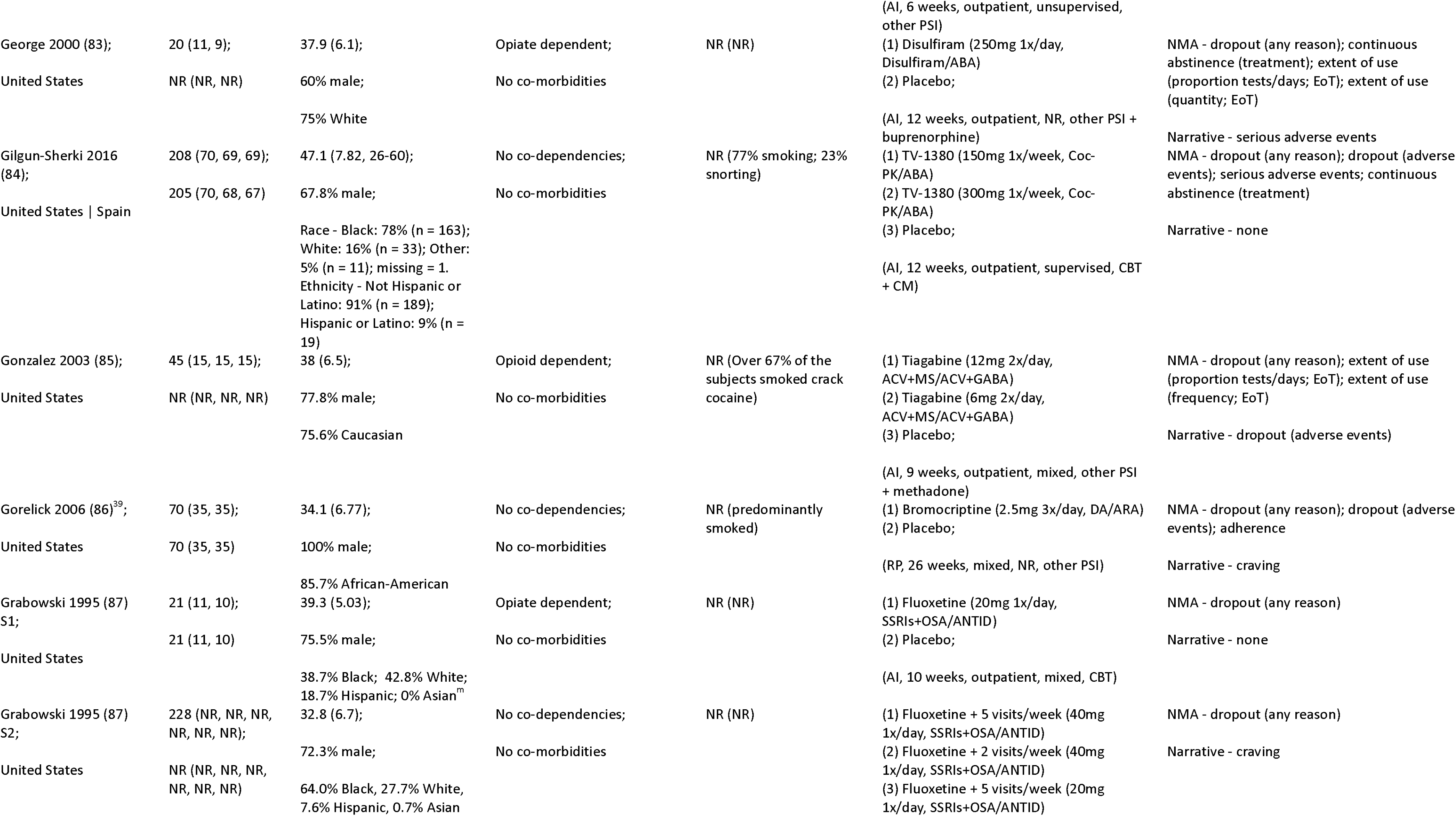
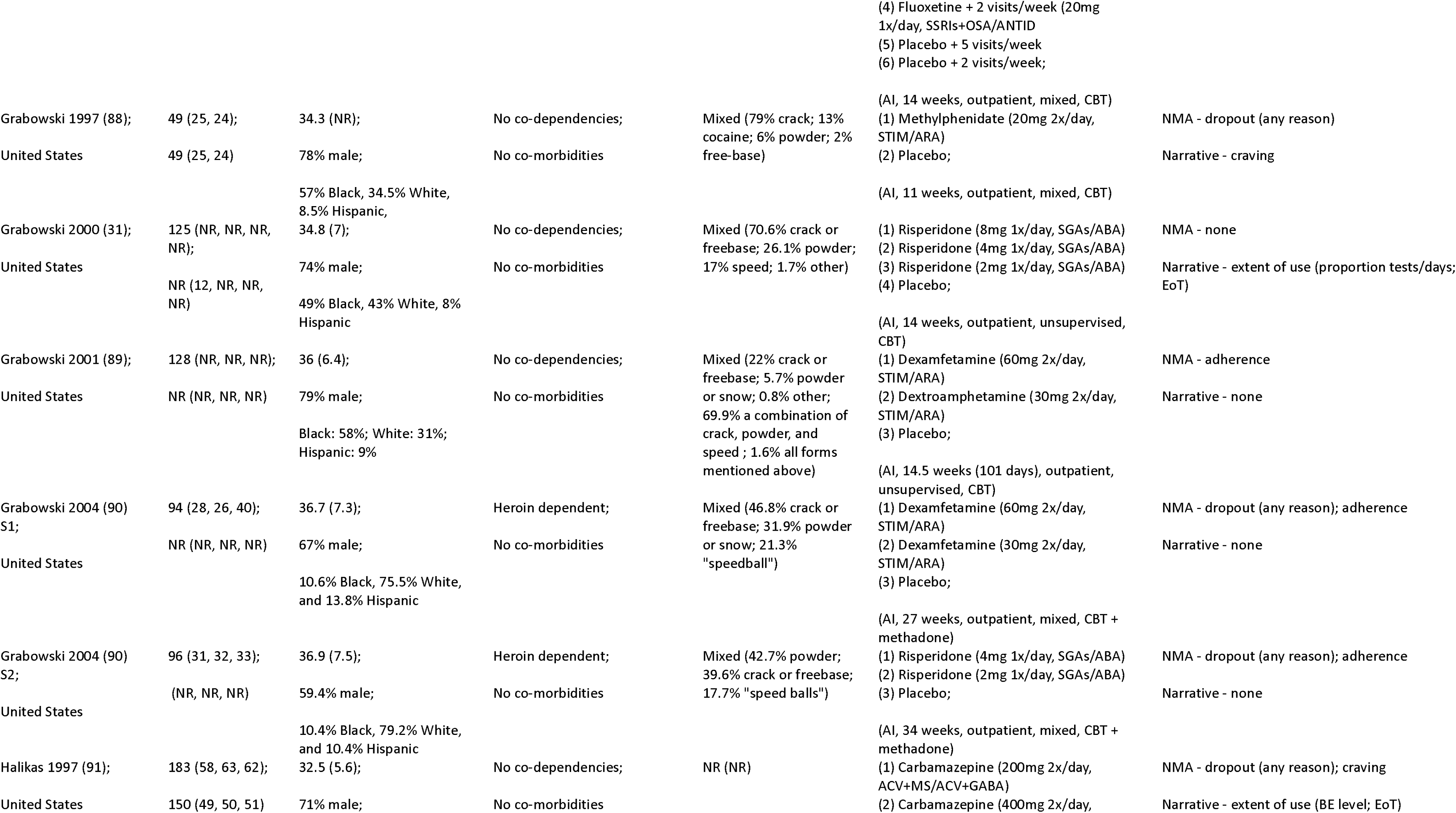
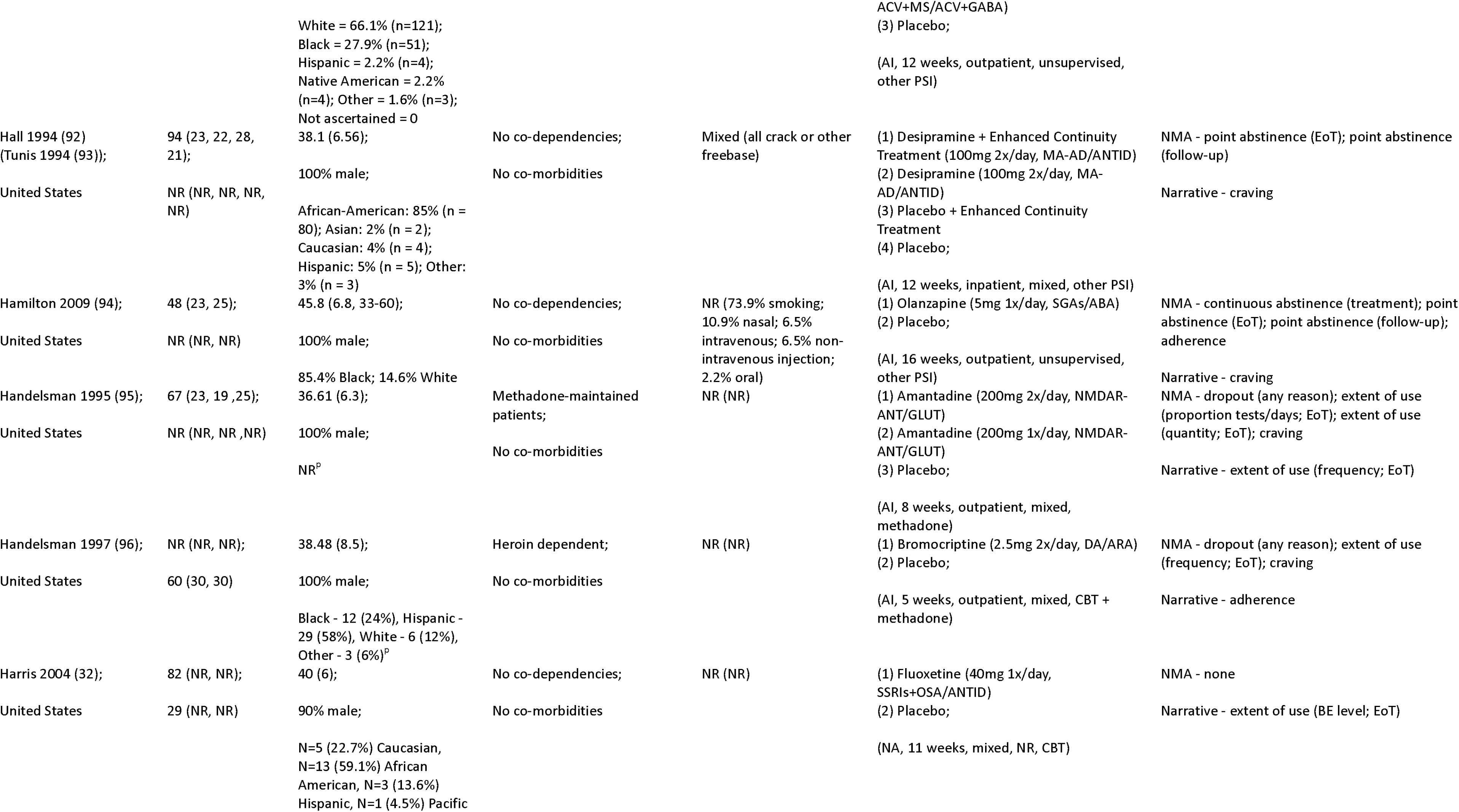
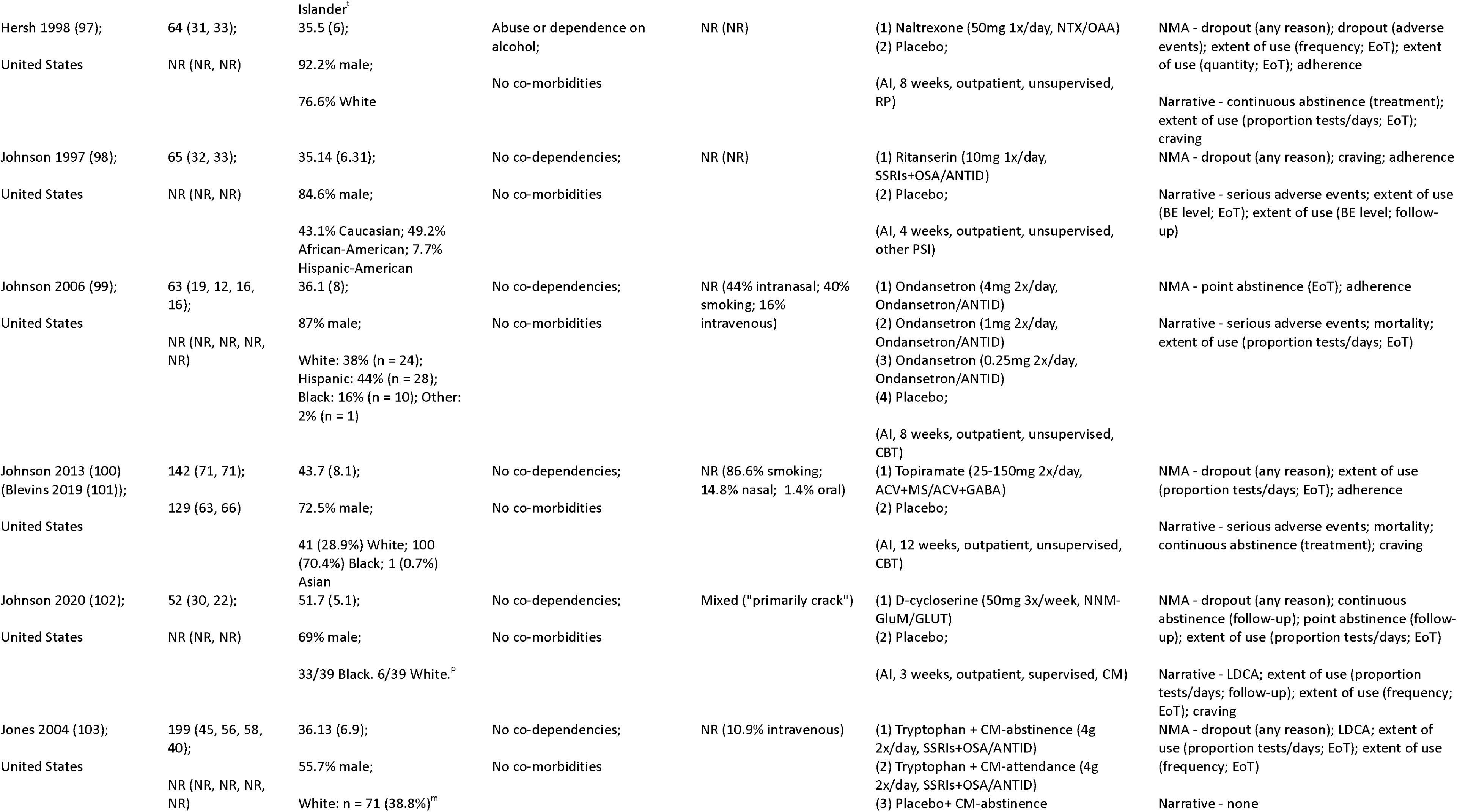
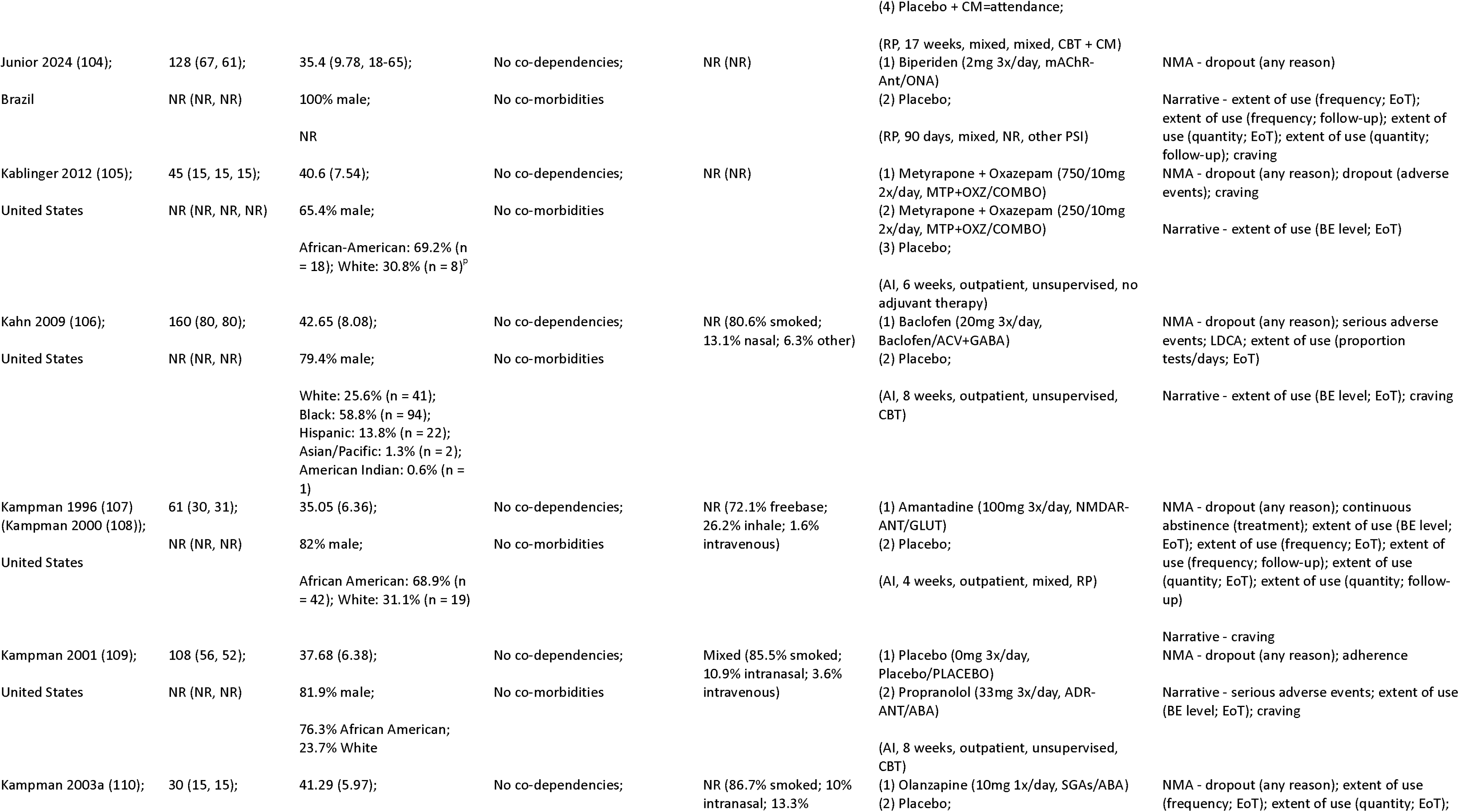
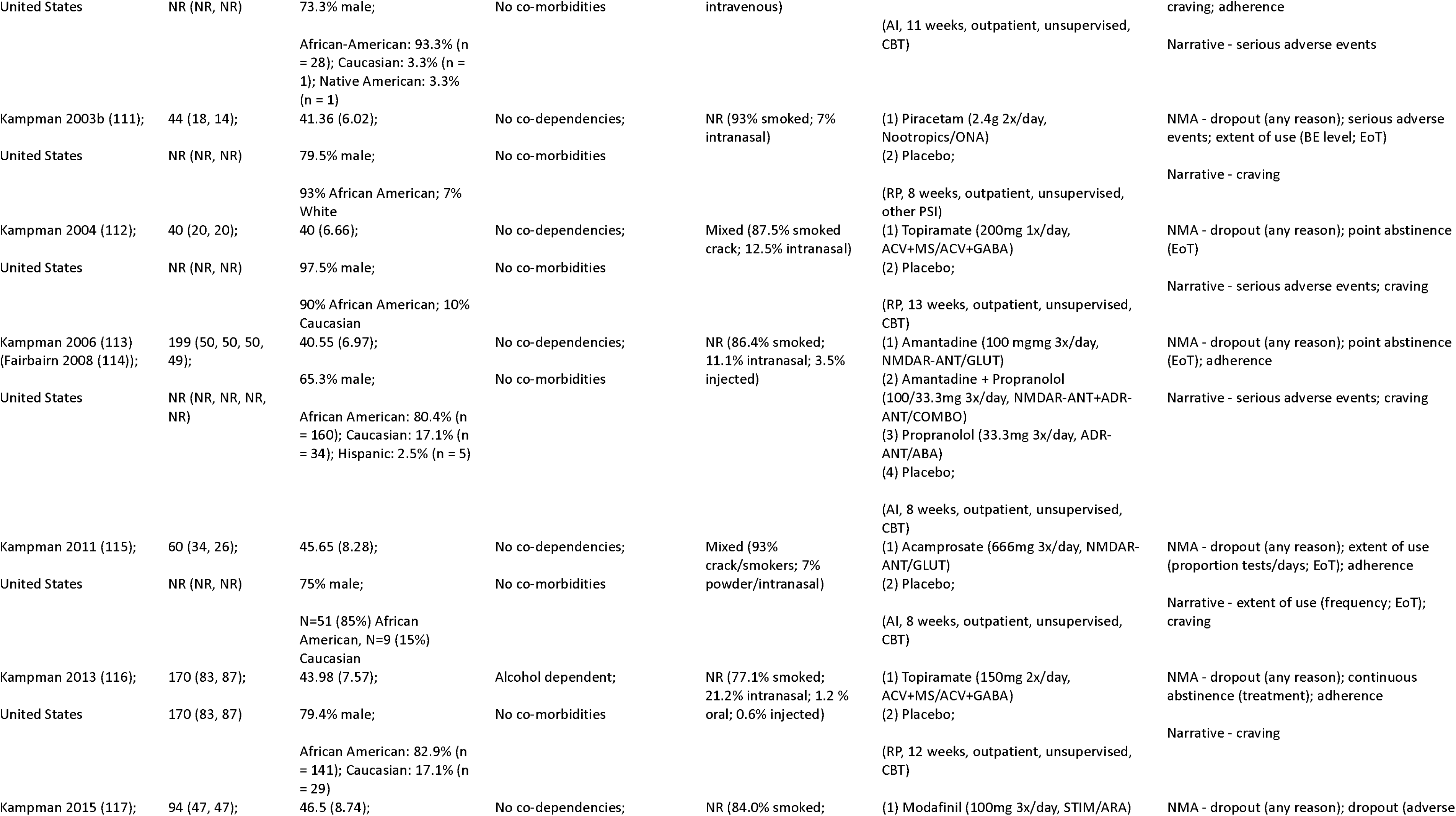
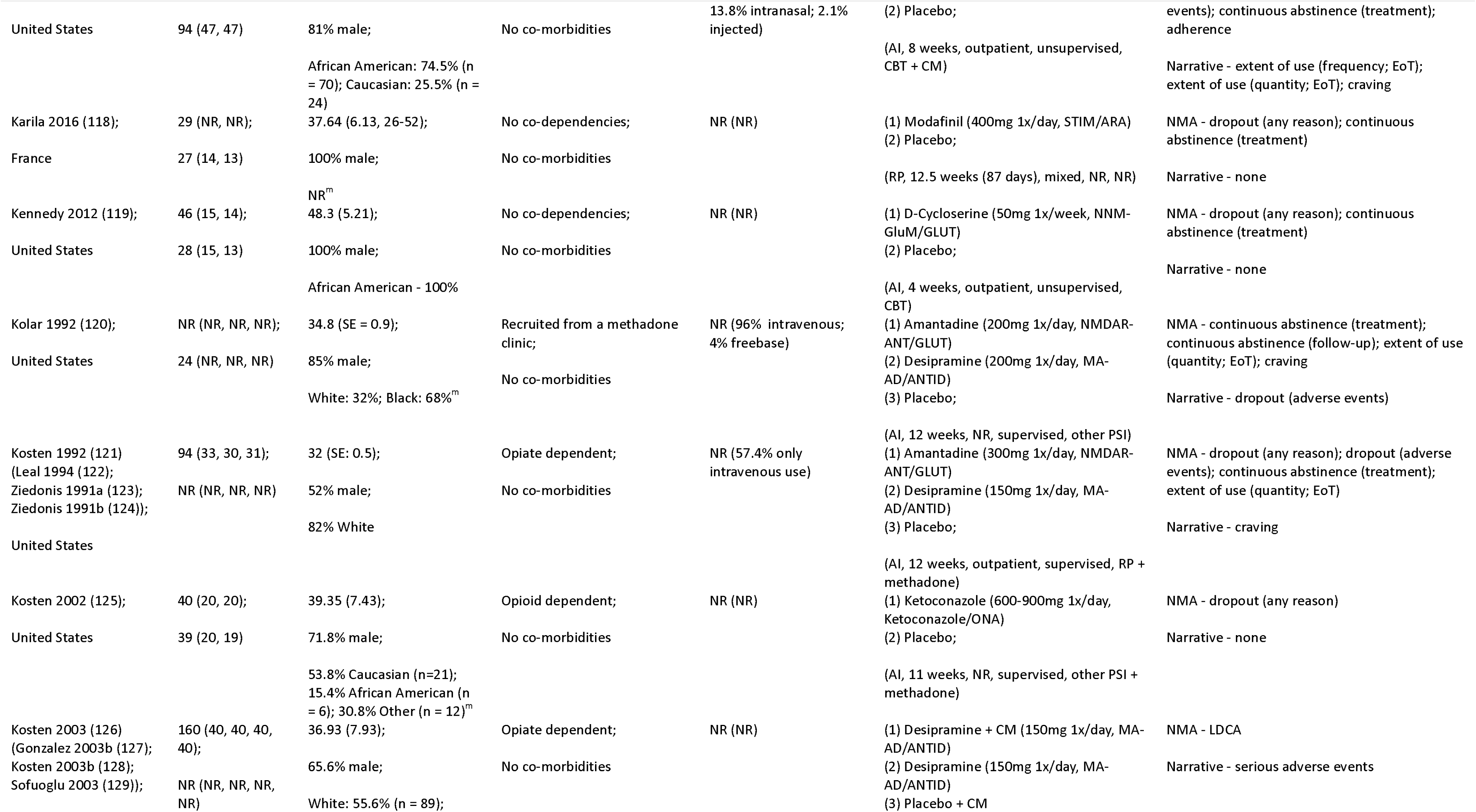
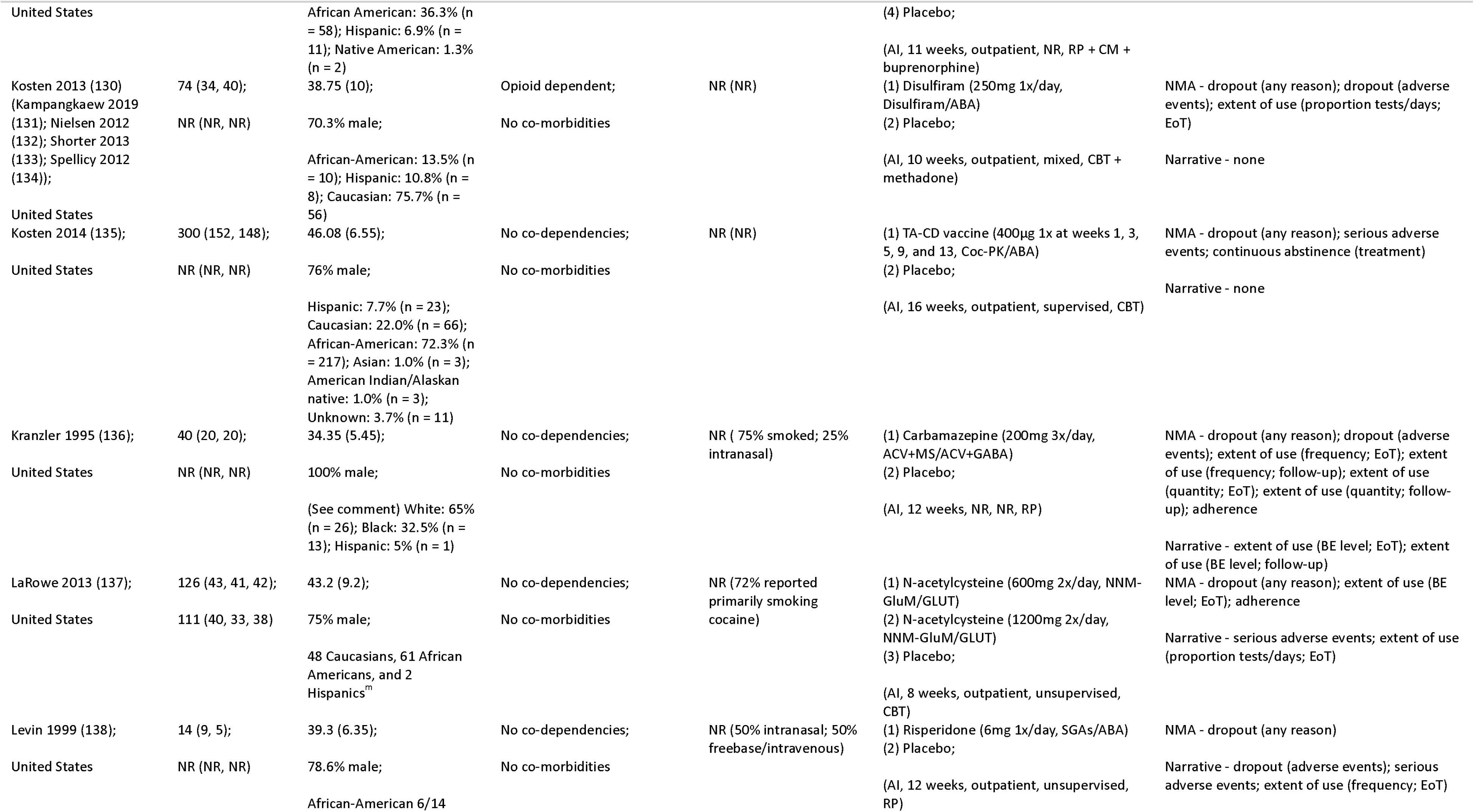
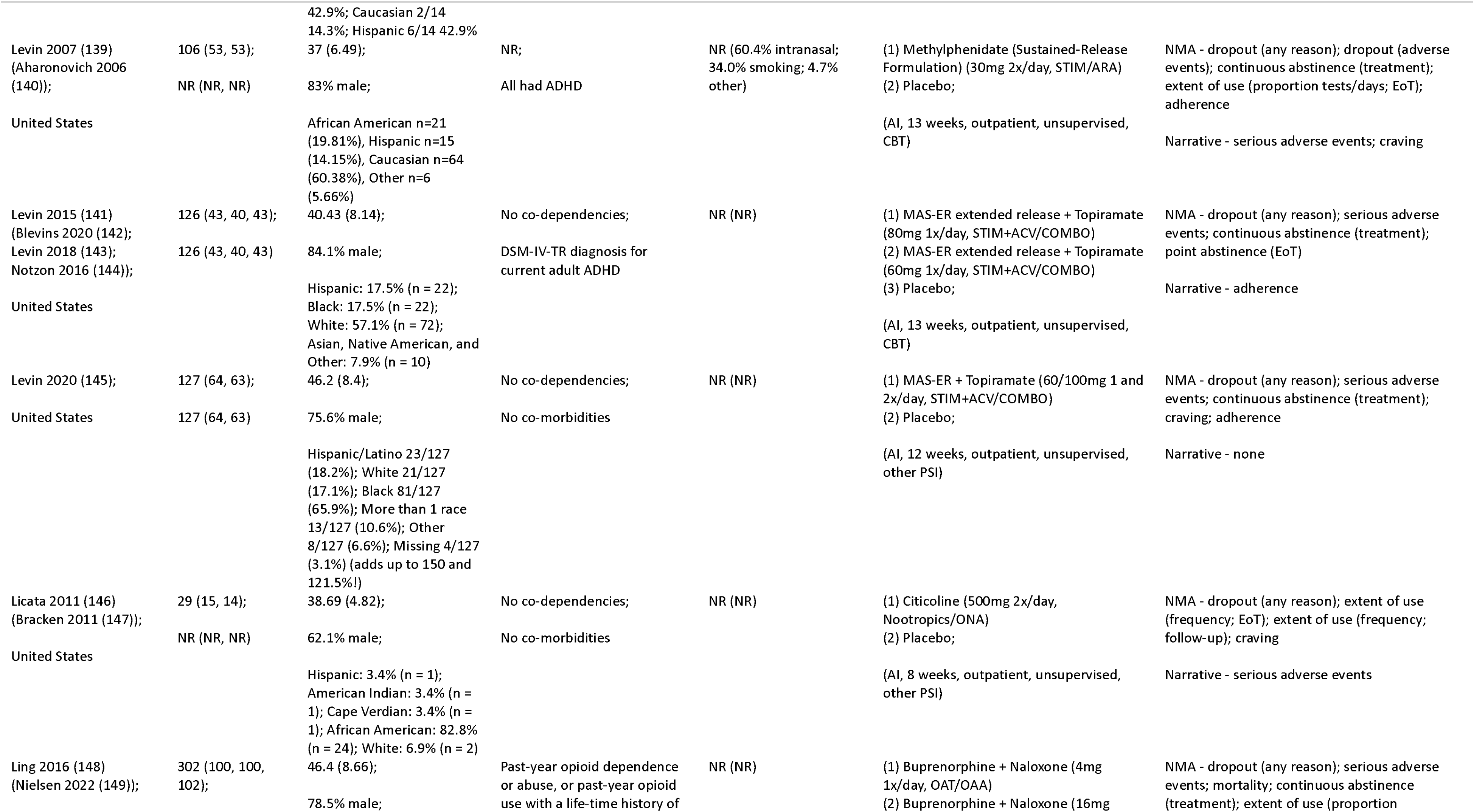
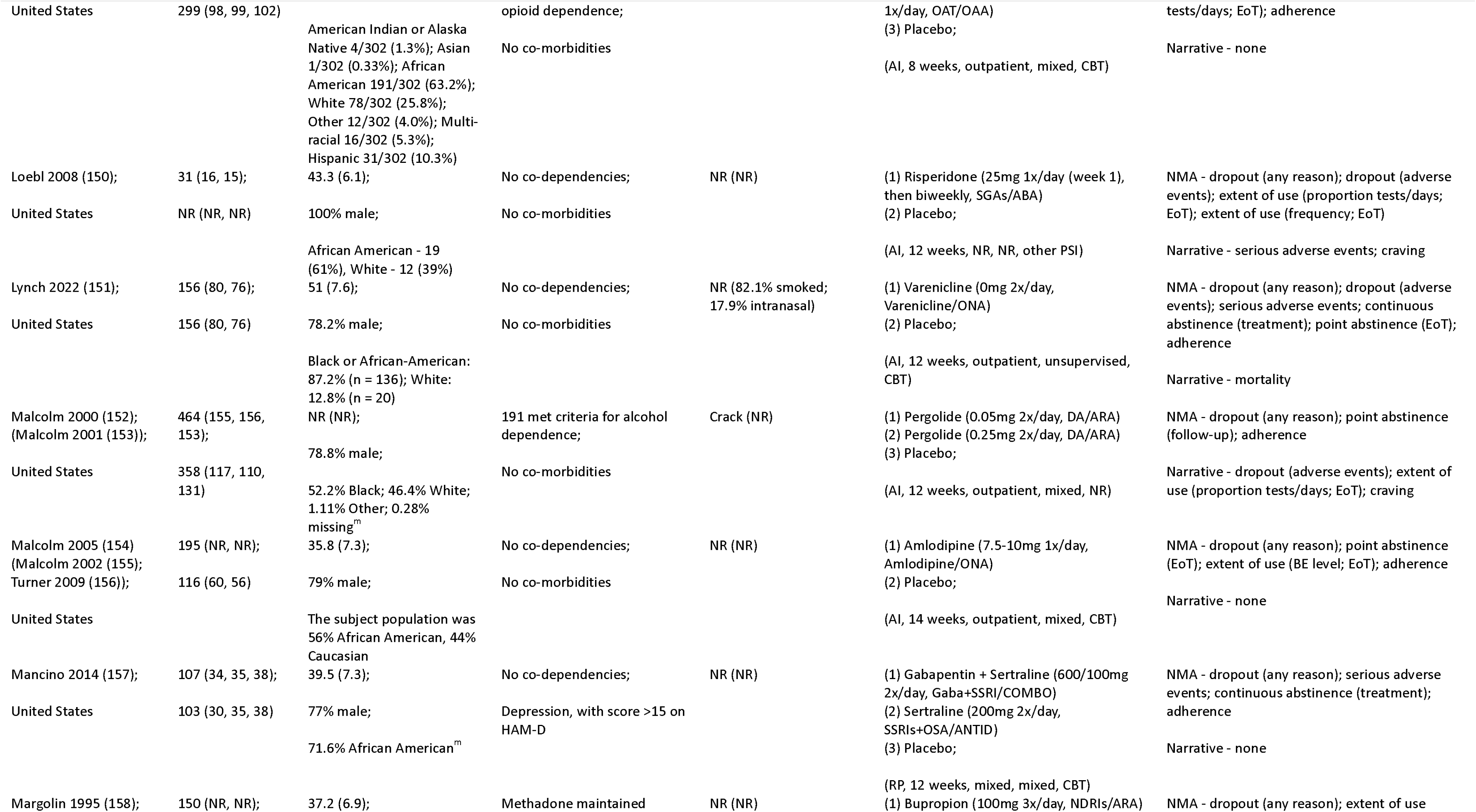
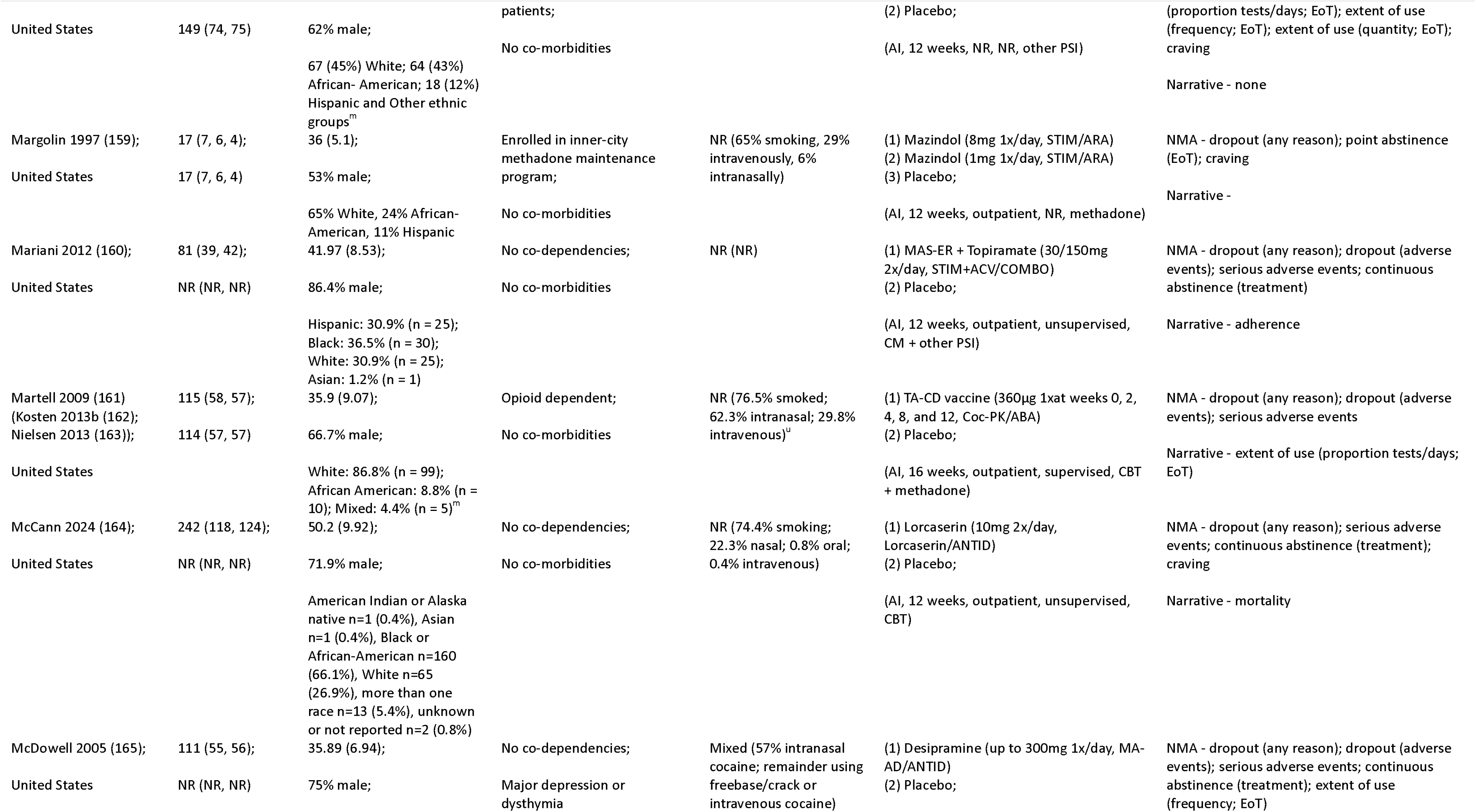
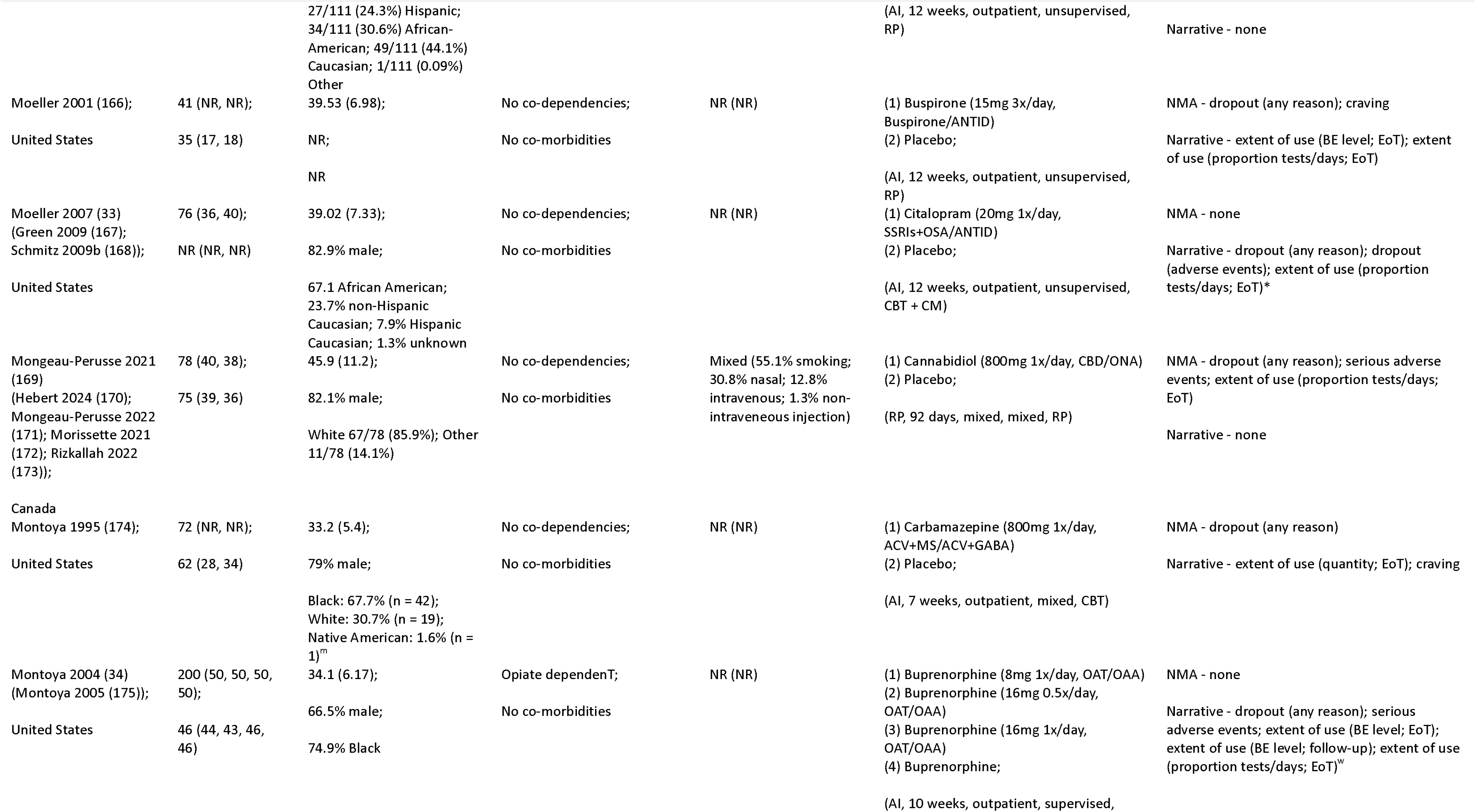
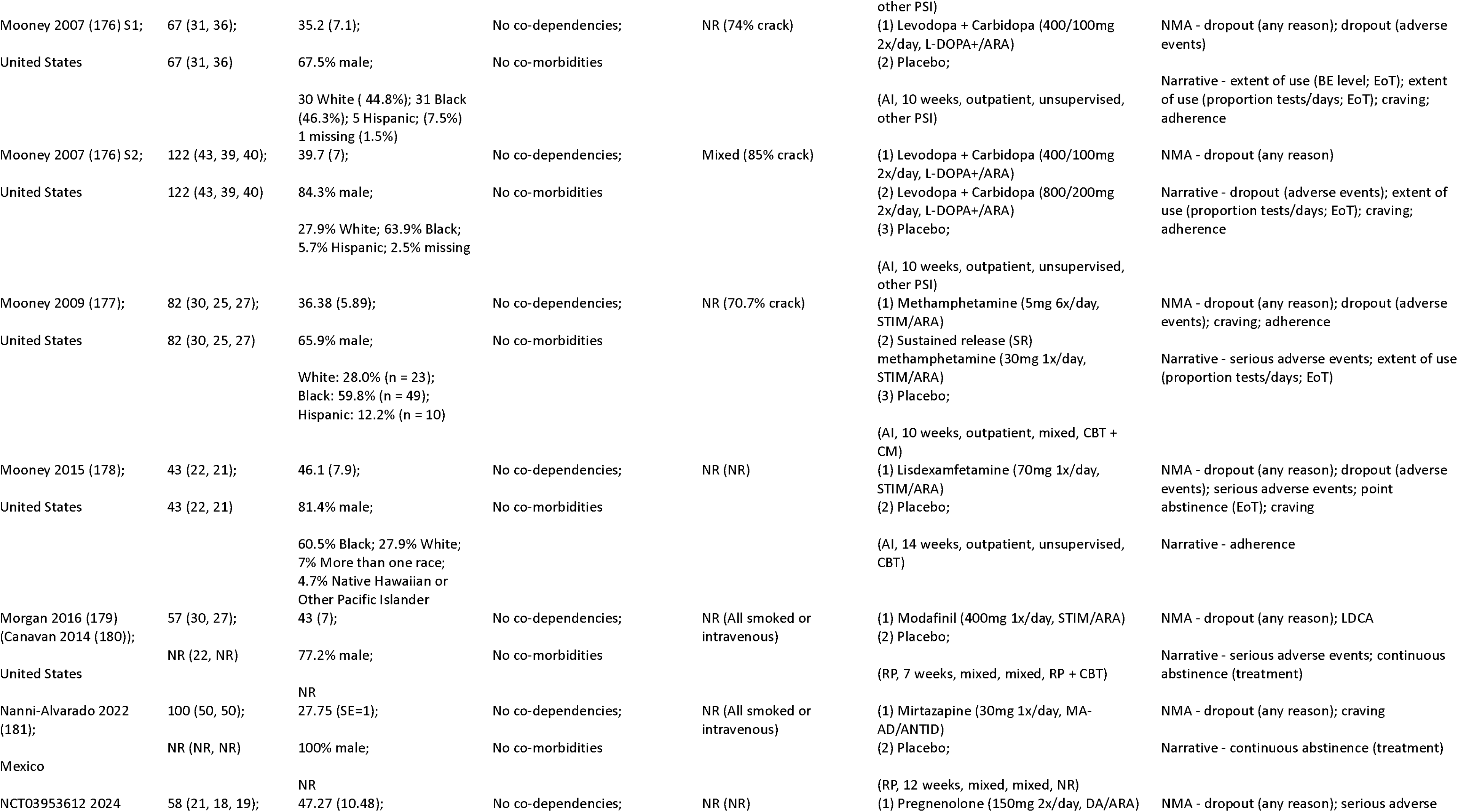
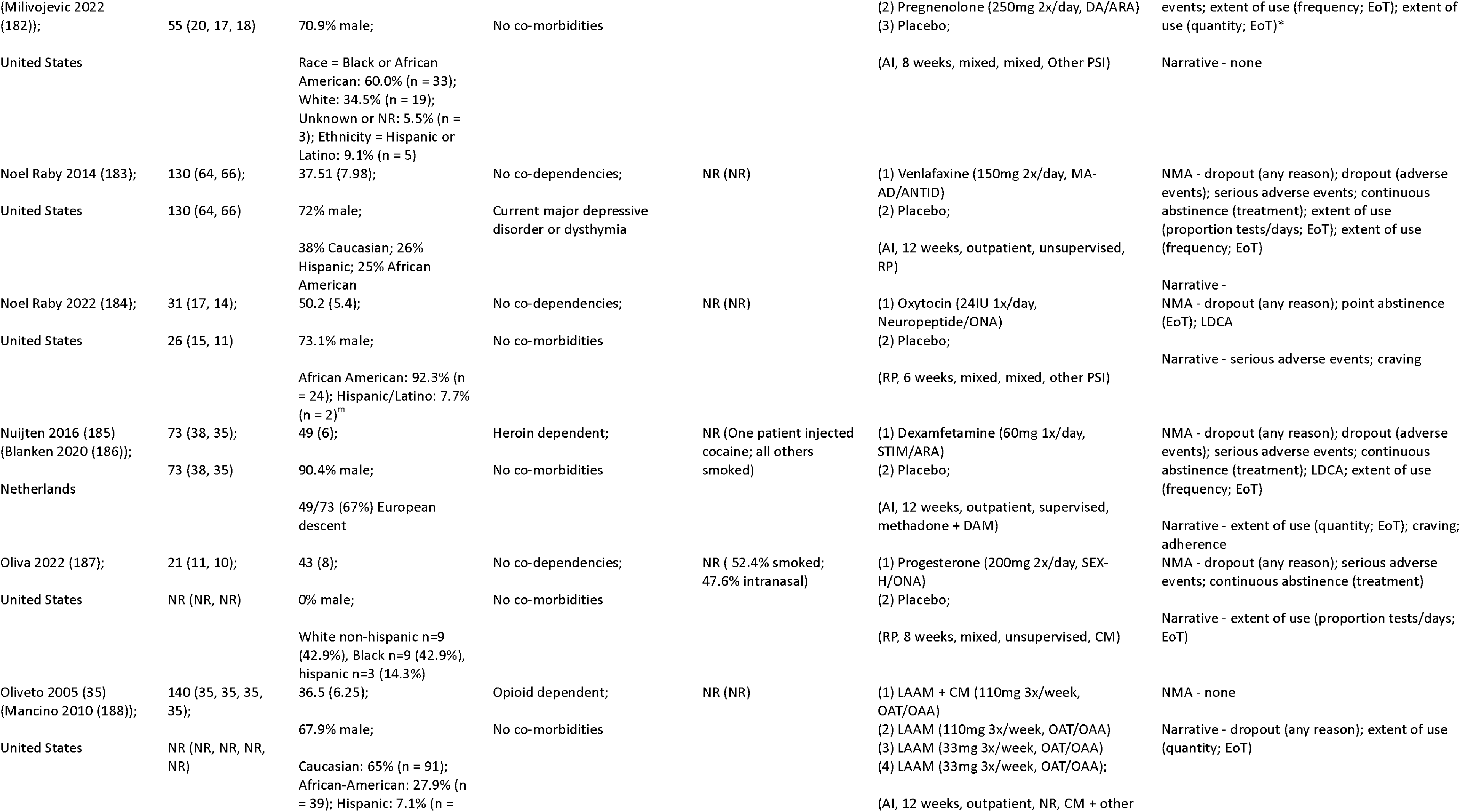
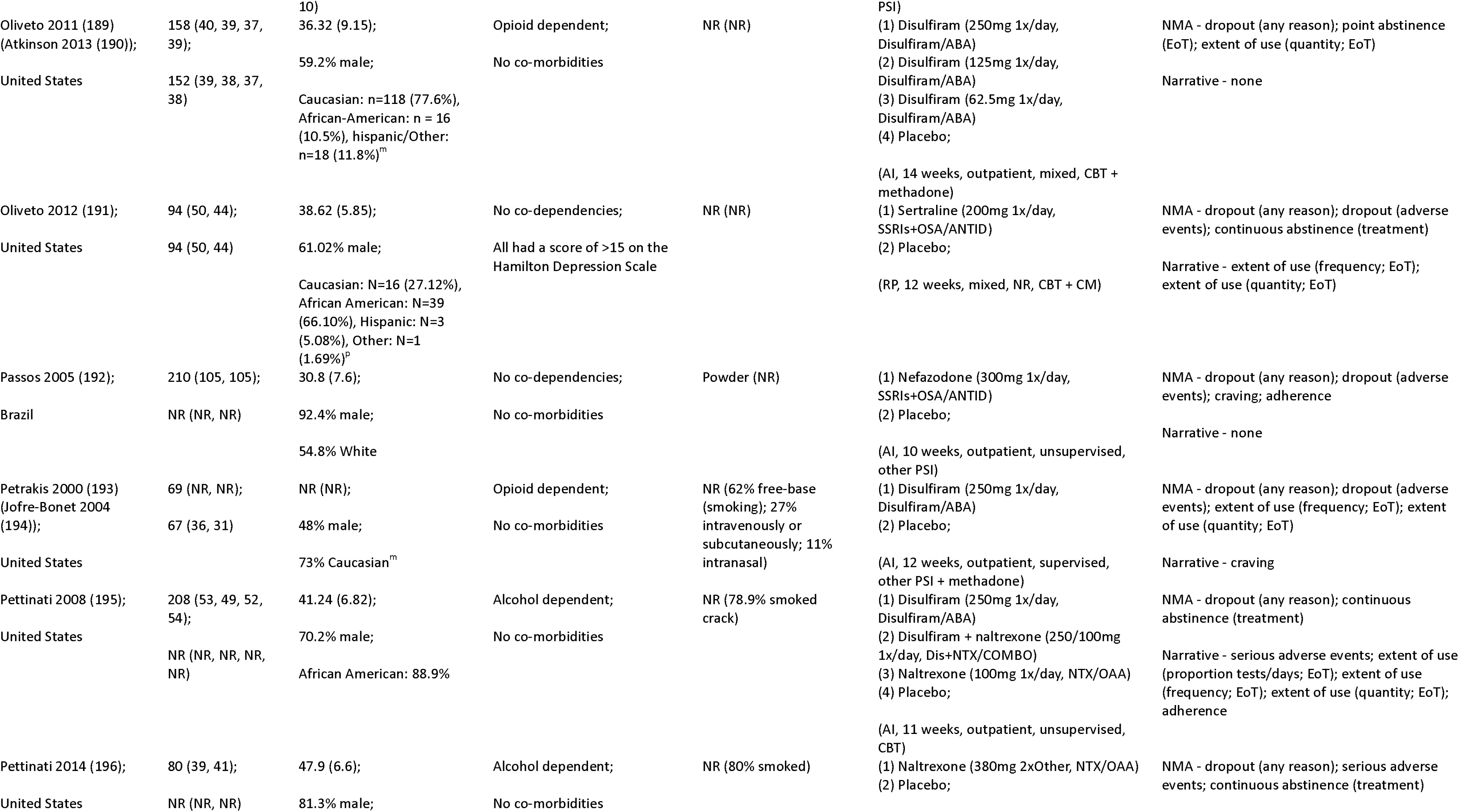
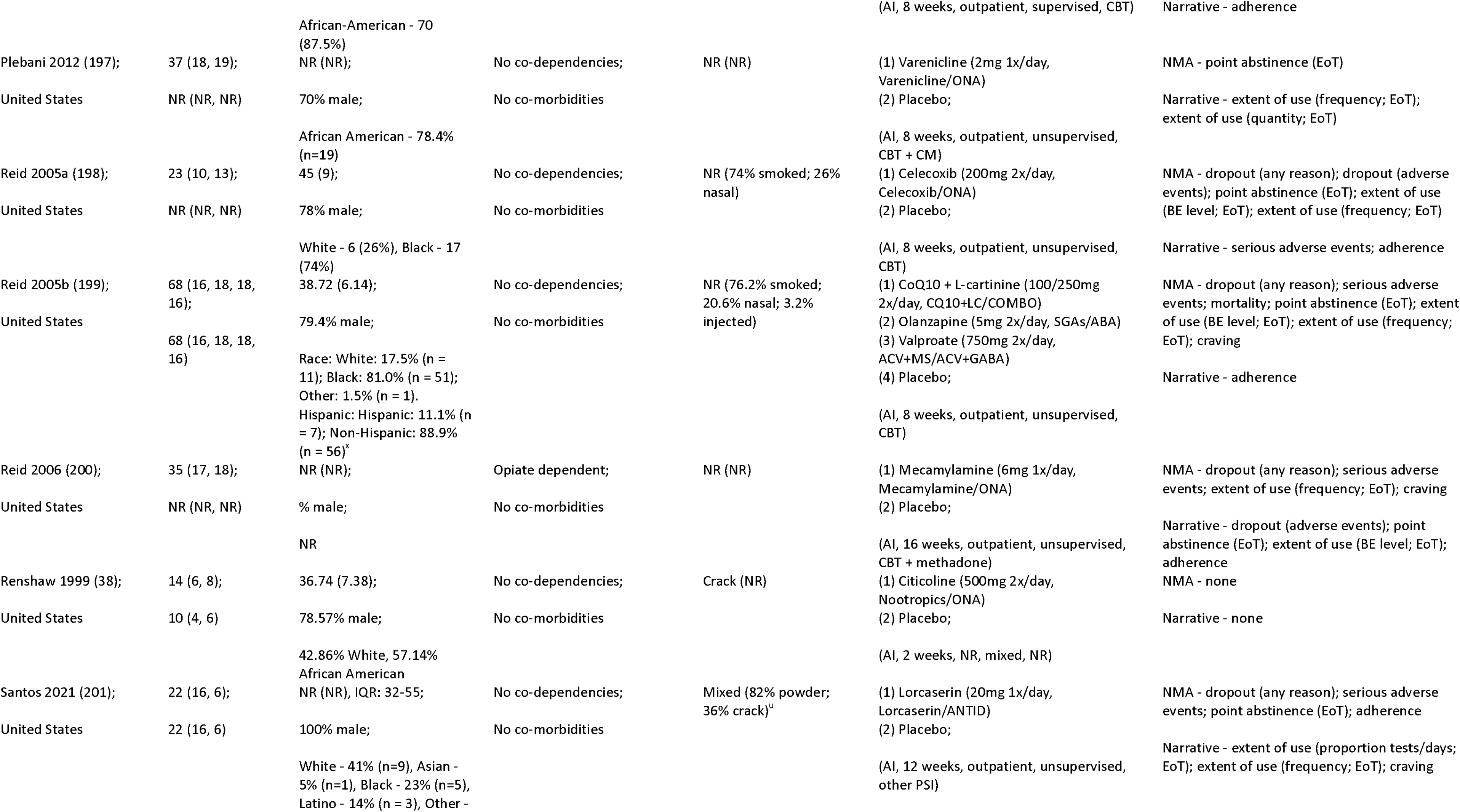
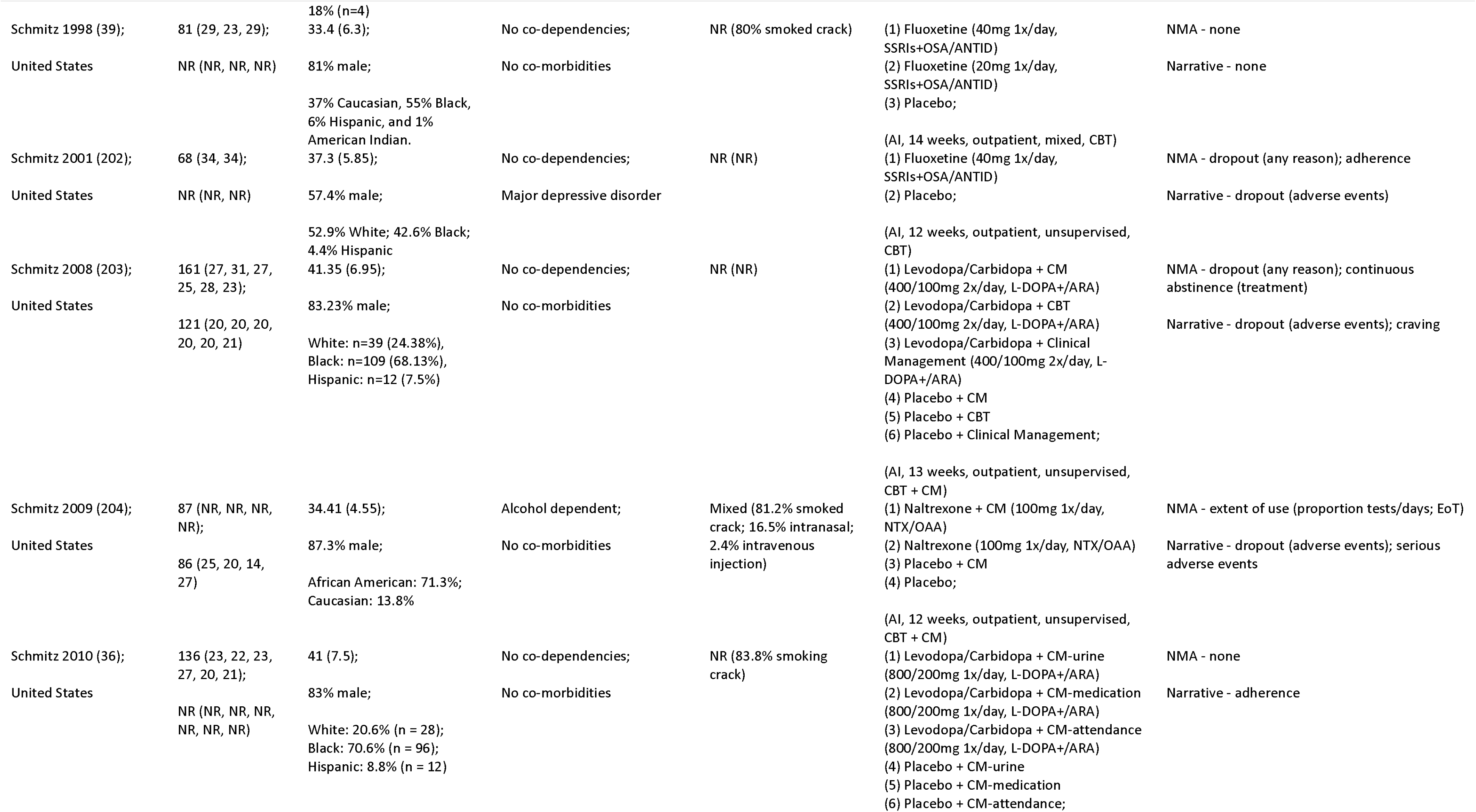
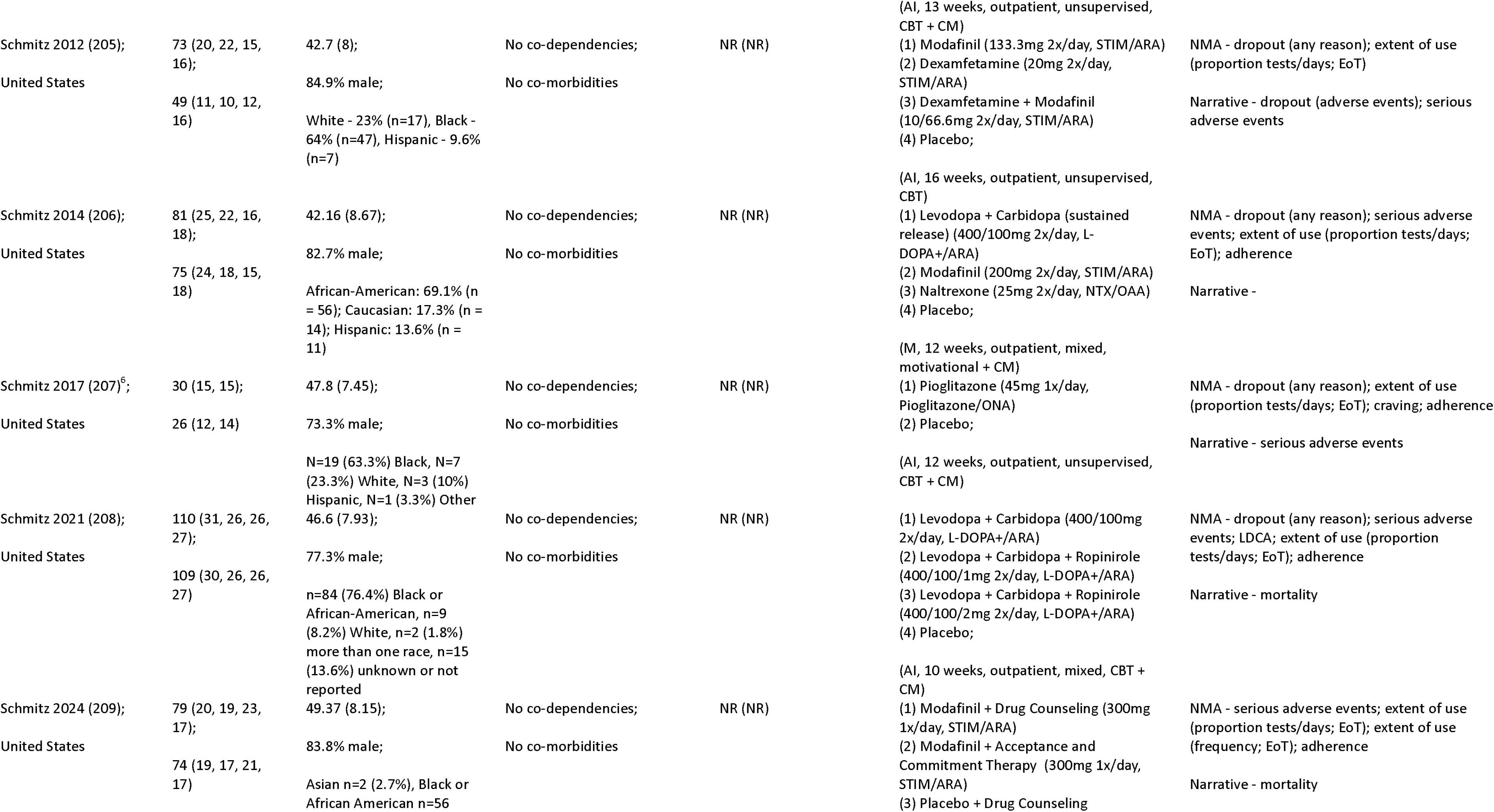
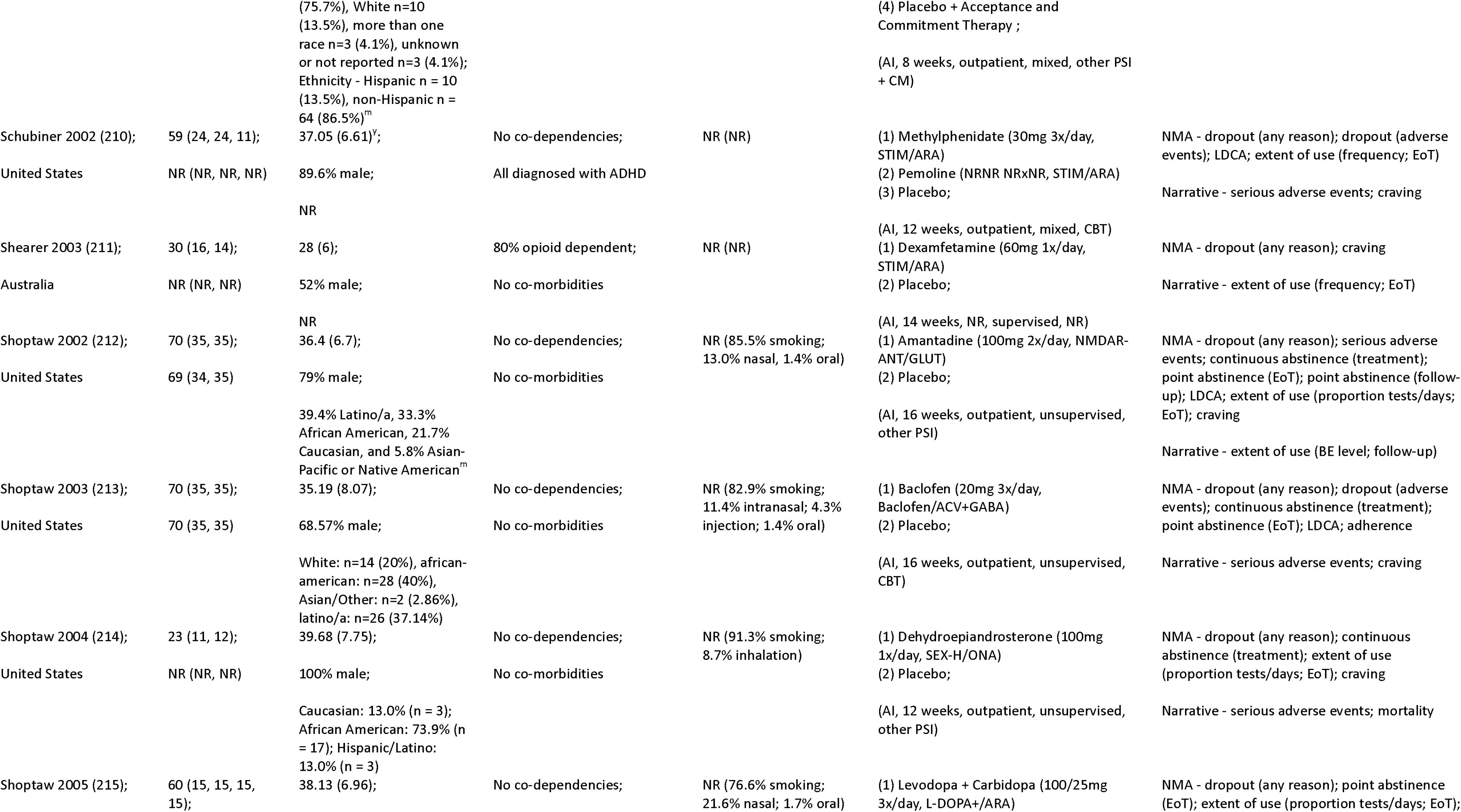
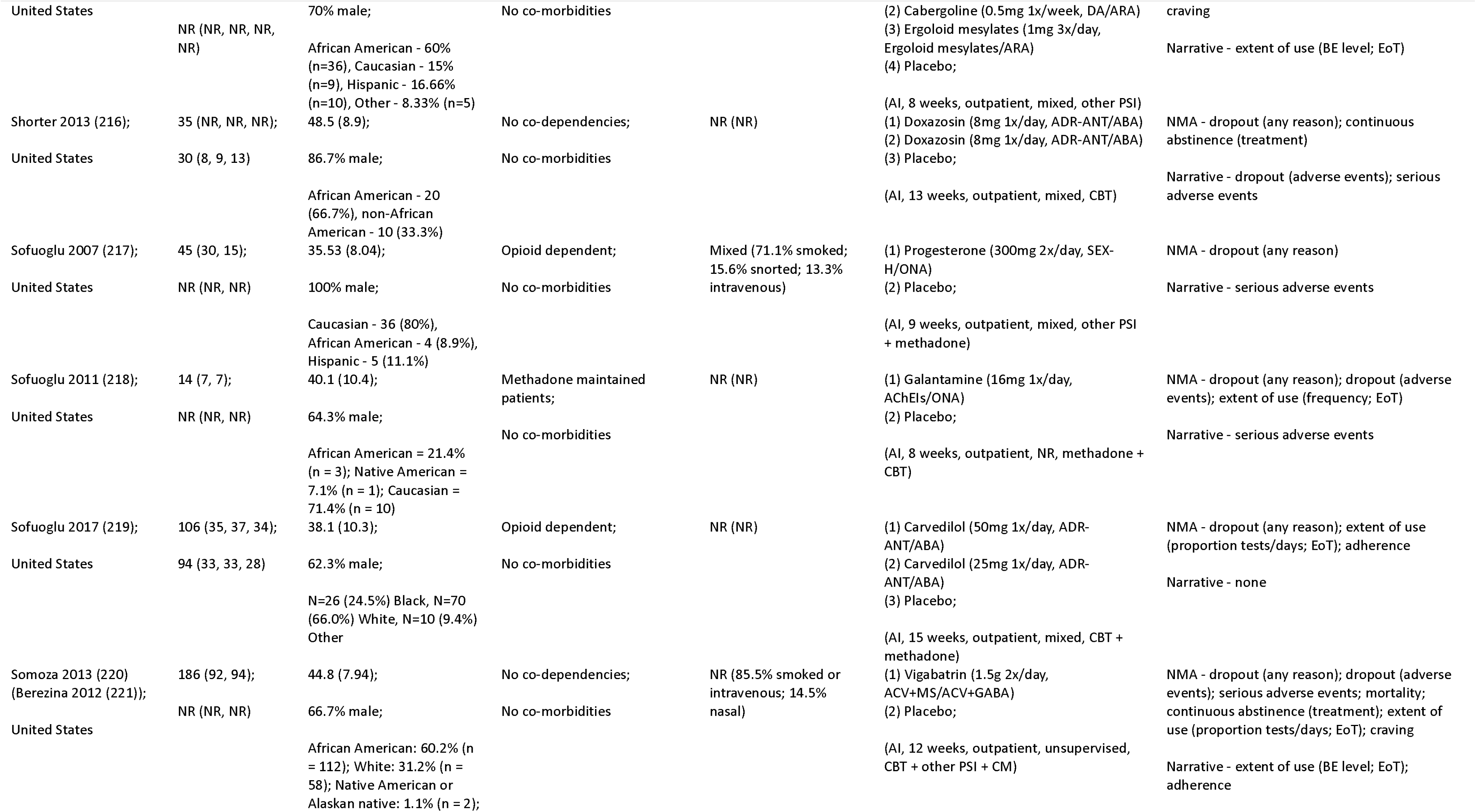
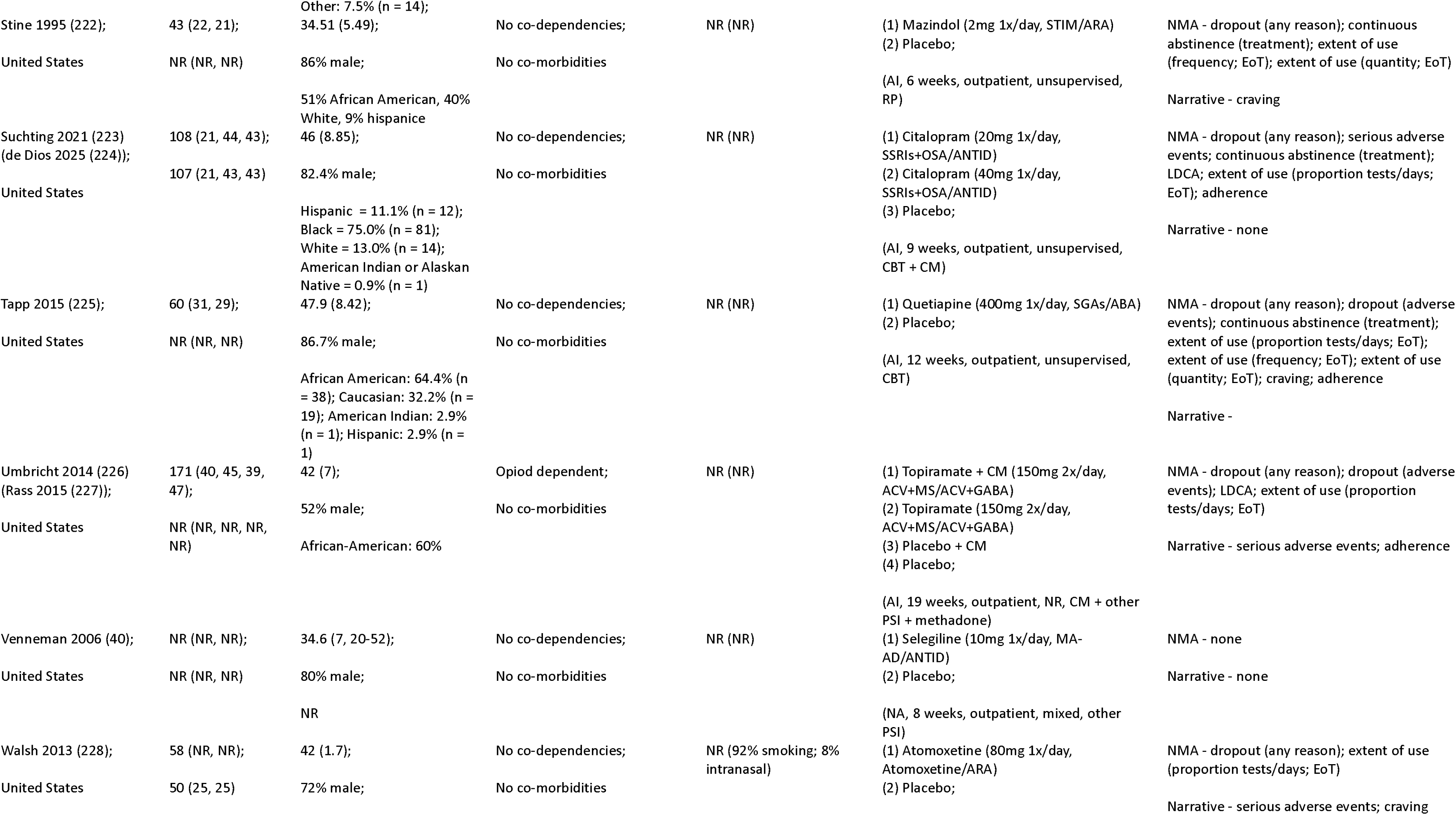
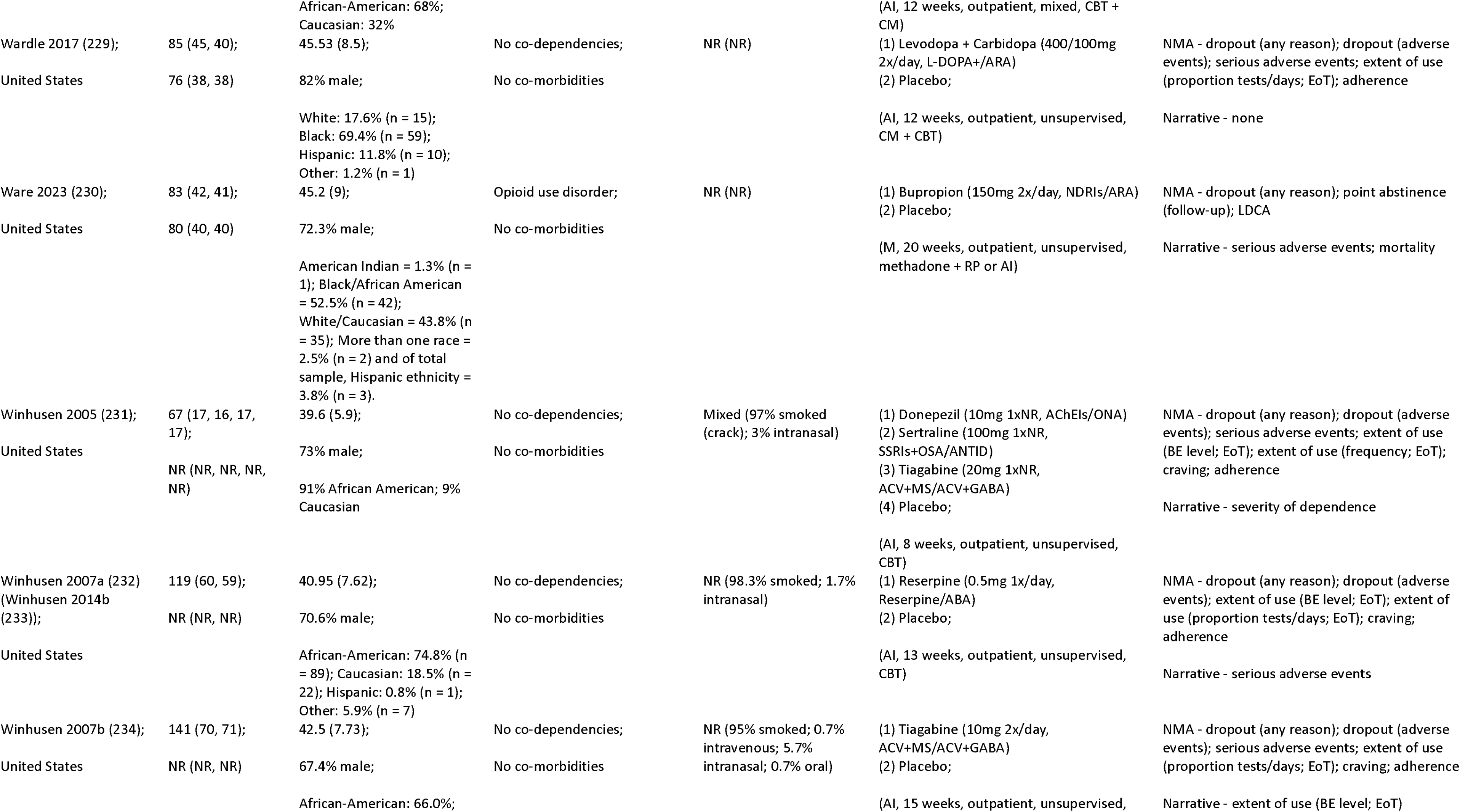
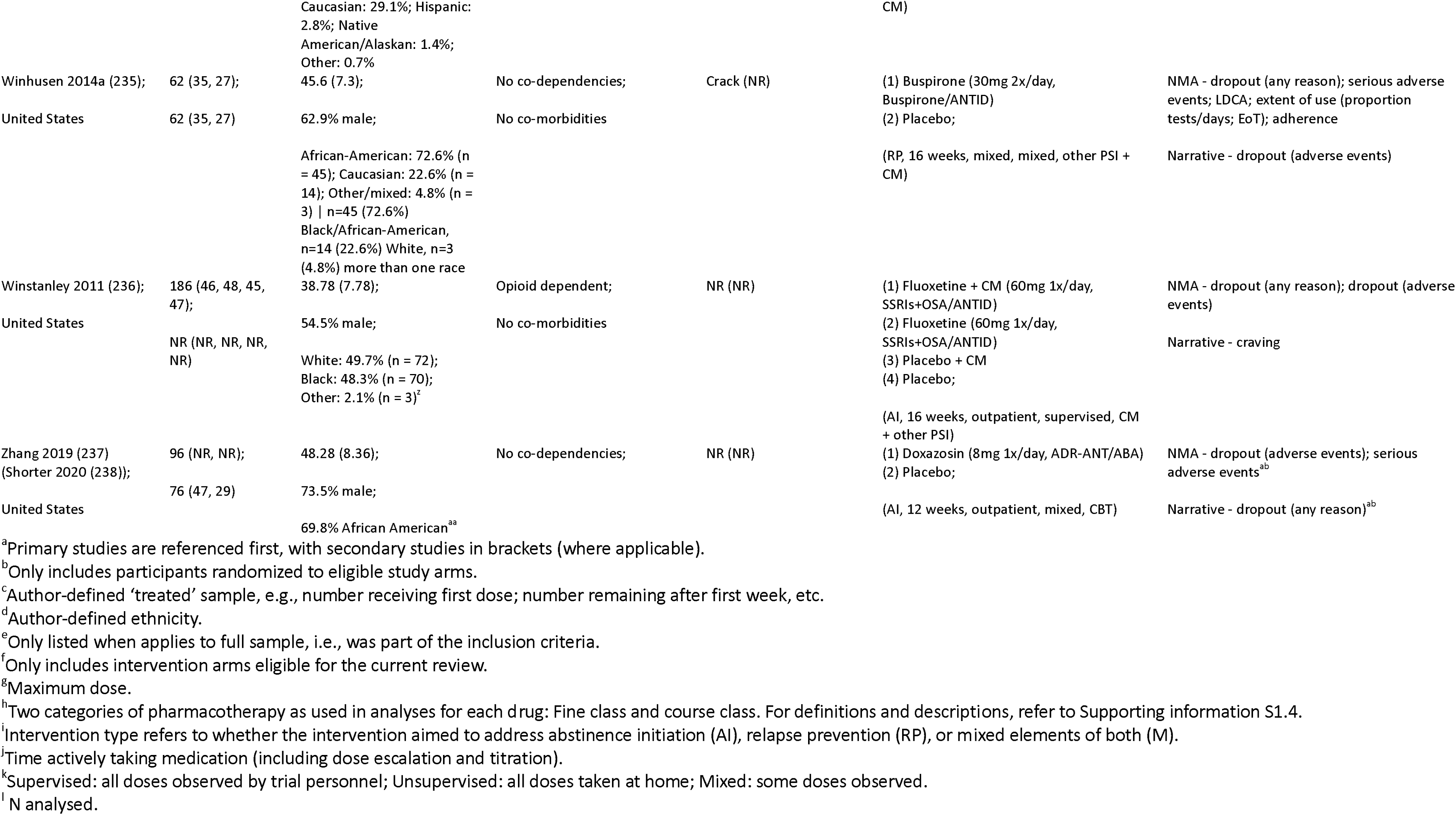
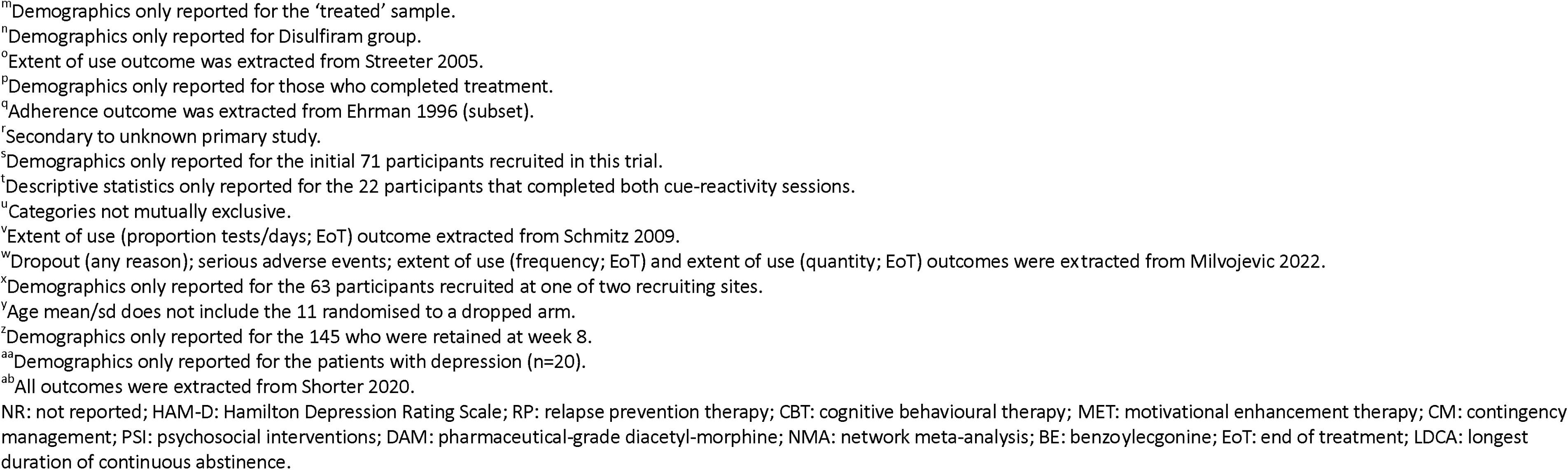
Characteristics of included studies.

In total, 89 unique medications were included across all studies. As in previous reviews of pharmacological treatments for SUD, we grouped medications according to type and mechanism of action for the purpose of analysis (239). Two versions of drug categories were created: (i) therapeutic strategy and (ii) mechanistic. The therapeutic strategy categorization pragmatically consolidated medications into a smaller set of broader intervention nodes, consisting of eight classes in total (Supporting information S1.5). In developing these categories, we sought to preserve major therapeutic approaches relevant to CUD (e.g., dopaminergic agonist/replacement strategies or dopamine-blocking/antagonist approaches) (240) and to reflect the focus of previous reviews (e.g., anticonvulsants, antidepressants) (21,241).

The mechanistic categorization captured 39 mechanistic groupings that clustered related pharmacological classes (e.g., NMDA receptor antagonists, opioid agonist therapies. Further details, including lists of medications within each therapeutic strategy and mechanistic category, are also provided in the supplementary materials (Supporting information S1.5). This dual approach aimed to preserve pharmacological similarity in how interventions were grouped, while pragmatically consolidating nodes to support network connectivity and facilitate NMA. Results from the mechanistic categorization are not presented in the main text as the large number of nodes with small study counts limits interpretability, although we have presented them in the supplement for completeness (Supporting information S1.6).

### Outcomes

Results for primary outcomes are presented in the text. Quantitative data were available for all outcomes. We conducted NMA for all outcomes except ‘extent of use at follow-up (urine benzoylecgonine [BE] levels)’ and ‘severity of dependence’ where insufficient data were available for analyses. Where outcomes were reported in a format that was not suitable for NMA or insufficient data were available, we present results narratively in Supporting information S2.

### Risk of bias

We assessed a total of 590 results included in the NMAs. We judged 48% to be at high risk of bias, 36% to have some concerns, and 16% to be at low risk of bias. The principal concerns were risk of bias due to failure to follow intention-to-treat principles in the analysis (covered in the domain on deviations from the intended interventions) and risk of bias due to missing outcome data. Detailed assessments for each outcome are presented in Supporting information S1.7.

### Results of the syntheses

We considered transitivity across the included studies to be plausible, so we conducted NMAs on outcomes for which the available comparisons formed a connected network. Results from random-effects models are presented for most outcomes. Unless otherwise stated, the results from common-effect sensitivity analyses and direct estimates from inconsistency random-effects models were consistent with the primary NMAs. For models with fewer than 5 degrees of freedom for estimating the amount of heterogeneity, results of common-effect models are presented without sensitivity analyses. R-hat values were all <1.01, suggesting good convergence. Network plots for all analyses are presented in Supporting information 3A-AF. We conducted subgroup analyses by intervention type (abstinence initiation vs relapse prevention) and presence of co-occurring SUD/comorbidity for continuous abstinence during treatment, point abstinence at end of treatment, and dropout for any reason. Several pre-specified subgroup analyses were not possible. The predominant cocaine type used by participants was seldom reported. Around 95% of studies employed co-occurring psychosocial interventions (e.g., counselling) and the vast majority of studies were conducted in unsupervised, outpatient settings, which prevented meaningful subgroup analysis.

Results are primarily expressed relative to placebo and relative effects for all outcomes are available in Supporting information 3A-AF. In line with the CINeMA approach of considering outcomes in the context of a MCID, we estimated the probability (reported as Pr) that the relative effect exceeded a MCID by calculating the proportion of posterior draws above or below that value from each NMA (depending on the direction of beneficial effect). The results within the manuscript represent the therapeutic strategy categorization. Results from the mechanistic categorization are presented in Supporting information S1.6).

#### Effectiveness

##### Continuous abstinence during treatment

NMA consisted of 46 studies of 9 classes, including direct evidence from a maximum of 11 studies per contrast (*antidepressants and other serotonergic agents*). Periods of continuous abstinence ranged from 1 to 16 weeks, most commonly 3 weeks (29/46 studies). A majority of studies (25/46) assessed a period of continuous abstinence up to and including end of treatment, with the remaining 21 studies assessing a period of continuous abstinence at any point during the treatment period. Results of random-effects NMA suggest that *antidepressants and other serotonergic agents* (OR 1.74, 95% CrI: 1.02, 3.13, Pr = 0.89) and *combination pharmacotherapy* (OR 4.38, 95% CrI: 2.04, 9.94, Pr = 0.99) may have beneficial effects relative to placebo (**Error! Reference source not found.**A). Other placebo comparisons were consistent with no effect. Relative effects of different active treatments suggest that *combination pharmacotherapy* may be more beneficial than all treatments except *anticonvulsants and GABAergic agents* and *agonist/replacement approaches*. All other contrasts between interventions were consistent with the possibility of no effect. All estimates were rated as very low certainty.

Results of subgroup analysis by presence of co-occurring SUD/comorbidity (Figure 3A) suggests that *combination pharmacotherapy* may be beneficial in both groups, and *antidepressants and other serotonergic agents* may be more beneficial in those with co-occurring SUD/comorbidity (OR 2.27, 95% CrI: 1.00, 5.45) than those without (OR 1.19, 95% CrI: 0.48, 2.89). Subgroup analysis by intervention type (Figure 3B) suggest that *combination pharmacotherapy* may have a beneficial effect relative to placebo in abstinence initiation studies (OR 5.16, 95% CrI: 2.39, 11.7) but there was substantial uncertainty in relapse prevention studies (2.70, 95% CrI: 0.56, 13.8). Additionally, whilst *agonist/replacement approaches* may have a beneficial effect in abstinence initiation studies relative to placebo (OR 2.19, 95% CrI: 1.23, 4.01), that effect, although imprecise, may be reversed in relapse prevention studies, resulting in reduced abstinence relative to placebo (OR 0.03, 95% CrI: 0.00, 0.32).

**Figure 1.** PRISMA flow diagram

**Figure 2.**
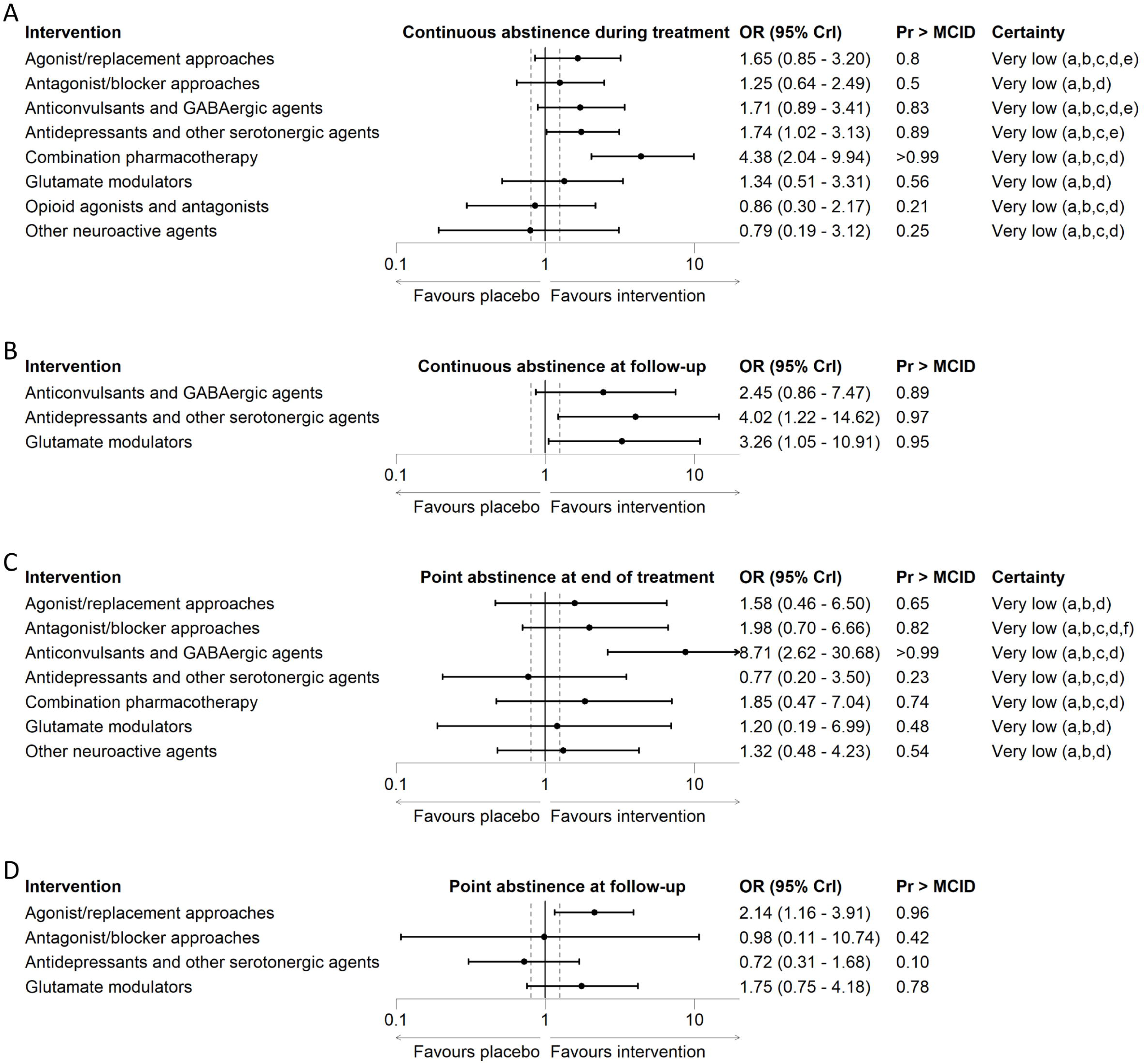
Forest plots of primary effectiveness outcomes. (A) continuous abstinence during treatment, (B) continuous abstinence at follow-up, (C) point abstinence at end of treatment, (D) point abstinence at follow-up.

**Figure 3.**
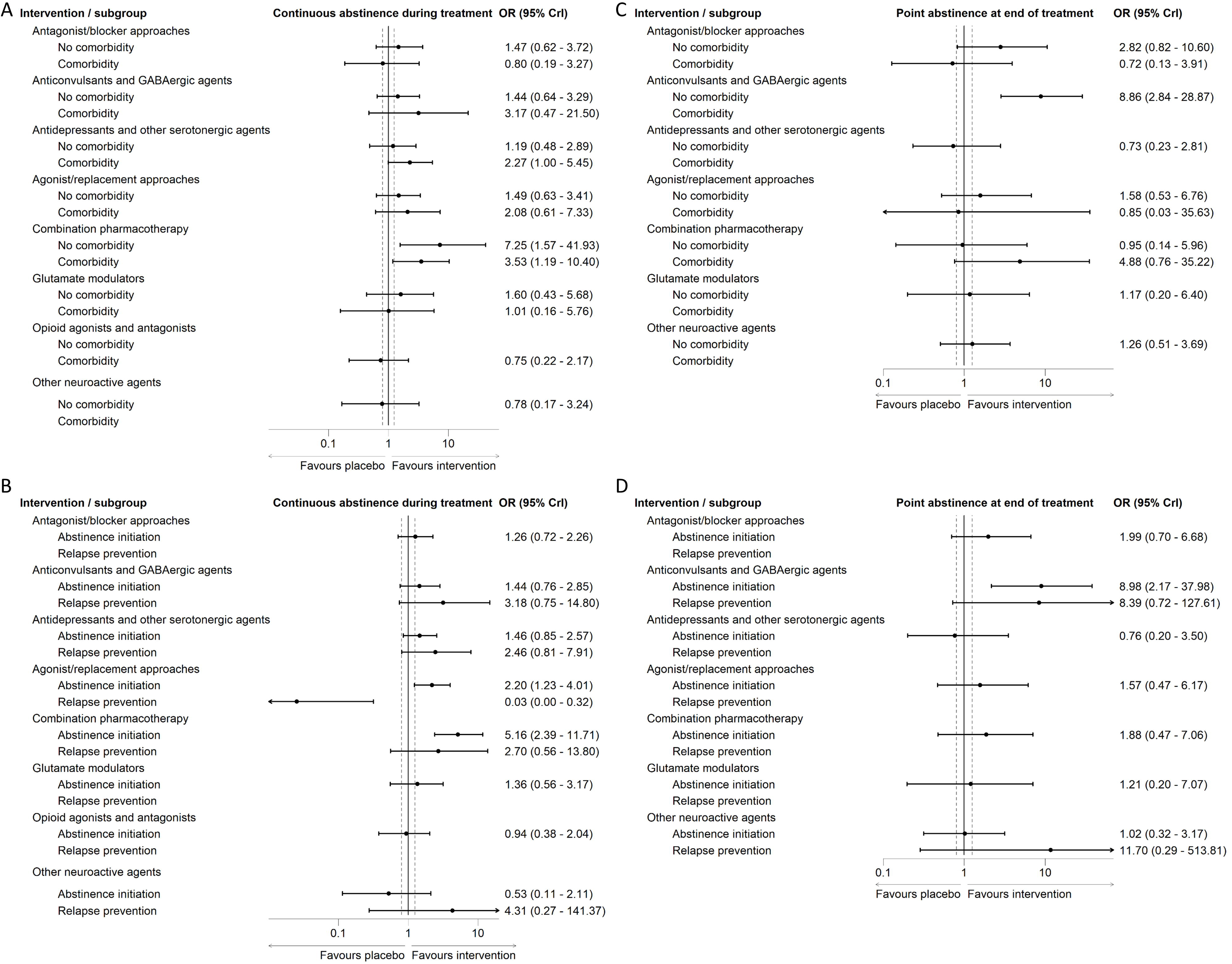
Forest plots of subgroup analyses for primary effectiveness outcomes. (A) continuous abstinence during treatment by presence of co-occurring SUD/comorbidity, (B) continuous abstinence during treatment by intervention type, (C) point abstinence at end of treatment by presence of co-occurring SUD/comorbidity, (D) point abstinence at end of treatment by intervention type.

##### Continuous abstinence at follow-up

NMA consisted of 5 studies of 4 classes, including direct evidence from a maximum of three studies per contrast (*glutamate modulators*). Studies assessed continuous periods of abstinence ranging from 2 to 24 weeks. Results of common-effect NMA suggest that *antidepressants and other serotonergic agents* (OR 4.02, 95% CrI: 1.22, 14.6) and *glutamate modulators* (OR 3.26, 95% CrI: 1.05, 10.9) may have beneficial effects relative to placebo (Figure 2B**Error! Reference source not found.**).

##### Point abstinence at end of treatment

NMA consisted of 22 studies of 8 classes, including direct evidence from a maximum of 6 studies per contrast (*agonist/replacement approaches*). All measures were confirmed with urinalysis and represented abstinence at the final assessment point during the treatment period. Results of random-effects NMA suggest that *anticonvulsants and GABAergic agents* may lead to meaningful improvements relative to placebo (OR 8.71, 95% CrI: 2.62, 30.7, Pr = 0.99) (**Error! Reference source not found.**2C**Error! Reference source not found.**). All other placebo and active treatment comparisons were consistent with the possibility of no effect relative to placebo. All estimates were rated as very low certainty.

Results of subgroup analyses were consistent with the primary models for studies in people without co-occurring SUD/comorbidity and abstinence initiation studies (Figure 3C and 3D). Due to very few relapse prevention studies and studies in people with co-occurring SUD/comorbidity, the effect estimates in these subgroups were very imprecise.

##### Point abstinence at end of follow-up

NMA consisted of 7 studies of 5 classes, including direct evidence from a maximum of three studies per contrast (*glutamate modulators*). Results of common-effect NMA suggest that *agonist/replacement approaches* may have beneficial effects relative to placebo (OR 2.14, 95% CrI: 1.16, 3.91) (Figure 2D**Error! Reference source not found.**). All other estimates showed evidence consistent with the possibility of no effect.

#### Acceptability

##### Dropout for any reason

NMA consisted of 135 studies of 9 classes, including direct evidence from a maximum of 34 studies per contrast (*agonist/replacement approaches*). All estimates were rated as very low certainty. All placebo comparisons from random-effects NMA showed evidence consistent with the possibility of no effect (Figure 4). Relative effects of different active treatments suggest that *antagonist/blocker approaches* may reduce dropout compared with *other neuroactive agents* while all other contrasts were consistent with the possibility of no effect.

**Figure 4.**
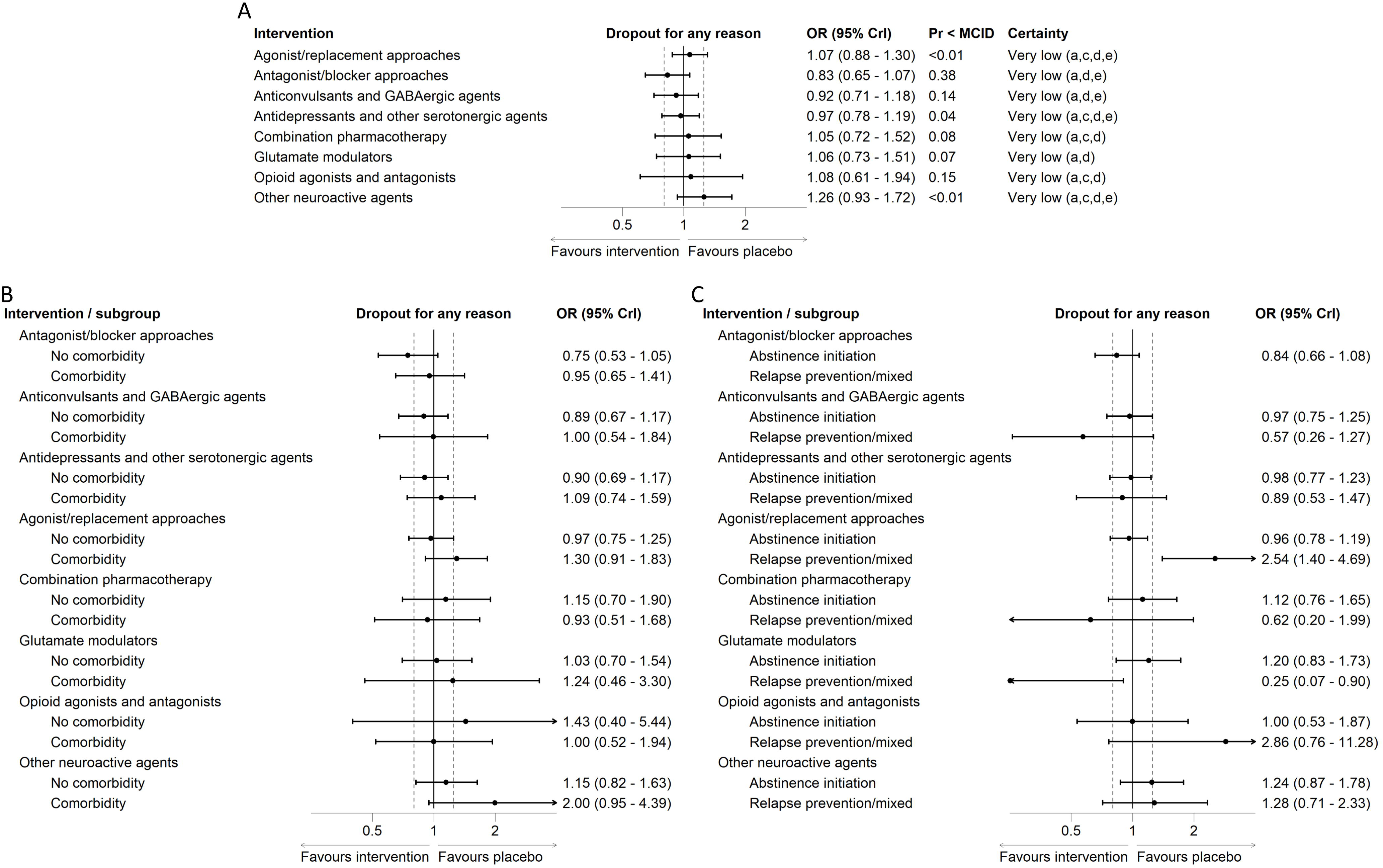
Forest plots of primary acceptability outcome, dropout for any reason, with subgroup analyses. (A) primary model, (B) subgroup by presence of co-occurring SUD/comorbidity, (C) subgroup by intervention type.

Results of subgroup analysis by presence of co-occurring SUD/comorbidity were consistent with the results of the main analysis (Figure 4B). Intervention type subgroup analysis results were largely consistent with the primary model with the exception of *agonist/replacement approaches*. In contrast to the primary analysis, *agonist/replacement approaches* appeared to result in greater dropout than placebo in relapse prevention/mixed studies (Figure 4C).

#### Safety

##### Serious adverse events

NMA consisted of 41 studies of 9 classes, including direct evidence from a maximum of 10 studies per contrast (*antidepressants and other serotonergic agents*). The number of reported serious adverse events across studies, and subsequently different nodes, was typically low, contributing to imprecise estimates with wide 95% CrI. No consistent evidence of effect was observed across any placebo or active treatment comparisons, either in the primary or subgroup analyses (Figure 5A-C).

**Figure 5.**
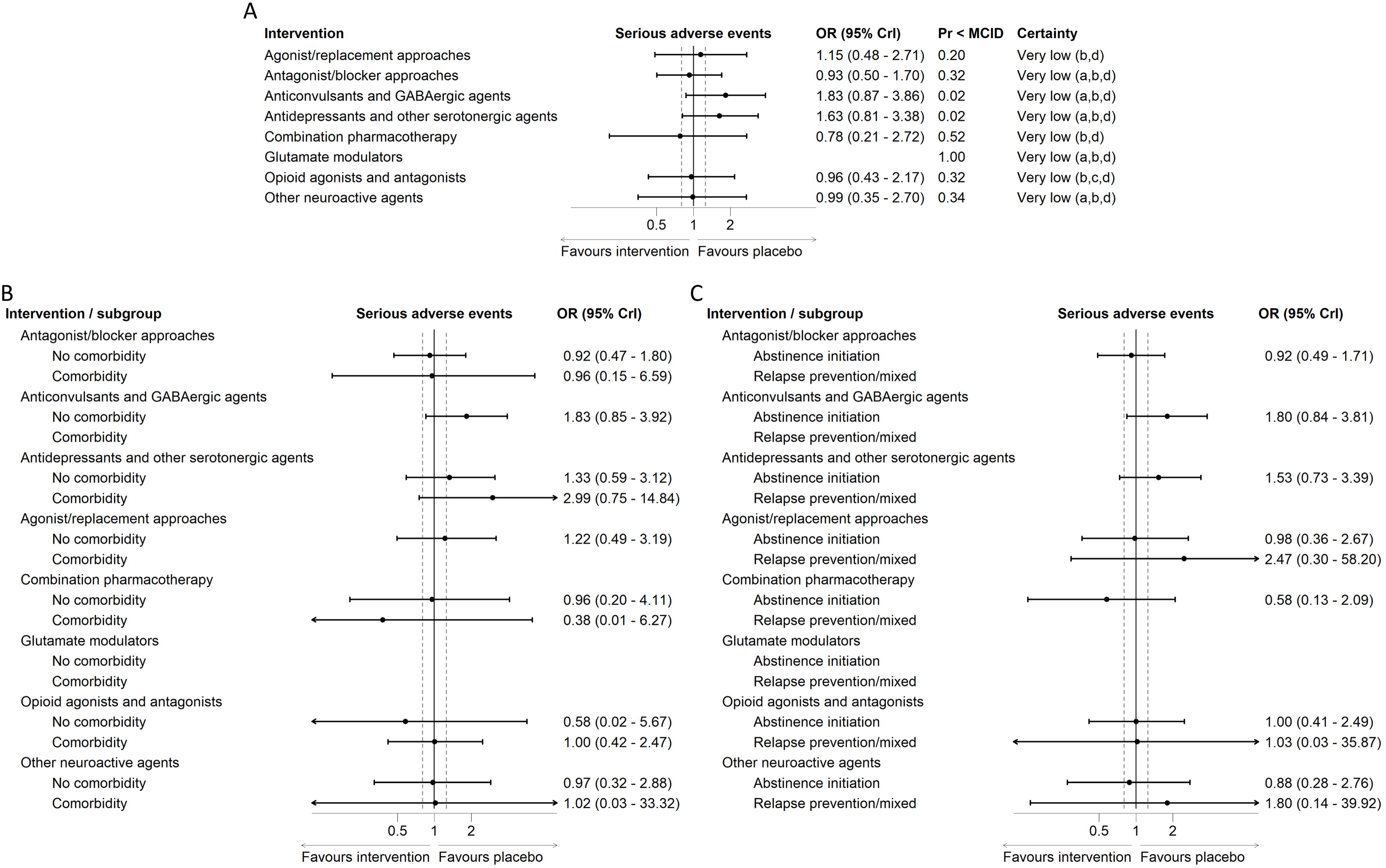
Forest plots of primary safety outcome, serious adverse events, with subgroup analyses. (A) primary model, (B) subgroup by presence of co-occurring SUD/comorbidity, (C) subgroup by intervention type.

The results for all secondary effectiveness, acceptability, and safety outcomes are summarized in Table 3.

**Table 3.**
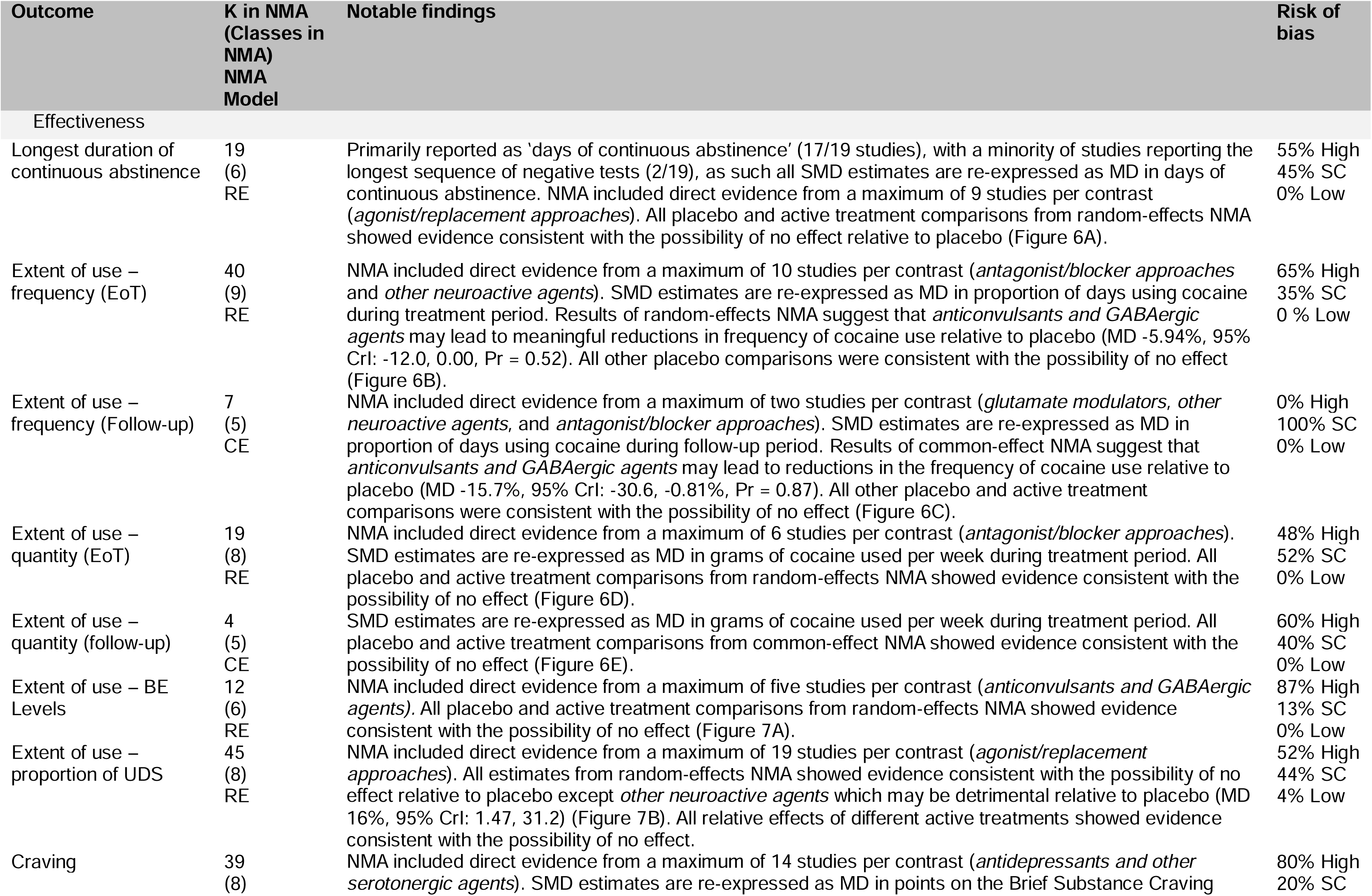
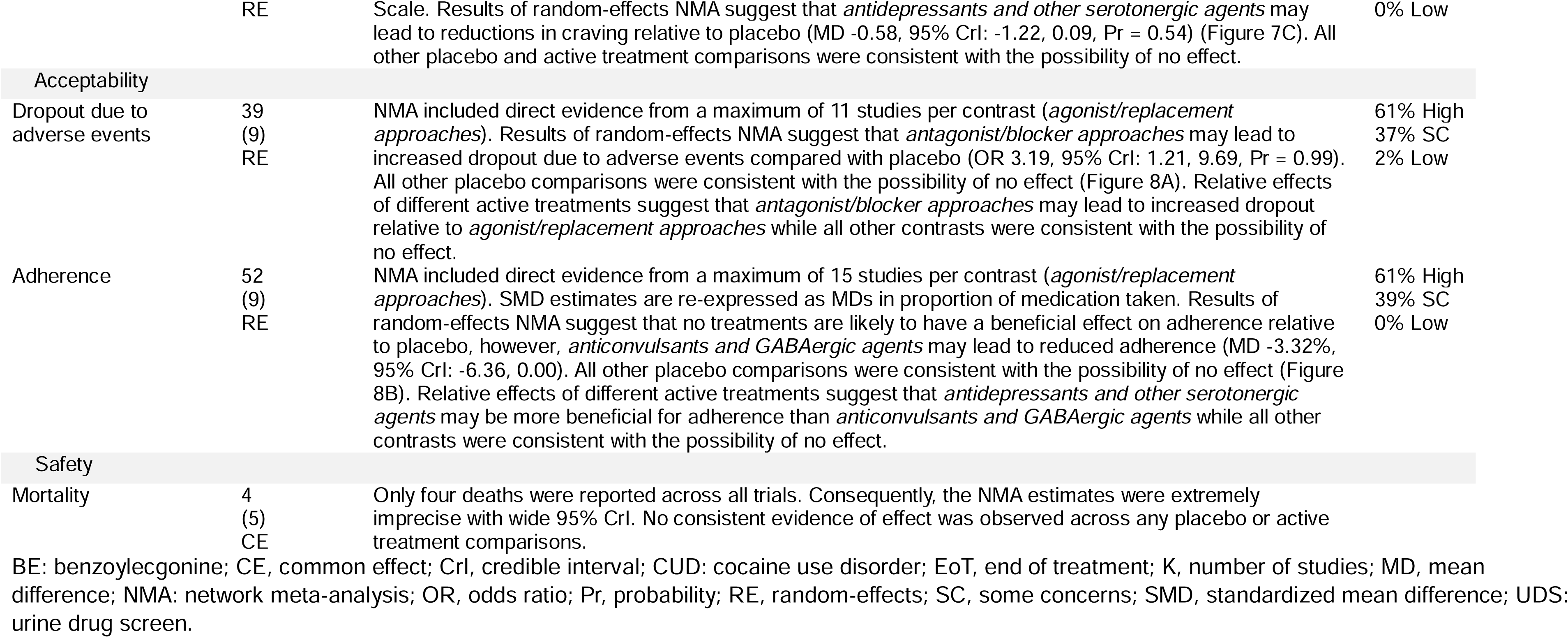
Secondary outcomes.

## DISCUSSION

### Summary of findings

Our systematic review and NMA sought to determine the comparative effectiveness, safety, and acceptability of different pharmacological interventions for the treatment of CUD and prevention of relapse. The 163 included RCTs with 148871 participants, evaluated 89 pharmacological agents, and our NMAs addressed 12 effectiveness outcomes, three acceptability outcomes, and two safety outcomes. Overall, there were limited and inconsistent signals from the data across outcomes with the certainty of evidence ranging from very low to low due to concerns regarding the risk of bias and imprecise and heterogenous results.

#### Effectiveness

The mixed signals from effectiveness outcomes, in tandem with risk of bias concerns and very low certainty, cast doubt on any potentially promising results for a given therapeutic strategy. For example, antidepressants and other serotonergic agents show a potentially promising effect for continuous abstinence during treatment and follow-up. However, there is limited evidence of effect for point abstinence or any extent of use outcomes (Figure 9). These findings seem incompatible as one would expect that if continuous abstinence was achieved for a specified period of follow-up, this beneficial effect would also be seen for point abstinence in the same time period (although this would not necessarily be expected in reverse). Similarly, anticonvulsants and GABAergic agents show promising results for point abstinence at end of treatment and follow-up, but we do not see such benefit across any extent of use outcomes. Even if none of these outcomes directly predict long-term changes in cocaine use behaviour, we would expect some congruence between related outcomes on this acute time scale. These divergent findings across effectiveness outcomes may be evidence of limited construct validity, or they may be indicative of spurious or chance findings. As such, any conclusions should be made with caution.

**Figure 6.**
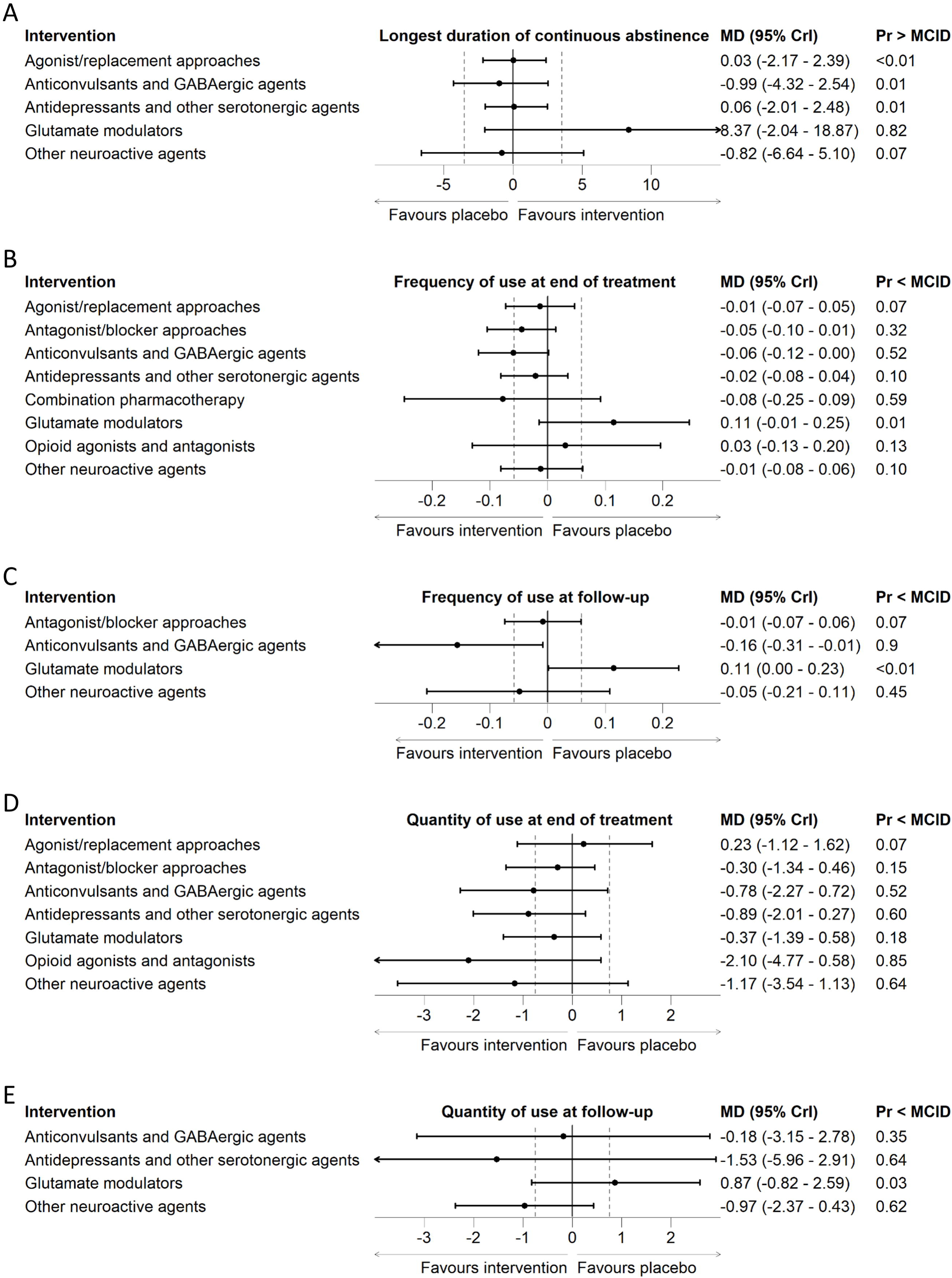
Forest plots of secondary effectiveness outcomes. (A) longest duration of continuous abstinence, (B) extent of use – frequency at end of treatment, (C) extent of use - frequency at end of follow-up, (D) extent of use – quantity at end of treatment, (E) extent of use - quantity at end of follow-up.

**Figure 7.**
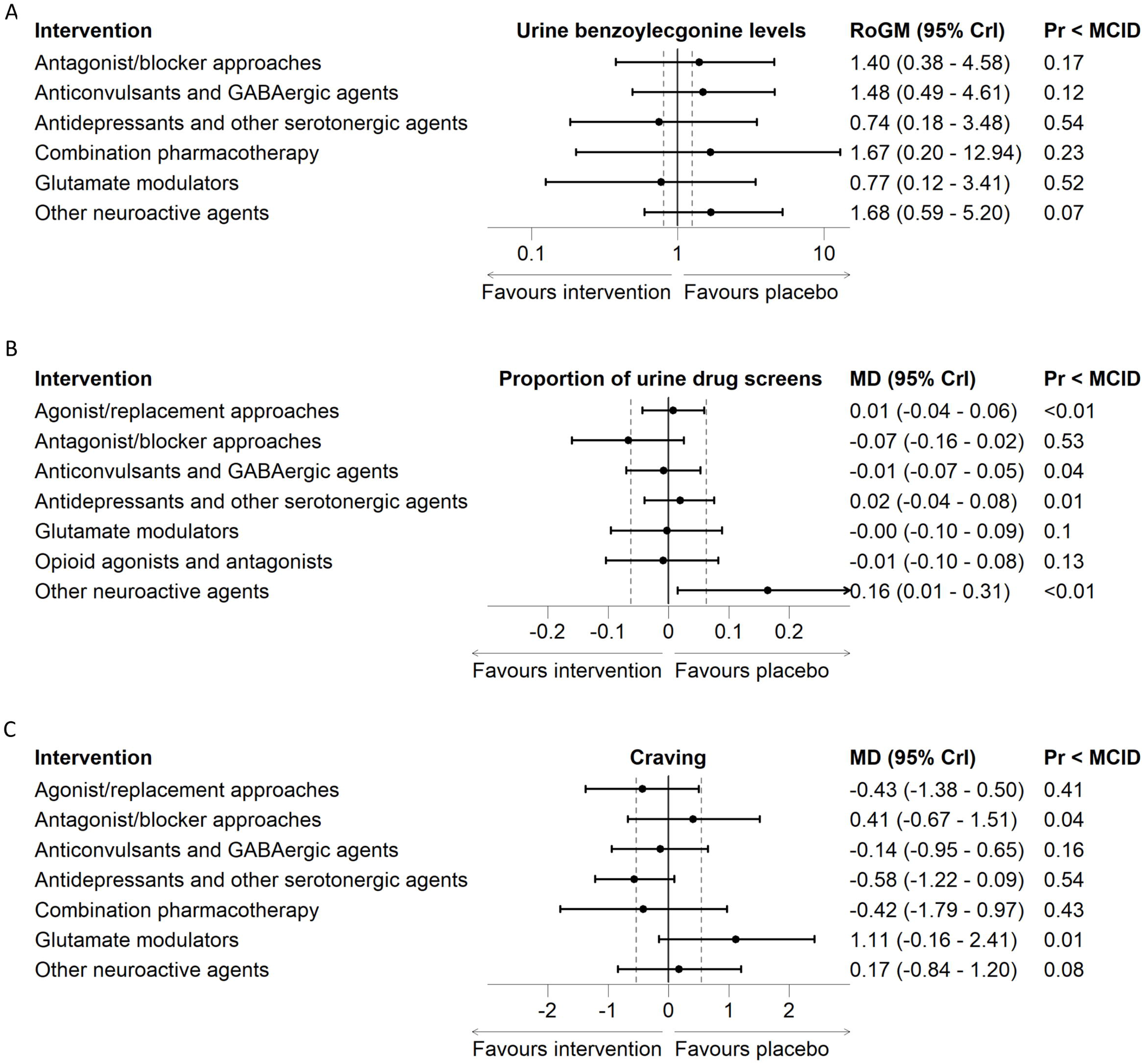
Forest plots of secondary effectiveness outcomes. (A) extent of use – urine benzoylecgonine levels, (B) extent of use - proportion of urine drug screens, (C) craving.

**Figure 8.**
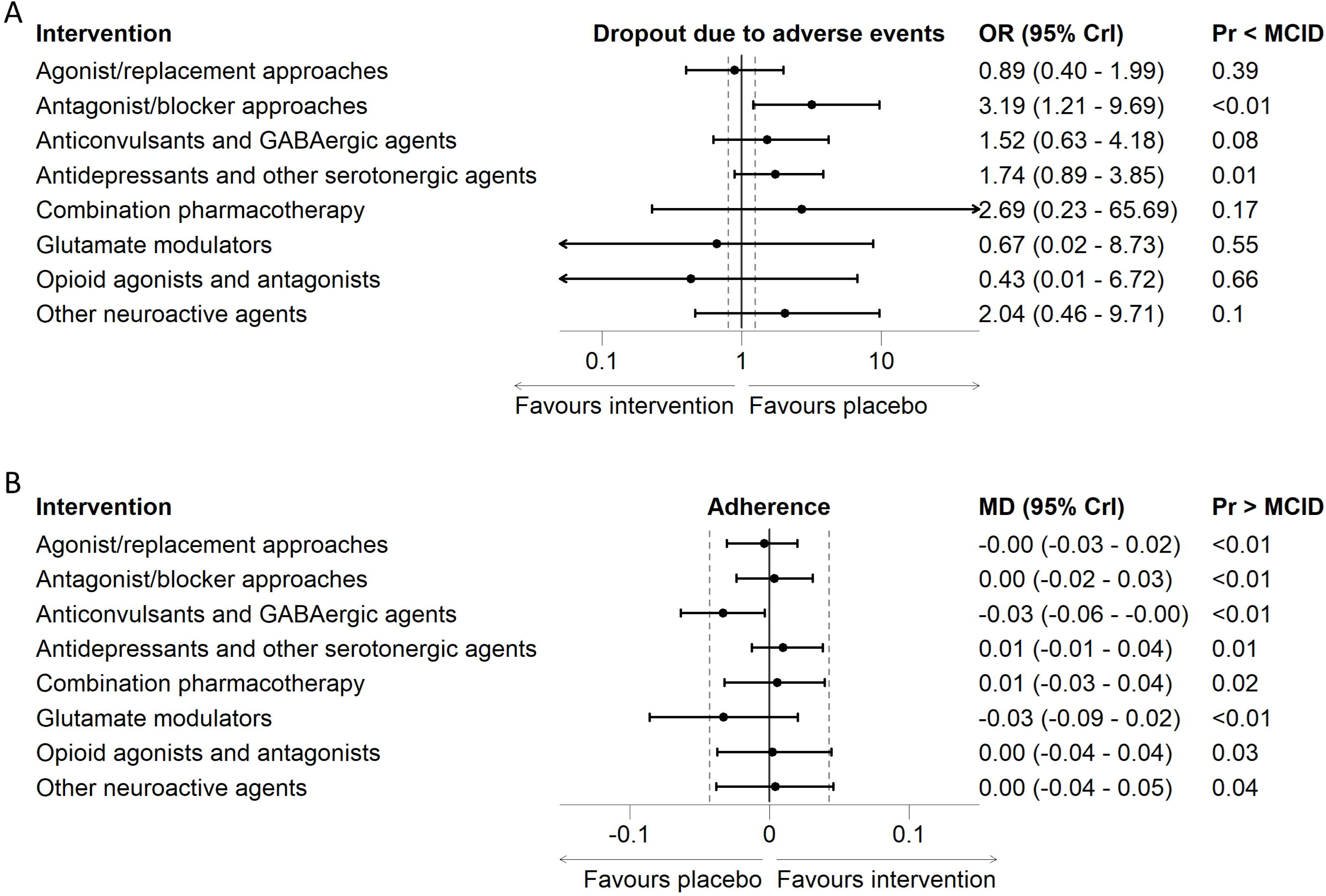
Forest plots of secondary acceptability outcomes. (A) dropout due to adverse events, (B) adherence.

**Figure 9.**
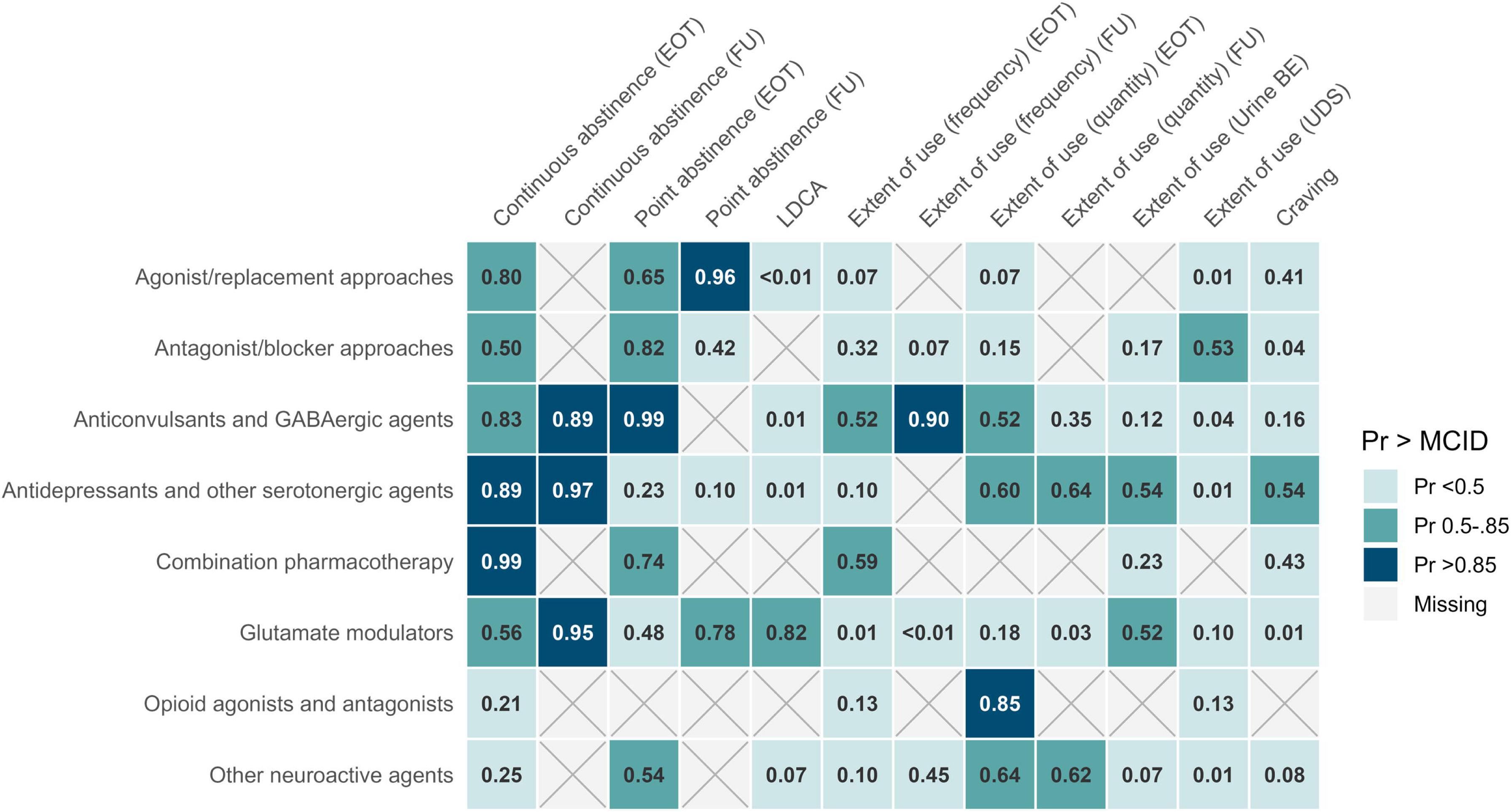
Heatmap of probabilities that point estimates for effectiveness outcomes exceed minimum clinically important differences.

#### Acceptability

Our primary analysis does not suggest that any one medication type is likely to influence dropout. All estimates were graded as very low certainty, due to risk of bias, imprecision, and indirectness concerns. We found no evidence that presence or absence of comorbidities affected the relative acceptability of pharmacotherapies. However, analysis by intervention type did suggest that agonist/replacement approaches used in relapse prevention studies may result in increased dropout relative to placebo but the reason for this is unclear.

#### Safety

Collectively, the results from this review do not suggest that any particular class of drugs is likely to be associated with increased incidence of serious adverse events or mortality. This is consistent with previous meta-analyses of different medication types, but the low number of events prevented precise estimations of intervention effects, such that any inferences should be made with caution.

### Strengths and limitations of the review

To our knowledge, this is the most comprehensive review and NMA of pharmacological interventions for CUD to date. Previous reviews have filtered by drug type (16) have restricted their synthesis to a single or a limited set of outcomes (11,242). By contrast, our review assessed a broad range of effectiveness, safety, and acceptability outcomes from double-blinded RCTs without limiting by medication type or by subpopulation of those living with CUD. We also applied NMA methods to enable simultaneous synthesis of different classes of drugs, the recommended approach when the evidence base consists of numerous diverse therapeutic approaches. Despite the rigorous methods employed in this review, many of its limitations stem from weaknesses with the broader evidence base. Firstly, we grouped studies by therapeutic strategy to facilitate NMA which may have missed some nuance afforded by differences in dosing or exact mechanism of action (22). However, as noted above, 163 identified studies assessed 89 unique medications, which indicates minimal replication across this research area. As such, any nuance that was missed by consolidating studies into broader therapeutic strategies may be outweighed by the potential for identifying potential future research directions. For completeness, results of a more granular grouping approach, informed by mechanism of action are presented in the supplementary material (Supporting Information S1.6). Secondly, in addition to heterogenous interventions, studies assessed broad and inconsistent effectiveness outcomes which made synthesis challenging. Such varying outcomes are perhaps understandable in the absence of established proxy outcomes for long-term behaviour change in people living with CUD. However, the lack of standardized outcomes may dilute the evidence base with outcomes that do not truly represent marked changes in cocaine use or lasting behaviour change. Thirdly, whilst we planned to conduct subgroup analyses by the presence of co-occurring SUD/comorbidity and predominant cocaine type, in practice so few trials adequately reported these factors, and thus we were unable to fully explore these potential differences.

### Implications for future research

This review highlights a lack of coherence across pharmacological trials for CUD. The absence of validated proxy measures for long-term behaviour change in people living with CUD has led to the use of heterogeneous and incongruent indicators of effectiveness. Equally, demographic and clinical characteristics are typically poorly and inconsistently reported. Given the high prevalence of co-occurring substance use and mental health disorders among individuals with CUD (243) and the potentially different treatment needs for people using different types of cocaine (i.e., powder or crack) (244), the under-reporting or exclusion of such factors reduces practicality and generalisability of findings. Establishing core outcomes and demographic reporting standards would enable greater comparability across trials and facilitate stronger conclusions.

Future work may also consider investigating different treatment paradigms. Most studies in this review were classified as abstinence initiation studies, whereby participants are actively using cocaine at the time of enrolment and the therapeutic goals are initiating and maintaining abstinence. However, for some SUDs, one approach is to employ an inpatient detoxification period before using pharmacological therapies to prevent relapse (rather than initiate abstinence) (245). Based on input from clinical experts, we had planned to compare findings from the two different treatment paradigms in a subgroup analysis, however, the paucity of studies using a relapse prevention design prohibited this. Future research should address this evidence gap in pharmacological treatments for longer-term prevention of relapse in people who have achieved initial abstinence from cocaine.

## CONCLUSIONS

Results from this systematic review and NMAs suggest that there is currently little compelling evidence for the adoption of pharmacological treatments for CUD. Whilst some treatment strategies such as *antidepressants and other serotonergic agents* and *anticonvulsants and GABAergic agents* showed promise for some effectiveness outcomes, these beneficial findings were not consistent across all effectiveness outcomes. There was limited evidence regarding safety of different pharmacotherapies and some were associated with poor acceptability. Additionally, all estimates were rated as “very low” or “low” certainty with common concerns around bias. Future work should seek to develop a set of core outcomes and demographics to improve the congruency of future research.

## Data Availability

All data produced in the present work are contained in the manuscript

## CRediT statement

Conceptualization: JSav, JPTH, DMC, FS; Data Curation: CP, TP, CB, MH, HG, JCP, RW, KW, FS; Formal analysis: MH, HG; Funding acquisition: JPTH, DMC, JSav; Investigation: CP, TP, CB, MH, HG, JCP, RW, KW, FS, SD; Methodology: CP, JSco, JPTH, JSav; Project administration: CP; Software: MH; Supervision: DMC, JPTH, SS, LAH; Validation: MH, TP, KW; Visualization: MH, HG, CP; Writing – Original draft: CP, TP, CB; Writing – reviewing and editing: CP, TP, CB, MH, HG, JCP, RW, KW, FS, SD, DMC, JSco, JPTH, JSav.

## Acknowledgements

We would like to thank our patient and public contributors from the Bristol Drugs Project for their invaluable insight during the development of the protocol for this review.

## Declarations of competing interest

None to declare

## Primary funding

This review was funded by National Institute for Health and Care Research (NIHR165378). The views expressed are those of the authors and not necessarily those of the NIHR or the Department of Health and Social Care.

## PROSPERO Registration

CRD42024596434

## Supporting Information

Supporting information S1.1 – Hierarchy for extracting outcomes with multiple data points

Supporting information S1.2 – Literature search strategy

Supporting information S1.3 – Additional analysis details, model parameters, and sensitivity analyses details

Supporting information S1.4 - Description of interventions

Supporting information S1.5 – Network meta-analysis results for mechanistic treatment classification

Supporting information S1.6 – Risk of bias assessments for included studies

Supporting information S1.7 – Data preparation methods

Supporting information S2 – Narrative results for all outcomes

Supporting information S3 – Network plots and all relative effects from network meta-analysis models of pharmacotherapies on each outcome

## Notes

### Competing Interest Statement

The authors have declared no competing interest.

## References

1. United Nations: Office on Drugs and Crime [Internet]. [cited 2025 Sept 19]. World Drug Report 2025 - Key findings. Available from: http://www.unodc.org/unodc/en/data-and-analysis/world-drug-report-2025-key-findings.html

2. Schwartz EKC, Wolkowicz NR, De Aquino JP, MacLean RR, Sofuoglu M. Cocaine Use Disorder (CUD): Current Clinical Perspectives. Subst Abuse Rehabil. 2022 Sept 3;13:25–46.

3. Hasin DS, O’Brien CP, Auriacombe M, Borges G, Bucholz K, Budney A, et al. DSM-5 Criteria for Substance Use Disorders: Recommendations and Rationale. Am J Psychiatry. 2013 Aug 1;170(8):834–51.

4. O’Brien MS, Anthony JC. Risk of Becoming Cocaine Dependent: Epidemiological Estimates for the United States, 2000–2001. Neuropsychopharmacol. 2005 May;30(5):1006–18.

5. Substance Abuse and Mental Health Services Administration. Key Substance Use and Mental Health Indicators in the United States: Results from the 2019 National Survey on Drug use and Health [Internet]. Rockville, MD: Center for Behavioral Health Statistics and Quality, Substance Abuse and Mental Health Administration; 2020. Report No.: PEP20-07-01–001. Available from: https://www.samhsa.gov/data/sites/default/files/reports/rpt29393/2019NSDUHFFRPDFWHTML/2019NSDUHFFR1PDFW090120.pdf

6. Stankowski RV, Kloner RA, Rezkalla SH. Cardiovascular consequences of cocaine use. Trends in Cardiovascular Medicine. 2015 Aug 1;25(6):517–26.

7. Jovanovski D, Erb S, Zakzanis KK. Neurocognitive deficits in cocaine users: a quantitative review of the evidence. J Clin Exp Neuropsychol. 2005 Feb;27(2):189–204.

8. Farrell M, Martin NK, Stockings E, Bórquez A, Cepeda JA, Degenhardt L, et al. Responding to global stimulant use: challenges and opportunities. The Lancet. 2019 Nov 2;394(10209):1652–67.

9. Daldegan-Bueno D, Fischer B. The association between cocaine product use and violence outcomes in Brazil: A comprehensive, systematized review. Aggression and Violent Behavior. 2024 Jan 1;74:101891.

10. Cocaine – the current situation in Europe (European Drug Report 2024) | www.euda.europa.eu [Internet]. [cited 2025 Sept 19]. Available from: https://www.euda.europa.eu/publications/european-drug-report/2024/cocaine_en#level-5

11. Bentzley BS, Han SS, Neuner S, Humphreys K, Kampman KM, Halpern CH. Comparison of treatments for cocaine use disorder among adults: a systematic review and meta-analysis. JAMA network open. 2021;4(5):e218049–e218049.

12. Rösner S, Hackl-Herrwerth A, Leucht S, Lehert P, Vecchi S, Soyka M. Acamprosate for alcohol dependence - Rösner, S - 2010 | Cochrane Library. [cited 2024 Sept 4]; Available from: https://www.cochranelibrary.com/cdsr/doi/10.1002/14651858.CD004332.pub2/full

13. Rösner S, Hackl-Herrwerth A, Leucht S, Vecchi S, Srisurapanont M, Soyka M. Opioid antagonists for alcohol dependence - Rösner, S - 2010 | Cochrane Library. [cited 2024 Sept 4]; Available from: https://www.cochranelibrary.com/cdsr/doi/10.1002/14651858.CD001867.pub3/full

14. Mattick RP, Breen C, Kimber J, Davoli M. Methadone maintenance therapy versus no opioid replacement therapy for opioid dependence - Mattick, RP - 2009 | Cochrane Library. [cited 2024 Sept 4]; Available from: https://www.cochranelibrary.com/cdsr/doi/10.1002/14651858.CD002209.pub2/full

15. Chan B, Kondo K, Freeman M, Ayers C, Montgomery J, Kansagara D. Pharmacotherapy for Cocaine Use Disorder-a Systematic Review and Meta-analysis. J Gen Intern Med. 2019 Dec;34(12):2858–73.

16. Traccis F, Minozzi S, Trogu E, Vacca R, Vecchi S, Pani PP, et al. Disulfiram for the treatment of cocaine dependence. Cochrane Database Syst Rev. 2024 Jan 5;1(1):CD007024.

17. Hutton B, Salanti G, Caldwell DM, Chaimani A, Schmid CH, Cameron C, et al. The PRISMA Extension Statement for Reporting of Systematic Reviews Incorporating Network Meta-analyses of Health Care Interventions: Checklist and Explanations. Ann Intern Med. 2015 June 2;162(11):777–84.

18. Castells X, Cunill R, Pérez-Mañá C, Vidal X, Capellà D. Psychostimulant drugs for cocaine dependence. Cochrane Database Syst Rev. 2016 Sept 27;9(9):CD007380.

19. Chan B, Freeman M, Ayers C, Korthuis PT, Paynter R, Kondo K, et al. A systematic review and meta-analysis of medications for stimulant use disorders in patients with co-occurring opioid use disorders. Drug Alcohol Depend. 2020 Nov 1;216:108193.

20. Lee-Cheong S, Ludgate SA, Epp TCM, Schütz CG. The effectiveness of oxytocin in the treatment of stimulant use disorders: a systematic review. Behav Pharmacol. 2023 Oct 1;34(7):381–92.

21. Minozzi S, Cinquini M, Amato L, Davoli M, Farrell MF, Pani PP, et al. Anticonvulsants for cocaine dependence. Cochrane Database Syst Rev. 2015 Apr 17;2015(4):CD006754.

22. Tardelli VS, Bisaga A, Arcadepani FB, Gerra G, Levin FR, Fidalgo TM. Prescription psychostimulants for the treatment of stimulant use disorder: a systematic review and meta-analysis. Psychopharmacology (Berl). 2020 Aug;237(8):2233–55.

23. Dias SW, Sutton A, Caldwell D, Lu G, Ades A. NICE DSU Technical Support Document 4: Inconsistency in the networks of evidence based on randomised controlled trials [Internet]. 2014. Available from: https://www.sheffield.ac.uk/media/34181/download?attachment

24. Sterne JA, Savović J, Page MJ, Elbers RG, Blencowe NS, Boutron I, et al. RoB 2: a revised tool for assessing risk of bias in randomised trials. bmj [Internet]. 2019 [cited 2024 Sept 4];366. Available from: https://www.bmj.com/content/366/bmj.l4898.short?casa_token=v15LhimsNoEAAAAA:9pT_5xjK_ROPkXPLkZ4xFpixs7akmfdqyK-m-grywU_6AdIzXablcBu-kXlcapuvoJFVYHuo_Ns

25. Salanti G. Indirect and mixed-treatment comparison, network, or multiple-treatments meta-analysis: many names, many benefits, many concerns for the next generation evidence synthesis tool. Research Synthesis Methods. 2012;3(2):80–97.

26. Phillippo DM, Perren SJ. multinma: Bayesian Network Meta-Analysis of Individual and Aggregate Data.

27. R Core Team. R: A Language and Environment for Statistical Computing [Internet]. Vienna, Austria; 2020. Available from: https://www.R-project.org/

28. Nikolakopoulou A, Higgins JP, Papakonstantinou T, Chaimani A, Del Giovane C, Egger M, et al. CINeMA: an approach for assessing confidence in the results of a network meta-analysis. PLoS medicine. 2020;17(4):e1003082.

29. Covi L, Hess JM, Kreiter NA, Haertzen CA. Effects of combined fluoxetine and counseling in the outpatient treatment of cocaine abusers. Am J Drug Alcohol Abuse. 1995 Aug;21(3):327–44.

30. Epstein D.H., Schmittner J., Umbricht A., Schroeder J.R., Moolchan E.T., Preston K.L. Promoting abstinence from cocaine and heroin with a methadone dose increase and a novel contingency. Drug Alcohol Depend. 2009;101(1–2):92–100.

31. Grabowski J., Rhoades H., Silverman P., Schmitz J.M., Stotts A., Creson D., et al. Risperidone for the treatment of cocaine dependence: Randomized, double-blind trial. J Clin Psychopharmacol. 2000;20(3):305–10.

32. Harris D.S., Batki S.L., Berger S.P. Fluoxetine attenuates adrenocortical but not subjective responses to cocaine cues. Am J Drug Alcohol Abuse. 2004;30(4):765–82.

33. Moeller FG, Schmitz JM, Steinberg JL, Green CM, Reist C, Lai LY, et al. Citalopram combined with behavioral therapy reduces cocaine use: a double-blind, placebo-controlled trial. Am J Drug Alcohol Abuse. 2007;33(3):367–78.

34. Montoya ID, Gorelick DA, Preston KL, Schroeder JR, Umbricht A, Cheskin LJ, et al. Randomized trial of buprenorphine for treatment of concurrent opiate and cocaine dependence. Clin Pharmacol Ther. 2004 Jan;75(1):34–48.

35. Oliveto A, Poling J, Sevarino KA, Gonsai KR, McCance-Katz EF, Stine SM, et al. Efficacy of dose and contingency management procedures in LAAM-maintained cocaine-dependent patients. Drug Alcohol Depend. 2005 Aug;79(2):157–65.

36. Schmitz J.M., Lindsay J.A., Stotts A.L., Green C.E., Moeller F.G. Contingency management and levodopa-carbidopa for cocaine treatment: A comparison of three behavioral targets. Exp Clin Psychopharmacol. 2010;18(3):238–44.

37. Delucchi KL, Batki SL, Moon J, Jacob P 3rd, Jones RT. Urine toxicology samples in cocaine treatment trials: how many need to be tested? J Addict Dis. 2002;21(2):17–26.

38. Renshaw P.F., Daniels S., Lundahl L.H., Rogers V., Lukas S.E. Short-term treatment with citicoline (CDP-choline) attenuates some measures of craving in cocaine-dependent subjects: A preliminary report. Psychopharmacology. 1999;142(2):132–8.

39. Schmitz JM, Rhoades HM, Elk R, Creson D, Hussein I, Grabowski J. Medication take-home doses and contingency management. Exp Clin Psychopharmacol. 1998 May;6(2):162–8.

40. Venneman S, Leuchter A, Bartzokis G, Beckson M, Simon SL, Schaefer M, et al. Variation in neurophysiological function and evidence of quantitative electroencephalogram discordance: predicting cocaine-dependent treatment attrition. J Neuropsychiatry Clin Neurosci. 2006;18(2):208–16.

41. Afshar M., Knapp C.M., Sarid-Segal O., Devine E., Colaneri L.S., Tozier L., et al. The efficacy of mirtazapine in the treatment of cocaine dependence with comorbid depression. Am J Drug Alcohol Abuse. 2012;38(2):181–6.

42. Alterman A.I., Droba M., Antelo R.E., Cornish J.W., Sweeney K.K., Parikh G.A., et al. Amantadine may facilitate detoxification of cocaine addicts. DRUG ALCOHOL DEPEND. 1992;31(1):19–29.

43. Anderson A.L., Reid M.S., Li S.-H., Holmes T., Shemanski L., Slee A., et al. Modafinil for the treatment of cocaine dependence. Drug Alcohol Depend. 2009;104(1–2):133–9.

44. Baldacara L., Diniz T.A., Parreira B.L., Milhomem J.J., de Almeida L.J.C., Fernandes C.C. Could disulfiram be a new treatment for crack cocaine dependence? A pilot study. Rev Bras Psiquiatr. 2013;35(1):97–8.

45. Baldacara L., Cogo-Moreira H., Parreira B.L., Diniz T.A., Milhomem J.J., Fernandes C.C., et al. Efficacy of topiramate in the treatment of crack cocaine dependence: A double-blind, randomized, placebo-controlled trial. J Clin Psychiatry. 2016;77(3):398–406.

46. Batki S.L., Washburn A.M., Delucchi K., Jones R.T. A controlled trial of fluoxetine in crack cocaine dependence. DRUG ALCOHOL DEPEND. 1996;41(2):137–42.

47. Becker J.E., Price J.L., Leonard D., Suris A., Kandil E., Shaw M., et al. The Efficacy of Lidocaine in Disrupting Cocaine Cue-Induced Memory Reconsolidation. Drug Alcohol Depend. 2020;212((Becker, Suris, Shaw, Brown) Department of Psychiatry, UT Southwestern Medical Center, Dallas, TX, United States):108062.

48. Berger S.P., Winhusen T.M., Somoza E.C., Harrer J.M., Mezinskis J.P., Leiderman D.B., et al. A medication screening trial evaluation of reserpine, gabapentin and lamotrigine pharmacotherapy of cocaine dependence. Addiction. 2005;100(SUPPL. 1):58–67.

49. Bisaga A., Aharonovich E., Garawi F., Levin F.R., Rubin E., Raby W.N., et al. A randomized placebo-controlled trial of gabapentin for cocaine dependence. Drug Alcohol Depend. 2006;81(3):267–74.

50. Bisaga A, Aharonovich E, Garawi F, Levin FR, Rubin E, Raby WN, et al. Utility of lead-in period in cocaine dependence pharmacotherapy trials. Drug and Alcohol Dependence. 2005;77(1):7 EP–11.

51. Bisaga A., Aharonovich E., Cheng W.Y., Levin F.R., Mariani J.J., Raby W.N., et al. A placebo-controlled trial of memantine for cocaine dependence with high-value voucher incentives during a pre-randomization lead-in period. Drug Alcohol Depend. 2010;111(1–2):97–104.

52. Blevins D, Seneviratne C, Wang XQ, Johnson BA, Ait-Daoud N. A randomized, double-blind, placebo-controlled trial of ondansetron for the treatment of cocaine use disorder with post hoc pharmacogenetic analysis. Drug Alcohol Depend. 2019;228:109074.

53. Brady KT, Sonne SC, Malcolm RJ, Randall CL, Dansky BS, Simpson K, et al. Carbamazepine in the treatment of cocaine dependence: subtyping by affective disorder. Exp Clin Psychopharmacol. 2002 Aug;10(3):276–85.

54. Brodie J.D., Case B.G., Figueroa E., Dewey S.L., Robinson J.A., Wanderling J.A., et al. Randomized, double-blind, placebo-controlled trial of vigabatrin for the treatment of cocaine dependence in Mexican parolees. Am J Psychiatry. 2009;166(11):1269–77.

55. Campbell J, Nickel EJ, Penick EC, Wallace D, Gabrielli WF, Rowe C, et al. Comparison of desipramine or carbamazepine to placebo for crack cocaine-dependent patients. The American Journal on Addictions. 2003;12(2):122–36.

56. Campbell J.L., Thomas H.M., Gabrielli W., Liskow B.I., Powell B.J. Impact of desipramine or carbamazepine on patient retention in outpatient cocaine treatment: Preliminary findings. J ADDICT DIS. 1994;13(4):191–9.

57. Carpenter KM, Choi CJ, Basaraba C, Pavlicova M, Brooks DJ, Brezing CA, et al. Mixed amphetamine salts-extended release (MAS-ER) as a behavioral treatment augmentation strategy for cocaine use disorder: A randomized clinical trial. Exp Clin Psychopharmacol. 2024 Feb;32(1):112–27.

58. Carroll KM, Rounsaville BJ, Gordon LT, Nich C, Jatlow P, Bisighini RM, et al. Psychotherapy and pharmacotherapy for ambulatory cocaine abusers. Arch Gen Psychiatry. 1994 Mar;51(3):177–87.

59. Carroll KM, Rounsaville BJ, Nich C, Gordon LT, Wirtz PW, Gawin F. One-year follow-up of psychotherapy and pharmacotherapy for cocaine dependence. Delayed emergence of psychotherapy effects. Arch Gen Psychiatry. 1994 Dec;51(12):989–97.

60. Carroll KM, Nich C, Rounsaville BJ. Differential symptom reduction in depressed cocaine abusers treated with psychotherapy and pharmacotherapy. J Nerv Ment Dis. 1995 Apr;183(4):251–9.

61. Carroll K.M., Fenton L.R., Ball S.A., Nich C., Frankforter T.L., Shi J., et al. Efficacy of Disulfiram and Cognitive Behavior Therapy in Cocaine-Dependent Outpatients: A Randomized Placebo-Controlled Trial. Arch Gen Psychiatry. 2004;61(3):264–72.

62. Carroll K.M., Nich C., Shi J.M., Eagan D., Ball S.A. Efficacy of disulfiram and Twelve Step Facilitation in cocaine-dependent individuals maintained on methadone: A randomized placebo-controlled trial. Drug Alcohol Depend. 2012;126(1–2):224–31.

63. Carroll K.M., Nich C., Petry N.M., Eagan D.A., Shi J.M., Ball S.A. A randomized factorial trial of disulfiram and contingency management to enhance cognitive behavioral therapy for cocaine dependence. Drug Alcohol Depend. 2016;160((Carroll, Nich, Eagan, Shi, Ball) Yale University School of Medicine, Department of Psychiatry, United States):135–42.

64. DeVito EE, Dong G, Kober H, Xu J, Carroll KM, Potenza MN. Functional neural changes following behavioral therapies and disulfiram for cocaine dependence. Psychology of Addictive Behaviors. 2017;31(5):534–47.

65. Carroll KM, Nich C, DeVito EE, Shi JM, Sofuoglu M. Galantamine and Computerized Cognitive Behavioral Therapy for Cocaine Dependence: A Randomized Clinical Trial. J Clin Psychiatry [Internet]. 2018 Jan;79(1). Available from: https://libkey.io/libraries/1108/openurl?genre=article&aulast=Carroll&issn=0160-6689&title=Journal+of+Clinical+Psychiatry&atitle=Galantamine+and+Computerized+Cognitive+Behavioral+Therapy+for+Cocaine+Dependence%3A+A+Randomized+Clinical+Trial.&volume=79&issue=1&spage=17m11669&epage=&date=2018&doi=10.4088%2FJCP.17m11669&pmid=29286595&sid=OVID:medline

66. McCurdy LY, DeVito EE, Loya JM, Nich C, Zhai ZW, Kiluk BD, et al. Structural brain changes associated with cocaine use and digital cognitive behavioral therapy in cocaine use disorder treatment. Drug and Alcohol Dependence Reports. 2024;11:100246.

67. Ciraulo D.A., Sarid-Segal O., Knapp C.M., Ciraulo A.M., Locastro J., Bloch D.A., et al. Efficacy screening trials of paroxetine, pentoxifylline, riluzole, pramipexole and venlafaxine in cocaine dependence. Addiction. 2005;100(SUPPL. 1):12–22.

68. Streeter CC, Hennen J, Ke Y, Jensen JE, Sarid-Segal O, Nassar LE, et al. Prefrontal GABA levels in cocaine-dependent subjects increase with pramipexole and venlafaxine treatment. Psychopharmacology (Berl). 2005 Nov;182(4):516–26.

69. Ciraulo D.A., Knapp C., Rotrosen J., Sarid-Segal O., Ciraulo A.M., LoCastro J., et al. Nefazodone treatment of cocaine dependence with comorbid depressive symptoms. Addiction. 2005;100(SUPPL. 1):23–31.

70. Cornish J.W., Maany I., Fudala P.J., Neal S., Poole S.A., Volpicelli P., et al. Carbamazepine treatment for cocaine dependence. DRUG ALCOHOL DEPEND. 1995;38(3):221–7.

71. Cornish J.W., Maany I., Fudala P.J., Ehrman R.N., Robbins S.J., O’Brien C.P. A randomized, double-blind, placebo-controlled study of ritanserin pharmacotherapy for cocaine dependence. Drug Alcohol Depend. 2001;61(2):183–9.

72. Ehrman RN, Robbins SJ, Cornish JW, Childress AR, O’Brien CP. Failure of ritanserin to block cocaine cue reactivity in humans. Drug Alcohol Depend. 1996;42(3):167–74.

73. Dackis C.A., Kampman K.M., Lynch K.G., Pettinati H.M., O’Brien C.P. A double-blind, placebo-controlled trial of modafinil for cocaine dependence. Neuropsychopharmacology. 2005;30(1):205–11.

74. Dackis C.A., Kampman K.M., Lynch K.G., Plebani J.G., Pettinati H.M., Sparkman T., et al. A double-blind, placebo-controlled trial of modafinil for cocaine dependence. J Subst Abuse Treat. 2012;43(3):303–12.

75. Dakwar E, Nunes EV, Hart CL, Foltin RW, Mathew SJ, Carpenter KM, et al. A Single Ketamine Infusion Combined With Mindfulness-Based Behavioral Modification to Treat Cocaine Dependence: A Randomized Clinical Trial. Am J Psychiatry. 11 01;176(11):923–30.

76. DeVito EE, Carroll KM, Babuscio T, Nich C, Sofuoglu M. Randomized placebo-controlled trial of galantamine in individuals with cocaine use disorder. Journal of substance abuse treatment. 2019;107:29.

77. DeVito EE, Poling J, Babuscio T, Nich C, Carroll KM, Sofuoglu M. Modafinil Does Not Reduce Cocaine Use in Methadone-Maintained Individuals. Drug Alcohol Depend Rep [Internet]. 2022 Mar;2. Available from: https://libkey.io/libraries/1108/openurl?genre=article&aulast=DeVito&issn=2772-7246&title=Drug+and+Alcohol+Dependence+Reports&atitle=Modafinil+Does+Not+Reduce+Cocaine+Use+in+Methadone-Maintained+Individuals.&volume=2&issue=&spage=&epage=&date=2022&doi=10.1016%2Fj.dadr.2022.100032&pmid=36310662&sid=OVID:medline

78. Dieckmann L.H.J., Ramos A.C., Silva E.A., Justo L.P., Sabioni P., Frade I.F., et al. Effects of biperiden on the treatment of cocaine/crack addiction: A randomised, double-blind, placebo-controlled trial. Eur Neuropsychopharmacol. 2014;24(8):1196–202.

79. Dursteler-Macfarland K.M., Farronato N.S., Strasser J., Boss J., Kuntze M.F., Petitjean S.A., et al. A randomized, controlled, pilot trial of methylphenidate and cognitive-behavioral group therapy for cocaine dependence in heroin prescription. J Clin Psychopharmacol. 2013;33(1):104–8.

80. Elkashef A., Fudala P.J., Gorgon L., Li S.-H., Kahn R., Chiang N., et al. Double-blind, placebo-controlled trial of selegiline transdermal system (STS) for the treatment of cocaine dependence. Drug Alcohol Depend. 2006;85(3):191–7.

81. Gawin FH, Kleber HD, Byck R, Rounsaville BJ, Kosten TR, Jatlow PI, et al. Desipramine facilitation of initial cocaine abstinence. Archives of General Psychiatry. 1989;46(2):117–21.

82. Kosten TR, Gawin FH, Kosten TA, Morgan C, Rounsaville BJ, Schottenfeld R, et al. Six-month follow-up of short-term pharmacotherapy for cocaine dependence. The American Journal on Addictions. 1992;1(1):40–9.

83. George TP, Chawarski MC, Pakes J, Carroll KM, Kosten TR, Schottenfeld RS. Disulfiram versus placebo for cocaine dependence in buprenorphine-maintained subjects: a preliminary trial. Biol Psychiatry. 2000;47(12):1080–6.

84. Gilgun-Sherki Y., Eliaz R.E., McCann D.J., Loupe P.S., Eyal E., Blatt K., et al. Placebo-controlled evaluation of a bioengineered, cocaine-metabolizing fusion protein, TV-1380 (AlbuBChE), in the treatment of cocaine dependence. Drug Alcohol Depend. 2016;166((Gilgun-Sherki) Formerly Clinical Development&Medicine Section Teva Pharmaceuticals, Petach Tikva, Israel):13–20.

85. Gonzalez G., Sevarino K., Sofuoglu M., Poling J., Oliveto A., Gonsai K., et al. Tiagabine increases cocaine-free urines in cocaine-dependent methadone-treated patients: Results of a randomized pilot study. Addiction. 2003;98(11):1625–32.

86. Gorelick D.A., Wilkins J.N. Bromocriptine treatment for cocaine addiction: Association with plasma prolactin levels. Drug Alcohol Depend. 2006;81(2):189–95.

87. Grabowski J, Rhoades H, Elk R, Schmitz J, Davis C, Creson D, et al. Fluoxetine is ineffective for treatment of cocaine dependence or concurrent opiate and cocaine dependence: two placebo-controlled double-blind trials. J Clin Psychopharmacol. 1995;15(3):163–74.

88. Grabowski J., Roache J.D., Schmitz J.M., Rhoades H., Creson D., Korszun A. Replacement medication for cocaine dependence: Methylphenidate. J Clin Psychopharmacol. 1997;17(6):485–8.

89. Grabowski J., Rhoades H., Schmitz J., Stotts A., Daruzska L.A., Creson D., et al. Dextroamphetamine for cocaine-dependence treatment: A double-blind randomized clinical trial. J Clin Psychopharmacol. 2001;21(5):522–6.

90. Grabowski J., Rhoades H., Stotts A., Cowan K., Kopecky C., Dougherty A., et al. Agonist-like or antagonist-like treatment for cocaine dependence with methadone for heroin dependence: Two double-blind randomized clinical trials. Neuropsychopharmacology. 2004;29(5):969–81.

91. Halikas J.A., Crosby R.D., Pearson V.L., Graves N.M. A randomized double-blind study of carbamazepine in the treatment of cocaine abuse. CLIN PHARMACOL THER. 1997;62(1):89–105.

92. Hall SM, Tunis S, Triffleman E, Banys P, Clark HW, Tusel D, et al. Continuity of care and desipramine in primary cocaine abusers. J Nerv Ment Dis. 1994 Oct;182(10):570–5.

93. Tunis SL, Delucchi KL, Hall SM. Assessing thoughts about cocaine and their relationship to short-term treatment outcome. Experimental and Clinical Psychopharmacology. 1994;2(2):184–93.

94. Hamilton J.D., Nguyen Q.X., Gerber R.M., Rubio N.B. Olanzapine in cocaine dependence: A double-blind, placebo-controlled trial. Am J Addict. 2009;18(1):48–52.

95. Handelsman L, Limpitlaw L, Williams D, Schmeidler J, Paris P, Stimmel B. Amantadine does not reduce cocaine use or craving in cocaine-dependent methadone maintenance patients. Drug and Alcohol Dependence. 1995 Oct 1;39(3):173–80.

96. Handelsman L., Limpitlaw L., Williams D., Schmeidler J., Paris P., Stimmel B. Amantadine does not reduce cocaine use or craving in cocaine-dependent methadone maintenance patients. DRUG ALCOHOL DEPEND. 1995;39(3):173–80.

97. Hersh D., Van Kirk J.R., Kranzler H.R. Naltrexone treatment of comorbid alcohol and cocaine use disorders. Psychopharmacology. 1998;139(1–2):44–52.

98. Johnson B.A., Chen Y.R., Swann A.C., Schmitz J., Lesser J., Ruiz P., et al. Ritanserin in the treatment of cocaine dependence. BIOL PSYCHIATRY. 1997;42(10):932–40.

99. Johnson B.A., Roache J.D., Ait-Daoud N., Javors M.A., Harrison J.M., Elkashef A., et al. A preliminary randomized, double-blind, placebo-controlled study of the safety and efficacy of ondansetron in the treatment of cocaine dependence. Drug Alcohol Depend. 2006;84(3):256–63.

100. Johnson BA, Ait-Daoud N, Wang XQ, Penberthy JK, Javors MA, Seneviratne C, et al. Topiramate for the treatment of cocaine addiction: a randomized clinical trial. JAMA Psychiatry. 2013;70(12):1338–46.

101. Blevins D, Wang XQ, Sharma S, Ait-Daoud N. Impulsiveness as a predictor of topiramate response for cocaine use disorder. The American Journal on Addictions. 2019;28(2):71–6.

102. Johnson M.W., Bruner N.R., Johnson P.S., Silverman K., Berry M.S. Randomized Controlled Trial of D-Cycloserine in Cocaine Dependence: Effects on Contingency Management and Cue-Induced Cocaine Craving in a Naturalistic Setting. Exp Clin Psychopharmacol. 2020;28(2):157–68.

103. Jones H.E., Johnson R.E., Bigelow G.E., Silverman K., Mudric T., Strain E.C. Safety and efficacy of L-tryptophan and behavioral incentives for treatment of cocaine dependence: A randomized clinical trial. Am J Addict. 2004;13(5):421–37.

104. Junior MSC, Bezerra AG, Curado DF, Gregório RP, Galduróz JCF. Preliminary investigation of the administration of biperiden to reduce relapses in individuals with cocaine/crack user disorder: A randomized controlled clinical trial. Pharmacology Biochemistry and Behavior. 2024 Apr 1;237:173725.

105. Kablinger A.S., Lindner M.A., Casso S., Hefti F., Demuth G., Fox B.S., et al. Effects of the combination of metyrapone and oxazepam on cocaine craving and cocaine taking: A double-blind, randomized, placebo-controlled pilot study. J Psychopharmacol. 2012;26(7):973–81.

106. Kahn R., Biswas K., Childress A.-R., Shoptaw S., Fudala P.J., Gorgon L., et al. Multi-center trial of baclofen for abstinence initiation in severe cocaine-dependent individuals. Drug Alcohol Depend. 2009;103(1–2):59–64.

107. Kampman K., Volpicelli J.R., Alterman A., Cornish J., Weinrieb R., Epperson L., et al. Amantadine in the early treatment of cocaine dependence: A double-blind, placebo-controlled trial. DRUG ALCOHOL DEPEND. 1996;41(1):25–33.

108. Kampman KM, Volpicelli JR, Alterman AI, Cornish J, O’Brien CP. Amantadine in the Treatment of Cocaine-Dependent Patients With Severe Withdrawal Symptoms. AJP. 2000 Dec 1;157(12):2052–4.

109. Kampman KM, Volpicelli JR, Mulvaney F, Alterman AI, Cornish J, Gariti P, et al. Effectiveness of propranolol for cocaine dependence treatment may depend on cocaine withdrawal symptom severity. Drug Alcohol Depend. 2001;63(1):69–78.

110. Kampman KM, Pettinati H, Lynch KG, Sparkman T, O’Brien CP. A pilot trial of olanzapine for the treatment of cocaine dependence. Drug and Alcohol Dependence. 2003 June 5;70(3):265–73.

111. Kampman K, Majewska MD, Tourian K, Dackis C, Cornish J, Poole S, et al. A pilot trial of piracetam and ginkgo biloba for the treatment of cocaine dependence. Addict Behav. 2003 Apr;28(3):437–48.

112. Kampman K.M., Pettinati H., Lynch K.G., Dackis C., Sparkman T., Weigley C., et al. A pilot trial of topiramate for the treatment of cocaine dependence. Drug Alcohol Depend. 2004;75(3):233–40.

113. Kampman K.M., Dackis C., Lynch K.G., Pettinati H., Tirado C., Gariti P., et al. A double-blind, placebo-controlled trial of amantadine, propranolol, and their combination for the treatment of cocaine dependence in patients with severe cocaine withdrawal symptoms. Drug Alcohol Depend. 2006;85(2):129–37.

114. Fairbairn CE, Dundon WD, Xie H, Plebani JG, Kampman KM, Lynch KG. Study blinding and correlations between perceived group assignment and outcome in a cocaine pharmacotherapy trial. Am J Addict. 2008 Sept;17(5):387–91.

115. Kampman K.M., Dackis C., Pettinati H.M., Lynch K.G., Sparkman T., O’Brien C.P. A double-blind, placebo-controlled pilot trial of acamprosate for the treatment of cocaine dependence. Addict Behav. 2011;36(3):217–21.

116. Kampman K.M., Pettinati H.M., Lynch K.G., Spratt K., Wierzbicki M.R., O’Brien C.P. A double-blind, placebo-controlled trial of topiramate for the treatment of comorbid cocaine and alcohol dependence. Drug Alcohol Depend. 2013;133(1):94–9.

117. Kampman K.M., Lynch K.G., Pettinati H.M., Spratt K., Wierzbicki M.R., Dackis C., et al. A double blind, placebo controlled trial of modafinil for the treatment of cocaine dependence without co-morbid alcohol dependence. Drug Alcohol Depend. 2015;155((Kampman, Lynch, Pettinati, Dackis, O’Brien) Department of Psychiatry, Perelman School of Medicine, University of Pennsylvania, 3900 Chestnut Street, Philadelphia, PA 19104, United States):105–10.

118. Karila L., Leroy C., Dubol M., Trichard C., Mabondo A., Marill C., et al. Dopamine transporter correlates and occupancy by modafinil in cocaine-dependent patients: A controlled study with high-resolution PET and [11 C]-PE2I. Neuropsychopharmacology. 2016;41(9):2294–302.

119. Kennedy AP, Gross RE, Whitfield N, Drexler KP, Kilts CD. A controlled trial of the adjunct use of D-cycloserine to facilitate cognitive behavioral therapy outcomes in a cocaine-dependent population. Addict Behav. 2012 Aug;37(8):900–7.

120. Kolar A.F., Brown B.S., Weddington W.W., Haertzen C.C., Michaelson B.S., Jaffe J.H. Treatment of cocaine dependence in methadone maintenance clients: A pilot study comparing the efficacy of desipramine and amantadine. INT J ADDICT. 1992;27(7):849–68.

121. Kosten TR, Morgan CM, Falcione J, Schottenfeld RS. Pharmacotherapy for cocaine-abusing methadone-maintained patients using amantadine or desipramine. Archives of General Psychiatry. 1992;49(11):894–8.

122. Leal J, Ziedonis D, Kosten T. Antisocial personality disorder as a prognostic factor for pharmacotherapy of cocaine dependence. Drug Alcohol Depend. 1994 Mar;35(1):31–5.

123. Ziedonis DM, Kosten TR. Depression as a prognostic factor for pharmacological treatment of cocaine dependence. Psychopharmacol Bull. 1991;27(3):337–43.

124. Ziedonis DM, Kosten TR. Pharmacotherapy Improves Treatment Outcome in Depressed Cocaine Addicts. Journal of Psychoactive Drugs. 1991 Oct 1;23(4):417–25.

125. Kosten TR, Oliveto A, Sevarino KA, Gonsai K, Feingold A. Ketoconazole increases cocaine and opioid use in methadone maintained patients. Drug Alcohol Depend. 2002;66(2):173–80.

126. Kosten T, Oliveto A, Feingold A, Poling J, Sevarino K, McCance-Katz E, et al. Desipramine and contingency management for cocaine and opiate dependence in buprenorphine maintained patients. Drug Alcohol Depend. 2003;70(3):315–25.

127. Gonzalez G, Feingold A, Oliveto A, Gonsai K, Kosten TR. Comorbid major depressive disorder as a prognostic factor in cocaine-abusing buprenorphine-maintained patients treated with desipramine and contingency management. Am J Drug Alcohol Abuse. 2003 Aug;29(3):497–514.

128. Kosten T, Poling J, Oliveto A. Effects of reducing contingency management values on heroin and cocaine use for buprenorphine- and desipramine-treated patients. Addiction. 2003 May;98(5):665–71.

129. Sofuoglu M, Gonzalez G, Poling J, Kosten TR. Prediction of treatment outcome by baseline urine cocaine results and self-reported cocaine use for cocaine and opioid dependence. Am J Drug Alcohol Abuse. 2003;29(4):713–27.

130. Kosten T.R., Wu G., Huang W., Harding M.J., Hamon S.C., Lappalainen J., et al. Pharmacogenetic randomized trial for cocaine abuse: Disulfiram and dopamine beta-hydroxylase. Biol Psychiatry. 2013;73(3):219–24.

131. Kampangkaew JP, Spellicy CJ, Nielsen EM, Harding MJ, Ye A, Hamon SC, et al. Pharmacogenetic role of dopamine transporter (SLC6A3) variation on response to disulfiram treatment for cocaine addiction. Am J Addict. 7 AD;28(4):311–7.

132. Nielsen DA, Harding MJ, Hamon SC, Huang W, Kosten TR. Modifying the role of serotonergic 5-HTTLPR and TPH2 variants on disulfiram treatment of cocaine addiction: a preliminary study. Genes Brain Behav. 2012 Nov;11(8):1001–8.

133. Shorter D, Nielsen DA, Huang W, Harding MJ, Hamon SC, Kosten TR. Pharmacogenetic randomized trial for cocaine abuse: disulfiram and alpha1A-adrenoceptor gene variation. Eur Neuropsychopharmacol. 2013 Nov;23(11):1401–7.

134. Spellicy CJ, Kosten TR, Hamon SC, Harding MJ, Nielsen DA. The MTHFR C677T Variant is Associated with Responsiveness to Disulfiram Treatment for Cocaine Dependency. Front Psychiatr. 2012;3:109.

135. Kosten T.R., Domingo C.B., Shorter D., Orson F., Green C., Somoza E., et al. Vaccine for cocaine dependence: A randomized double-blind placebo-controlled efficacy trial. Drug Alcohol Depend. 2014;140((Kosten, Domingo, Shorter, Orson) Michael E. DeBakey VA Medical Center, Baylor College of Medicine, 2002 Holcombe, Bldg. 121, Rm 141, Houston, TX 77030, United States):42–7.

136. Kranzler HR, Bauer LO, Hersh D, Klinghoffer V. Carbamazepine treatment of cocaine dependence: a placebo-controlled trial. Drug Alcohol Depend. 1995;38(3):203–11.

137. LaRowe S.D., Kalivas P.W., Nicholas J.S., Randall P.K., Mardikian P.N., Malcolm R.J. A double-blind placebo-controlled trial of N-acetylcysteine in the treatment of cocaine dependence. Am J Addict. 2013;22(5):443–52.

138. Levin FR, McDowell D, Evans SM, Brooks D, Spano C, Nunes EV. Pergolide mesylate for cocaine abuse: a controlled preliminary trial. Am J Addict. 1999;8(2):120–7.

139. Levin F.R., Evans S.M., Brooks D.J., Garawi F. Treatment of cocaine dependent treatment seekers with adult ADHD: Double-blind comparison of methylphenidate and placebo. Drug Alcohol Depend. 2007;87(1):20–9.

140. Aharonovich E, Garawi F, Bisaga A, Brooks D, Raby WN, Rubin E, et al. Concurrent cannabis use during treatment for comorbid ADHD and cocaine dependence: effects on outcome. Am J Drug Alcohol Abuse. 2006;32(4):629–35.

141. Levin F.R., Mariani J.J., Specker S., Mooney M., Mahony A., Brooks D.J., et al. Extended-release mixed amphetamine salts vs placebo for comorbid adult attention-deficit/hyperactivity disorder and cocaine use disorder a randomized clinical trial. JAMA Psychiatry. 2015;72(6):593–602.

142. Blevins D, Choi CJ, Pavlicova M, Martinez D, Mariani JJ, Grabowski J, et al. Impulsiveness as a moderator of amphetamine treatment response for cocaine use disorder among ADHD patients. Drug Alcohol Depend. 2020 Aug;213:108082.

143. Levin FR, Choi CJ, Pavlicova M, Mariani JJ, Mahony A, Brooks DJ, et al. How treatment improvement in ADHD and cocaine dependence are related to one another: A secondary analysis. Drug and Alcohol Dependence. 2018 July 1;188:135–40.

144. Notzon DP, Mariani JJ, Pavlicova M, Glass A, Mahony AL, Brooks DJ, et al. Mixed-amphetamine salts increase abstinence from marijuana in patients with co-occurring attention-deficit/hyperactivity disorder and cocaine dependence. Am J Addict. 12 AD;25(8):666–72.

145. Levin F.R., Mariani J.J., Pavlicova M., Choi C.J., Mahony A.L., Brooks D.J., et al. Extended release mixed amphetamine salts and topiramate for cocaine dependence: A randomized clinical replication trial with frequent users. Drug Alcohol Depend. 2020;206((Levin, Mariani, Mahony, Brooks, Bisaga, Dakwar, Carpenter, Naqvi, Nunes) New York State Psychiatric Institute, Division on Substance Use Disorders, 1051 Riverside Drive, New York, NY 10032, United States):107700.

146. Licata S.C., Penetar D.M., Ravichandran C., Rodolico J., Palmer C., Berko J., et al. Effects of daily treatment with citicoline: A double-blind, placebo-controlled study in cocaine-dependent volunteers. J Addict Med. 2011;5(1):57–64.

147. Bracken BK, Penetar DM, Rodolico J, Ryan ET, Lukas SE. Eight weeks of citicoline treatment does not perturb sleep/wake cycles in cocaine-dependent adults. Pharmacol Biochem Behav. 2011 June;98(4):518–24.

148. Ling W, Hillhouse MP, Saxon AJ, Mooney LJ, Thomas CM, Ang A, et al. Buprenorphine + naloxone plus naltrexone for the treatment of cocaine dependence: The Cocaine Use Reduction with Buprenorphine (CURB) study. Addiction. 2016;111(8):1416–27.

149. Nielsen DA, Walker R, Graham DP, Nielsen EM, Hamon SC, Hillhouse M, et al. Moderation of buprenorphine therapy for cocaine dependence efficacy by variation of the Prodynorphin gene. Eur J Clin Pharmacol. 2022 June;78(6):965–73.

150. Loebl T., Angarita G.A., Pachas G.N., Huang K.-L., Lee S.H., Nino J., et al. A randomized, double-blind, placebo-controlled trial of long-acting risperidone in cocaine-dependent men. J Clin Psychiatry. 2008;69(3):480–6.

151. Lynch KG, Plebani J, Spratt K, Morales M, Tamminga M, Feibush P, et al. Varenicline for the Treatment of Cocaine Dependence. J Addict Med. 2022 Mar;16(2):157–63.

152. Malcolm R, Kajdasz DK, Herron J, Anton RF, Brady KT. A double-blind, placebo-controlled outpatient trial of pergolide for cocaine dependence. Drug Alcohol Depend. 2000;60(2):161–8.

153. Malcolm R., Herron J., Sutherland S.E., Brady K.T. Adverse outcomes in a controlled trial of pergolide for cocaine dependence. J Addict Dis. 2001;20(1):81–92.

154. Malcolm R., LaRowe S., Cochran K., Moak D., Herron J., Brady K., et al. A controlled trial of amlodipine for cocaine dependence: A negative report. J Subst Abuse Treat. 2005;28(2):197–204.

155. Malcolm R, Liao J, Michel M, Cochran K, Pye W, Yeager D, et al. Amlodipine reduces blood pressure and headache frequency in cocaine-dependent outpatients. J Psychoactive Drugs. 2002 Oct;34(4):415–9.

156. Turner TH, LaRowe S, Horner MD, Herron J, Malcolm R. Measures of cognitive functioning as predictors of treatment outcome for cocaine dependence. J Subst Abuse Treat. 2009 Dec;37(4):328–34.

157. Mancino M.J., McGaugh J., Chopra M.P., Guise J.B., Cargile C., Williams D.K., et al. Clinical efficacy of sertraline alone and augmented with gabapentin in recently abstinent cocaine-dependent patients with depressive symptoms. J Clin Psychopharmacol. 2014;34(2):234–9.

158. Margolin A., Kosten T.R., Kelly Avants S., Wilkins J., Ling W., Beckson M., et al. A multicenter trial of bupropion for cocaine dependence in methadone-maintained patients. DRUG ALCOHOL DEPEND. 1995;40(2):125–31.

159. Margolin A., Avants S.K., Malison R.T., Kosten T.R. High- and low-dose mazindol for cocaine dependence in methadone-maintained patients: A preliminary evaluation. SUBST ABUSE. 1997;18(3):125–31.

160. Mariani J.J., Pavlicova M., Bisaga A., Nunes E.V., Brooks D.J., Levin F.R. Extended-release mixed amphetamine salts and topiramate for cocaine dependence: A randomized controlled trial. Biol Psychiatry. 2012;72(11):950–6.

161. Martell B.A., Orson F.M., Poling J., Mitchell E., Rossen R.D., Gardner T., et al. Cocaine vaccine for the treatment of cocaine dependence in methadone-maintained patients: A randomized, double-blind, placebo-controlled efficacy trial. Arch Gen Psychiatry. 2009;66(10):1116–23.

162. Kosten TR, Domingo CB, Hamon SC, Nielsen DA. DBH gene as predictor of response in a cocaine vaccine clinical trial. Neurosci Lett. 2013 Apr 29;541:29–33.

163. Nielsen DA, Hamon SC, Kosten TR. The kappa-opioid receptor gene as a predictor of response in a cocaine vaccine clinical trial. Psychiatr Genet. 2013 Dec;23(6):225–32.

164. McCann DJ, Chen HH, Devine EG, Gyaw S, Ramey T. Results of a randomized, double-blind, placebo-controlled trial of lorcaserin in cocaine use disorder. Drug Alcohol Depend. 2024 Feb;255:111063.

165. McDowell D., Nunes E.V., Seracini A.M., Rothenberg J., Vosburg S.K., Ma G.J., et al. Desipramine treatment of cocaine-dependent patients with depression: A placebo-controlled trial. Drug Alcohol Depend. 2005;80(2):209–21.

166. Moeller FG, Dougherty DM, Barratt ES, Schmitz JM, Swann AC, Grabowski J. The impact of impulsivity on cocaine use and retention in treatment. J Subst Abuse Treat. 2001 Dec;21(4):193–8.

167. Green CE, Moeller FG, Schmitz JM, Lucke JF, Lane SD, Swann AC, et al. Evaluation of heterogeneity in pharmacotherapy trials for drug dependence: a Bayesian approach. Am J Drug Alcohol Abuse. 2009;35(2):95–102.

168. Schmitz JM, Mooney ME, Green CE, Lane SD, Steinberg JL, Swann AC, et al. Baseline neurocognitive profiles differentiate abstainers and non-abstainers in a cocaine clinical trial. J Addict Dis. 2009 July;28(3):250–7.

169. Mongeau-Perusse V., Brissette S., Bruneau J., Conrod P., Dubreucq S., Gazil G., et al. Cannabidiol as a treatment for craving and relapse in individuals with cocaine use disorder: a randomized placebo-controlled trial. Addiction. 2021;116(9):2431–42.

170. Hebert FO, Mongeau-Perusse V, Rizkallah E, Mahroug A, Bakouni H, Morissette F, et al. Absence of Evidence for Sustained Effects of Daily Cannabidiol Administration on Anandamide Plasma Concentration in Individuals with Cocaine Use Disorder: Exploratory Findings from a Randomized Controlled Trial. Cannabis Cannabinoid Res. 2024 May 21;21:21.

171. Mongeau-Perusse V, Rizkallah E, Morissette F, Brissette S, Bruneau J, Dubreucq S, et al. Cannabidiol Effect on Anxiety Symptoms and Stress Response in Individuals With Cocaine Use Disorder: Exploratory Results From a Randomized Controlled Trial. J Addict Med. 2022 Sept;16(5):521–6.

172. Morissette F, Mongeau-Perusse V, Rizkallah E, Thebault P, Lepage S, Brissette S, et al. Exploring cannabidiol effects on inflammatory markers in individuals with cocaine use disorder: a randomized controlled trial. Neuropsychopharmacology. 11 AD;46(12):2101–11.

173. Rizkallah E, Mongeau-Perusse V, Lamanuzzi L, Castenada-Ouellet S, Stip E, Juteau LC, et al. Cannabidiol effects on cognition in individuals with cocaine use disorder: Exploratory results from a randomized controlled trial. Pharmacol Biochem Behav. 5 AD;216:173376.

174. Montoya I.D., Levin F.R., Fudala P.J., Gorelick D.A. Double-blind comparison of carbamazepine and placebo for treatment of cocaine dependence. DRUG ALCOHOL DEPEND. 1995;38(3):213–9.

175. Montoya ID, Schroeder JR, Preston KL, Covi L, Umbricht A, Contoreggi C, et al. Influence of psychotherapy attendance on buprenorphine treatment outcome. J Subst Abuse Treat. 2005 Apr;28(3):247–54.

176. Mooney ME, Schmitz JM, Moeller FG, Grabowski J. Safety, tolerability and efficacy of levodopa-carbidopa treatment for cocaine dependence: two double-blind, randomized, clinical trials. Drug Alcohol Depend. 2007;88(2–3):214–23.

177. Mooney M.E., Herin D.V., Schmitz J.M., Moukaddam N., Green C.E., Grabowski J. Effects of oral methamphetamine on cocaine use: A randomized, double-blind, placebo-controlled trial. Drug Alcohol Depend. 2009;101(1–2):34–41.

178. Mooney M.E., Herin D.V., Specker S., Babb D., Levin F.R., Grabowski J. Pilot study of the effects of lisdexamfetamine on cocaine use: A randomized, double-blind, placebo-controlled trial. Drug Alcohol Depend. 2015;153((Mooney, Herin, Specker, Babb, Grabowski) Department of Psychiatry, University of Minnesota, Minneapolis, United States):94–103.

179. Morgan P.T., Angarita G.A., Canavan S., Pittman B., Oberleitner L., Malison R.T., et al. Modafinil and sleep architecture in an inpatient-outpatient treatment study of cocaine dependence. Drug Alcohol Depend. 2016;160((Morgan, Angarita, Canavan, Pittman, Oberleitner, Malison, Hodges, Easton, McKee, Bessette, Forselius) Connecticut Mental Health Center, Yale University Department of Psychiatry, 34 Park Street, New Haven, CT 06519, United States):49–56.

180. Canavan SV, Forselius EL, Bessette AJ, Morgan PT. Preliminary evidence for normalization of risk taking by modafinil in chronic cocaine users. Addict Behav. 2014 June;39(6):1057–61.

181. Nanni-Alvarado R, Gonzalez M, Lima C, Marin-Navarrete R, Barbosa-Mendez S, Salazar-Juarez A. Effect of mirtazapine on craving in cocaine-dependent patients. International Journal of Mental Health and Addiction. 2022;20(5):2770–86.

182. Milivojevic V, Charron L, Fogelman N, Hermes G, Sinha R. Pregnenolone Reduces Stress-Induced Craving, Anxiety, and Autonomic Arousal in Individuals with Cocaine Use Disorder. Biomolecules. 10 29;12(11):29.

183. Raby W.N., Rubin E.A., Garawi F., Cheng W., Mason E., Sanfilippo L., et al. A randomized, double-blind, placebo-controlled trial of venlafaxine for the treatment of depressed cocaine-dependent patients. Am J Addict. 2014;23(1):68–75.

184. Noel Raby W, Heller M, Milliaressis D, Jean Choi C, Basaraba C, Pavlicova M, et al. Intranasal oxytocin may improve odds of abstinence in cocaine-dependent patients: results from a preliminary study. Drug Alcohol Depend Rep. 2022 Mar;2:100016.

185. Nuijten M., Blanken P., Van De Wetering B., Nuijen B., Van Den Brink W., Hendriks V.M. Sustained-release dexamfetamine in the treatment of chronic cocaine-dependent patients on heroin-assisted treatment: A randomised, double-blind, placebo-controlled trial. Lancet. 2016;387(10034):2226–34.

186. Blanken P, Nuijten M, van den Brink W, Hendriks VM. Clinical effects beyond cocaine use of sustained-release dexamphetamine for the treatment of cocaine dependent patients with comorbid opioid dependence: secondary analysis of a double-blind, placebo-controlled randomized trial. Addiction. 2020;115(5):917–23.

187. Oliva A, Reed SC, Brooks DJ, Levin FR, Evans SM. Safety and tolerability of progesterone treatment for women with cocaine use disorder: a pilot treatment trial. Am J Drug Alcohol Abuse. 2022;48(5):586–95.

188. Mancino MJ, McGaugh J, Feldman Z, Poling J, Oliveto A. Effect of PTSD diagnosis and contingency management procedures on cocaine use in dually cocaine- and opioid-dependent individuals maintained on LAAM: a retrospective analysis. Am J Addict. 2010 Mar;19(2):169–77.

189. Oliveto A., Poling J., Mancino M.J., Feldman Z., Cubells J.F., Pruzinsky R., et al. Randomized, double blind, placebo-controlled trial of disulfiram for the treatment of cocaine dependence in methadone-stabilized patients. Drug Alcohol Depend. 2011;113(2–3):184–91.

190. Atkinson TS, Sanders N, Mancino M, Oliveto A. Effects of disulfiram on QTc interval in non-opioid-dependent and methadone-treated cocaine-dependent patients. J Addict Med. 2013 July;7(4):243–8.

191. Oliveto A, Poling J, Mancino MJ, Williams DK, Thostenson J, Pruzinsky R, et al. Sertraline delays relapse in recently abstinent cocaine-dependent patients with depressive symptoms. Addiction. 2012;107(1):131–41.

192. Passos S.R.L., Camacho L.A.B., Lopes C.S., Dos Santos M.A.B. Nefazodone in out-patient treatment of inhaled cocaine dependence: A randomized double-blind placebo-controlled trial. Addiction. 2005;100(4):489–94.

193. Petrakis I.L., Carroll K.M., Nich C., Gordon L.T., Mccance-Katz E.F., Frankforter T., et al. Disulfiram treatment for cocaine dependence in methadone-maintained opioid addicts. Addiction. 2000;95(2):219–28.

194. Jofre-Bonet M, Sindelar JL, Petrakis IL, Nich C, Frankforter T, Rounsaville BJ, et al. Cost effectiveness of disulfiram: treating cocaine use in methadone-maintained patients. J Subst Abuse Treat. 2004 Apr;26(3):225–32.

195. Pettinati H.M., Kampman K.M., Lynch K.G., Xie H., Dackis C., Rabinowitz A.R., et al. A double blind, placebo-controlled trial that combines disulfiram and naltrexone for treating co-occurring cocaine and alcohol dependence. Addict Behav. 2008;33(5):651–67.

196. Pettinati HM, Kampman KM, Lynch KG, Dundon WD, Mahoney EM, Wierzbicki MR, et al. A pilot trial of injectable, extended-release naltrexone for the treatment of co-occurring cocaine and alcohol dependence. The American Journal on Addictions. 2014;23(6):591–7.

197. Plebani J.G., Lynch K.G., Yu Q., Pettinati H.M., O’Brien C.P., Kampman K.M. Results of an initial clinical trial of varenicline for the treatment of cocaine dependence. Drug Alcohol Depend. ((Plebani, Lynch, Yu, Pettinati, O’Brien, Kampman) University of Pennsylvania, Department of Psychiatry, Treatment Research Center, 3900 Chestnut Street, Philadelphia, PA 19104, United States).

198. Reid M.S., Angrist B., Baker S., Woo C., Schwartz M., Montgomery A., et al. A placebo-controlled screening trial of celecoxib for the treatment of cocaine dependence. Addiction. 2005;100(SUPPL. 1):32–42.

199. Reid M.S., Casadonte P., Baker S., Sanfilipo M., Braunstein D., Hitzemann R., et al. A placebo-controlled screening trial of olanzapine, valproate, and coenzyme Q10/L-carnitine for the treatment of cocaine dependence. Addiction. 2005;100(SUPPL. 1):43–57.

200. Reid M.S., Angrist B., Baker S.A., O’Leary S., Stone J., Schwartz M., et al. A placebo controlled, double-blind study of mecamylamine treatment for cocaine dependence in patients enrolled in an opiate replacement program. Subst Abuse. 2006;26(2):5–14.

201. Santos GM, Ikeda J, Coffin P, Walker JE, Matheson T, McLaughlin M, et al. Pilot study of extended-release lorcaserin for cocaine use disorder among men who have sex with men: A double-blind, placebo-controlled randomized trial. PLoS ONE. 2021;16(7).

202. Schmitz JM, Averill P, Stotts AL, Moeller FG, Rhoades HM, Grabowski J. Fluoxetine treatment of cocaine-dependent patients with major depressive disorder. Drug Alcohol Depend. 2001;63(3):207–14.

203. Schmitz J.M., Mooney M.E., Moeller F.G., Stotts A.L., Green C., Grabowski J. Levodopa pharmacotherapy for cocaine dependence: Choosing the optimal behavioral therapy platform. Drug Alcohol Depend. 2008;94(1–3):142–50.

204. Schmitz J.M., Lindsay J.A., Green C.E., Herin D.V., Stotts A.L., Gerard Moeller F. High-dose naltrexone therapy for cocaine-alcohol dependence. Am J Addict. 2009;18(5):356–62.

205. Schmitz J.M., Rathnayaka N., Green C.E., Moeller F.G., Dougherty A.E., Grabowski J. Combination of modafinil and d-amphetamine for the treatment of cocaine dependence: A preliminary investigation. Front Psychiatry. 2012;3(AUG):Article 77.

206. Schmitz J.M., Green C.E., Stotts A.L., Lindsay J.A., Rathnayaka N.S., Grabowski J., et al. A two-phased screening paradigm for evaluating candidate medications for cocaine cessation or relapse prevention: Modafinil, levodopa-carbidopa, naltrexone. Drug Alcohol Depend. 2014;136(1):100–7.

207. Schmitz J.M., Green C.E., Hasan K.M., Vincent J., Suchting R., Weaver M.F., et al. PPAR-gamma agonist pioglitazone modifies craving intensity and brain white matter integrity in patients with primary cocaine use disorder: a double-blind randomized controlled pilot trial. Addiction. 2017;112(10):1861–8.

208. Schmitz JM, Suchting R, Green CE, Webber HE, Vincent J, Moeller FG, et al. The effects of combination levodopa-ropinirole on cognitive improvement and treatment outcome in individuals with cocaine use disorder: A bayesian mediation analysis. Drug and Alcohol Dependence. 2021 Aug 1;225:108800.

209. Schmitz JM, Stotts AL, Vujanovic AA, Yoon JH, Webber HE, Lane SD, et al. Contingency management plus acceptance and commitment therapy for initial cocaine abstinence: Results of a sequential multiple assignment randomized trial (SMART). Drug Alcohol Depend. 2024 Mar;256:111078.

210. Schubiner H., Saules K.K., Arfken C.L., Johanson C.-E., Schuster C.R., Lockhart N., et al. Double-blind placebo-controlled trial of methylphenidate in the treatment of adult ADHD patients with comorbid cocaine dependence. Exp Clin Psychopharmacol. 2002;10(3):286–94.

211. Shearer J., Wodak A., Van Beek I., Mattick R.P., Lewis J. Pilot randomized double blind placebo-controlled study of dexamphetamine for cocaine dependence. Addiction. 2003;98(8):1137–41.

212. Shoptaw S., Kintaudi P.C., Charuvastra C., Ling W. A screening trial of amantadine as a medication for cocaine dependence. Drug Alcohol Depend. 2002;66(3):217–24.

213. Shoptaw S., Yang X., Rotheram-Fuller E.J., Hsieh Y.-C.M., Kintaudi P.C., Charuvastra V.C., et al. Randomized placebo-controlled trial of baclofen for cocaine dependence: Preliminary effects for individuals with chronic patterns of cocaine use. J Clin Psychiatry. 2003;64(12):1440–8.

214. Shoptaw S, Majewska MD, Wilkins J, Twitchell G, Yang X, Ling W. Participants receiving dehydroepiandrosterone during treatment for cocaine dependence show high rates of cocaine use in a placebo-controlled pilot study. Experimental and clinical psychopharmacology. 2004;12(2):126.

215. Shoptaw S., Watson D.W., Reiber C., Rawson R.A., Montgomery M.A., Majewska M.D., et al. Randomized controlled pilot trial of cabergoline, hydergine and levodopa/carbidopa: Los Angeles Cocaine Rapid Efficacy Screening Trial (CREST). Addiction. 2005;100(SUPPL. 1):78–90.

216. Shorter D, Lindsay JA, Kosten TR. The alpha-1 adrenergic antagonist doxazosin for treatment of cocaine dependence: A pilot study. Drug and Alcohol Dependence. 2013 July 1;131(1):66–70.

217. Sofuoglu M., Poling J., Gonzalez G., Gonsai K., Oliveto A., Kosten T.R. Progesterone Effects on Cocaine Use in Male Cocaine Users Maintained on Methadone: A Randomized, Double-Blind, Pilot Study. Exp Clin Psychopharmacol. 2007;15(5):453–60.

218. Sofuoglu M, Carroll KM. Effects of galantamine on cocaine use in chronic cocaine users. Am J Addict. 2011 May;20(3):302–3.

219. Sofuoglu M., Poling J., Babuscio T., Gonsai K., Severino K., Nich C., et al. Carvedilol does not reduce cocaine use in methadone-maintained cocaine users. J Subst Abuse Treat. 2017;73((Sofuoglu, Poling, Babuscio, Gonsai, Severino, Nich, Carroll) VA Connecticut Healthcare System, West Haven, CT, United States):63–9.

220. Somoza E.C., Winship D., Gorodetzky C.W., Lewis D., Ciraulo D.A., Galloway G.P., et al. A multisite, double-blind, placebo-controlled clinical trial to evaluate the safety and efficacy of vigabatrin for treating cocaine dependence. JAMA Psychiatry. 2013;70(6):630–7.

221. Berezina TL, Khouri AS, Winship MD, Fechtner RD. Visual field and ocular safety during short-term vigabatrin treatment in cocaine abusers. Am J Ophthalmol. 2012 Aug;154(2):326–332.e2.

222. Stine S.M., Krystal J.H., Kosten T.R., Charney D.S. Mazindol treatment for cocaine dependence. DRUG ALCOHOL DEPEND. 1995;39(3):245–52.

223. Suchting R, Green CE, de Dios C, Vincent J, Moeller FG, Lane SD, et al. Citalopram for treatment of cocaine use disorder: A Bayesian drop-the-loser randomized clinical trial. Drug Alcohol Depend. 2021;228:109054.

224. de Dios C, Suchting R, Green CE, Webber HE, Moeller FG, Lane SD, et al. The role of Iowa gambling task performance in response to citalopram treatment for cocaine use disorder. Am J Drug Alcohol Abuse. 2025 Jan 24;1–12.

225. Tapp A., Wood A.E., Kennedy A., Sylvers P., Kilzieh N., Saxon A.J. Quetiapine for the treatment of cocaine use disorder. Drug Alcohol Depend. 2015;149((Tapp) VA Puget Sound Health Care System, American Lake Division (A-116), University of Washington, 9600 Veterans Drive SW, Tacoma, WA 98493, United States):18–24.

226. Umbricht A., DeFulio A., Winstanley E.L., Tompkins D.A., Peirce J., Mintzer M.Z., et al. Topiramate for cocaine dependence during methadone maintenance treatment: A randomized controlled trial. Drug Alcohol Depend. 2014;140((Umbricht, DeFulio, Winstanley, Tompkins, Peirce, Mintzer, Strain, Bigelow) Behavioral Pharmacology Research Unit, Department of Psychiatry and Behavioral Sciences, Johns Hopkins School of Medicine, 5510 Nathan Shock Drive, Baltimore, MD 21224, United States):92–100.

227. Rass O, Umbricht A, Bigelow GE, Strain EC, Johnson MW, Mintzer MZ. Topiramate impairs cognitive function in methadone-maintained individuals with concurrent cocaine dependence. Psychol Addict Behav. 2015 Mar;29(1):237–46.

228. Walsh S.L., Middleton L.S., Wong C.J., Nuzzo P.A., Campbell C.L., Rush C.R., et al. Atomoxetine does not alter cocaine use in cocaine dependent individuals: A double blind randomized trial. Drug Alcohol Depend. 2013;130(1–3):150–7.

229. Wardle M.C., Vincent J.N., Suchting R., Green C.E., Lane S.D., Schmitz J.M. Anhedonia Is Associated with Poorer Outcomes in Contingency Management for Cocaine Use Disorder. J Subst Abuse Treat. 2017;72((Wardle, Vincent, Suchting, Green, Lane, Schmitz) Center for Neurobehavioral Research on Addiction, University of Texas Medical Center at Houston, 1941 East Rd., Houston, TX 77054, United States):32–9.

230. Ware OD, Sweeney MM, Cunningham C, Umbricht A, Stitzer M, Dunn KE. Bupropion Slow Release vs Placebo With Adaptive Incentives for Cocaine Use Disorder in Persons Receiving Methadone for Opioid Use Disorder: A Randomized Clinical Trial. JAMA Netw Open. 2023 Mar 15;6(3):e232278.

231. Winhusen TM, Somoza EC, Harrer JM, Mezinskis JP, Montgomery MA, Goldsmith RJ, et al. A placebo-controlled screening trial of tiagabine, sertraline and donepezil as cocaine dependence treatments. Addiction. 2005 Mar;100 Suppl 1:68–77.

232. Winhusen T, Somoza E, Sarid-Segal O, Goldsmith RJ, Harrer JM, Coleman FS, et al. A double-blind, placebo-controlled trial of reserpine for the treatment of cocaine dependence. Drug and Alcohol Dependence. 2007 Dec 1;91(2):205–12.

233. Winhusen TM, Lewis DF, Somoza EC, Horn P. Pharmacodynamics must inform statistics: an example from a cocaine dependence pharmacotherapy trial. ISRN Addict. 2014;2014:927290.

234. Winhusen T., Somoza E., Ciraulo D.A., Harrer J.M., Goldsmith R.J., Grabowski J., et al. A double-blind, placebo-controlled trial of tiagabine for the treatment of cocaine dependence. Drug Alcohol Depend. 2007;91(2–3):141–8.

235. Winhusen TM, Kropp F, Lindblad R, Douaihy A, Haynes L, Hodgkins C, et al. Multisite, randomized, double-blind, placebo-controlled pilot clinical trial to evaluate the efficacy of buspirone as a relapse-prevention treatment for cocaine dependence. J Clin Psychiatry. 2014 July;75(7):757–64.

236. Winstanley E.L., Bigelow G.E., Silverman K., Johnson R.E., Strain E.C. A randomized controlled trial of fluoxetine in the treatment of cocaine dependence among methadone-maintained patients. J Subst Abuse Treat. 2011;40(3):255–64.

237. Zhang X, Nielsen DA, Domingo CB, Shorter DI, Nielsen EM, Kosten TR. Pharmacogenetics of *Dopamine* β-*Hydroxylase* in cocaine dependence therapy with doxazosin. Addiction Biology. 2019 May;24(3):531–8.

238. Shorter DI, Zhang X, Domingo CB, Nielsen EM, Kosten TR, Nielsen DA. Doxazosin treatment in cocaine use disorder: pharmacogenetic response based on an alpha-1 adrenoreceptor subtype D genetic variant. Am J Drug Alcohol Abuse. 2020;46(2):184–93.

239. Spiga F, Parkhouse T, Tang VM, Savović J, Foll BL, Nielsen S. Pharmacotherapies for cannabis use disorder - Spiga, F - 2025 | Cochrane Library. [cited 2025 Oct 31]; Available from: https://www.cochranelibrary.com/cdsr/doi/10.1002/14651858.CD008940.pub4/full

240. Brandt L, Chao T, Comer SD, Levin FR. Pharmacotherapeutic strategies for treating cocaine use disorder—what do we have to offer? Addiction. 2021;116(4):694–710.

241. Pani PP, Trogu E, Vecchi S, Amato L. Antidepressants for cocaine dependence and problematic cocaine use - Pani, PP - 2011 | Cochrane Library. [cited 2025 Sept 18]; Available from: https://www.cochranelibrary.com/cdsr/doi/10.1002/14651858.CD002950.pub3/full

242. Lassi DLS, Malbergier A, Negrao AB, Florio L, De Aquino JP, Castaldelli-Maia JM. Pharmacological treatments for cocaine craving: what is the way forward? A systematic review. Brain Sciences. 2022;12(11):1546.

243. Jiménez-Silvestre K, Rodríguez-Kuri SE, Fernández-Cáceres C, Marín-Navarrete R. Comorbidities in patients with cocaine use disorders. Journal of Substance Use [Internet]. 2024 Dec 26 [cited 2025 Sept 30]; Available from: https://www.tandfonline.com/doi/abs/10.1080/14659891.2024.2444217

244. Palamar JJ, Davies S, Ompad DC, Cleland CM, Weitzman M. Powder cocaine and crack use in the United States: An examination of risk for arrest and socioeconomic disparities in use. Drug and Alcohol Dependence. 2015 Apr 1;149:108–16.

245. Kirchoff RW, Mohammed NM, McHugh J, Markota M, Kingsley T, Leung J, et al. Naltrexone Initiation in the Inpatient Setting for Alcohol Use Disorder: A Systematic Review of Clinical Outcomes. Mayo Clin Proc Innov Qual Outcomes. 2021 Apr 8;5(2):495–501.

