## Supplement 1 for "Efficacy and safety of pharmacological interventions for the treatment of cocaine use disorder: a systematic review and network meta-analysis"

**Supporting information S1**

### S1.1 Hierarchy for extracting outcomes with multiple data points

Where reports of studies included multiple results for the same outcome (e.g., different measurement scales, timeframes, thresholds) we implemented a standardized approach to avoid ‘double-counting’ (i.e., measures are correlated as they are measured on the same participants). Where appropriate, we performed a within-trial synthesis to average across multiple measures and avoid excluding data. For most outcomes, we implemented a hierarchy of extraction with specified preferences which are detailed blow.

Continuous abstinence at end of treatment

1. Longest period of abstinence up to and including end of treatment period
2. Longest period of abstinence at any point in the treatment period

Extent of cocaine use

*Proportion of UDS*

1. Proportion of urine samples positive for cocaine
2. Urine verified proportion of timeframe (e.g., days, weeks) used

*Quantity of use*

1. Grams of cocaine consumed over longest period of time
2. $ spent on cocaine over longest period of time

*Frequency of use*

3. Self-reported days of use over longest period of time

Self-reported instances/times used over longest period of timeCraving

We performed within-study synthesis toaverage craving outcome measures (where they had been assessed using a validated scale). Details are reported in S1.8.

1. AdherenceDirectly observed ingestion
2. Biochemical confirmation (e.g., riboflavin in urine)
3. MEMs (electronic bottles) or equivalent
4. Pill counts
5. Self-reported adherence

### S1.2 Literature search strategy

Ovid MEDLINE(R) ALL <1946 to February 19, 2025>

1	[Population]

2	Cocaine-Related Disorders/	9221

3	Cocaine Smoking/	26

4	*Cocaine/ or Cocaine/dt	19377

5	Crack Cocaine/	1518

6	cocain*.ti,kf. or crackcocain*.mp.	25063

7	(cocain* adj3 (abus* or addict* or depend* or misus* or overus* or problem* use* or use* disorder* or craving* or withdraw* or abstin*)).ab.	10009

8	(cocain* adj3 use* adj3 (aberran* or frequent* or habitual* or heavy or high or regular*)).tw,kf.	516

9	(((substance* or stimulant* or psychostimulant*) adj3 disorder*) and cocain*).mp.	11795

10	or/2-9	36923

11	[Study Design]

12	exp Randomized Controlled Trial/	633662

13	randomized controlled trial.pt.	631912

14	(randomi#ed or randomi#ation or randomi#ing).tw,kf.	913301

15	(RCT or "at random" or (random* adj3 (administ* or allocat* or assign* or class* or cluster or crossover or cross-over or control* or determine* or divide* or division or distribut* or expose* or fashion or number* or place* or pragmatic or quasi or recruit* or split or substitut* or treat*))).tw,kf.	815470

16	Random Allocation/	108125

17	randomly.ab.	453329

18	double-blind method/ or single-blind method/	216190

19	((single or double or triple or treble) adj2 (blind* or mask* or dummy)).tw,kf.	213259

20	trial.ti.	329171

21	placebo.ab.	256069

22	or/12-21	1670911

23	exp animal/ not humans/	5308102

24	((animal model* or mouse or mice or murine* or rat or rats or rodent* or muridae or murids or rabbit* or leporine* or leporidae or guineapig* or cavies or caviidae or hamster* or cricetidae or gerbil* or gerbillinae) not human*).ti.	1720023

25	or/23-24	5579292

26	22 not 25	1485320

27	10 and 26	2299

28	[Intervention: Drug Therapy]

29	Drug Therapy/	31187

30	drug therapy.fs.	2782507

31	((pharma* or psychopharma* or drug) and (therap* or treat* or (relapse* and prevent*) or maintenance)).ti.	89073

32	((pharma* or psychopharma* or drug?) adj5 treat* adj5 addiction).tw,kf.	1853

33	((prescrip* or prescrib*) adj5 stimulant*).tw,kf.	1129

34	Central Nervous System Stimulants/tu [Therapeutic Use]	6617

35	Dopamine Uptake Inhibitors/tu	740

36	((dopamin* adj (uptake or reuptake) adj inhibitor*) and (therap* or pharmacotherap* or treat* or prevent*)).mp.	3679

37	methylphenidate/ or dexmethylphenidate hydrochloride/	8054

38	(methylphenidat* or methyl-phenidat* or dexmethylphenidat* or dexmethyl-phenidat* or dex-methylphenidat* or dex-methyl-phenidat*).tw,kf.	8568

39	((amphetamine or amfetamine) adj salts).tw,kf.	213

40	dextroamphetamine/ or lisdexamfetamine dimesylate/	7233

41	(dextroamphetamin* or dextro-amphetamin* or dextroamfetamin* or dextro-amfetamin* or dexamphetamin* or dexamfetamin* or "d amphetamin*" or "d amfetamin*").tw,kf.	6186

42	(lisdexamfetamin* or lis-dexamfetamin* or lisdexamphetamin* or lis-dexamphetamin*).tw,kf.	596

43	((oral or therapeutic or "treat* with" or "to treat" or effectiveness) adj3 (methamphetamin* or metamfetamin* or methylamphetamin* or methyl-amphetamin*)).tw,kf.	426

44	(bupropion* or amfebutamon* or mazindol* or nomifensin* or modafinil*).mp.	10564

45	(nootropic* or psychotropic*).mp.	43585

46	exp "serotonin and noradrenaline reuptake inhibitors"/ or exp selective serotonin reuptake inhibitors/	50804

47	((serotonin* or noradrenalin* or nor-adrenalin* or norepinephrin* or nor-epinephrin*) adj (uptake or reuptake) adj inhibitor*).mp.	31760

48	exp Antidepressive Agents/	165739

49	exp Monoamine Oxidase Inhibitors/ or monoamine oxidase inhibit*.tw,kf.	24598

50	(antidepress* or anti depress* or MAOI* or NDRI* or NRI* or SSRI* or SNRI* or TCA* or TeCA*).tw,kf.	118925

51	(Agomelatin* or Alaproclat* or Amoxapin* or Amineptin* or Amitriptylin* or Amitriptylinoxid* or Atomoxetin* or Befloxaton* or Benactyzin* or Binospiron* or Brofaromin* or (Bupropion* or Amfebutamon*) or Butriptylin* or Caroxazon* or Cianopramin* or Cilobamin* or Cimoxaton* or Citalopram* or (Chlorimipramin* or Clomipramin* or Chlomipramin* or Clomipramine) or Clorgylin* or Clovoxamin* or Demexiptilin* or Deprenyl* or (Desipramin* or Pertofran*) or Desvenlafaxin* or Dibenzepin* or Diclofensin* or Dimetacrin* or Dosulepin* or Dothiepin* or Doxepin* or Duloxetin* or Desvenlafaxin* or Edivoxetin* or Escitalopram* or Etoperidon* or Femoxetin* or Fluotracen* or Fluoxetin* or Fluvoxamin* or (Hyperforin* or Hypericum* or St John*) or Imipramin* or Iprindol* or Iproniazid* or Ipsapiron* or Isocarboxazid* or Levomilnacipran* or Lofepramin* or Lorpiprazol* or Maprotilin* or Melitracen* or Mepiprazol* or Metapramin* or Mianserin* or Milnacipran* or Minaprin* or Mirtazapin* or Moclobemid* or Nefazodon* or Nialamid* or Nitroxazepin* or Nomifensin* or Norfenfluramin* or Nortriptylin* or Noxiptilin* or Opipramol* or Oxaflozan* or Paroxetin* or Phenelzin* or Pheniprazin* or Pipofezin* or Pirlindol* or Pivagabin* or Pizotylin* or Propizepin* or Protriptylin* or Quinupramin* or Reboxetin* or Ritanserin* or Rolipram* or Scopolamin* or Selegilin* or Sertralin* or Setiptilin* or Teciptilin* or Thozalinon* or Tianeptin* or Toloxaton* or Tranylcypromin* or Trazodon* or Trimipramin* or (Tryptophan not depletion) or Tyrima or Venlafaxin* or Viloxazin* or Vilazodon* or Vortioxetin* or Viqualin* or Zimelidin*).mp.	188783

52	exp Anticonvulsants/	158554

53	(anticonvulsant* or anti-convulsant* or anticonvulsiv* or anti-convulsiv* or antiepileptic* or anti-epileptic* or acetazolamid* or bromides or carbamazepin* or chlormethiazol* or clomethiazol* or clobazam* or clonazepam* or clorazepat* or diazepam* or dimethadion* or estazolam* or ethosuximid* or felbamat* or fenfluramin* or flunarizin* or gabapentin* or lacosamid* or lamotrigin* or levetiracetam* or lorcaserin* or lorazepam* or magnesium sulfate or magnesium sulphate or medazepam* or mephenytoin* or mephobarbital* or meprobamate* or midazolam* or nitrazepam* or oxcarbazepin* or paraldehyd* or phenobarb* or phenytoin* or pregabalin* or primidon* or riluzol* or thiopental* or tiagabin* or tiletamin* or topiramat* or trimethadion* or valproic acid or valproat* or divalproex* or vigabatrin* or zonisamid*).mp.	241511

54	(CBD or cannabinoid* or cannabidiol* or cannabinol* or dronabinol*).mp.	45828

55	exp Antipsychotic Agents/	131512

56	(antipsychotics or anti-psychotics or neuroleptics or ((antipsychotic* or anti-psychotic* or neuroleptic*) adj3 (agent* or drug? or pharma* or medication*))).tw,kf.	53087

57	(chlorpromazin* or chlorprothixen* or clopenthixol* or droperidol* or flupenthixol* or fluphenazin* or fluspirilen* or haloperidol* or loxapin* or mesoridazin* or methotrimeprazin* or molindon* or ondansetron* or penfluridol* or perazin* or perphenazin* or pimozid* or prochlorperazin* or promazin* or reserpin* or spiperon* or thioridazin* or thiothixen* or trifluoperazin* or triflupromazin*).mp.	87558

58	(atypical-antipsychotic* or amisulprid* or aripiprazol* or asenapin* or brexpiprazol* or cariprazin* or clozapin* or iloperidon* or lumateperon* or lurasidon* or olanzapin* or quetiapin* or paliperidon* or pimavanserin* or prosulprid* or quetiapin* or risperidon* or sertindol* or sulpirid* or ziprasidon* or zotepin*).mp.	49768

59	exp Lithium Compounds/ or lithium.tw,kf.	73680

60	(("5 hydroxytryptamin*" or "5 HT*") adj3 (antagonist* or block* or reversal agent*)).tw,kf.	15754

61	(("5 hydroxytryptamin*" or "5 HT*") adj3 agonist*).tw,kf. or lorcaserin*.mp.	11880

62	exp Anti-Anxiety Agents/	72557

63	(anxiolytic* or ((anti anxiety or antianxiety) adj1 (agent* or drug? or pharma* or medication*))).tw,kf.	19240

64	exp Benzodiazepines/	71462

65	(benzo* or adinazolam* or alprazolam* or bentazepam* or bretazenil* or bromazepam* or brotizolam* or camazepam* or chldiazepoxid* or cinolazepam* or clazepat* or clobazam* or clonazepam* or clotiazepam* or cloxazolam* or delorazepam* or devazepid* or diazepam* or estazolam* or ethyl loflazepat* or etizolam* or fludiazepam* or flumazenil* or flunitrazepam* or flurazepam* or flutoprazepam* or halazepam* or haloxazolam* or ketazolam* or loflazepat* or loprazolam* or lorazepam* or lormetazepam* or medazepam* or metaclazepam* or mexazolam* or midazolam* or nimetazepam* or nitrazepam* or nordazepam* or oxazepam* or oxazolam* or phenazepam* or pinazepam* or potassium clorazepat* or prazepam* or premazepam* or propazepam* or quazepam* or ripazepam* or serazepin* or temazepam* or tetrazepam* or tofisopam* or triazolam*).mp.	312262

66	(azapiron* or aeptapiron* or alnespiron* or binospiron* or buspiron* or enilospiron* or eptapiron* or gepiron* or ipsapiron* or lesopitron* or revospiron* or tandospiron* or zalospiron* or hydroxyzin*).mp.	5942

67	exp "Hypnotics and Sedatives"/	133764

68	(hypnotic* or nonbenzo* or non benzo* or zolpidem* or zaleplon* or zopiclon* or eszopiclon* or z drug*).mp.	46237

69	exp Receptors, Opioid/ag [Agonists]	5925

70	Analgesics, Opioid/tu [Therapeutic Use]	26772

71	((narcotic* or opioid* or muopioid*) adj3 agonist*).tw,kf.	11550

72	Methadone/tu	8106

73	(buprenorphin* or nalbuphin* or (methadon* adj3 (maintenance or treat*))).mp.	19047

74	exp Narcotic Antagonists/	43035

75	((narcotic* or opioid* or muopioid*) adj3 (antagonist* or block* or reversal agent*)).tw,kf.	12694

76	(naltrexon* or naloxon* or nalmefen*).mp.	40015

77	exp dopamine agonists/tu	9605

78	(dopamin* adj3 agonist*).tw,kf.	15250

79	(amantadin* or apomorphin* or aripiprazol* or bromocriptin* or cabergolin* or dihydroergocornin* or dihydroergocryptin* or dihydroergotamin* or dihydroergotoxin* or fenoldopam* or hydergin* or levodopa* or l-dopa* or carbidopa* or lisurid* or memantin* or metergolin* or pergolid* or piribedil* or pramipexol* or quinpirol*).mp.	76938

80	exp Adrenergic alpha-Agonists/tu or exp *Adrenergic alpha-Agonists/	89116

81	(alpha adj3 (adrenergic or adrenoceptor) adj3 (agent* or agonist* or stimula*)).tw,kf.	10729

82	(brimonidin* or clonidin* or dexmedetomidin* or guanabenz* or guanfacin* or medetomidin* or methyldopa* or rilmenidine* or xylazin*).mp.	41684

83	exp adrenergic alpha-antagonists/	54330

84	(alpha* adj3 adrenergic adj3 (antagonist* or block* or reversal agent*)).mp.	22036

85	(propranolol or yohimbin*).tw,id.	41252

86	nicotinic agonists/ or varenicline/ or nicotine/tu	9724

87	((nicotin* adj3 agonist*) or vareniclin*).tw,kf.	5172

88	(nicotin* adj3 (drug or treat* or therap* or medication*)).mp.	12241

89	exp Nicotinic Antagonists/	26177

90	((nicotin* adj3 (antagonist* or block* or reversal agent*)) or mecamylamin*).tw,kf.	6059

91	(acetaldehyde dehydrogenase inhibitor* or disulfiram*).mp.	5150

92	GABA Modulators/tu [Therapeutic Use]	624

93	exp GABA Uptake Inhibitors/	645

94	(((GABA or gabaergic or gaba-ergic or gamma aminobutyric acid) adj5 (uptake or reuptake) adj inhibitor*) or modulator*).mp.	141491

95	(gamma-aminobutyric acid or glutaminergic*).mp.	57990

96	Acetylcysteine/ or (N-acetylcystein* or NAC).tw,kf.	42101

97	Cycloserine.hw. or d-cycloserin*.tw,kf.	3073

98	((N-methyl-D-aspartate or NMDA) adj3 (antagonist* or block* or reversal agent*)).mp.	19793

99	acamprosate.mp.	1050

100	(ketamin* or esketamin*).mp.	26740

101	exp neuroprotective agents/ or exp neuropeptides/ or (orexin* or hypocretin*).mp.	450739

102	exp Pituitary Hormones/ or oxytocin*.mp.	302079

103	exp Gonadal Steroid Hormones/ or (dehydroepiandrosteron* or DHEA or prasteron* or progesteron*).mp.	298913

104	[Other]	0

105	(amlodipin* or baclofen* or biperiden* or citicolin* or doxazosin* or isradipin* or nimodipin* or carvedilol or celecoxib or ecopipam or chlorophenylpiperazine or metachlorophenylpiperazine or m-CPP or lidocain* or magnesium* or metyrapon* or pioglitazon* or RBP-8000 or succinylnorcocaine or TV-1380 or AlbuBChE).mp.	211610

106	(thiazolidinedion* or pioglitazon* or rosiglitazon* or troglitazon*).mp.	20010

107	(cocaine adj3 (vaccin* or antibod*)).mp.	201

108	or/29-107	4885571

109	27 and 108	1358

110	Cocaine-Related Disorders/dt [Drug Therapy]	1077

111	Substance-Related Disorders/dt [Drug Therapy]	3282

112	(cocain* or crackcocain*).mp.	48827

113	111 and 112	583

114	((pharma* or psychopharma* or psycho-pharma* or drug) adj5 (therap* or treat* or prevent* or maintenance) adj5 (cocain* or crackcocain*)).mp.	614

115	110 or 113 or 114	2060

116	27 and 115	484

117	109 or 116	1376

118	limit 117 to english language	1357

119	("1319961" or "1330470" or "1444728" or "1862788" or "2492422" or "2672061" or "7555620" or "7555621" or "7555622" or "7635993" or "7734469" or "7734470" or "8556965" or "8556974" or "8612392" or "8745134" or "8793307" or "8809502" or "8912799" or "9097872" or "9246023" or "9283506" or "9359980" or "9408812" or "9768541" or "10102764" or "10723850" or "10831016" or "10862808" or "10940543" or "11097979" or "11137283" or "11286433" or "11418225" or "11593078" or "11906804" or "12028741" or "12062456" or "12233989" or "12703668" or "12746087" or "12757964" or "12757969" or "12873248" or "14616189" or "14728105" or "14749690" or "14993114" or "15039761" or "15122957" or "15283944" or "15525998" or "15624548" or "15677600" or "15730346" or "15730347" or "15730348" or "15730349" or "15730350" or "15730352" or "15764421" or "15780550" or "15784063" or "15913920" or "16040376" or "16051446" or "16169160" or "16461866" or "16631323" or "16687365" or "16687365" or "16697124" or "16730924" or "16930857" or "16930863" or "17127551" or "17134849" or "17174102" or "17628352" or "17629631" or "17924779" or "18079068" or "18164144" or "18294021" or "18551884" or "19058926" or "19101098" or "19219665" or "19414226" or "19560290" or "19651710" or "19805702" or "19874153" or "20537812" or "20545388" or "20716302" or "20828943" or "21112155" or "21266301" or "21707811" or "21769048" or "21925806" or "22221171" or "22236504" or "22377391" or "22695473" or "22795453" or "22906516" or "22969732" or "23200303" or "23277248" or "23306096" or "23567610" or "23575810" or "23810644" or "23952889" or "24132249" or "24313244" or "24424425" or "24462581" or "24525654" or "24793366" or "24809448" or "24814607" or "24911028" or "24974353" or "25251201" or "25682480" or "25887096" or "26116930" or "26320827" or "26501788" or "26777774" or "26817621" or "26892922" or "26948856" or "27015909" or "27046312" or "27394932" or "27646197" or "28017186" or "28498501" or "31368770" or "31753736" or "32480252" or "33464660" or "34265007" or "34265007" or "34265007").ui.	147

120	118 and 119	147 [Recapture of Known Studies]

*************************************************

Ovid APA PsycInfo <1806 to February 2025 Week 2>

1	[Population]	0

2	Cocaine/ and (Drug Abuse/ or Drug Addiction/ or Drug Dependency/ or Substance Abuse Addiction.cc. or Drug Abstinence/ or Drug Withdrawal/ or Craving/)	7092

3	crack cocaine/	709

4	cocain*.ti,id. or crackcocain*.tw,id.	15189

5	(cocain* adj3 (abus* or addict* or depend* or misus* or overus* or problem* use* or use* disorder* or craving* or withdraw* or abstin*)).ab.	7407

6	(cocain* adj3 use* adj3 (aberran* or frequent* or habitual* or heavy or high or regular*)).tw,id.	352

7	(((substance* or stimulant* or psychostimulant*) adj3 disorder*) and cocain*).tw,id,hw.	2338

8	or/2-7	18003

9	[Study Design]	0

10	(clinical trial and quantitative study).md.	39368

11	randomized clinical trials/	602

12	(randomi#ed or randomi#ation or randomi#ing).tw,id.	121645

13	(RCT or "at random" or (random* adj3 (administ* or allocat* or assign* or class* or cluster or crossover or cross-over or control* or determine* or divide* or division or distribut* or expose* or fashion or number* or place* or pragmatic or quasi or recruit* or split or substitut* or treat*))).tw,id.	141387

14	((single or double or triple or treble) adj2 (blind* or mask* or dummy)).tw,id.	30738

15	trial.ti.	42671

16	placebo*.tw,id,hw.	46363

17	or/10-16	227037

18	((animal model* or mouse or mice or murine* or rat or rats or rodent* or muridae or murids or rabbit* or leporine* or leporidae or guineapig* or cavies or caviidae or hamster* or cricetidae or gerbil* or gerbillinae) not human*).ti.	141451

19	17 not 18	223916

20	8 and 19	1430

21	[Intervention]	0

22	Drug Therapy/	155237

23	((pharma* or psychopharma* or drug) and (therap* or treat* or (relapse* and prevent*) or maintenance)).ti.	12578

24	((pharma* or psychopharma* or drug?) adj5 treat* adj5 addiction).tw,id.	1734

25	((prescrip* or prescrib*) adj5 stimulant*).tw,id.	951

26	exp cns stimulating drugs/	26812

27	(dopamin* adj (uptake or reuptake) adj inhibitor*).tw,id.	305

28	(methylphenidat* or methyl-phenidat* or dexmethylphenidat* or dexmethyl-phenidat* or dex-methylphenidat* or dex-methyl-phenidat*).tw,id,hw.	5993

29	((amphetamine or amfetamine) adj salts).tw,id.	146

30	(dextroamphetamin* or dextro-amphetamin* or dextroamfetamin* or dextro-amfetamin* or dexamphetamin* or dexamfetamin* or "d amphetamin*" or "d amfetamin*").tw,id,hw.	4355

31	(lisdexamfetamin* or lis-dexamfetamin* or lisdexamphetamin* or lis-dexamphetamin*).tw,id,hw.	315

32	((oral or therapeutic or "treat* with" or "to treat" or effectiveness) adj3 (methamphetamin* or metamfetamin* or methylamphetamin* or methyl-amphetamin*)).tw,id.	188

33	(bupropion* or amfebutamon* or mazindol* or nomifensin* or modafinil*).tw,id,hw.	3950

34	nootropic*.tw,id,hw.	1337

35	exp psychotropic drugs/ or psychotropic*.tw,id.	110598

36	exp serotonin norepinephrine reuptake inhibitors/ or exp serotonin reuptake inhibitors/	16017

37	((serotonin* or noradrenalin* or nor-adrenalin* or norepinephrin* or nor-epinephrin*) adj (uptake or reuptake) adj inhibitor*).tw,id.	11384

38	exp antidepressant drugs/	43201

39	exp monoamine oxidase inhibitors/ or monoamine oxidase inhibit*.tw,id.	3032

40	(antidepress* or anti depress* or MAOI* or NDRI* or NRI* or SSRI* or SNRI* or TCA* or TeCA*).tw,id.	52754

41	exp anticonvulsive drugs/	13200

42	(Agomelatin* or Alaproclat* or Amoxapin* or Amineptin* or Amitriptylin* or Amitriptylinoxid* or Atomoxetin* or Befloxaton* or Benactyzin* or Binospiron* or Brofaromin* or (Bupropion* or Amfebutamon*) or Butriptylin* or Caroxazon* or Cianopramin* or Cilobamin* or Cimoxaton* or Citalopram* or (Chlorimipramin* or Clomipramin* or Chlomipramin* or Clomipramine) or Clorgylin* or Clovoxamin* or Demexiptilin* or Deprenyl* or (Desipramin* or Pertofran*) or Desvenlafaxin* or Dibenzepin* or Diclofensin* or Dimetacrin* or Dosulepin* or Dothiepin* or Doxepin* or Duloxetin* or Desvenlafaxin* or Edivoxetin* or Escitalopram* or Etoperidon* or Femoxetin* or Fluotracen* or Fluoxetin* or Fluvoxamin* or (Hyperforin* or Hypericum* or St John*) or Imipramin* or Iprindol* or Iproniazid* or Ipsapiron* or Isocarboxazid* or Levomilnacipran* or Lofepramin* or Lorpiprazol* or Maprotilin* or Melitracen* or Mepiprazol* or Metapramin* or Mianserin* or Milnacipran* or Minaprin* or Mirtazapin* or Moclobemid* or Nefazodon* or Nialamid* or Nitroxazepin* or Nomifensin* or Norfenfluramin* or Nortriptylin* or Noxiptilin* or Opipramol* or Oxaflozan* or Paroxetin* or Phenelzin* or Pheniprazin* or Pipofezin* or Pirlindol* or Pivagabin* or Pizotylin* or Propizepin* or Protriptylin* or Quinupramin* or Reboxetin* or Ritanserin* or Rolipram* or Scopolamin* or Selegilin* or Sertralin* or Setiptilin* or Teciptilin* or Thozalinon* or Tianeptin* or Toloxaton* or Tranylcypromin* or Trazodon* or Trimipramin* or (Tryptophan not depletion) or Tyrima or Venlafaxin* or Viloxazin* or Vilazodon* or Vortioxetin* or Viqualin* or Zimelidin*).tw,id,hw.	43510

43	exp anticonvulsive drugs/	13200

44	(anticonvulsant* or anti-convulsant* or anticonvulsiv* or anti-convulsiv* or antiepileptic* or anti-epileptic* or acetazolamid* or bromides or carbamazepin* or chlormethiazol* or clomethiazol* or clobazam* or clonazepam* or clorazepat* or diazepam* or dimethadion* or estazolam* or ethosuximid* or felbamat* or fenfluramin* or flunarizin* or gabapentin* or lacosamid* or lamotrigin* or levetiracetam* or lorcaserin* or lorazepam* or magnesium sulfate or magnesium sulphate or medazepam* or mephenytoin* or mephobarbital* or meprobamate* or midazolam* or nitrazepam* or oxcarbazepin* or paraldehyd* or phenobarb* or phenytoin* or pregabalin* or primidon* or riluzol* or thiopental* or tiagabin* or tiletamin* or topiramat* or trimethadion* or valproic acid or valproat* or divalproex* or vigabatrin* or zonisamid*).tw,id,hw.	35146

45	exp cannabinoids/	7297

46	(CBD or cannabinoid* or cannabidiol* or cannabinol* or dronabinol*).tw,id.	8093

47	exp neuroleptic drugs/	35812

48	(antipsychotics or anti-psychotics or neuroleptics or ((antipsychotic* or anti-psychotic* or neuroleptic*) adj3 (agent* or drug? or pharma* or medication*))).tw,id.	36755

49	(chlorpromazin* or chlorprothixen* or clopenthixol* or droperidol* or flupenthixol* or fluphenazin* or fluspirilen* or haloperidol* or loxapin* or mesoridazin* or methotrimeprazin* or molindon* or ondansetron* or penfluridol* or perazin* or perphenazin* or pimozid* or prochlorperazin* or promazin* or reserpin* or spiperon* or thioridazin* or thiothixen* or trifluoperazin* or triflupromazin*).tw,id,hw.	17852

50	(atypical-antipsychotic* or amisulprid* or aripiprazol* or asenapin* or brexpiprazol* or cariprazin* or clozapin* or iloperidon* or lumateperon* or lurasidon* or olanzapin* or quetiapin* or paliperidon* or pimavanserin* or prosulprid* or quetiapin* or risperidon* or sertindol* or sulpirid* or ziprasidon* or zotepin*).tw,id,hw.	28195

51	exp lithium/ or lithium.tw,id.	12149

52	(("5 hydroxytryptamin*" or "5 HT*") adj3 (antagonist* or block* or reversal agent*)).tw,id.	4467

53	(("5 hydroxytryptamin*" or "5 HT*") adj3 agonist*).tw,id. or lorcaserin*.tw,id,hw.	4153

54	exp anxiolytic drugs/	50864

55	(anxiolytic* or ((anti anxiety or antianxiety) adj1 (agent* or drug? or pharma* or medication*))).tw,id.	9473

56	exp Benzodiazepines/	11737

57	(benzo* or adinazolam* or alprazolam* or bentazepam* or bretazenil* or bromazepam* or brotizolam* or camazepam* or chldiazepoxid* or cinolazepam* or clazepat* or clobazam* or clonazepam* or clotiazepam* or cloxazolam* or delorazepam* or devazepid* or diazepam* or estazolam* or ethyl loflazepat* or etizolam* or fludiazepam* or flumazenil* or flunitrazepam* or flurazepam* or flutoprazepam* or halazepam* or haloxazolam* or ketazolam* or loflazepat* or loprazolam* or lorazepam* or lormetazepam* or medazepam* or metaclazepam* or mexazolam* or midazolam* or nimetazepam* or nitrazepam* or nordazepam* or oxazepam* or oxazolam* or phenazepam* or pinazepam* or potassium clorazepat* or prazepam* or premazepam* or propazepam* or quazepam* or ripazepam* or serazepin* or temazepam* or tetrazepam* or tofisopam* or triazolam*).tw,id,hw.	24714

58	(azapiron* or aeptapiron* or alnespiron* or binospiron* or buspiron* or enilospiron* or eptapiron* or gepiron* or ipsapiron* or lesopitron* or revospiron* or tandospiron* or zalospiron* or hydroxyzin*).tw,id,hw.	2080

59	exp hypnotic drugs/	6092

60	(hypnotic* or nonbenzo* or non benzo* or zolpidem* or zaleplon* or zopiclon* or eszopiclon* or z drug*).tw,id,hw.	14245

61	exp narcotic agonists/	1931

62	((narcotic* or opioid* or muopioid*) adj3 agonist*).tw,id.	3459

63	Opioid Analgesics/	1013

64	methadone/ or methadone maintenance/	5912

65	(buprenorphin* or nalbuphin* or (methadon* adj3 (maintenance or treat*))).tw,id,hw.	9552

66	exp narcotic antagonists/	6707

67	((narcotic* or opioid* or muopioid*) adj3 (antagonist* or block* or reversal agent*)).tw,id.	3493

68	(naltrexon* or naloxon* or nalmefen*).tw,id.	10322

69	exp dopamine agonists/	26502

70	(dopamin* adj3 agonist*).tw,id.	4768

71	(amantadin* or apomorphin* or aripiprazol* or bromocriptin* or cabergolin* or dihydroergocornin* or dihydroergocryptin* or dihydroergotamin* or dihydroergotoxin* or fenoldopam* or hydergin* or levodopa* or l-dopa* or carbidopa* or lisurid* or memantin* or metergolin* or pergolid* or piribedil* or pramipexol* or quinpirol*).tw,id,hw.	17701

72	exp adrenergic drugs/	21054

73	((adrenergic or adrenoceptor) adj3 (agent* or agonist* or stimula*)).tw,id.	1921

74	(brimonidin* or clonidin* or dexmedetomidin* or guanabenz* or guanfacin* or medetomidin* or methyldopa* or rilmenidine* or xylazin*).tw,id,hw.	3635

75	exp adrenergic blocking drugs/	4114

76	(alpha* adj3 adrenergic adj3 (antagonist* or block* or reversal agent*)).tw,id.	635

77	(propranolol or yohimbin*).tw,id.	3175

78	((nicotin* adj3 agonist*) or vareniclin*).tw,id,hw.	1712

79	((nicotin* adj3 (antagonist* or block* or reversal agent*)) or mecamylamin*).tw,id,hw.	1621

80	(nicotin* adj3 (drug or treat* or therap* or medication*)).tw,id.	4038

81	(acetaldehyde dehydrogenase inhibitor* or disulfiram*).tw,id,hw.	804

82	gamma aminobutyric acid/ or exp gamma aminobutyric acid agonists/	10650

83	(((GABA or gabaergic or gaba-ergic or gamma aminobutyric acid) adj5 (uptake or reuptake) adj inhibitor*) or modulator*).tw,id.	12666

84	(gamma-aminobutyric acid or glutaminergic*).tw,id.	7729

85	Cysteine/ or (N-acetylcystein* or NAC).tw,id.	5114

86	d-cycloserin*.tw,id.	657

87	n-methyl-d-aspartate/	9960

88	((N-methyl-D-aspartate or NMDA) adj3 (antagonist* or block* or reversal agent*)).tw,id,hw.	6456

89	exp gamma aminobutyric acid agonists/	1842

90	acamprosate.tw,id,hw.	574

91	psychedelic assisted therapy/	210

92	(ketamin* or esketamin*).tw,id,hw.	5388

93	exp neuropeptides/ or (orexin* or hypocretin*).tw,id,hw.	35706

94	exp hormones/ or (dehydroepiandrosteron* or DHEA or prasteron* or oxytocin* or progesteron*).tw,id,hw.	80148

95	[Other]	0

96	(amlodipin* or baclofen* or biperiden* or citicolin* or doxazosin* or isradipin* or lidocain* or magnesium* or nimodipin* or carvedilol or celecoxib or ecopipam or chlorophenylpiperazine or metachlorophenylpiperazine or m-CPP or metyrapon* or pioglitazon* or RBP-8000 or succinylnorcocaine or TV-1380 or AlbuBChE).tw,id,hw.	6774

97	(thiazolidinedion* or pioglitazon* or rosiglitazon* or troglitazon*).tw,id,hw.	456

98	(cocaine adj3 (vaccin* or antibod*)).tw,id.	45

99	or/22-98	442971

100	20 and 99	925

101	limit 100 to english language	903

102	("1319961" or "1330470" or "1444728" or "1862788" or "2492422" or "2672061" or "7555620" or "7555621" or "7555622" or "7635993" or "7734469" or "7734470" or "8556965" or "8556974" or "8612392" or "8745134" or "8793307" or "8809502" or "8912799" or "9097872" or "9246023" or "9283506" or "9359980" or "9408812" or "9768541" or "10102764" or "10723850" or "10831016" or "10862808" or "10940543" or "11097979" or "11137283" or "11286433" or "11418225" or "11593078" or "11906804" or "12028741" or "12062456" or "12233989" or "12703668" or "12746087" or "12757964" or "12757969" or "12873248" or "14616189" or "14728105" or "14749690" or "14993114" or "15039761" or "15122957" or "15283944" or "15525998" or "15624548" or "15677600" or "15730346" or "15730347" or "15730348" or "15730349" or "15730350" or "15730352" or "15764421" or "15780550" or "15784063" or "15913920" or "16040376" or "16051446" or "16169160" or "16461866" or "16631323" or "16687365" or "16687365" or "16697124" or "16730924" or "16930857" or "16930863" or "17127551" or "17134849" or "17174102" or "17628352" or "17629631" or "17924779" or "18079068" or "18164144" or "18294021" or "18551884" or "19058926" or "19101098" or "19219665" or "19414226" or "19560290" or "19651710" or "19805702" or "19874153" or "20537812" or "20545388" or "20716302" or "20828943" or "21112155" or "21266301" or "21707811" or "21769048" or "21925806" or "22221171" or "22236504" or "22377391" or "22695473" or "22795453" or "22906516" or "22969732" or "23200303" or "23277248" or "23306096" or "23567610" or "23575810" or "23810644" or "23952889" or "24132249" or "24313244" or "24424425" or "24462581" or "24525654" or "24793366" or "24809448" or "24814607" or "24911028" or "24974353" or "25251201" or "25682480" or "25887096" or "26116930" or "26320827" or "26501788" or "26777774" or "26817621" or "26892922" or "26948856" or "27015909" or "27046312" or "27394932" or "27646197" or "28017186" or "28498501" or "31368770" or "31753736" or "32480252" or "33464660" or "34265007" or "34265007" or "34265007").pm.	123

103	101 and 102	123 [Recapture of Known Studies]

<https://ovidsp.ovid.com/ovidweb.cgi?T=JS&NEWS=N&PAGE=main&SHAREDSEARCHID=394Eq5GRu6Qr25xStuqzuWEahnzvbdP30PTldIBwsj50qXtIDbrNcV2bKJK4LQr1X>

*************************************************

Cochrane Central Register of Controlled Trials (Cochrane Library, Issue 2 of 12, 2025)

Search Name:	B-ESG Cocaine Use Disorder

Date Run:	20/02/2025

ID	Search	Hits

#1	MeSH descriptor: [Cocaine-Related Disorders] this term only	1227

#2	MeSH descriptor: [Crack Cocaine] this term only	92

#3	(cocain* NEAR/3 (abus* or addict* or depend* or misus* or overus* or (problem* NEXT use*) or (use* NEXT disorder*) or craving* or withdraw* or abstin*)):ti,ab,kw	2240

#4	crackcocain*:ti,ab,kw	48

#5	(cocain* near/3 use* near/3 (aberran* or frequent* or habitual* or heavy or high or regular*)):ti,ab,kw	72

#6	(((substance* or stimulant* or psychostimulant*) NEAR/3 disorder*) and cocain*):ti,ab,kw	869

#7	(#1 OR #2 OR #3 OR #4 OR #5 OR #6)	2821

#8	MeSH descriptor: [Drug Therapy] this term only	480

#9	((pharma* or psychopharma* or drug) and (therap* or treat* or (relapse* and prevent*) or maintenance)):ti	12653

#10	((pharma* or psychopharma* or drug or drugs) NEAR treat* NEAR addiction):ti,ab,kw	346

#11	((prescrip* or prescrib*) NEAR stimulant*):ti,ab,kw	95

#12	MeSH descriptor: [Central Nervous System Stimulants] explode all trees and with qualifier(s): [therapeutic use - TU]	1295

#13	MeSH descriptor: [Dopamine Uptake Inhibitors] explode all trees and with qualifier(s): [therapeutic use - TU]	197

#14	((dopamin* NEXT (uptake or reuptake) NEXT inhibitor*) and (therap* or pharmacotherap* or treat* or prevent*)):ti,ab,kw	413

#15	MeSH descriptor: [Methylphenidate] explode all trees	1983

#16	(methylphenidat* or methyl-phenidat* or dexmethylphenidat* or dexmethyl-phenidat* or dex-methylphenidat* or dex-methyl-phenidat*).:ti,ab,kw	3225

#17	((amphetamine or amfetamine) NEXT salts):ti,ab,kw	110

#18	MeSH descriptor: [Dextroamphetamine] explode all trees	872

#19	(dextroamphetamin* or dextro-amphetamin* or dextroamfetamin* or dextro-amfetamin* or dexamphetamin* or dexamfetamin* or d-amphetamin* or d-amfetamin*):ti,ab,kw	1127

#20	(lisdexamfetamin* or lis-dexamfetamin* or lisdexamphetamin* or lis-dexamphetamin*):ti,ab,kw	371

#21	((oral or therapeutic or (treat* NEXT with) or "to treat" or effectiveness) NEAR/3 (methamphetamin* or metamfetamin* or methylamphetamin* or methyl-amphetamin*)):ti,ab,kw	79

#22	(bupropion* or amfebutamon* or mazindol* or nomifensin* or modafinil*):ti,ab,kw	3309

#23	(nootropic* or psychotropic*):ti,ab,kw	4234

#24	MeSH descriptor: [Neurotransmitter Uptake Inhibitors] explode all trees	4339

#25	((serotonin* or noradrenalin* or nor-adrenalin* or norepinephrin* or nor-epinephrin*) NEXT (uptake or reuptake) NEXT inhibitor*):ti,ab,kw	6232

#26	MeSH descriptor: [Antidepressive Agents] explode all trees	7542

#27	MeSH descriptor: [Monoamine Oxidase Inhibitors] explode all trees	461

#28	(monoamine NEXT oxidase NEXT inhibit*):ti,ab,kw	734

#29	(antidepress* or (anti NEXT depress*) or MAOI* or NDRI* or NRI* or SSRI* or SNRI* or TCA* or TeCA*):ti,ab,kw	22951

#30	(Agomelatin* or Alaproclat* or Amoxapin* or Amineptin* or Amitriptylin* or Amitriptylinoxid* or Atomoxetin* or Befloxaton* or Benactyzin* or Binospiron* or Brofaromin* or (Bupropion* or Amfebutamon*) or Butriptylin* or Caroxazon* or Cianopramin* or Cilobamin* or Cimoxaton* or Citalopram* or (Chlorimipramin* or Clomipramin* or Chlomipramin* or Clomipramine) or Clorgylin* or Clovoxamin* or Demexiptilin* or Deprenyl* or (Desipramin* or Pertofran*) or Desvenlafaxin* or Dibenzepin* or Diclofensin* or Dimetacrin* or Dosulepin* or Dothiepin* or Doxepin* or Duloxetin* or Desvenlafaxin* or Edivoxetin* or Escitalopram* or Etoperidon* or Femoxetin* or Fluotracen* or Fluoxetin* or Fluvoxamin* or (Hyperforin* or Hypericum* or St John*) or Imipramin* or Iprindol* or Iproniazid* or Ipsapiron* or Isocarboxazid* or Levomilnacipran* or Lofepramin* or Lorpiprazol* or Maprotilin* or Melitracen* or Mepiprazol* or Metapramin* or Mianserin* or Milnacipran* or Minaprin* or Mirtazapin* or Moclobemid* or Nefazodon* or Nialamid* or Nitroxazepin* or Nomifensin* or Norfenfluramin* or Nortriptylin* or Noxiptilin* or Opipramol* or Oxaflozan* or Paroxetin* or Phenelzin* or Pheniprazin* or Pipofezin* or Pirlindol* or Pivagabin* or Pizotylin* or Propizepin* or Protriptylin* or Quinupramin* or Reboxetin* or Ritanserin* or Rolipram* or Scopolamin* or Selegilin* or Sertralin* or Setiptilin* or Teciptilin* or Thozalinon* or Tianeptin* or Toloxaton* or Tranylcypromin* or Trazodon* or Trimipramin* or (Tryptophan not depletion) or Tyrima or Venlafaxin* or Viloxazin* or Vilazodon* or Vortioxetin* or Viqualin* or Zimelidin*):ti,ab,kw	30456

#31	MeSH descriptor: [Anticonvulsants] explode all trees	3225

#32	(anticonvulsant* or anti-convulsant* or anticonvulsiv* or anti-convulsiv* or antiepileptic* or anti-epileptic* or acetazolamid* or bromides or carbamazepin* or chlormethiazol* or clomethiazol* or clobazam* or clonazepam* or clorazepat* or diazepam* or dimethadion* or estazolam* or ethosuximid* or felbamat* or fenfluramin* or flunarizin* or gabapentin* or lacosamid* or lamotrigin* or levetiracetam* or lorcaserin* or lorazepam* or magnesium sulfate or magnesium sulphate or medazepam* or mephenytoin* or mephobarbital* or meprobamate* or midazolam* or nitrazepam* or oxcarbazepin* or paraldehyd* or phenobarb* or phenytoin* or pregabalin* or primidon* or riluzol* or thiopental* or tiagabin* or tiletamin* or topiramat* or trimethadion* or valproic acid or valproat* or divalproex* or vigabatrin* or zonisamid*):ti,ab,kw	43341

#33	MeSH descriptor: [Cannabinoids] explode all trees	1556

#34	(CBD or cannabinoid* or cannabidiol* or cannabinol* or dronabinol*):ti,ab,kw	3688

#35	MeSH descriptor: [Antipsychotic Agents] explode all trees	6135

#36	(antipsychotics or anti-psychotics or neuroleptics or ((antipsychotic* or anti-psychotic* or neuroleptic*) NEAR/3 (agent* or drug or drugs or pharma* or medication*))):ti,ab,kw	11484

#37	(chlorpromazin* or chlorprothixen* or clopenthixol* or droperidol* or flupenthixol* or fluphenazin* or fluspirilen* or haloperidol* or loxapin* or mesoridazin* or methotrimeprazin* or molindon* or ondansetron* or penfluridol* or perazin* or perphenazin* or pimozid* or prochlorperazin* or promazin* or reserpin* or spiperon* or thioridazin* or thiothixen* or trifluoperazin* or triflupromazin*).mp.	87555

58	(atypical-antipsychotic* or amisulprid* or aripiprazol* or asenapin* or brexpiprazol* or cariprazin* or clozapin* or iloperidon* or lumateperon* or lurasidon* or olanzapin* or quetiapin* or paliperidon* or pimavanserin* or prosulprid* or quetiapin* or risperidon* or sertindol* or sulpirid* or ziprasidon* or zotepin*):ti,ab,kw	0

#38	MeSH descriptor: [Lithium Compounds] explode all trees	720

#39	lithium:ti,ab,kw	3410

#40	(("5-hydroxytryptamine" or "5-HT") NEAR/3 (antagonist* or block* or (reversal NEXT agent*))):ti,ab,kw	473

#41	((("5-hydroxytryptamine" or "5-HT") NEAR/3 (agonist*)) or lorcaserin*):ti,ab,kw	444

#42	MeSH descriptor: [Anti-Anxiety Agents] explode all trees	2587

#43	(anxiolytic* or ((anti-anxiety or antianxiety) NEAR/2 (agent* or drug or drugs or pharma* or medication*))):ti,ab,kw	5261

#44	MeSH descriptor: [Benzodiazepines] explode all trees	11664

#45	(benzo* or adinazolam* or alprazolam* or bentazepam* or bretazenil* or bromazepam* or brotizolam* or camazepam* or chldiazepoxid* or cinolazepam* or clazepat* or clobazam* or clonazepam* or clotiazepam* or cloxazolam* or delorazepam* or devazepid* or diazepam* or estazolam* or ethyl loflazepat* or etizolam* or fludiazepam* or flumazenil* or flunitrazepam* or flurazepam* or flutoprazepam* or halazepam* or haloxazolam* or ketazolam* or loflazepat* or loprazolam* or lorazepam* or lormetazepam* or medazepam* or metaclazepam* or mexazolam* or midazolam* or nimetazepam* or nitrazepam* or nordazepam* or oxazepam* or oxazolam* or phenazepam* or pinazepam* or potassium clorazepat* or prazepam* or premazepam* or propazepam* or quazepam* or ripazepam* or serazepin* or temazepam* or tetrazepam* or tofisopam* or triazolam*):ti,ab,kw	30032

#46	(azapiron* or aeptapiron* or alnespiron* or binospiron* or buspiron* or enilospiron* or eptapiron* or gepiron* or ipsapiron* or lesopitron* or revospiron* or tandospiron* or zalospiron* or hydroxyzin*):ti,ab,kw	1564

#47	MeSH descriptor: [Hypnotics and Sedatives] explode all trees	4828

#48	(hypnotic* or nonbenzo* or (non NEXT benzo*) or zolpidem* or zaleplon* or zopiclon* or eszopiclon* or z-drug*):ti,ab,kw	8667

#49	MeSH descriptor: [Receptors, Opioid] explode all trees and with qualifier(s): [agonists - AG]	137

#50	MeSH descriptor: [Analgesics, Opioid] explode all trees and with qualifier(s): [therapeutic use - TU]	4546

#51	((narcotic* or opioid* or muopioid*) NEAR/3 agonist*):ti,ab,kw	1378

#52	MeSH descriptor: [Methadone] explode all trees and with qualifier(s): [therapeutic use - TU]	993

#53	(buprenorphin* or nalbuphin* or (methadon* NEAR/3 (maintenance or treat*))):ti,ab,kw	6033

#54	MeSH descriptor: [Narcotic Antagonists] explode all trees	1657

#55	((narcotic* or opioid* or muopioid*) NEAR/3 (antagonist* or block* or (reversal NEXT agent*))):ti,ab,kw	3348

#56	(naltrexon* or naloxon* or nalmefen*):ti,ab,kw	5762

#57	MeSH descriptor: [Dopamine Agonists] explode all trees and with qualifier(s): [therapeutic use - TU]	424

#58	(dopamin* NEAR/3 agonist*):ti,ab,kw	1983

#59	(amantadin* or apomorphin* or aripiprazol* or bromocriptin* or cabergolin* or dihydroergocornin* or dihydroergocryptin* or dihydroergotamin* or dihydroergotoxin* or fenoldopam* or hydergin* or levodopa* or l-dopa* or carbidopa* or lisurid* or memantin* or metergolin* or pergolid* or piribedil* or pramipexol* or quinpirol*):ti,ab,kw	11325

#60	MeSH descriptor: [Adrenergic alpha-Agonists] explode all trees	1305

#61	(alpha NEAR/3 (adrenergic or adrenoceptor) NEAR/3 (agent* or agonist* or stimula*)):ti,ab,kw	2218

#62	(brimonidin* or clonidin* or dexmedetomidin* or guanabenz* or guanfacin* or medetomidin* or methyldopa* or rilmenidine* or xylazin*):ti,ab,kw	16986

#63	MeSH descriptor: [Adrenergic alpha-Antagonists] explode all trees	1516

#64	(alpha* NEAR/3 adrenergic NEAR/3 (antagonist* or block* or reversal agent*)):ti,ab,kw	2138

#65	(propranolol or yohimbin*):ti,ab,kw	5736

#66	MeSH descriptor: [Nicotinic Agonists] explode all trees	901

#67	MeSH descriptor: [Nicotine] explode all trees	3385

#68	MeSH descriptor: [Varenicline] explode all trees	730

#69	((nicotin* near/3 agonist*) or vareniclin*):ti,ab,kw	1962

#70	(nicotin* NEAR/3 (drug or treat* or therap* or medication*)):ti,ab,kw	3706

#71	MeSH descriptor: [Nicotinic Antagonists] explode all trees	61

#72	((nicotin* NEAR/3 (antagonist* or block* or reversal agent*)) or mecamylamin*):ti,ab,kw	320

#73	((acetaldehyde NEXT dehydrogenase NEXT inhibitor*) or disulfiram*):ti,ab,kw	338

#74	MeSH descriptor: [GABA Modulators] explode all trees	278

#75	MeSH descriptor: [GABA Uptake Inhibitors] explode all trees	7

#76	((GABA or gabaergic or gaba-ergic or "gamma aminobutyric acid") NEAR ((uptake or reuptake) NEXT inhibitor*) or modulator*):ti,ab,kw	6479

#77	("gamma aminobutyric acid" or glutaminergic*):ti,ab,kw	1945

#78	MeSH descriptor: [Acetylcysteine] this term only	1448

#79	(N-acetylcystein* or NAC):ti,ab,kw	3553

#80	MeSH descriptor: [Cycloserine] this term only	298

#81	d-cycloserin*:ti,ab,kw	450

#82	((N-methyl-D-aspartate or NMDA) NEAR/3 (antagonist* or block* or reversal agent*)):ti,ab,kw	1583

#83	acamprosate:ti,ab,kw	379

#84	(ketamin* or esketamin*):ti,ab,kw	8925

#85	MeSH descriptor: [Neuroprotective Agents] explode all trees	1261

#86	(orexin* or hypocretin*):ti,ab,kw	549

#87	MeSH descriptor: [Hormones] explode all trees	78166

#88	(dehydroepiandrosteron* or DHEA or prasteron* or oxytocin* or progesteron*):ti,ab,kw	17386

#89	(amlodipin* or baclofen* or biperiden* or citicolin* or doxazosin* or isradipin* or lidocain* or magnesium* or nimodipin* or carvedilol or celecoxib or ecopipam or chlorophenylpiperazine or metachlorophenylpiperazine or m-CPP or metyrapon* or pioglitazon* or RBP-8000 or succinylnorcocaine or TV-1380 or AlbuBChE):ti,ab,kw	41038

#90	(thiazolidinedion* or pioglitazon* or rosiglitazon* or troglitazon*):ti,ab,kw	4326

#91	(cocaine NEAR/3 (vaccin* or antibod*)):ti,ab,kw	16

#92	(#8 OR #9 OR #10 OR #11 OR #12 OR #13 OR #14 OR #15 OR #16 OR #17 OR #18 OR #19 OR #20 OR #21 OR #22 OR #23 OR #24 OR #25 OR #26 OR #27 OR #28 OR #29 OR #30 OR #31 OR #32 OR #33 OR #34 OR #35 OR #36 OR #37 OR #38 OR #39 OR #40 OR #41 OR #42 OR #43 OR #44 OR #45 OR #46 OR #47 OR #48 OR #49 OR #50 OR #51 OR #52 OR #53 OR #54 OR #55 OR #56 OR #57 OR #58 OR #59 OR #60 OR #61 OR #62 OR #63 OR #64 OR #65 OR #66 OR #67 OR #68 OR #69 OR #70 OR #71 OR #72 OR #73 OR #74 OR #75 OR #76 OR #77 OR #78 OR #79 OR #80 OR #81 OR #82 OR #83 OR #84 OR #85 OR #86 OR #87 OR #88 OR #89 OR #90 OR #91)	308827

#93	(#7 AND #92)	1687

#94	conference proceeding:pt	253565

#95	#93 NOT #94	1572

[74 addn conference abstracts removed in EndNote)

*************************************************

Ovid Embase (precise search)<1974 to 2025 February 19>

1	[Population]	0

2	cocaine dependence/	14842

3	cocaine/ and (drug abuse/ or substance abuse/ or drug dependence/ or drug craving/ or drug withdrawal/ or withdrawal syndrome/)	22285

4	(cocain* adj3 (abus* or addict* or depend* or misus* or overus* or problem* use* or use* disorder* or craving* or withdraw* or abstin*)).tw,kf.	14522

5	(cocain* adj3 use* adj3 (aberran* or frequent* or habitual* or heavy or high or regular*)).tw,kf.	687

6	Crack Cocaine.mp.	2164

7	((substance* or stimulant* or psychostimulant*) adj3 disorder*).ti,kf,hw. and cocain*.mp.	2681

8	or/2-7	38971

9	[Study Design]	0

10	randomized controlled trial/	865110

11	randomization/	100166

12	(randomi#ed or randomi#ation or randomi#ing).tw,kf.	1300346

13	(RCT or "at random" or (random* adj3 (administ* or allocat* or assign* or class* or cluster or crossover or cross-over or control* or determine* or divide* or division or distribut* or expose* or fashion or number* or place* or pragmatic or quasi or recruit* or split or substitut* or treat*))).tw,kf.	1106399

14	single blind procedure/ or double blind procedure/ or triple blind procedure/	284072

15	((single or double or triple or treble) adj2 (blind* or mask* or dummy)).tw,kf.	300864

16	trial.ti. or control* trial.ab,kf.	562749

17	((placebo adj5 (control* or group* or trial or versus or vs)) or "or placebo").tw,kf.	291829

18	(drug therapy and placebo).mp.	364535

19	or/10-18	2183556

20	(animal.hw. or nonhuman.sh.) not human.sh.	7167974

21	((animal model* or mouse or mice or murine* or rat or rats or rodent* or muridae or murids or rabbit* or leporine* or leporidae or guineapig* or cavies or caviidae or hamster* or cricetidae or gerbil* or gerbillinae) not human*).ti.	1890640

22	20 or 21	7475968

23	19 not 22	1966059

24	8 and 23	2950

25	[Intervention/Embase Section Heading]	0

26	"Drug literature ".ec.	8340765

27	24 and 26	1663

28	(cocain* or crackcocain*).ti.	26999

29	27 and 28	786

30	[Included - studies captured from reviews]	0

31	("1319961" or "1330470" or "1444728" or "1862788" or "2492422" or "2672061" or "7555620" or "7555621" or "7555622" or "7635993" or "7734469" or "7734470" or "8556965" or "8556974" or "8612392" or "8745134" or "8793307" or "8809502" or "8912799" or "9097872" or "9246023" or "9283506" or "9359980" or "9408812" or "9768541" or "10102764" or "10723850" or "10831016" or "10862808" or "10940543" or "11097979" or "11137283" or "11286433" or "11418225" or "11593078" or "11906804" or "12028741" or "12062456" or "12233989" or "12703668" or "12746087" or "12757964" or "12757969" or "12873248" or "14616189" or "14728105" or "14749690" or "14993114" or "15039761" or "15122957" or "15283944" or "15525998" or "15624548" or "15677600" or "15730346" or "15730347" or "15730348" or "15730349" or "15730350" or "15730352" or "15764421" or "15780550" or "15784063" or "15913920" or "16040376" or "16051446" or "16169160" or "16461866" or "16631323" or "16687365" or "16687365" or "16697124" or "16730924" or "16930857" or "16930863" or "17127551" or "17134849" or "17174102" or "17628352" or "17629631" or "17924779" or "18079068" or "18164144" or "18294021" or "18551884" or "19058926" or "19101098" or "19219665" or "19414226" or "19560290" or "19651710" or "19805702" or "19874153" or "20537812" or "20545388" or "20716302" or "20828943" or "21112155" or "21266301" or "21707811" or "21769048" or "21925806" or "22221171" or "22236504" or "22377391" or "22695473" or "22795453" or "22906516" or "22969732" or "23200303" or "23277248" or "23306096" or "23567610" or "23575810" or "23810644" or "23952889" or "24132249" or "24313244" or "24424425" or "24462581" or "24525654" or "24793366" or "24809448" or "24814607" or "24911028" or "24974353" or "25251201" or "25682480" or "25887096" or "26116930" or "26320827" or "26501788" or "26777774" or "26817621" or "26892922" or "26948856" or "27015909" or "27046312" or "27394932" or "27646197" or "28017186" or "28498501" or "31368770" or "31753736" or "32480252" or "33464660" or "34265007" or "34265007" or "34265007").pm.	142

32	[Embase not MEDLINE records]

33	limit 31 to embase status	138

34	29 and 33	138 [Recapture of Known Studies]

*************************************************

Noel-Storr AH, Dooley G, Wisniewski S, Glanville J, Thomas J, Cox S, Featherstone R, Foxlee R. Cochrane Centralised Search Service showed high sensitivity identifying randomized controlled trials: A retrospective analysis. J Clin Epidemiol. 2020 Nov;127:142-150. doi: 10.1016/j.jclinepi.2020.08.008. Epub 2020 Aug 13. PMID: 32798713.

*************************************************

### S1.3 Additional analysis details, model parameters, and sensitivity analyses details

Non-informative prior distributions were initially applied for the intercept and intervention effects (normal distribution with mean = 0 and SD = 10), and for between-study heterogeneity SD (half-normal distribution with SD = 5), on the log-OR and SMD scales. Each model was run with four chains of 5000 warm-up (burn-in) and 5000 post-warm-up iterations. Convergence was assessed using R-hat rank-based diagnostics (comparing the between- and within-chain estimates for model parameters), with values close to 1 and <1.1 considered acceptable. In cases of numerical instability, convergence, mixing, or sampling failures, we first re-checked the data and evidence structure, then considered adjusting sampling parameters, or specifying alternative prior distributions. Where we suspected identifiability problems due to low degrees of freedom for estimating *τ* (<5), we used informative prior distributions for heterogeneity variance (*τ*^2^) based on empirical estimates from a meta-epidemiological study of meta-analyses. If there were remaining convergence or identifiability problems, we presented the results based on a common-effect model. Model fit was assessed using the posterior mean of the residual deviance and the deviance information criterion, which accounts for both model fit and complexity. For model selection, a difference of ≥5 points in the posterior mean residual deviance and DIC was considered meaningful, with lower values indicating better fit.

Subgroup effects were investigated for primary outcomes through network meta-regression on a dichotomous covariate (intervention type or comorbidity). The regression formula specified an interaction of the covariate with treatment and we used a vague prior distribution for the regression coefficient (normal distribution with mean = 0 and SD = 10). The remaining model parameters matched those of the selected primary analysis model. Pre-specified subgroup analyses were conducted on primary outcomes only.

### S1.4 Data preparation methods

For continuous outcomes, numerical outcome data were extracted primarily at arm-level as baseline and end-of-treatment (EoT) means and standard deviations (SDs). Statistical analysis was performed on contrast-level standardised mean difference (SMD) in change from baseline (CfB) data, where available.

If number analysed was unclear it was conservatively assumed that the number of participants with available outcome data (i.e., retained at EOT, or follow-up assessment) were used in the analysis. Missing SDs were derived from other within-study statistics (e.g., standard errors [SEs], 95% confidence intervals [CIs]) where possible. Otherwise, missing SDs were imputed based on the available means using a linear regression. The exception to this approach was analysis of urine benzoylecgonine (BE) levels, where estimates were reported on a log-transformed scale and the average SD for baseline log-transformed urine BE level was used. If outcomes were reported as a median, these were converted to a mean provided studies reported the range (i.e., min-max values) to account for skew in conversion^1^.

Arm-level CfB was rarely reported across outcomes. Where available, we used baseline and EOT means and SDs to estimate CfB and its SD, assuming empirically derived external estimate of pre-post correlation (r = 0.59)^2^. EOT means and SDs were only used where: (i) it was not possible to estimate CfB; (ii) the outcome was only relevant to the entire treatment duration (i.e., LDCA, adherence); (iii) the majority of studies reported measures of the outcome relevant to the entire treatment duration (i.e., proportion of UDS positive); or (iv) evidence suggested that the assumed pre-post correlation was inappropriate (i.e., urine BE level). For urine BE level, evidence suggests that for urine samples collected within a given week the within-participant correlation for estimates of log-transformed BE level is low^3^ so EOT means were compared instead of calculating CfB.

Relative effects were summarised using mean difference (MD) in CfB or EOT values. As it was not possible to align all different measures of adherence, craving, frequency of use, LDCA, proportion of UDS and quantity of use, these relative effects were expressed as SMD to maximise the number of datapoints included in the analysis. Following NICE TSU guidance^4^ to obtain internal measure-specific SD we used the average of the pooled (across treatment arms) baseline SDs reported by studies included in the analysis that used the corresponding outcome measure (e.g., median of pooled baseline SDs of all studies reporting dollars/week spent; **Table S6.1**). Imputed baseline SDs were not included in the data used to derive the measure-specific SD.

For each contrast, MD and its corresponding SE were rescaled by dividing each by the internal measure-specific baseline SD, to obtain SMD and its SE, which were used in the NMA. The same internal measure-specific SDs were used later to re-express the NMA results as MDs on the most common measure (by multiplying SMDs by measure-specific SD).

Table S6.1. Internal measure-specific standard deviations

| **Outcome** | **Measure** | **SD** | **N studies** |
| --- | --- | --- | --- |
| Quantity of use | Dollars spent per week ^a^ | 226.704 | 6 |
|  | Grams used per week ^a^ | 3.764 | 10 |
| Frequency of use | Halikas DIRS cocaine use item ^a^ | 0.915 | 1 |
|  | Number of uses per week ^a^ | 10.933 | 1 |
|  | Proportion of days used ^a^ | 0.292 | 24 |
| LDCA | Longest consecutive number of days ^b^ | 17.613 | 17 |
|  | Longest consecutive number of urine samples ^b^ | 11.796 | 2 |
| Adherence | Proportion of medication taken ^a^ | 0.214 | 11 |
|  | Proportion of treatment days adherent ^a^ | 0.163 | 4 |
|  | Proportion of urine samples adherent ^a^ | 0.182 | 8 |
|  | Proportion of supervised visits attended ^a^ | 0.459 | 1 |
| Craving | Brief Substance Craving Scale (BSCS) ^a^ | 2.706 | 11 |
|  | Cocaine Craving Questionnaire (CCQ) ^a^ | 1.382 | 3 |
|  | Numerical Rating Scale (NRS; 0 – 10) ^a^ | 2.664 | 13 |
| UDS proportion | Proportion of urine samples positive ^a^ | 0.313 | 24 |
|  | Proportion of days used (UDS verified) ^a^ | 0.238 | 4 |
|  | Proportion of weeks used (UDS verified) ^a^ | 0.290 | 1 |
| ^a^ Median SD; ^b^ Mean SD; DIRS: Drug Impairment Rating Scale^5^; LDCA: longest duration of continuous abstinence; UDS: urine drug screen | | | |

*Level of cocaine use (non-urine)*

Frequency of cocaine use was reported using various measures across the studies (e.g., number of days used in past week, proportion of days used, number of uses per week). Where feasible, we converted the reported values to those most commonly available (i.e., proportion of days used). Different measures of quantity of cocaine use were also reported, including dollars spent, ‘dime’ bags and grams. Based on reported estimates (i.e., dime bag ≈ $10 street worth of cocaine or crack cocaine)^6^, dime bags were first converted to dollars spent. Where quantities were expressed over different periods of time, these were converted to the most commonly available timeframe (i.e., quantity used per week).

*Longest duration of continuous abstinence (LDCA)*

LDCA was primarily reported using an indication of time (e.g., days, weeks) abstinent during treatment. In contrast to the point and continuous abstinence outcomes, these reports did not have to be verified by a urine sample. Where time abstinent was expressed using different metrics, this was converted to the most commonly available measure (i.e., continuous abstinence in days). Some studies reported LDCA using number of consecutive urine samples that were negative (e.g., BE <300 ng/ml) for cocaine.

*Benzoylecgonine (BE) level*

BE level (ng/mL) was primarily reported by studies on the (natural) log-transformed scale as this measure was typically skewed. Based on recommendations for performing meta-analysis with skewed data, to maintain fidelity to the available data^7^ we log-transformed raw estimates using available formulae for conversion^4^. Where studies presented arm-level means and SDs on the raw scale, their log transformed MD and corresponding SE was estimated using a Taylor series approximation that assumes a pooled SD on the log-transformed scale^7^. MDs on the log scale and their 95% credible intervals were re-expressed on the raw scale by using exponential function, as ratio of geometric means.

*Adherence*

Adherence was primarily reported as means and SDs, using various measures across the studies (e.g., proportion of medication taken as assessed by pill count, proportion of urine samples fluoresced for riboflavin). A minority of studies reported results as a proportion of the sample ‘adherent’ (e.g., took at least 80% of the prescribed medication, author defined as compliant). Accordingly, the meta-analysis was performed on the continuous outcome as this was most reported. To maximise use of available data, trials reporting the proportion of the sample adherent in each arm were combined with other trials reporting additive effects on the assumption that the underlying individual responses have a logistic distribution.

*Craving*

Per protocol, we included craving assessments only when measured with a validated tool (e.g., Cocaine Craving Questionnaire [CCQ]^8^, Brief Substance Craving Scale [BSCS]^9^, Obsessive Compulsive Drug Use Scale [OCDUS]^10^) or a clearly documented validated implementation for craving assessments (e.g., Minnesota Cocaine Craving Scale [MCCS]-Q1^11^, Visual Analogue Scales [VAS]). Ad-hoc single items including frequency measures (e.g., ‘days craving’) and composite scores without validation evidence were excluded. VAS and other numerical rating scales (NRS; e.g., 0–10) were checked for common anchor points (i.e., low = least possible craving, high = most possible craving) – whereby low values represent less craving. If appropriate, these were rescaled linearly to a common 0–10 metric. Following recommendations for analysing continuous outcome data a within-trial synthesis was conducted for trials reporting more than one craving outcome (e.g., NRS and BSCS) on a validated scale, taking into account the correlations between outcomes. MDs were first standardized using the internal-measure specific SDs (Table S6.1). Correlations between outcomes were assumed to be 0.70 and used to generate composite MDs.

*UDS proportion*

Based on clinical guidance, urine sample verified indicators of cocaine use were kept separate in the analyses. Most studies in this outcome category reported the proportion of urine samples positive for cocaine throughout the duration of treatment. Where feasible, for studies reporting alternative measures (e.g., number of tests positive during treatment), we converted the reported values to those most commonly available (i.e., proportion of urine samples positive). A minority of studies reported a Treatment Effectiveness Score (TES)^12^ or similar (i.e., number of urine samples negative for cocaine during treatment). To align with the direction of other measures (i.e., higher proportion indicates greater cocaine use) these were inverted, assuming missing urine samples were positive for cocaine. Trials reporting the proportion of the sample achieving a reduction in UDS verified cocaine use in each arm were combined with other trials reporting additive effects on the assumption that the underlying individual responses have a logistic distribution.

Dichotomous outcomes

Dichotomous outcome data were extracted as number of participants with event and total number of participants in each study arm. The same arm-level data was included in the NMA.

### S1.5 Description of interventions

As specified in the protocol (CRD42024596434), studies examining any pharmacological intervention were included. Eligible comparators were placebo or any other eligible pharmacological intervention.

For analysis, we collapsed estimates from studies reporting arm-level data for multiple doses of the same medication (e.g., fluoxetine 20 mg, fluoxetine 40 mg) into a single estimate within the relevant node (e.g., antidepressants and other serotonergic agents). We also collapsed arm-level data reported separately for various population subgroups (e.g., split by genotype, presence of a mental health condition). Similarly, intervention arms that combined the same medication with different adjuvant psychosocial therapies (e.g., contingency management conditions) were collapsed into a single estimate within the relevant node, as our focus was on pharmacological rather than behavioural components of treatment. For continuous outcomes, arm-level data were combined using the formulae for combining summary statistics across groups reported in Table 6.5.a of the Cochrane Handbook^1^. For dichotomous outcomes, arm-level data were combined by summing events and sample sizes across relevant arms.

The tables below provide the intervention categories used in the analysis, showing both the mechanistic (Table S5.1) and therapeutic strategy (Table S5.2) groupings, the specific medications included within each node and a brief description. Appropriate classification of each medication was based on information gathered across several resources: (i) the Anatomical Therapeutic Chemical (ATC) classification system; (ii) DrugBank (<https://go.drugbank.com/>); (iii) the KEGG DRUG Database (<https://www.genome.jp/kegg/drug/>); and (iv) the British National Formulary (BNF; <https://bnf.nice.org.uk/>).

Table S5.1 Therapeutic strategy categories

| **Intervention** | **Description** | **Specific medications** |
| --- | --- | --- |
| Agonist/Replacement Approaches (ARA) | Drugs that act primarily through dopaminergic and/or stimulant-like mechanisms that may mimic the subjective or reinforcing effects of cocaine. Includes amphetamine (e.g., dexamfetamine) and non-amphetamine CNS stimulants (e.g., methylphenidate), norepinephrine-dopamine reuptake inhibitors (NDRIs; e.g., bupropion), selective norepinephrine reuptake inhibitors (sNRIs; e.g., atomoxetine), dopamine receptor agonists (e.g., cabergoline), and dopamine precursors or combination therapies (e.g., levodopa plus carbidopa). | Atomoxetine  Bromocriptine  Bupropion  Cabergoline  Dexamfetamine  Dexamfetamine + Modafinil  Ergoloid mesylates  Levodopa + Carbidopa  Levodopa, Carbidopa + Ropinirole  Lisdexamfetamine  MAS-ER  Mazindol  Methamphetamine  Methylphenidate  Modafinil  Pemoline  Pergolide  Pramipexole  Ropinirole |
| Antidepressants and Other Serotonergic Agents (ANTID) | Drugs primarily developed for the treatment of depression, together with other agents whose pharmacological actions include modulation of serotonergic neurotransmission. This group encompasses selective serotonin reuptake inhibitors (e.g., fluoxetine, sertraline), serotonin–norepinephrine reuptake inhibitors (e.g., venlafaxine), serotonin receptor antagonists and reuptake inhibitors (e.g., nefazodone), noradrenergic and specific serotonergic antidepressants (e.g., mirtazapine), tricyclic antidepressants (e.g., desipramine), monoamine oxidase inhibitors (e.g., selegiline), serotonergic receptor agonists or antagonists used for other indications (e.g., buspirone, lorcaserin), and serotonergic modulators or precursors (e.g., tryptophan). | Buspirone  Citalopram  Desipramine  Fluoxetine  Lorcaserin  Mirtazapine  Nefazodone  Ondansetron  Paroxetine  Ritanserin  Selegiline  Sertraline  Tryptophan  Venlafaxine |
| Glutamate Modulators (GLUT) | Drugs that act primarily via glutamatergic mechanisms, including NMDA receptor antagonists (e.g., memantine, ketamine), NMDA partial agonists (e.g., D-cycloserine), and modulators of extracellular glutamate concentrations or release (e.g., riluzole, N-acetylcysteine). | Acamprosate  Amantadine  Ketamine  Memantine  D-cycloserine  Riluzole  N-acetylcysteine |
| Anticonvulsants and GABAergic Agents (ACV+GABA) | Drugs primarily used as anticonvulsants or muscle relaxants, as well as other agents with overlapping mechanisms such as sodium channel blockade or calcium channel modulation. This includes drugs acting via the GABA system (e.g., baclofen, tiagabine, topiramate), sodium channel blockers (e.g., carbamazepine, lamotrigine), calcium channel modulators (e.g., valproate), and mood stabilisers (e.g., lithium carbonate). | Baclofen  Carbamazepine  Gabapentin  Lamotrigine  Lithium carbonate  Tiagabine  Topiramate  Valproate  Vigabatrin |
| Antagonist/Blocker Approaches (ABA) | Drugs and biological interventions designed to reduce or prevent the psychoactive effects of cocaine, either by blocking central dopamine signalling (e.g., antipsychotics, dopamine β-hydroxylase inhibitors [disulfiram]), inhibiting adrenergic pathways (e.g., α- or β-adrenergic receptor antagonists, reserpine), or by altering cocaine pharmacokinetics through enzyme-based therapies (e.g., TV-1380) or immunotherapies (e.g., TA-CD vaccine). | Aripiprazole  Carvedilol  Disulfiram  Doxazosin  Olanzapine  Propranolol  Quetiapine  Reserpine  Risperidone  TA-CD vaccine  TV-1380 |
| Opioid Agonists and Antagonists (OAA) | Drugs that act as agonists, partial agonists, or antagonists at opioid receptors. This group includes agents used in opioid substitution therapy (e.g., methadone, LAAM, buprenorphine), opioid receptor partial agonists (e.g., buprenorphine), and opioid receptor antagonists used for relapse prevention (e.g., naltrexone). | Buprenorphine  Buprenorphine + Naloxone  LAAM  Methadone  Naltrexone |
| Combination Pharmacotherapy (COMBO) | Combinations of two medications which don't otherwise fit into existing node structures (e.g., both agonist replacement therapy) or include drugs from one or more existing nodes, which may achieve additive or synergistic effects. | Amantadine + Propranolol  Coenzyme Q10 + L-carnitine  Disulfiram + Naltrexone  Gabapentin + Sertraline  MAS-ER + Topiramate  Metyrapone + Oxazepam |
| Other Neuroactive Agents (ONA) | Pharmacologically diverse agents grouped pragmatically because their primary mechanisms do not align closely with other intervention categories. Includes nicotinic receptor modulators (e.g., varenicline, mecamylamine), acetylcholinesterase inhibitors (e.g., donepezil, galantamine), cannabinoids (e.g., cannabidiol), calcium channel blockers (e.g., amlodipine), hormonal agents (e.g., dehydroepiandrosterone, pregnenolone, progesterone), anti-inflammatory agents (e.g., celecoxib), nootropic agents (e.g., piracetam, citicoline), local anaesthetics with central sodium channel blocking effects (e.g., lidocaine), and other unique mechanisms including a neuropeptide (e.g., oxytocin), a xanthine derivative (e.g., pentoxifylline), a PPAR-γ agonist (e.g., pioglitazone), and a steroidogenesis inhibitor (e.g., ketoconazole). | Amlodipine  Biperiden  Cannabidiol  Celecoxib  Citicoline  Dehydroepiandrosterone  Donepezil  Galantamine  Ketoconazole  Lidocaine  Mecamylamine  Oxytocin  Pentoxifylline  Pioglitazone  Piracetam  Pregnenolone  Progesterone  Varenicline |

Table S5.1 Mechanistic intervention categories

| **Intervention** | **Description and specific medications** |
| --- | --- |
| AChEIs | Acetylcholinesterase Inhibitors (AChEIs)  Donepezil, galantamine |
| ACV+MS | Anticonvulsants + mood stabilisers (ACV+MS)  Carbamazepine, lamotrigine, lithium, tiagabine, topiramate, valproate, vigabatrin |
| Adrenergic antagonists (ADR-ANT) | Drugs that bind to and inhibit the functions of alpha (α) or beta (β) adrenergic receptors  Carvedilol, doxazosin, propranolol |
| Amlodipine | Dihydropyridine calcium-channel blocker  Amlodipine |
| Atomoxetine | Selective norepinephrine reuptake inhibitor (sNRI)  Atomoxetine |
| Baclofen | Gamma-aminobutyric acid type B (GABA-B) receptor agonist  Baclofen |
| Buspirone | Anxiolytic (serotonin 5-HT1A partial agonist)  Buspirone |
| Cannabidiol  (CBD) | Non-psychoactive cannabinoid; modulates endocannabinoid and serotonin systems, with low affinity for CB1/CB2 receptors  Cannabidiol |
| Celecoxib | Non-steroidal anti-inflammatory drug (NSAID)  Celecoxib |
| Coc-PK | Cocaine targeting pharmacokinetic interventions (Coc-PK)  TA-CD vaccine, TV-1380 (AlbuChE) |
| CQ10+LC | Mitochondrial cofactor (CoQ10) + carrier molecule in the transport of long chain fatty acids across the inner mitochondrial membrane (l-carnitine)  Coenzyme Q10 + levocarnitine |
| Disulfiram | Aldehyde dehydrogenase (ALDH) inhibitor  Disulfiram |
| Dopamine agonists (DA) | Drugs that bind to and activate dopamine receptors  Bromocriptine, cabergoline, pergolide, pramipexole, ropinirole |
| Ergoloid mesylates | Mixed agonist/antagonist effects at adrenergic, dopaminergic and serotonergic receptors  Ergoloid mesylates |
| Gabapentin | Gabapentinoid that targets the the α2δ-1 auxiliary subunit of voltage-gated Ca2+ channels  Gabapentin |
| Ketoconazole | Imidazole derivative which acts as a potent inhibitor of cortisol and aldosterone synthesis  Ketoconazole |
| L-DOPA+ | Drugs used in combination with levodopa; a dopamine precursor  Levodopa + carbidopa, levodopa + ropinirole |
| Lidocaine | Local anaesthetic that blocks voltage-gated Na ^+^ channels (VGSC/NaVs)  Lidocaine |
| Lorcaserin | Highly selective serotonin (5-HT2C) receptor agonist  Lorcaserin |
| mAchR antagonists  (mAChR-Ant) | Muscarinic cholinergic receptor (mAchR) antagonist  Biperiden |
| Mixed action antidepressants (MA-AD) | Monoamine-oxidase inhibitors (MAOIs), noradrenaline and specific serotonergic antidepressants (NaSSAs), serotonin-noradrenaline reuptake inhibitors (SNRIs) and tricyclic antidepressants (TCAs)  Selegiline, mirtazipine, venlafaxine, desipramine |
| Mecamylamine | A non-competitive antagonist of nicotine receptor gated ion channels  Mecamylamine |
| Metyrapone + Oxazepam (MTP+OXZ) | A steroid 11-beta-monooxygenase inhibitor (metyrapone) + benzodiazepine (oxazepam)  Metyrapone + Oxazepam |
| NDRIs | Norepinephrine-dopamine reuptake inhibitors (NDRIs)  Bupropion |
| Neuropeptide | Neuropeptide  Oxytocin |
| Non-NMDAR glutamate modulators  (NNM-GluM) | Class of drugs that influence the activity of glutamate receptors in the brain, excluding the NMDA receptor subtype  D-cycloserine, n-acetylcysteine, riluzole |
| NMDAR antagonists (NMDAR-ANT) | Drugs that block or inhibit the action of N-Methyl-D-aspartate receptors (NMDARs)  Acamprosate, amantadine, ketamine, memantine |
| Nootropics | Non-stimulant cognitive enhancers  Piracetam, citicoline |
| Naltrexone (NTX) | Naltrexone (NTX); opioid antagonist with a high affinity to mu-opioid receptors  Naltrexone |
| OAT | Opioid Agonist Treatment (OAT)  Buprenorphine, buprenorphine + naloxone, levacetylmethadol (LAAM), methadone |
| Ondansetron | Serotonin 5HT-3 receptor antagonist which blocks receptors in the gastro-intestinal tract and in the CNS  Ondansetron |
| Pioglitazone | Peroxisome proliferator-activated receptor gamma (PPAR-γ) agonist  Pioglitazone |
| Pentoxifylline | Synthetic dimethylxanthine derivative  Pentoxifylline |
| Reserpine | Adrenergic blocking agent via the disruption of norepinephrine vesicular storage  Reserpine |
| Sex hormones (SEX-H) | Sex hormones  Dehydroepiandrosterone (DHEA), pregnenolone, progesterone |
| SGAs | Second-generation or ‘atypical’ antipsychotics (SGAs)  Aripiprazole, olanzapine, quetiapine, risperidone |
| SSRIs and other serotonergic agents  (SSRI+OSA) | Selective serotonin reuptake inhibitor (SSRI) antidepressants, ritanserin (5-HT2 receptor antagonist, serotonin antagonist and reuptake inhibitors (SARIs; nefazodone), tryptophan (serotonin precursor)  Citalopram, fluoxetine, paroxetine, sertraline, ritanserin, nefazodone, tryptophan |
| Stimulants (STIM) | Amphetamine and non-amphetamine central nervous system (CNS) stimulants  Dexamfetamine, MAS-ER, mazindol, methamphetamine, modafinil, lisdexamfetamine, methylphenidate, pemoline |
| Varenicline | Selective nicotinic acetylcholine receptor partial agonist  Varenicline |

### S1.6 Network meta-analysis results for mechanistic treatment classification

Results of NMAs using mechanistic treatment classifications are detailed below. Statistical approaches are the same as reported in the main manuscript. Results are primarily reported relative to placebo, however, full contrasts for each outcome are available in Supplement X. Network plots are presented in Supplement 2. Forest plots are presented for primary outcomes only.

Table S1.6.1. List of mechanistic treatments included in NMAs

| **Treatment** | **Abbreviation** | **Studies** |
| --- | --- | --- |
| Acetylcholinesterase inhibitors | AChEIs | 4 |
| Anticonvulsants and mood stabilisers | ACV+MS | 15 |
| Adrenergic antagonists | ADR-ANT | 3 |
| Amlodipine |  | 1 |
| Atomoxetine |  | 1 |
| Baclofen |  | 2 |
| Buspirone |  | 2 |
| Cannabidiol | CBD | 1 |
| Celecoxib |  | 1 |
| Cocaine targeting pharmacokinetic interventions | Coc-PK | 3 |
| Co-enzyme Q10 and l-carnitine | CQ10+LC | 1 |
| Disulfiram |  | 9 |
| Dopamine agonists | DA | 5 |
| Ergoloid mesylates |  | 1 |
| Gabapentin |  | 2 |
| Ketoconazole |  | 1 |
| Drugs used in combination with levodopa | L-DOPA+ | 8 |
| Lidocaine |  | 1 |
| Lorcaserin |  | 2 |
| Muscarinic cholinergic receptor antagonist | mAChR-Ant | 2 |
| Mixed action antidepressants | MA-AD | 9 |
| Mecamylamine |  | 1 |
| Metyrapone and oxazepam | MTP+OXZ | 1 |
| Norepinephrine-dopamine reuptake inhibitors | NDRIs | 2 |
| Non-NMDAR glutamate modulators | NNM-GluM | 3 |
| N-Methyl-D-aspartate receptor antagonists | NMDAR-ANT | 10 |
| Nootropics |  | 3 |
| Naltrexone | NTX | 3 |
| Opioid agonist treatment | OAT | 5 |
| Ondansetron |  | 2 |
| Pioglitazone |  | 1 |
| Pentoxifylline |  | 1 |
| Reserpine |  | 2 |
| Sex hormones | SEX-H | 3 |
| Second-generation or ‘atypical’ antipsychotics | SGAs | 7 |
| Selective serotonin reuptake inhibitor and other serotonergic agents | SSRI+OSA | 16 |
| Stimulants | STIM | 24 |
| Varenicline |  | 2 |

Effectiveness

Continuous abstinence during treatment

NMA included 46 studies of 22 treatments, including direct evidence from a maximum of 9 studies per contrast (stimulants). Results of random-effects NMA suggest that stimulants + anticonvulsants may have beneficial effects relative to placebo (OR 6.16, 95% CrI: 1.66, 26.7, Pr = 0.99) (Figure S1.6.1). Other placebo comparisons were consistent with no effect. Note that while L-DOPA+ was included in the NMA, the estimate was extremely imprecise and is not displayed in Figure S1.6.1 for clarity. The estimate is presented alongside other contrasts in Supplement 2 but should not be interpreted with confidence. Relative effects of different active treatments suggest that stimulants + anticonvulsants may be more beneficial than disulfiram and naltrexone while all other contrasts were consistent with the possibility of no effect. All estimates were rated as very low certainty.

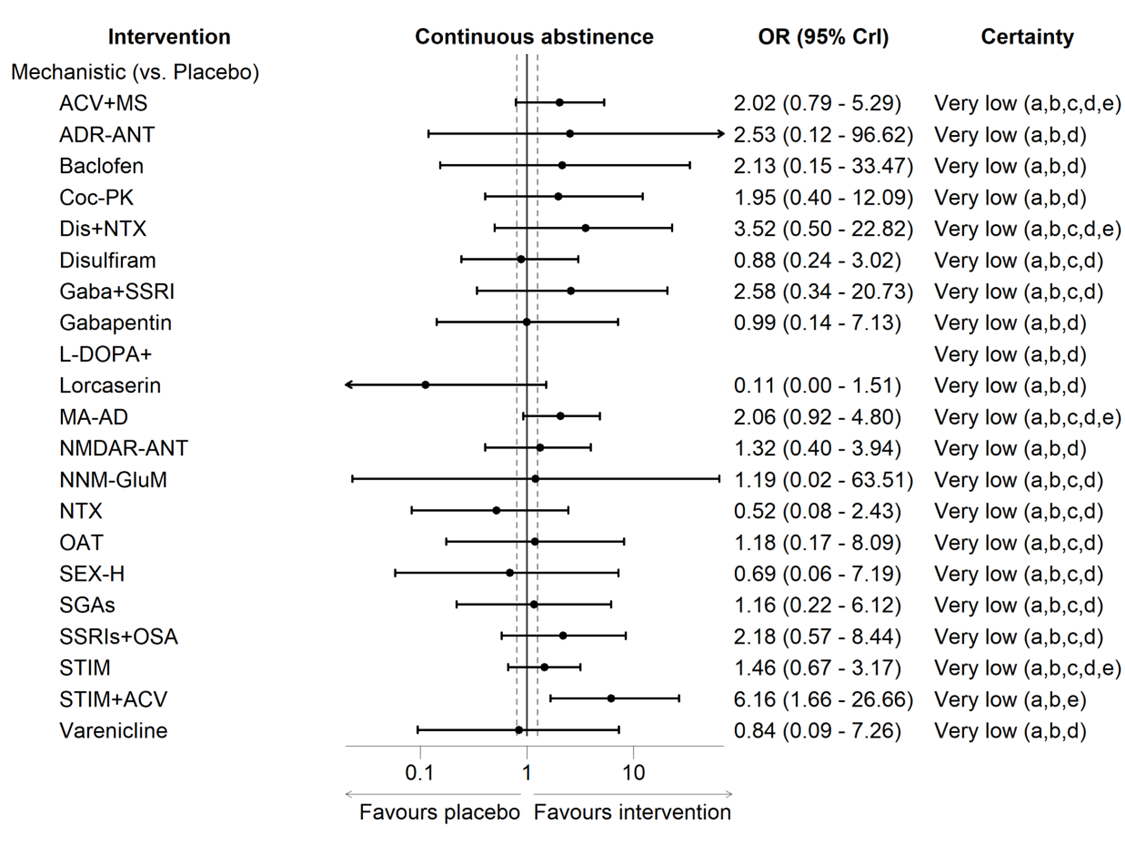

Figure S1.6.1. NMA results for continuous abstinence during treatment

Continuous abstinence at follow-up

NMA included 5 studies of 5 treatments, including direct evidence from a maximum of 2 studies per contrast (NMDA receptor antagonists, mixed-action antidepressants, and anticonvulsants + mood stabilizers). Results of common-effect NMA suggest that mixed-action antidepressants may have beneficial effects relative to placebo (OR 4.04, 95% CrI: 1.20, 14.6, Pr = 0.99). Other placebo comparisons were consistent with no effect. All relative effects of different active treatments showed evidence consistent with the possibility of no effect.

Point abstinence at end of treatment

NMA included 22 studies of 21 treatments, including direct evidence from a maximum of 3 studies per contrast (stimulants and anticonvulsants + mood stabilisers). Results of random-effects NMA suggest that anticonvulsants + mood stabilisers may lead to meaningful improvements relative to placebo (OR 12.2, 95% CrI: 3.43, 49.7, Pr = 0.99) (Figure S1.6.2). DA and ondansetron were included in NMA and showed potentially beneficial effects. However, the estimates were extremely imprecise and should not be interpreted with confidence. To that end, estimates for L-DOPA+ and ondansetron are not displayed in Figure S1.6.2 for clarity but are presented alongside other contrasts in Supplement X for completeness. All other placebo and active treatment comparisons were consistent with the possibility of no effect. All estimates were rated as very low certainty.

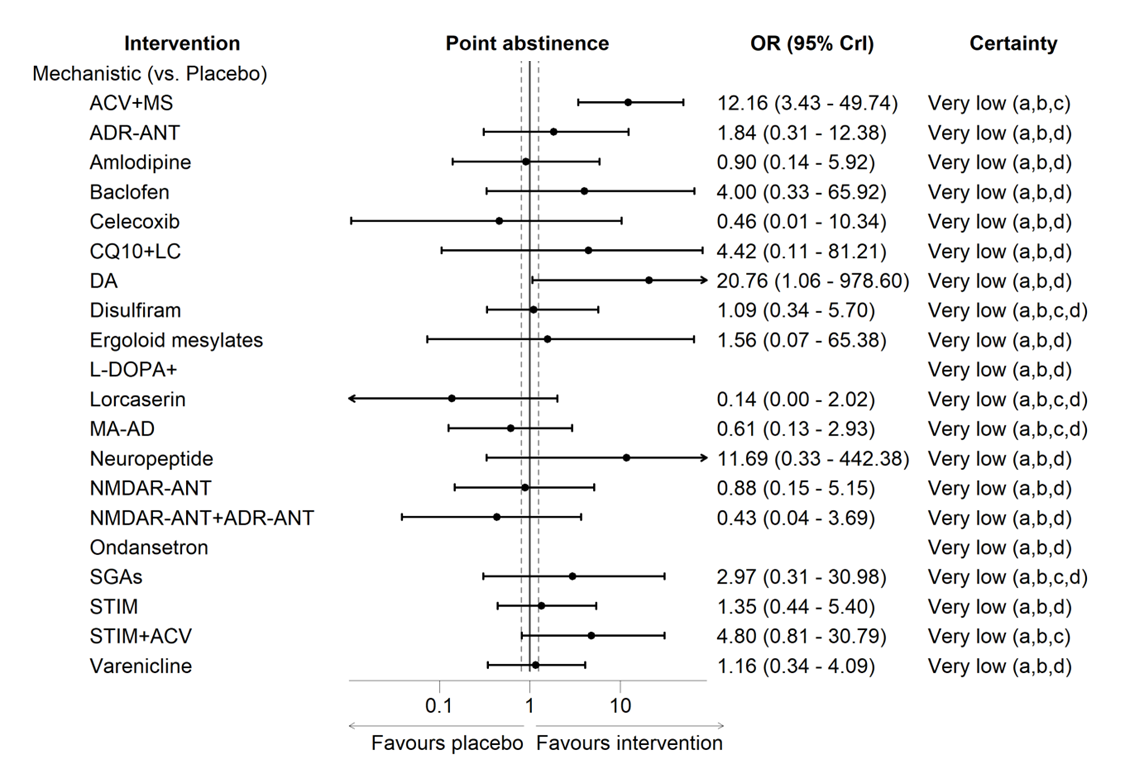

Figure S1.6.1. NMA results for point abstinence at the end of treatment

Point abstinence at end of follow-up

NMA included 7 studies of 7 treatments, including direct evidence from a maximum of 2 studies per contrast (NMDA-receptor antagonists). Results of common-effect NMA suggest that NDRIs (OR 3.19, 95% CrI: 1.32, 8.02) and NMDA-receptor antagonists (OR 4.29, 95% CrI: 1.41, 14.6) may have beneficial effects relative to placebo (Table 6).

Longest duration of continuous abstinence

NMA included 19 studies of 12 treatments were assessed, including direct evidence from a maximum of 5 studies per contrast (*stimulants*). All placebo and active treatment comparisons from random-effects NMA showed evidence consistent with the possibility of no effect relative to placebo (Table 7).

Extent of use – frequency

NMA included 40 studies of 22 treatments, including direct evidence from a maximum of 8 studies per contrast (anticonvulsants + mood stabilisers). Results of random-effects NMA suggest that anticonvulsants + mood stabilisers (MD -5.49%, 95% CrI: -12.5, -1.40, Pr = 0.99) and disulfiram (MD -7.98, 95% Cr I: -16.3, 0.00, Pr = 0.71) may lead to reductions in frequency of cocaine use relative to placebo. All other placebo and active treatment comparisons were consistent with the possibility of no effect.

Extent of use at follow-up – frequency

NMA included 7 studies of 6 treatments, including direct evidence from a maximum of 2 studies per contrast (disulfiram and NMDA-receptor antagonists). Results of common-effect NMA suggest that anticonvulsants + mood stabilisers may lead to reductions in the frequency of cocaine use relative to placebo (MD -15.8%, 95% CrI: -30.8, -1.09, Pr = 0.86). All other placebo and active treatment comparisons were consistent with the possibility of no effect.

Extent of use – quantity

NMA included 19 studies of 11 treatments including direct evidence from a maximum of 4 studies per contrast (NMDA-receptor antagonists and disulfiram). All placebo and active treatment comparisons from random-effects NMA showed evidence consistent with the possibility of no effect.

Extent of use at follow-up – quantity

For the therapeutic strategy and mechanistic classifications, the same four studies assessed five classes/treatments. As such, the analysed data for each analysis was exactly the same, but with different classification labels. Consequently, the results of both analyses were identical. All placebo and active treatment comparisons from common-effect NMA showed evidence consistent with the possibility of no effect. SMD estimates are re-expressed as grams of cocaine used per week during treatment period. All placebo and active treatment comparisons from random-effects NMA showed evidence consistent with the possibility of no effect (Tables 5 and 7).

Extent of use – urine benzoylecgonine levels

NMA included 12 studies of 14 treatments including direct evidence from a maximum of 4 studies per contrast (anticonvulsants + mood stabilisers). All placebo and active treatment comparisons from random-effects NMA showed evidence consistent with the possibility of no effect.

Extent of use – proportion of urinary drug screenings

NMA included 45 studies of 25 treatments including direct evidence from a maximum of 8 studies per contrast (STIM). All placebo and active treatment comparisons from random-effects NMA showed evidence consistent with the possibility of no effect.

Craving

NMA included 39 studies of 26 treatments including direct evidence from a maximum of 8 studies per contrast (anticonvulsants + mood stabilisers). Results of random-effects NMA suggest that STIM may lead to meaningful reductions in craving relative to placebo (MD -1.33, 95% CrI: -2.72, 0.07, Pr = 0.88). All other placebo comparisons were consistent with the possibility of no effect. Relative effects of different active treatments suggest that stimulants may be more beneficial than NMDAR antagonists, non-NMDAR glutamate modulators, and *second generation or atypical antipsychotics* while all other contrasts were consistent with the possibility of no effect.

Acceptability

Dropout for any reason

NMA included 135 studies of 43 treatments including direct evidence from a maximum of 18 studies per contrast (anticonvulsants + mood stabilisers). All estimates were rated as very low certainty, except acetylcholinesterase inhibitors, non-NMDAR glutamate modulators, and reserpine which were rated as low certainty. Results of random-effects NMA suggest that adrenergic antagonists may result in reduced dropout for any reason compared with placebo (OR 0.56, 95% CrI: 0.32, 0.96, Pr = 0.91) (Figure S1.6.3). All other placebo comparisons were consistent with the possibility of no effect. Note that while opioid agonist treatment (OAT) was included in the NMA, the estimate was extremely imprecise and is not displayed in Figure S1.6.3 for clarity. The estimate is presented alongside other results in Supplement X for completeness. Relative effects of different active treatments suggest that adrenergic antagonists may also be more beneficial than acetylcholinesterase inhibitors.

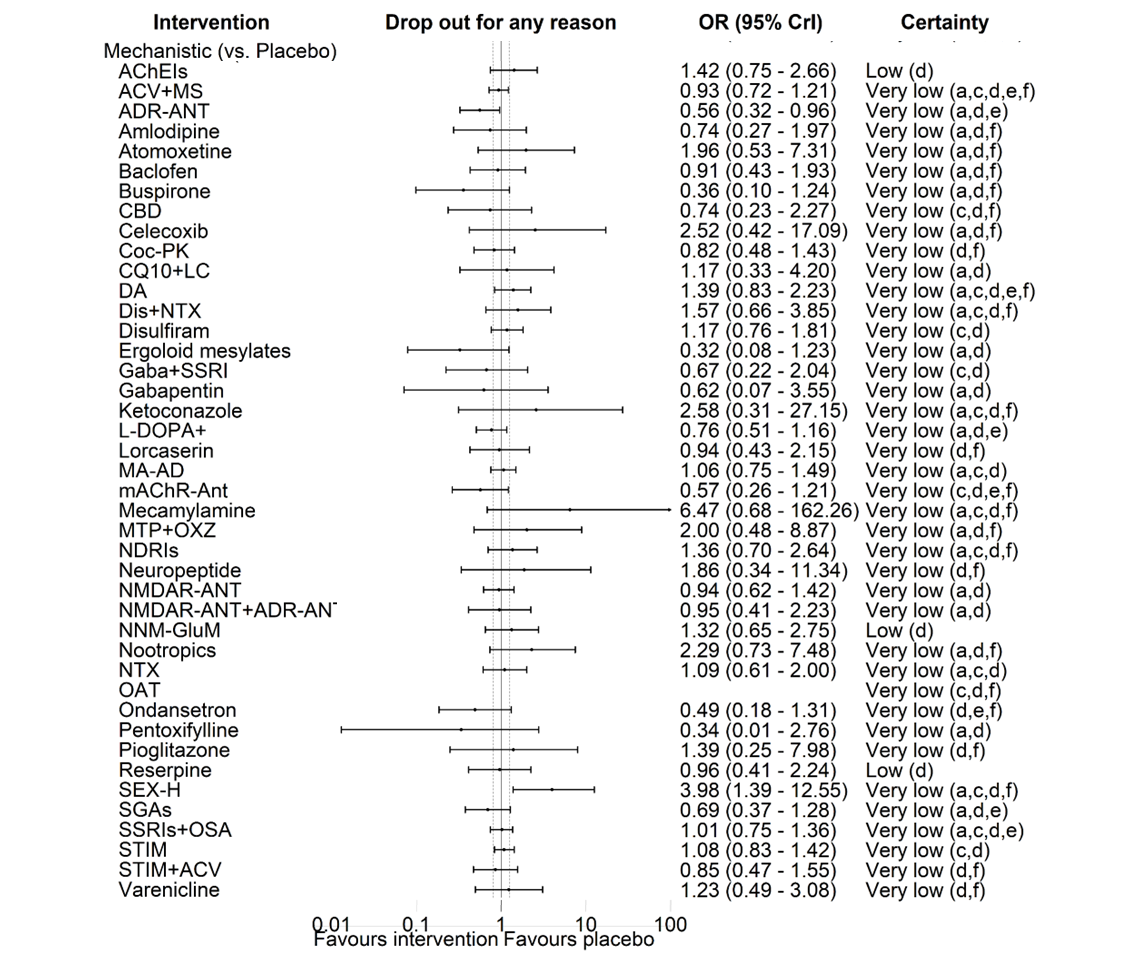

Figure S1.6.3 NMA results for dropout for any reason

Dropout due to adverse events

NMA included 39 studies of 21 treatments including direct evidence from a maximum of 9 studies per contrast (STIM). All estimates from random-effects NMA showed evidence consistent with the possibility of no effect relative to placebo.

Adherence

NMA included 52 studies of 27 treatments including direct evidence from a maximum of 8 studies per contrast (STIM). All placebo comparisons from random-effects NMA showed evidence consistent with the possibility of no effect. Relative effects of different active treatments suggest that SSRIs + other serotonergic agents may be beneficial relative to NMDA-receptor antagonists and anticonvulsants + mood stabilisers while all other contrasts were consistent with the possibility of no effect.

Safety

Serious adverse events

NMA included 41 studies of 27 treatments including direct evidence from a maximum of 6 studies per contrast (anticonvulsants + mood stabilisers). As above, the number of reported serious adverse events across studies was typically low, contributing to very imprecise estimates. No consistent evidence of effect was observed across any placebo or active treatment comparisons.

Mortality

For the therapeutic strategy and mechanistic classifications, 4 studies of 5 classes/treatments were included in NMA. Only four deaths were reported across all trials. Consequently, the NMA estimates were extremely imprecise with wide 95% CrI. No consistent evidence of effect was observed across any placebo or active treatment comparisons.

### S1.7 Risk of bias assessments for included studies

| **Study** | **Interventions** | **Specific contrasts** | **Outcome** | **Domain 1** | **Domain 2** | **Domain 3** | **Domain 4** | **Domain 5** | **Overall** | **Comments** |
| --- | --- | --- | --- | --- | --- | --- | --- | --- | --- | --- |
| Afshar 2012 | 1. Placebo 2. Mirtazapine | All | Adherence | Some concerns | High | Some concerns | Low | Some concerns | High | D1: Limited information regarding randomization and allocation process. Baseline demographics only reported for subset of randomised participants. D2: Analysis was restricted to N treated (i.e., not ITT or mITT). D3: Some missing data but unlikely that missingness depended on the true value of outcome. D5: No evidence of a pre-specified analysis plan. |
|  |  | All | Extent of use (BE level; EoT) | Some concerns | High | Some concerns | Low | Some concerns | High | D1: Limited information regarding randomization and allocation process. Baseline demographics only reported for subset of randomised participants. D2: Analysis was restricted to N treated (i.e., not ITT or mITT). D3: Some missing data but unlikely that missingness depended on the true value of outcome. D5: No evidence of a pre-specified analysis plan. |
|  |  | All | Craving | Some concerns | High | Some concerns | Low | Some concerns | High | D1: Limited information regarding randomization and allocation process. Baseline demographics only reported for subset of randomised participants. D2: Analysis was restricted to N treated (i.e., not ITT or mITT). D3: Some missing data but unlikely that missingness depended on the true value of outcome. D5: No evidence of a pre-specified analysis plan. |
| Alterman 1992 | 1. Placebo 2. Amantadine | All | Dropout (any reason) | Some concerns | Low | Low | Low | Low | Some concerns | D1: Limited information regarding allocation process. |
|  |  | All | Point abstinence (follow-up) | Some concerns | Low | Low | Low | Some concerns | Some concerns | D1: Limited information regarding allocation process. D5: No evidence of a pre-specified analysis plan. |
|  |  | All | Extent of use (frequency; follow-up) | Some concerns | Low | Low | Low | Some concerns | Some concerns | D1: Limited information regarding allocation process. D5: No evidence of a pre-specified analysis plan. |
| Anderson 2009 | 1. Placebo 2. Modafinil (200mg) 3. Modafinil (400mg) | All | Dropout (any reason) | Low | Low | Low | Low | Low | Low |  |
|  |  | All | Dropout (adverse events) | Low | Low | Some concerns | Low | Low | Some concerns | D3: Some missing data but unlikely that missingness depended on the true value of outcome. |
|  |  | All | Longest duration of continuous abstinence | Low | Some concerns | Some concerns | Low | Some concerns | Some concerns | D2: Analysis restricted to those receiving first dose - but judged unlikely to have substantial impact on the results. D3: Some missing data but unlikely that missingness depended on the true value of outcome. D5: No evidence of a pre-specified analysis plan. |
| Baldacara 2013 | 1. Placebo 2. Disulfiram | All | Dropout (any reason) | Low | Low | Low | Low | Low | Low |  |
|  |  | All | Point abstinence (EoT) | Low | Low | High | Low | High | High | D3: Some missing data and likely that it could depend on the true value of the outcome. D5: No evidence of a pre-specified analysis plan. The registration history suggests that the result could have been selected from multiple eligible outcome measurements. |
|  |  | All | Extent of use (frequency; EoT) | Low | Some concerns | High | Low | High | High | D2: Method for analysis unclear - but judged unlikely to have substantial impact on the results. D3: Some missing data and likely that it could depend on the true value of the outcome. D5: No evidence of a pre-specified analysis plan. The registration history suggests that the result could have been selected from multiple eligible outcome measurements. |
|  |  | All | Extent of use (quantity; EoT) | Low | Some concerns | High | Low | High | High | D2: Method for analysis unclear - but judged unlikely to have substantial impact on the results. D3: Some missing data and likely that it could depend on the true value of the outcome. D5: No evidence of a pre-specified analysis plan. The registration history suggests that the result could have been selected from multiple eligible outcome measurements. |
| Baldacara 2016 | 1. Placebo 2. Topiramate | All | Dropout (any reason) | Some concerns | Low | Low | Low | Low | Low | D1: Limited information regarding allocation process. |
|  |  | All | Point abstinence (EoT) | Some concerns | Low | Low | Low | Some concerns | Some concerns | D1: Limited information regarding allocation process. D5: No evidence of a pre-specified analysis plan. |
|  |  | All | Extent of use (frequency; EoT) | Some concerns | Low | Low | Low | Some concerns | Some concerns | D1: Limited information regarding allocation process. D5: No evidence of a pre-specified analysis plan. |
|  |  | All | Extent of use (quantity; EoT) | Some concerns | Low | Low | Low | Some concerns | Some concerns | D1: Limited information regarding allocation process. D5: No evidence of a pre-specified analysis plan. |
| Batki 1996 | 1. Placebo 2. Fluoxetine | All | Dropout (any reason) | Some concerns | Low | Low | Low | Low | Some concerns | D1: Limited information regarding randomization and allocation process. |
|  |  | All | Dropout (adverse events) | Some concerns | Low | High | Low | Low | High | D1: Limited information regarding randomization and allocation process. D3: Some missing data and likely that it could depend on the true value of the outcome. |
| Becker 2020 | 1. Placebo 2. Lidocaine | All | Extent of use (frequency; follow-up) | Low | Some concerns | Some concerns | Low | Low | Some concerns | D2: Analysis restricted to those receiving first dose - but judged unlikely to have substantial impact on the results. D3: Some missing data but unlikely that missingness depended on the true value of outcome. |
|  |  | All | Extent of use (quantity; follow-up) | Low | Some concerns | Some concerns | Low | Low | Some concerns | D2: Analysis restricted to those receiving first dose - but judged unlikely to have substantial impact on the results. D3: Some missing data but unlikely that missingness depended on the true value of outcome. |
| Berger 2005 | 1. Placebo 2. Gabapentin 3. Reserpine 4. Lamotrigine | All | Dropout (any reason) | Some concerns | High | Low | Low | Low | High | D1: Limited information regarding allocation process. D2: Part of CREST protocol is modified blind, meaning participants and people responsible for intervention delivery likely unblinded. |
|  |  | 2 vs 1;  4 vs 2;  4 vs 3 | Dropout (adverse events) | Some concerns | High | Some concerns | Some concerns | Low | High | D1: Limited information regarding allocation process. D2: Part of CREST protocol is modified blind, meaning participants and people responsible for intervention delivery likely unblinded. D3: Some missing data but unlikely that missingness depended on the true value of outcome. D4: The outcome was self-reported and participants may be aware of their group allocation, but unlikely to be a substantial portion of participants. |
|  |  | 3 vs 1;  4 vs 1;  3 vs 2 | Dropout (adverse events) | Some concerns | High | High | Some concerns | Low | High | D1: Limited information regarding allocation process. D2: Part of CREST protocol is modified blind, meaning participants and people responsible for intervention delivery likely unblinded. D3: Some missing data and likely that it could depend on the true value of the outcome. D4: The outcome was self-reported and participants may be aware of their group allocation, but unlikely to be a substantial portion of participants. |
|  |  | 2 vs 1;  4 vs 2;  4 vs 3 | Adherence | Some concerns | High | Some concerns | Low | Some concerns | High | D1: Limited information regarding allocation process. D2: Part of CREST protocol is modified blind, meaning participants and people responsible for intervention delivery likely unblinded. D3: Some missing data but unlikely that missingness depended on the true value of outcome. D5: No evidence of a pre-specified analysis plan. |
|  |  | 3 vs 1;  4 vs 1;  3 vs 2 | Adherence | Some concerns | High | High | Low | Some concerns | High | D1: Limited information regarding allocation process. D2: Part of CREST protocol is modified blind, meaning participants and people responsible for intervention delivery likely unblinded. D3: Some missing data and likely that it could depend on the true value of outcome. D5: No evidence of a pre-specified analysis plan. |
|  |  | All | Serious adverse events | Some concerns | High | Low | Low | Low | High | D1: Limited information regarding allocation process. D2: Part of CREST protocol is modified blind, meaning participants and people responsible for intervention delivery likely unblinded. |
|  |  | 2 vs 1;  4 vs 2;  4 vs 3 | Extent of use (BE level; EoT) | Some concerns | High | Some concerns | Low | Some concerns | High | D1: Limited information regarding allocation process. D2: Part of CREST protocol is modified blind, meaning participants and people responsible for intervention delivery likely unblinded. D3: Some missing data but unlikely that missingness depended on the true value of outcome. D5: No evidence of a pre-specified analysis plan. |
|  |  | 3 vs 1;  4 vs 1;  3 vs 2 | Extent of use (BE level; EoT) | Some concerns | High | High | Low | Some concerns | High | D1: Limited information regarding allocation process. D2: Part of CREST protocol is modified blind, meaning participants and people responsible for intervention delivery likely unblinded. D3: Some missing data and likely that it could depend on the true value of the outcome. D5: No evidence of a pre-specified analysis plan. |
|  |  | 2 vs 1;  4 vs 2;  4 vs 3 | Extent of use (frequency; EoT) | Some concerns | High | Some concerns | Some concerns | Some concerns | High | D1: Limited information regarding allocation process. D2: Part of CREST protocol is modified blind, meaning participants and people responsible for intervention delivery likely unblinded. D3: Some missing data but unlikely that missingness depended on the true value of outcome. D4: The outcome was self-reported and participants may be aware of their group allocation, but unlikely to be a substantial portion of participants. D5: No evidence of a pre-specified analysis plan. |
|  |  | 3 vs 1;  4 vs 1;  3 vs 2 | Extent of use (frequency; EoT) | Some concerns | High | High | Some concerns | Some concerns | High | D1: Limited information regarding allocation process. D2: Part of CREST protocol is modified blind, meaning participants and people responsible for intervention delivery likely unblinded. D3: Some missing data but unlikely that missingness depended on the true value of outcome. D4: The outcome was self-reported and participants may be aware of their group allocation, but unlikely to be a substantial portion of participants. D5: No evidence of a pre-specified analysis plan. |
|  |  | 2 vs 1;  4 vs 2;  4 vs 3 | Craving | Some concerns | High | Some concerns | Some concerns | Some concerns | High | D1: Limited information regarding allocation process. D2: Part of CREST protocol is modified blind, meaning participants and people responsible for intervention delivery likely unblinded. D3: Some missing data but unlikely that missingness depended on the true value of outcome. D4: The outcome was self-reported and participants may be aware of their group allocation, but unlikely to be a substantial portion of participants. D5: No evidence of a pre-specified analysis plan. |
|  |  | 3 vs 1;  4 vs 1;  3 vs 2 | Craving | Some concerns | High | High | Some concerns | Some concerns | High | D1: Limited information regarding allocation process. D2: Part of CREST protocol is modified blind, meaning participants and people responsible for intervention delivery likely unblinded. D3: Some missing data and likely that it could depend on the true value of the outcome. D4: The outcome was self-reported and participants may be aware of their group allocation, but unlikely to be a substantial portion of participants. D5: No evidence of a pre-specified analysis plan. |
| Bisaga 2006 | 1. Placebo 2. Gabapentin | All | Serious adverse events | Low | Low | Low | Low | Low | Low |  |
|  |  | All | Continuous abstinence (treatment) | Low | Low | High | Low | Some concerns | High | D3: Some missing data and likely that it could depend on the true value of the outcome. D5: No evidence of a pre-specified analysis plan. |
|  |  | All | Extent of use (proportion tests/days; EoT) | Low | Low | Low | Low | Some concerns | Some concerns | D5: No evidence of a pre-specified analysis plan. |
| Bisaga 2010 | 1. Placebo 2. Memantine | All | Dropout (any reason) | Low | Low | Low | Low | Low | Low |  |
|  |  | All | Adherence | Low | Low | Some concerns | Low | Some concerns | Some concerns | D3: Some missing data but unlikely that missingness depended on the true value of outcome. D5: No evidence of a pre-specified analysis plan. |
|  |  | All | Serious adverse events | Low | Low | Low | Low | Low | Low |  |
|  |  | All | Continuous abstinence (treatment) | Low | Low | Some concerns | Low | Some concerns | Some concerns | D3: Some missing data but unlikely that missingness depended on the true value of outcome. D5: No evidence of a pre-specified analysis plan. |
| Blevins 2021 | 1. Placebo 2. Ondansetron | All | Dropout (any reason) | Low | Low | Low | Low | Low | Low |  |
|  |  | All | Adherence | Low | High | High | Low | Some concerns | High | D2: Analysis was restricted to N treated (i.e., not ITT or mITT).  D3: Some missing data and likely that it could depend on the true value of the outcome. D5: No evidence of a pre-specified analysis plan. |
|  |  | All | Serious adverse events | Low | High | Low | Low | Low | High | D2: Analysis was restricted to N treated (i.e., not ITT or mITT). |
|  |  | All | Extent of use (proportion tests/days; EoT) | Low | High | High | Low | Low | High | D2: Analysis was restricted to N treated (i.e., not ITT or mITT).  D3: Some missing data and likely that it could depend on the true value of the outcome. |
| Brady 2002 | 1. Placebo 2. Carbamazepine | All | Adherence | Some concerns | High | High | Low | Some concerns | High | D1: Limited information regarding allocation process. D2: Analysis was restricted to N treated (i.e., not ITT or mITT).  D3: Some missing data and likely that it could depend on the true value of the outcome. D5: No evidence of a pre-specified analysis plan. |
|  |  | All | Extent of use (frequency; EoT) | Some concerns | High | High | Low | Some concerns | High | D1: Limited information regarding allocation process. Baseline demographics only reported for subset of randomised participants. D2: Analysis was restricted to N treated (i.e., not ITT or mITT).  D3: Some missing data and likely that it could depend on the true value of the outcome. D5: No evidence of a pre-specified analysis plan. |
|  |  | All | Craving | Some concerns | High | High | Low | Some concerns | High | D1: Limited information regarding allocation process. Baseline demographics only reported for subset of randomised participants. D2: Analysis was restricted to N treated (i.e., not ITT or mITT).  D3: Some missing data and likely that it could depend on the true value of the outcome. D5: No evidence of a pre-specified analysis plan. |
| Brodie 2009 | 1. Placebo 2. Vigabatrin | All | Dropout (any reason) | Some concerns | High | Low | Low | Low | High | D1: Limited information regarding randomization and allocation process. Baseline demographics only reported for subset of randomised participants. D2: Analysis was restricted to N treated (i.e., not ITT or mITT). |
|  |  | All | Continuous abstinence (treatment) | Some concerns | High | High | Low | Some concerns | High | D1: Limited information regarding randomization and allocation process. Baseline demographics only reported for subset of randomised participants. D2: Analysis was restricted to N treated (i.e., not ITT or mITT).  D3: Some missing data and likely that it could depend on the true value of the outcome. D5: No evidence of a pre-specified analysis plan. |
|  |  | All | Continuous abstinence (follow-up) | Some concerns | High | High | Low | Some concerns | High | D1: Limited information regarding randomization and allocation process. Baseline demographics only reported for subset of randomised participants. D2: Analysis was restricted to N treated (i.e., not ITT or mITT).  D3: Some missing data and likely that it could depend on the true value of the outcome. D5: No evidence of a pre-specified analysis plan. |
|  |  | All | Extent of use (proportion tests/days; EoT) | Some concerns | High | High | Low | Some concerns | High | D1: Limited information regarding randomization and allocation process. Baseline demographics only reported for subset of randomised participants. D2: Analysis was restricted to N treated (i.e., not ITT or mITT).  D3: Some missing data and likely that it could depend on the true value of the outcome. D5: No evidence of a pre-specified analysis plan. |
| Campbell 2003 | 1. Placebo 2. Desipramine 3. Carbamazepine | All | Dropout (any reason) | Some concerns | Low | Low | Low | Low | Some concerns | D1: Limited information regarding randomization and allocation process. |
|  |  | All | Continuous abstinence (treatment) | Some concerns | High | High | Low | Some concerns | High | D1: Limited information regarding randomization and allocation process. D2: Analysis was restricted to N treated (i.e., not ITT or mITT).  D3: Some missing data and likely that it could depend on the true value of the outcome. D5: No evidence of a pre-specified analysis plan. |
|  |  | All | Extent of use (frequency; EoT) | Some concerns | High | High | Low | Some concerns | High | D1: Limited information regarding randomization and allocation process. D2: Analysis was restricted to N treated (i.e., not ITT or mITT).  D3: Some missing data and likely that it could depend on the true value of the outcome. D5: No evidence of a pre-specified analysis plan. |
| Carpenter 2024 | 1. Placebo 2. MAS-ER | All | Dropout (any reason) | Low | Low | Low | Low | Low | Low |  |
|  |  | All | Serious adverse events | Low | Low | Low | Low | Low | Low |  |
|  |  | All | Continuous abstinence (treatment) | Low | Low | Some concerns | Low | Low | Some concerns | D3: Some missing data but unlikely that missingness depended on the true value of outcome. |
| Carroll 1994 | 1. Placebo 2. Desipramine | All | Longest duration of continuous abstinence | Some concerns | Some concerns | Low | Low | Some concerns | Some concerns | D1: Limited information regarding randomization and allocation process. Baseline demographics only reported for subset of randomised participants. D2: Appears that a per protocol analysis was used - but judged unlikely to have substantial impact on the results. D5: No evidence of a pre-specified analysis plan. |
|  |  | All | Extent of use (proportion tests/days; EoT) | Some concerns | Some concerns | Low | Low | Some concerns | Some concerns | D1: Limited information regarding randomization and allocation process. Baseline demographics only reported for subset of randomised participants. D2: Appears that a per protocol analysis was used - but judged unlikely to have substantial impact on the results. D5: No evidence of a pre-specified analysis plan. |
| Carroll 2004 | 1. Placebo 2. Disulfiram | All | Adherence | Low | Low | Low | Low | Some concerns | Some concerns | D5: No evidence of a pre-specified analysis plan. |
|  |  | All | Extent of use (frequency; EoT) | Low | Low | Low | Low | Some concerns | Some concerns | D5: No evidence of a pre-specified analysis plan. |
| Carroll 2012 | 1. Placebo 2. Disulfiram | All | Dropout (any reason) | Low | Low | Low | Low | Low | Low |  |
|  |  | All | Extent of use (frequency; EoT) | Low | Low | Low | Low | Some concerns | Some concerns | D5: No evidence of a pre-specified analysis plan. |
|  |  | All | Extent of use (frequency; follow-up) | Low | Low | Low | Low | Some concerns | Some concerns | D5: No evidence of a pre-specified analysis plan. |
| Carroll 2016 | 1. Placebo 2. Disulfiram | All | Dropout (any reason) | Low | Low | Low | Low | Low | Low |  |
|  |  | All | Adherence | Low | Low | Some concerns | Low | Some concerns | Some concerns | D3: Some missing data but unlikely that missingness depended on the true value of outcome. D5: No evidence of a pre-specified analysis plan. |
|  |  | All | Serious adverse events | Low | Low | Low | Low | Low | Low |  |
|  |  | All | Continuous abstinence (treatment) | Low | Low | Some concerns | Low | Some concerns | Some concerns | D3: Some missing data but unlikely that missingness depended on the true value of outcome. D5: No evidence of a pre-specified analysis plan. |
|  |  | All | Extent of use (proportion tests/days; EoT) | Low | Low | Some concerns | Low | Some concerns | Some concerns | D3: Some missing data but unlikely that missingness depended on the true value of outcome. D5: No evidence of a pre-specified analysis plan. |
|  |  | All | Extent of use (frequency; EoT) | Low | Low | Some concerns | Low | Some concerns | Some concerns | D3: Some missing data but unlikely that missingness depended on the true value of outcome. D5: No evidence of a pre-specified analysis plan. |
|  |  | All | Extent of use (frequency; follow-up) | Low | Low | Some concerns | Low | Some concerns | Some concerns | D3: Some missing data but unlikely that missingness depended on the true value of outcome. D5: No evidence of a pre-specified analysis plan. |
| Carroll 2018 | 1. Placebo 2. Galantamine | All | Dropout (any reason) | Low | Low | Low | Low | Low | Low |  |
|  |  | All | Adherence | Low | Low | Low | Low | Some concerns | Some concerns | D5: No evidence of a pre-specified analysis plan. |
|  |  | All | Extent of use (frequency; EoT) | Low | Low | Low | Low | Some concerns | Some concerns | D5: No evidence of a pre-specified analysis plan. |
| Ciraulo 2005a (study 1) | 1. Placebo 2. Paroxetine 3. Pentoxifylline 4. Riluzole | All | Dropout (any reason) | Some concerns | High | Low | Low | Low | High | D1: Limited information regarding randomization and allocation process. D2: Part of CREST protocol is modified blind, meaning participants and people responsible for intervention delivery likely unblinded; also unclear if denominator is N randomised or N treated. |
|  |  | 2 vs 1;  3 vs 1;  4 vs 1;  4 vs 2;  4 vs 3 | Extent of use (frequency; EoT) | Some concerns | High | Some concerns | Low | Some concerns | High | D1: Limited information regarding randomization and allocation process. D2: Part of CREST protocol is modified blind, meaning participants and people responsible for intervention delivery likely unblinded; also unclear if denominator is N randomised or N treated. D3: Some missing data but unlikely that missingness depended on the true value of outcome. D5: No evidence of a pre-specified analysis plan. |
|  |  | 3 vs 2 | Extent of use (frequency; EoT) | Some concerns | High | High | Low | Some concerns | High | D1: Limited information regarding randomization and allocation process. D2: Part of CREST protocol is modified blind, meaning participants and people responsible for intervention delivery likely unblinded; also unclear if denominator is N randomised or N treated. D3: Some missing data and likely that it could depend on the true value of the outcome. D5: No evidence of a pre-specified analysis plan. |
|  |  | 2 vs 1;  3 vs 1;  4 vs 1;  4 vs 2;  4 vs 3 | Craving | Some concerns | High | Some concerns | Some concerns | Some concerns | High | D1: Limited information regarding randomization and allocation process. D2: Part of CREST protocol is modified blind, meaning participants and people responsible for intervention delivery likely unblinded; also unclear if denominator is N randomised or N treated. D3: Some missing data but unlikely that missingness depended on the true value of outcome. D4: The outcome was self-reported and participants may be aware of their group allocation, but unlikely to be a substantial portion of participants. D5: No evidence of a pre-specified analysis plan. |
|  |  | 3 vs 2 | Craving | Some concerns | High | High | Some concerns | Some concerns | High | D1: Limited information regarding randomization and allocation process. D2: Part of CREST protocol is modified blind, meaning participants and people responsible for intervention delivery likely unblinded; also unclear if denominator is N randomised or N treated. D3: Some missing data and likely that it could depend on the true value of the outcome. D4: The outcome was self-reported and participants may be aware of their group allocation, but unlikely to be a substantial portion of participants. |
| Ciraulo 2005a (Study 2) | 1. Placebo 2. Pramipexole 3. Venlafaxine | All | Dropout (any reason) | Some concerns | High | Low | Low | Low | High | D1: Limited information regarding randomization and allocation process. D2: Part of CREST protocol is modified blind, meaning participants and people responsible for intervention delivery likely unblinded. |
|  |  | All | Extent of use (proportion tests/days; EoT)* | Some concerns | High | High | Low | Some concerns | High | D1: Limited information regarding allocation process. D2: Part of CREST protocol is modified blind, meaning participants and people responsible for intervention delivery likely unblinded. D3: Subset of the main trial. Unclear whether availability of data dependent on the true value.  D5: No evidence of a pre-specified analysis plan. |
|  |  | 2 vs 1 | Extent of use (frequency; EoT) | Some concerns | High | Some concerns | Some concerns | Some concerns | High | D1: Limited information regarding randomization and allocation process. D2: Part of CREST protocol is modified blind, meaning participants and people responsible for intervention delivery likely unblinded. D3: Some missing data but unlikely that missingness depended on the true value of outcome. D4: The outcome was self-reported and participants may be aware of their group allocation, but unlikely to be a substantial portion of participants. D5: No evidence of a pre-specified analysis plan. |
|  |  | 3 vs 1;  3 vs 2 | Extent of use (frequency; EoT) | Some concerns | High | High | Some concerns | Some concerns | High | D1: Limited information regarding randomization and allocation process. D2: Part of CREST protocol is modified blind, meaning participants and people responsible for intervention delivery likely unblinded. D3: Some missing data and likely that it could depend on the true value of the outcome. D4: The outcome was self-reported and participants may be aware of their group allocation, but unlikely to be a substantial portion of participants. D5: No evidence of a pre-specified analysis plan. |
|  |  | 2 vs 1 | Craving | Some concerns | High | Some concerns | Some concerns | Some concerns | High | D1: Limited information regarding randomization and allocation process. D2: Part of CREST protocol is modified blind, meaning participants and people responsible for intervention delivery likely unblinded. D3: Some missing data but unlikely that missingness depended on the true value of outcome. D4: The outcome was self-reported and participants may be aware of their group allocation, but unlikely to be a substantial portion of participants. D5: No evidence of a pre-specified analysis plan. |
|  |  | 3 vs 1;  3 vs 2 | Craving | Some concerns | High | High | Some concerns | Some concerns | High | D1: Limited information regarding randomization and allocation process. D2: Part of CREST protocol is modified blind, meaning participants and people responsible for intervention delivery likely unblinded. D3: Some missing data and likely that it could depend on the true value of the outcome. D4: The outcome was self-reported and participants may be aware of their group allocation, but unlikely to be a substantial portion of participants. D5: No evidence of a pre-specified analysis plan. |
| Cirualo 2005b | 1. Placebo 2. Nefazodone | All | Dropout (any reason) | Some concerns | Low | Low | Low | Low | Some concerns | D1: Limited information regarding randomization and allocation process. |
|  |  | All | Dropout (adverse events) | Some concerns | Low | High | Low | Low | High | D1: Limited information regarding randomization and allocation process. D3: Some missing data and likely that it could depend on the true value of the outcome. |
|  |  | All | Extent of use (BE level; EoT) | Some concerns | Low | High | Low | Some concerns | High | D1: Limited information regarding randomization and allocation process. D3: Some missing data and likely that it could depend on the true value of the outcome. D5: No evidence of a pre-specified analysis plan. |
|  |  | All | Extent of use (frequency; EoT) | Some concerns | Low | High | Low | Some concerns | High | D1: Limited information regarding randomization and allocation process. D3: Some missing data and likely that it could depend on the true value of the outcome. D5: No evidence of a pre-specified analysis plan. |
|  |  | All | Craving | Some concerns | Low | High | Low | Some concerns | High | D1: Limited information regarding randomization and allocation process. D3: Some missing data and likely that it could depend on the true value of the outcome. D5: No evidence of a pre-specified analysis plan. |
| Cornish 1995 | 1. Placebo 2. Carbamazepine | All | Dropout (any reason) | Some concerns | High | Low | Low | Low | High | D1: Limited information regarding randomization and allocation process. Baseline demographics only reported for subset of randomised participants. D2: Appears that a per protocol analysis was used (e.g., exclusions due to non-compliance). |
|  |  | All | Serious adverse events | Some concerns | High | Low | Low | Low | High | D1: Limited information regarding randomization and allocation process. Baseline demographics only reported for subset of randomised participants. D2: Appears that a per protocol analysis was used (e.g., exclusions due to non-compliance). |
|  |  | All | Mortality | Some concerns | High | Low | Low | Low | High | D1: Limited information regarding randomization and allocation process. Baseline demographics only reported for subset of randomised participants. D2: Appears that a per protocol analysis was used (e.g., exclusions due to non-compliance). |
|  |  | All | Extent of use (BE level; EoT) | Some concerns | High | High | Low | Some concerns | High | D1: Limited information regarding randomization and allocation process. Baseline demographics only reported for subset of randomised participants. D2: Appears that a per protocol analysis was used (e.g., exclusions due to non-compliance).  D3: Some missing data and likely that it could depend on the true value of the outcome. D5: No evidence of a pre-specified analysis plan. |
|  |  | All | Extent of use (proportion tests/days; EoT) | Some concerns | High | High | Low | Some concerns | High | D1: Limited information regarding randomization and allocation process. Baseline demographics only reported for subset of randomised participants. D2: Appears that a per protocol analysis was used (e.g., exclusions due to non-compliance).  D3: Some missing data and likely that it could depend on the true value of the outcome. D5: No evidence of a pre-specified analysis plan. |
| Cornish 2001 | 1. Placebo 2. Ritanserin | All | Dropout (any reason) | Some concerns | Low | Low | Low | Low | Some concerns | D1: Limited information regarding allocation process. |
|  |  | All | Adherence* | Some concerns | Low | Some concerns | Low | Some concerns | Some concerns | D1: Limited information regarding allocation process. D3: Some missing data but unlikely that missingness depended on the true value of outcome. D5: No evidence of a pre-specified analysis plan. |
|  |  | All | Extent of use (proportion tests/days; EoT) | Some concerns | Low | High | Low | Some concerns | High | D1: Limited information regarding allocation process. D3: Some missing data and likely that it could depend on the true value of the outcome. D5: No evidence of a pre-specified analysis plan. |
|  |  | All | Craving | Some concerns | Low | Some concerns | Low | Some concerns | Some concerns | D1: Limited information regarding allocation process. D3: Some missing data but unlikely that missingness depended on the true value of outcome. D5: No evidence of a pre-specified analysis plan. |
| Dackis 2005 | 1. Placebo 2. Modafinil | All | Dropout (any reason) | Low | Low | Low | Low | Low | Low |  |
|  |  | All | Continuous abstinence (treatment) | Low | Low | Some concerns | Low | Some concerns | Some concerns | D3: Some missing data but unlikely that missingness depended on the true value of outcome. D5: No evidence of a pre-specified analysis plan. |
|  |  | All | Extent of use (proportion tests/days; EoT) | Low | Low | Some concerns | Low | Some concerns | Some concerns | D3: Some missing data but unlikely that missingness depended on the true value of outcome. D5: No evidence of a pre-specified analysis plan. |
| Dackis 2012 | 1. Placebo 2. Modafinil (200 mg/day) 3. Modafinil (400 mg/day) | All | Dropout (any reason) | Low | Low | Low | Low | Low | Low |  |
|  |  | 2 vs 1;  3 vs 1 | Continuous abstinence (treatment) | Low | Low | High | Low | Some concerns | High | D3: Some missing data and likely that it could depend on the true value of the outcome. D5: No evidence of a pre-specified analysis plan. |
|  |  | 3 vs 2 | Continuous abstinence (treatment) | Low | Low | Some concerns | Low | Some concerns | Some concerns | D3: Some missing data but unlikely that missingness depended on the true value of outcome. D5: No evidence of a pre-specified analysis plan. |
|  |  | 2 vs 1;  3 vs 1 | Point abstinence (EoT) | Low | Low | High | Low | Some concerns | High | D3: Some missing data and likely that it could depend on the true value of the outcome. D5: No evidence of a pre-specified analysis plan. |
|  |  | 3 vs 2 | Point abstinence (EoT) | Low | Low | Some concerns | Low | Some concerns | Some concerns | D3: Some missing data but unlikely that missingness depended on the true value of outcome. D5: No evidence of a pre-specified analysis plan. |
| Dakwar 2019 | 1. Midazolam 2. Ketamine | All | Dropout (any reason) | Low | Some concerns | Low | Low | Low | Some concerns | D2: Potential issues with blinding of participants due to effects of drug. |
|  |  | All | Continuous abstinence (follow-up) | Low | Some concerns | High | Low | Low | High | D2: Potential issues with blinding of participants due to effects of drug. D3: Some missing data and likely that it could depend on the true value of the outcome. |
| DeVito 2019 | 1. Placebo 2. Galantamine (8mg/day) 3. Galantamine (16mg/day) | All | Dropout (any reason) | Low | Low | Low | Low | Low | Low |  |
|  |  | 2 vs 1 | Adherence | Low | Low | Some concerns | Low | Some concerns | Some concerns | D3: Some missing data but unlikely that missingness depended on the true value of outcome. D5: No evidence of a pre-specified analysis plan. |
|  |  | 3 vs 1;  3 vs 2 | Adherence | Low | Low | High | Low | Some concerns | High | D3: Some missing data and likely that it could depend on the true value of the outcome. D5: No evidence of a pre-specified analysis plan. |
|  |  | 2 vs 1 | Longest duration of continuous abstinence | Low | Low | Some concerns | Low | Some concerns | Some concerns | D3: Some missing data but unlikely that missingness depended on the true value of outcome. D5: No evidence of a pre-specified analysis plan. |
|  |  | 3 vs 1;  3 vs 2 | Longest duration of continuous abstinence | Low | Low | High | Low | Some concerns | High | D3: Some missing data and likely that it could depend on the true value of the outcome. D5: No evidence of a pre-specified analysis plan. |
|  |  | 2 vs 1 | Extent of use (frequency; EoT) | Low | Low | Some concerns | Low | Some concerns | Some concerns | D3: Some missing data but unlikely that missingness depended on the true value of outcome. D5: No evidence of a pre-specified analysis plan. |
|  |  | 3 vs 1;  3 vs 2 | Extent of use (frequency; EoT) | Low | Low | High | Low | Some concerns | High | D3: Some missing data and likely that it could depend on the true value of the outcome. D5: No evidence of a pre-specified analysis plan. |
| DeVito 2022 | 1. Placebo 2. Modafinil | All | Dropout (any reason) | Low | Low | Low | Low | Low | Low |  |
|  |  | All | Dropout (adverse events) | Low | Low | High | Low | Low | High | D3: Some missing data and likely that it could depend on the true value of the outcome. |
|  |  | All | Adherence | Low | Low | Low | Low | Some concerns | Some concerns | D5: No evidence of a pre-specified analysis plan. |
|  |  | All | Longest duration of continuous abstinence | Low | Low | High | Low | Some concerns | High | D3: Some missing data and likely that it could depend on the true value of the outcome. D5: No evidence of a pre-specified analysis plan. |
|  |  | All | Extent of use (proportion tests/days; EoT) | Low | Low | Low | Low | Some concerns | Some concerns | D5: No evidence of a pre-specified analysis plan. |
|  |  | All | Extent of use (frequency; EoT) | Low | Low | Low | Low | Some concerns | Some concerns | D5: No evidence of a pre-specified analysis plan. |
| Dieckmann 2014 | 1. Placebo 2. Biperiden | All | Dropout due to any reason | Low | Low | Low | Low | Low | Low |  |
|  |  | All | Extent of use (frequency; EoT) | Low | Low | Low | Low | High | High | D5: No evidence of a pre-specified analysis plan. The registration history suggests that the result could have been selected from multiple eligible outcome measurements. |
|  |  | All | Extent of use (quantity; EoT) | Low | Low | Low | Low | High | High | D5: No evidence of a pre-specified analysis plan. The registration history suggests that the result could have been selected from multiple eligible outcome measurements. |
|  |  | All | Craving | Low | Low | Low | Low | High | High | D5: No evidence of a pre-specified analysis plan. The registration history suggests that the result could have been selected from multiple eligible outcome measurements. |
| Dursteler-Macfarland 2013 | 1. Placebo 2. Methylphenidate | All | Dropout (any reason) | Low | Low | Low | Low | Low | Low |  |
|  |  | All | Continuous abstinence (treatment) | Low | Low | High | Low | Some concerns | High | D3: Some missing data and likely that it could depend on the true value of the outcome. D5: No evidence of a pre-specified analysis plan. |
|  |  | All | Extent of use (frequency; EoT) | Low | Low | High | Low | Some concerns | High | D3: Some missing data and likely that it could depend on the true value of the outcome. D5: No evidence of a pre-specified analysis plan. |
|  |  | All | Extent of use (quantity; EoT) | Low | Low | High | Low | Some concerns | High | D3: Some missing data and likely that it could depend on the true value of the outcome. D5: No evidence of a pre-specified analysis plan. |
| Elkashef 2006 | 1. Placebo 2. Selegiline Transdermal System | All | Dropout (any reason) | Low | Low | Low | Low | Low | Low |  |
|  |  | All | Serious adverse events | Low | Low | Low | Low | High | High | D5: Only medication-related SAEs reported. More SAEs were recorded but this result is not reported by intervention arm. |
|  |  | All | Continuous abstinence (treatment) | Low | Low | Some concerns | Low | Some concerns | Some concerns | D3: Some missing data but unlikely that missingness depended on the true value of outcome. D5: No evidence of a pre-specified analysis plan. |
|  |  | All | Longest duration of continuous abstinence | Low | Low | Some concerns | Low | Some concerns | Some concerns | D3: Some missing data but unlikely that missingness depended on the true value of outcome. D5: No evidence of a pre-specified analysis plan. |
|  |  | All | Extent of use (proportion tests/days; EoT) | Low | Low | Some concerns | Low | Some concerns | Some concerns | D3: Some missing data but unlikely that missingness depended on the true value of outcome. D5: No evidence of a pre-specified analysis plan. |
|  |  | All | Craving | Low | Low | Some concerns | Low | Some concerns | Some concerns | D3: Some missing data but unlikely that missingness depended on the true value of outcome. D5: No evidence of a pre-specified analysis plan. |
| Gawin 1989 | 1. Placebo 2. Lithium carbonate 3. Desipramine | All | Dropout (any reason) | High | Low | Low | Low | Low | High | D1: Concerns over the allocation process to support reaching balanced group sizes across the different interventions; Baseline demographics only reported for subset of randomised participants. |
|  |  | 2 vs 1 | Continuous abstinence (treatment) | High | High | Some concerns | Low | Some concerns | High | D1: Concerns over the allocation process to support reaching balanced group sizes across the different interventions; Baseline demographics only reported for subset of randomised participants. D2: Analysis was restricted to N treated (i.e., not ITT or mITT).  D3: Some missing data but unlikely that missingness depended on the true value of outcome. D5: No evidence of a pre-specified analysis plan. |
|  |  | 3 vs 1;  3 vs 2 | Continuous abstinence (treatment) | High | High | High | Low | Some concerns | High | D1: Concerns over the allocation process to support reaching balanced group sizes across the different interventions; Baseline demographics only reported for subset of randomised participants. D2: Analysis was restricted to N treated (i.e., not ITT or mITT).  D3: Some missing data and likely that it could depend on the true value of the outcome. D5: No evidence of a pre-specified analysis plan. |
|  |  | 2 vs 1;  3 vs 1 | Continuous abstinence (follow-up)* | High | High | High | Low | Some concerns | High | D1: Concerns over the allocation process to support reaching balanced group sizes across the different interventions; Baseline demographics only reported for subset of randomised participants. D2: Analysis was restricted to N treated (i.e., not ITT or mITT).  D3: Some missing data and likely that it could depend on the true value of the outcome. D5: No evidence of a pre-specified analysis plan. |
|  |  | 3 vs 2 | Continuous abstinence (follow-up)* | High | High | Some concerns | Low | Some concerns | High | D1: Concerns over the allocation process to support reaching balanced group sizes across the different interventions; Baseline demographics only reported for subset of randomised participants. D2: Analysis was restricted to N treated (i.e., not ITT or mITT).  D3: Some missing data but unlikely that missingness depended on the true value of outcome. D5: No evidence of a pre-specified analysis plan. |
|  |  | 2 vs 1 | Extent of use (quantity; EoT) | High | High | Some concerns | Low | Some concerns | High | D1: Concerns over the allocation process to support reaching balanced group sizes across the different interventions; Baseline demographics only reported for subset of randomised participants. D2: Analysis was restricted to N treated (i.e., not ITT or mITT).  D3: Some missing data but unlikely that missingness depended on the true value of outcome. D5: No evidence of a pre-specified analysis plan. |
|  |  | 3 vs 1;  3 vs 2 | Extent of use (quantity; EoT) | High | High | High | Low | Some concerns | High | D1: Concerns over the allocation process to support reaching balanced group sizes across the different interventions; Baseline demographics only reported for subset of randomised participants. D2: Analysis was restricted to N treated (i.e., not ITT or mITT).  D3: Some missing data and likely that it could depend on the true value of the outcome. D5: No evidence of a pre-specified analysis plan. |
|  |  | 2 vs 1;  3 vs 1 | Extent of use (quantity; follow-up)* | High | High | High | Low | Some concerns | High | D1: Concerns over the allocation process to support reaching balanced group sizes across the different interventions; Baseline demographics only reported for subset of randomised participants. D2: Analysis was restricted to N treated (i.e., not ITT or mITT).  D3: Some missing data and likely that it could depend on the true value of the outcome. D5: No evidence of a pre-specified analysis plan. |
|  |  | 3 vs 2 | Extent of use (quantity; follow-up)* | High | High | Some concerns | Low | Some concerns | High | D1: Concerns over the allocation process to support reaching balanced group sizes across the different interventions; Baseline demographics only reported for subset of randomised participants. D2: Analysis was restricted to N treated (i.e., not ITT or mITT).  D3: Some missing data but unlikely that missingness depended on the true value of outcome. D5: No evidence of a pre-specified analysis plan. |
|  |  | 2 vs 1 | Craving | High | High | Some concerns | Low | Some concerns | High | D1: Concerns over the allocation process to support reaching balanced group sizes across the different interventions; Baseline demographics only reported for subset of randomised participants. D2: Analysis was restricted to N treated (i.e., not ITT or mITT).  D3: Some missing data but unlikely that missingness depended on the true value of outcome. D5: No evidence of a pre-specified analysis plan. |
|  |  | 3 vs 1;  3 vs 2 | Craving | High | High | High | Low | Some concerns | High | D1: Concerns over the allocation process to support reaching balanced group sizes across the different interventions; Baseline demographics only reported for subset of randomised participants. D2: Analysis was restricted to N treated (i.e., not ITT or mITT).  D3: Some missing data and likely that it could depend on the true value of the outcome. D5: No evidence of a pre-specified analysis plan. |
| George 2000 | 1. Placebo 2. Disulfiram | All | Dropout (any reason) | Some concerns | Low | Low | Low | Low | Some concerns | D1: Limited information regarding allocation process. |
|  |  | All | Continuous abstinence (treatment) | Some concerns | Low | Some concerns | Low | Some concerns | Some concerns | D1: Limited information regarding allocation process. D3: Some missing data but unlikely that missingness depended on the true value of outcome. D5: No evidence of a pre-specified analysis plan. |
|  |  | All | Extent of use (proportion tests/days; EoT) | Some concerns | Low | Some concerns | Low | Some concerns | Some concerns | D1: Limited information regarding allocation process. D3: Some missing data but unlikely that missingness depended on the true value of outcome. D5: No evidence of a pre-specified analysis plan. |
|  |  | All | Extent of use (quantity; EoT) | Some concerns | Low | Some concerns | Low | Some concerns | Some concerns | D1: Limited information regarding allocation process. D3: Some missing data but unlikely that missingness depended on the true value of outcome. D5: No evidence of a pre-specified analysis plan. |
| Gilgun-Sherki 2016 | 1. Placebo 2. TV-1380 (300 mg) 3. TV-1380 (150 mg) | All | Dropout (any reason) | Low | Low | Low | Low | Low | Low |  |
|  |  | All | Dropout (adverse events) | Low | Low | Some concerns | Low | Low | Some concerns | D3: Some missing data but unlikely that missingness depended on the true value of outcome. |
|  |  | All | Serious adverse events | Low | Some concerns | Low | Low | Low | Some concerns | D2: Analysis restricted to those receiving first dose - but judged unlikely to have substantial impact on the results. |
|  |  | All | Continuous abstinence (treatment) | Low | Some concerns | Some concerns | Low | Low | Some concerns | D2: Analysis restricted to those receiving first dose - but judged unlikely to have substantial impact on the results. D3: Some missing data but unlikely that missingness depended on the true value of outcome. |
| Gonzalez 2003 | 1. Placebo 2. Tiagabine (6mg) 3. Tiagabine (12mg) | All | Dropout (any reason) | Some concerns | Low | Low | Low | Low | Some concerns | D1: Limited information regarding allocation process. |
|  |  | 2 vs 1 | Extent of use (proportion tests/days; EoT) | Some concerns | Low | High | Low | Some concerns | High | D1: Limited information regarding allocation process. D3: Some missing data and likely that it could depend on the true value of the outcome. |
|  |  | 3 vs 1;  3 vs 2 | Extent of use (proportion tests/days; EoT) | Some concerns | Low | Some concerns | Low | Some concerns | Some concerns | D1: Limited information regarding allocation process. D3: Some missing data but unlikely that missingness depended on the true value of outcome. |
|  |  | 2 vs 1 | Extent of use (frequency; EoT) | Some concerns | Low | High | Low | Some concerns | High | D1: Limited information regarding allocation process. D3: Some missing data and likely that it could depend on the true value of the outcome. |
|  |  | 3 vs 1;  3 vs 2 | Extent of use (frequency; EoT) | Some concerns | Low | Some concerns | Low | Some concerns | Some concerns | D1: Limited information regarding allocation process. D3: Some missing data but unlikely that missingness depended on the true value of outcome. |
| Gorelick 2006 | 1. Placebo 2. Bromocriptine | All | Dropout (any reason) | Low | Low | Low | Low | Low | Low |  |
|  |  | All | Dropout (adverse events) | Low | Low | High | Low | Low | High | D3: Some missing data and likely that it could depend on the true value of the outcome. |
|  |  | All | Adherence | Low | Low | High | Low | Some concerns | High | D3: Some missing data and likely that it could depend on the true value of the outcome. D5: No evidence of a pre-specified analysis plan. |
| Grabowski 1995 (study 1) | 1. Placebo 2. Fluoxetine (20mg) 3. Fluoxetine (40mg) | All | Dropout (any reason) | Some concerns | High | Low | Low | Low | High | D1: Limited information regarding allocation process. Baseline demographics only reported for subset of randomised participants. D2: Analysis was restricted to N treated (i.e., not ITT or mITT). |
| Grabowski 1995 (study 2) | 1. Placebo 2. Fluoxetine | All | Dropout (any reason) | Some concerns | Low | Low | Low | Low | Some concerns | D1: Limited information regarding allocation process. |
| Grabowski 1997 | 1. Placebo 2. Methylphenidate | All | Dropout (any reason) | Some concerns | Low | Low | Low | Low | Some concerns | D1: Limited information regarding allocation process. |
| Grabowski 2001 | 1. Placebo 2. Dextroamphetamine (30mg) 3. Dextroamphetamine (60mg) | All | Adherence | Some concerns | Low | High | Low | Some concerns | High | D1: Limited information regarding allocation process. D3: Some missing data and likely that it could depend on the true value of the outcome. D5: No evidence of a pre-specified analysis plan. |
| Grabowski 2004 (study 1) | 1. Placebo 2. Dextroamphetamine (30mg) 3. Dextroamphetamine (60mg) | All | Dropout (any reason) | Some concerns | Low | Low | Low | Low | Some concerns | D1: Limited information regarding allocation process. |
|  |  | 2 vs 1;  3 vs 1 | Adherence | Some concerns | High | High | Low | Some concerns | High | D1: Limited information regarding allocation process. D2: Analysis was restricted to N treated (i.e., not ITT or mITT).  D3: Some missing data and likely that it could depend on the true value of outcome. D5: No evidence of a pre-specified analysis plan. |
|  |  | 3 vs 2 | Adherence | Some concerns | High | Some concerns | Low | Some concerns | High | D1: Limited information regarding allocation process. D2: Analysis was restricted to N treated (i.e., not ITT or mITT).  D3: Some missing data but unlikely that missingness depended on the true value of outcome. D5: No evidence of a pre-specified analysis plan. |
| Grabowski 2004 (study 2) | 1. Placebo 2. Risperidone (2mg) 3. Risperidone (4mg) | All | Dropout (any reason) | Some concerns | Low | Low | Low | Low | Some concerns | D1: Limited information regarding allocation process. |
|  |  | All | Adherence | Some concerns | Low | High | Low | Some concerns | High | D1: Limited information regarding allocation process. D3: Some missing data and likely that it could depend on the true value of the outcome. D5: No evidence of a pre-specified analysis plan. |
| Halikas 1997 | 1. Placebo 2. Carbamazepine (800mg) 3. Carbamazepine (400mg) | All | Dropout (any reason) | High | Low | Low | Low | Low | High | D1: Concerns over the allocation process as early dropouts were replaced; Baseline demographics not reported by randomised group. |
|  |  | All | Craving | High | High | High | Low | Some concerns | High | D1: Concerns over the allocation process as early dropouts were replaced; Baseline demographics not reported by randomised group. D2: Analysis was restricted to N treated (i.e., not ITT or mITT).  D3: Some missing data and likely that it could depend on the true value of the outcome. D5: No evidence of a pre-specified analysis plan. |
| Hall 1994 | 1. Placebo 2. Desipramine | All | Point abstinence (EoT) | Some concerns | Low | Low | Low | Some concerns | Some concerns | D1: Limited information regarding randomization and allocation process. D5: No evidence of a pre-specified analysis plan. |
|  |  | All | Point abstinence (follow-up) | Some concerns | Low | Low | Low | Some concerns | Some concerns | D1: Limited information regarding randomization and allocation process. D5: No evidence of a pre-specified analysis plan. |
| Hamilton 2009 | 1. Placebo 2. Olanzapine | All | Adherence | Low | Low | High | Low | Some concerns | High | D3: Some missing data. Amount and difference between groups unclear. D5: No evidence of a pre-specified analysis plan. |
|  |  | All | Continuous abstinence (treatment) | Low | Low | High | Low | Some concerns | High | D3: Some missing data and likely that it could depend on the true value of the outcome. D5: No evidence of a pre-specified analysis plan. |
|  |  | All | Point abstinence (EoT) | Low | Low | High | Low | Some concerns | High | D3: Some missing data and likely that it could depend on the true value of the outcome. D5: No evidence of a pre-specified analysis plan. |
|  |  | All | Point abstinence (follow-up) | Low | Low | High | Low | Some concerns | High | D3: Some missing data and likely that it could depend on the true value of the outcome. D5: No evidence of a pre-specified analysis plan. |
| Handelsman 1995 | 1. Placebo 2. Amantadine (200mg) 3. Amantadine (400mg) | All | Dropout (any reason) | Low | Low | Low | Low | Low | Low |  |
|  |  | All | Extent of use (proportion tests/days; EoT) | Low | Low | Some concerns | Low | Some concerns | Some concerns | D3: Some missing data but unlikely that missingness depended on the true value of outcome. D5: No evidence of a pre-specified analysis plan. |
|  |  | All | Extent of use (quantity; EoT) | Low | Low | Some concerns | Low | Some concerns | Some concerns | D3: Some missing data but unlikely that missingness depended on the true value of outcome. D5: No evidence of a pre-specified analysis plan. |
|  |  | All | Craving | Low | Low | Some concerns | Low | Some concerns | Some concerns | D3: Some missing data but unlikely that missingness depended on the true value of outcome. D5: No evidence of a pre-specified analysis plan. |
| Handelsman 1997 | 1. Placebo 2. Bromocriptine | All | Dropout (any reason) | Some concerns | High | Low | Low | Low | High | D1: Limited information regarding randomisation and allocation process. Baseline demographics only reported for subset of randomised participants. D2: Unclear whether sample included in analysis is N randomised or N treated. |
|  |  | All | Extent of use (frequency; EoT) | Some concerns | High | Some concerns | Low | Some concerns | High | D1: Limited information regarding allocation process. Baseline demographics only reported for subset of randomised participants. D2: Unclear whether analysis includes N randomised or N treated. D3: Some missing data but unlikely that missingness depended on the true value of outcome. D5: No evidence of a pre-specified analysis plan. |
|  |  | All | Craving | Some concerns | High | Some concerns | Low | Some concerns | High | D1: Limited information regarding allocation process. Baseline demographics only reported for subset of randomised participants. D2: Unclear whether analysis includes N randomised or N treated. D3: Some missing data but unlikely that missingness depended on the true value of outcome. D5: No evidence of a pre-specified analysis plan. |
| Hersh 1998 | 1. Placebo 2. Naltrexone | All | Dropout (any reason) | Some concerns | Low | Low | Low | Low | Some concerns | D1: Limited information regarding randomization and allocation process. |
|  |  | All | Dropout (adverse events) | Some concerns | Low | Some concerns | Low | Low | Some concerns | D1: Limited information regarding randomization and allocation process. D3: Some missing data but unlikely that missingness depended on the true value of outcome. |
|  |  | All | Adherence | Some concerns | Low | Some concerns | Low | Some concerns | Some concerns | D1: Limited information regarding randomization and allocation process. D3: Some missing data but unlikely that missingness depended on the true value of outcome. D5: No evidence of a pre-specified analysis plan. |
|  |  | All | Extent of use (frequency; EoT) | Some concerns | Low | Some concerns | Low | Some concerns | Some concerns | D1: Limited information regarding randomization and allocation process. D3: Some missing data but unlikely that missingness depended on the true value of outcome. D5: No evidence of a pre-specified analysis plan. |
|  |  | All | Extent of use (quantity; EoT) | Some concerns | Low | Some concerns | Low | Some concerns | Some concerns | D1: Limited information regarding randomization and allocation process. D3: Some missing data but unlikely that missingness depended on the true value of outcome. D5: No evidence of a pre-specified analysis plan. |
| Johnson 1997 | 1. Placebo 2. Ritanserin | All | Dropout (any reason) | Some concerns | Low | Low | Low | Low | Some concerns | D1: Limited information regarding randomization and allocation process. |
|  |  | All | Adherence | Some concerns | High | Some concerns | Low | Low | High | D1: Limited information regarding randomization and allocation process. D2: Appears that a per protocol analysis was used (e.g., exclusions due to non-compliance) D3: Some missing data but unlikely that missingness depended on the true value of outcome. D5: No evidence of a pre-specified analysis plan. |
|  |  | All | Craving | Some concerns | High | Some concerns | Low | Some concerns | High | D1: Limited information regarding randomization and allocation process. D2: Appears that a per protocol analysis was used (e.g., exclusions due to non-compliance) D3: Some missing data but unlikely that missingness depended on the true value of outcome. D5: No evidence of a pre-specified analysis plan. |
| Johnson 2006 | 1. Placebo 2. Ondansetron (0.25mg) 3. Ondansetron (1mg) 4. Ondansetron (4mg) | All | Adherence | Low | Low | High | Low | Some concerns | High | D3: Some missing data. Amount and difference between groups unclear. D5: No evidence of a pre-specified analysis plan. |
|  |  | All | Point abstinence (EoT) | Low | Low | High | Low | Some concerns | High | D3: Some missing data. Amount and difference between groups unclear. D5: No evidence of a pre-specified analysis plan. |
| Johnson 2013 | 1. Placebo 2. Topiramate | All | Dropout (any reason) | Low | Low | Low | Low | Low | Low |  |
|  |  | All | Adherence | Low | Low | High | Low | Some concerns | High | D3: Some missing data and likely that it could depend on the true value of the outcome. D5: No evidence of a pre-specified analysis plan. |
|  |  | All | Extent of use (proportion tests/days; EoT) | Low | Low | High | Low | Some concerns | High | D3: Some missing data and likely that it could depend on the true value of the outcome. D5: No evidence of a pre-specified analysis plan. |
| Johnson 2020 | 1. Placebo 2. D-cycloserine | All | Dropout (any reason) | Low | Low | Low | Low | Low | Low |  |
|  |  | All | Continuous abstinence (follow-up) | Low | High | Some concerns | Low | High | High | D2: Appears that a per protocol analysis was used (e.g., exclusions due to non-compliance).  D3: Some missing data but unlikely that missingness depended on the true value of outcome. D5: Discrepancy between registry and report over outcome measure used. |
|  |  | All | Point abstinence (follow-up) | Low | High | Some concerns | Low | Low | High | D2: Appears that a per protocol analysis was used (e.g., exclusions due to non-compliance).  D3: Some missing data but unlikely that missingness depended on the true value of outcome. |
|  |  | All | Extent of use (proportion tests/days; EoT) | Low | High | Some concerns | Low | Some concerns | High | D2: Appears that a per protocol analysis was used (e.g., exclusions due to non-compliance).  D3: Some missing data but unlikely that missingness depended on the true value of outcome. D5: No evidence of a pre-specified analysis plan. |
| Jones 2004 | 1. Placebo 2. Tryptophan | All | Dropout (any reason) | Some concerns | Low | Low | Low | Some concerns | Some concerns | D1: Limited information regarding allocation process. D5: No evidence of a pre-specified analysis plan. |
|  |  | All | Longest duration of continuous abstinence | Some concerns | High | High | Low | Some concerns | High | D1: Limited information regarding allocation process. D2: Analysis was restricted to N treated (i.e., not ITT or mITT).  D3: Some missing data and likely that it could depend on the true value of the outcome. D5: No evidence of a pre-specified analysis plan. |
|  |  | All | Extent of use (proportion tests/days; EoT) | Some concerns | High | High | Low | Some concerns | High | D1: Limited information regarding allocation process. D2: Analysis was restricted to N treated (i.e., not ITT or mITT).  D3: Some missing data and likely that it could depend on the true value of the outcome. D5: No evidence of a pre-specified analysis plan. |
|  |  | All | Extent of use (frequency; EoT) | Some concerns | High | High | Low | Some concerns | High | D1: Limited information regarding allocation process. D2: Analysis was restricted to N treated (i.e., not ITT or mITT).  D3: Some missing data and likely that it could depend on the true value of the outcome. D5: No evidence of a pre-specified analysis plan. |
| Junior 2024 | 1. Placebo 2. Biperiden | All | Dropout (any reason) | Low | Low | Low | Low | Low | Low |  |
| Kablinger 2012 | 1. Placebo 2. Metyrapone + oxazepam (500/20mg) 3. Metyrapone + oxazepam (1500/20mg) | All | Dropout (any reason) | High | Low | Low | Low | Low | High | D1: Inappropriate randomization method and unlikely allocation concealment. |
|  |  | All | Dropout (adverse events) | High | Low | Some concerns | Low | Low | High | D1: Inappropriate randomization method and unlikely allocation concealment.  D3: Some missing data but unlikely that missingness depended on the true value of outcome. |
|  |  | All | Craving | High | High | High | Low | Some concerns | High | D1: Inappropriate randomization method and unlikely allocation concealment.  D2: Appears that a per protocol analysis was used (e.g., exclusions due to non-compliance). D3: Some missing data and likely that it could depend on the true value of the outcome.  D5: No evidence of a pre-specified analysis plan. |
| Kahn 2009 | 1. Placebo 2. Baclofen | All | Dropout (any reason) | Some concerns | Low | Low | Low | Low | Some concerns | D1: Limited information regarding allocation process. |
|  |  | All | Serious adverse events | Some concerns | Low | Low | Low | High | High | D1: Limited information regarding allocation process. D5: No evidence of a pre-specified analysis plan. The result could have been selected from multiple eligible outcome measurements. |
|  |  | All | Longest duration of continuous abstinence | Some concerns | Low | Some concerns | Low | Some concerns | Some concerns | D1: Limited information regarding allocation process. D3: Some missing data but unlikely that missingness depended on the true value of outcome.  D5: No evidence of a pre-specified analysis plan. |
|  |  | All | Extent of use (proportion tests/days; EoT) | Some concerns | Low | Some concerns | Low | Some concerns | Some concerns | D1: Limited information regarding allocation process. D3: Some missing data but unlikely that missingness depended on the true value of outcome.  D5: No evidence of a pre-specified analysis plan. |
| Kampman 1996 | 1. Placebo 2. Amantadine | All | Dropout (any reason) | Some concerns | Low | Low | Low | Low | Some concerns | D1: Limited information regarding allocation process. |
|  |  | All | Continuous abstinence (treatment) | Some concerns | Low | Some concerns | Low | Some concerns | Some concerns | D1: Limited information regarding allocation process. D3: Some missing data but unlikely that missingness depended on the true value of outcome.  D5: No evidence of a pre-specified analysis plan. |
|  |  | All | Extent of use (BE level; EoT) | Some concerns | Low | Some concerns | Low | Some concerns | Some concerns | D1: Limited information regarding allocation process. D3: Some missing data but unlikely that missingness depended on the true value of outcome.  D5: No evidence of a pre-specified analysis plan. |
|  |  | All | Extent of use (frequency; EoT) | Some concerns | Low | Some concerns | Low | Some concerns | Some concerns | D1: Limited information regarding allocation process. D3: Some missing data but unlikely that missingness depended on the true value of outcome.  D5: No evidence of a pre-specified analysis plan. |
|  |  | All | Extent of use (frequency; follow-up) | Some concerns | Low | Some concerns | Low | Some concerns | Some concerns | D1: Limited information regarding allocation process. D3: Some missing data but unlikely that missingness depended on the true value of outcome.  D5: No evidence of a pre-specified analysis plan. |
|  |  | All | Extent of use (quantity; EoT) | Some concerns | Low | High | Low | Some concerns | High | D1: Limited information regarding allocation process. D3: Some missing data and likely that it could depend on the true value of the outcome.  D5: No evidence of a pre-specified analysis plan. |
|  |  | All | Extent of use (quantity; follow-up) | Some concerns | Low | High | Low | Some concerns | High | D1: Limited information regarding allocation process. D3: Some missing data and likely that it could depend on the true value of the outcome.  D5: No evidence of a pre-specified analysis plan. |
| Kampman 2001 | 1. Placebo 2. Propranolol | All | Dropout (any reason) | Low | Low | Low | Low | Low | Low |  |
|  |  | All | Adherence | Low | Low | Some concerns | Low | Some concerns | Some concerns | D3: Some missing data but unlikely that missingness depended on the true value of outcome.  D5: No evidence of a pre-specified analysis plan. |
| Kampman 2003a | 1. Placebo 2. Olanzapine | All | Dropout (any reason) | Low | Low | Low | Low | Low | Low |  |
|  |  | All | Adherence | Low | Low | Low | Low | Some concerns | Some concerns | D5: No evidence of a pre-specified analysis plan. |
|  |  | All | Extent of use (frequency; EoT) | Low | Low | Low | Low | Some concerns | Some concerns | D5: No evidence of a pre-specified analysis plan. |
|  |  | All | Extent of use (quantity; EoT) | Low | Low | Low | Low | Some concerns | Some concerns | D5: No evidence of a pre-specified analysis plan. |
|  |  | All | Craving | Low | Low | Low | Low | Some concerns | Some concerns | D5: No evidence of a pre-specified analysis plan. |
| Kampman 2003b | 1. Placebo 2. Piracetam | All | Dropout (any reason) | Some concerns | Low | Low | Low | Low | Some concerns | D1: Limited information regarding randomization and allocation process. |
|  |  | All | Serious adverse events | Some concerns | Low | Low | Low | Low | Some concerns | D1: Limited information regarding randomization and allocation process. |
|  |  | All | Extent of use (BE level; EoT) | Some concerns | Low | High | Low | Some concerns | High | D1: Limited information regarding randomization and allocation process. D3: Some missing data and likely that it could depend on the true value of the outcome.  D5: No evidence of a pre-specified analysis plan. |
| Kampman 2004 | 1. Placebo 2. Topiramate | All | Dropout (any reason) | Some concerns | Low | Low | Low | Low | Some concerns | D1: Limited information regarding randomization and allocation process. |
|  |  | All | Point abstinence (EoT) | Some concerns | Low | Low | Low | Some concerns | Some concerns | D1: Limited information regarding randomization and allocation process. D5: No evidence of a pre-specified analysis plan. |
| Kampman 2006 | 1. Placebo 2. Propranolol 3. Amantadine 4. Amantadine + propranolol | All | Dropout (any reason) | Some concerns | Low | Low | Low | Low | Some concerns | D1: Limited information regarding allocation process. |
|  |  | 2 vs 1;  3 vs 2;  4 vs 2 | Adherence | Some concerns | Low | High | Low | Some concerns | High | D1: Limited information regarding allocation process. D3: Some missing data and likely that it could depend on the true value of the outcome. D5: No evidence of a pre-specified analysis plan. |
|  |  | 3 vs 1;  4 vs 1;  4 vs 3 | Adherence | Some concerns | Low | Some concerns | Low | Some concerns | Some concerns | D1: Limited information regarding allocation process. D3: Some missing data but unlikely that missingness depended on the true value of outcome.  D5: No evidence of a pre-specified analysis plan. |
|  |  | All | Point abstinence (EoT) | Some concerns | Low | Low | Low | Some concerns | Some concerns | D1: Limited information regarding allocation process. D5: No evidence of a pre-specified analysis plan. |
| Kampman 2011 | 1. Placebo 2. Acamprosate | All | Dropout (any reason) | Low | Low | Low | Low | Low | Low |  |
|  |  | All | Adherence | Low | Low | High | Low | Some concerns | High | D3: Some missing data and likely that it could depend on the true value of the outcome.  D5: No evidence of a pre-specified analysis plan. |
|  |  | All | Extent of use (proportion tests/days; EoT) | Low | Low | Low | Low | Some concerns | Some concerns | D5: No evidence of a pre-specified analysis plan. |
| Kampman 2013 | 1. Placebo 2. Topiramate | All | Dropout (any reason) | Some concerns | Low | Low | Low | Low | Some concerns | D1: Limited information regarding allocation process. |
|  |  | All | Adherence | Some concerns | Low | High | Low | Some concerns | High | D1: Limited information regarding allocation process. D3: Some missing data and likely that it could depend on the true value of the outcome.  D5: No evidence of a pre-specified analysis plan. |
|  |  | All | Continuous abstinence (treatment) | Some concerns | Low | Low | Low | Some concerns | Some concerns | D1: Limited information regarding allocation process. D5: No evidence of a pre-specified analysis plan. |
| Kampman 2015 | 1. Placebo 2. Modafinil | All | Dropout (any reason) | Low | Low | Low | Low | Low | Low |  |
|  |  | All | Dropout (adverse events) | Low | Low | Some concerns | Low | Low | Some concerns | D3: Some missing data but unlikely that missingness depended on the true value of outcome. |
|  |  | All | Adherence | Low | Low | Some concerns | Low | Some concerns | Some concerns | D3: Some missing data but unlikely that missingness depended on the true value of outcome.  D5: No evidence of a pre-specified analysis plan. |
|  |  | All | Continuous abstinence (treatment) | Low | Low | Low | Low | Some concerns | Some concerns | D5: No evidence of a pre-specified analysis plan. |
| Karila 2016 | 1. Placebo 2. Modafinil | All | Dropout (any reason) | Low | Low | Low | Low | Low | Low |  |
|  |  | All | Continuous abstinence (treatment) | Low | Low | High | Low | Some concerns | High | D3: Some missing data and likely that it could depend on the true value of the outcome.  D5: No evidence of a pre-specified analysis plan. |
| Kennedy 2012 | 1. Placebo 2. D-Cycloserine | All | Dropout due to any reason | Some concerns | Low | Low | Low | Low | Some concerns | D1: Limited information regarding allocation process. |
|  |  | All | Continuous abstinence (treatment) | Some concerns | Some concerns | Low | Low | Some concerns | Some concerns | D1: Limited information regarding allocation process. D2: Analysis restricted to those receiving first dose - but judged unlikely to have substantial impact on the results.  D5: No evidence of a pre-specified analysis plan. |
| Kolar 1992 | 1. Placebo 2. Desipramine 3. Amantadine | All | Continuous abstinence (treatment) | Low | High | High | Low | Some concerns | High | D2: Analysis was restricted to N treated (i.e., not ITT or mITT) and potential issues with blinding of participants and/or outcome assessors.  D3: Some missing data and likely that it could depend on the true value of the outcome.  D5: No evidence of a pre-specified analysis plan. |
|  |  | All | Continuous abstinence (follow-up) | Low | High | High | Low | Some concerns | High | D2: Analysis was restricted to N treated (i.e., not ITT or mITT) and potential issues with blinding of participants and/or outcome assessors.  D3: Some missing data and likely that it could depend on the true value of the outcome.  D5: No evidence of a pre-specified analysis plan. |
|  |  | All | Extent of use (quantity; EoT) | Low | High | High | High | Some concerns | High | D2: Analysis was restricted to N treated (i.e., not ITT or mITT) and potential issues with blinding of participants and/or outcome assessors.  D3: Some missing data and likely that it could depend on the true value of the outcome.  D4: The outcome was self-reported and participants were likely aware of their group allocation. D5: No evidence of a pre-specified analysis plan. |
|  |  | All | Craving | Low | High | High | High | Some concerns | High | D2: Analysis was restricted to N treated (i.e., not ITT or mITT) and potential issues with blinding of participants and/or outcome assessors.  D3: Some missing data and likely that it could depend on the true value of the outcome.  D4: The outcome was self-reported and participants were likely aware of their group allocation. D5: No evidence of a pre-specified analysis plan. |
| Kosten 1992 | 1. Placebo 2. Amantadine 3. Desipramine | All | Dropout (any reason) | Some concerns | Low | Low | Low | Low | Some concerns | D1: Limited information regarding randomization and allocation process. |
|  |  | All | Dropout (adverse events) | Some concerns | Low | Low | Low | Low | Some concerns | D1: Limited information regarding randomization and allocation process. |
|  |  | 2 vs 1;  3 vs 1 | Continuous abstinence (treatment) | Some concerns | Low | High | Low | Some concerns | High | D1: Limited information regarding randomization and allocation process. D3: Some missing data and likely that it could depend on the true value of the outcome.  D5: No evidence of a pre-specified analysis plan. |
|  |  | 3 vs 2 | Continuous abstinence (treatment) | Some concerns | Low | Some concerns | Low | Some concerns | Some concerns | D1: Limited information regarding randomization and allocation process. D3: Some missing data but unlikely that missingness depended on the true value of outcome.  D5: No evidence of a pre-specified analysis plan. |
|  |  | 2 vs 1;  3 vs 1 | Extent of use (quantity; EoT) | Some concerns | Low | High | Low | Some concerns | High | D1: Limited information regarding randomization and allocation process. D3: Some missing data and likely that it could depend on the true value of the outcome.  D5: No evidence of a pre-specified analysis plan. |
|  |  | 3 vs 2 | Extent of use (quantity; EoT) | Some concerns | Low | Some concerns | Low | Some concerns | Some concerns | D1: Limited information regarding randomization and allocation process. D3: Some missing data but unlikely that missingness depended on the true value of outcome.  D5: No evidence of a pre-specified analysis plan. |
| Kosten 2002 | 1. Placebo 2. Ketoconazole | All | Dropout (any reason) | Some concerns | Low | Low | Low | Low | Some concerns | D1: Limited information regarding randomization and allocation process. |
| Kosten 2003 | 1. Placebo 2. Desipramine | All | Longest duration of continuous abstinence | Some concerns | Low | High | Low | Some concerns | High | D1: Limited information regarding allocation process. D3: Some missing data. Amount and difference between groups unclear.  D5: No evidence of a pre-specified analysis plan. |
| Kosten 2013 | 1. Placebo 2. Disulfiram | All | Dropout (any reason) | Low | Low | Low | Low | Low | Low |  |
|  |  | All | Dropout (adverse events) | Low | Low | Low | Low | Low | Low |  |
|  |  | All | Extent of use (proportion tests/days; EoT) | Low | Low | Low | Low | Some concerns | Some concerns | D5: No evidence of a pre-specified analysis plan. |
| Kosten 2014 | 1. Placebo 2. TA-CD vaccine | All | Dropout (any reason) | Some concerns | Low | Low | Low | Low | Some concerns | D1: Limited information regarding allocation process. |
|  |  | All | Serious adverse events | Some concerns | Low | Low | Low | Low | Some concerns | D1: Limited information regarding allocation process. |
|  |  | All | Continuous abstinence (treatment) | Some concerns | Low | Some concerns | Low | High | High | D1: Limited information regarding allocation process. D3: Some missing data but unlikely that missingness depended on the true value of outcome.  D5: No evidence of a pre-specified analysis plan. The result could have been selected from multiple eligible outcome measurements. |
| Kranzler 1995 | 1. Placebo 2. Carbamazepine | All | Dropout (any reason) | Low | Low | Low | Low | Low | Low |  |
|  |  | All | Dropout (adverse events) | Low | Low | Some concerns | Low | Low | Some concerns | D3: Some missing data but unlikely that missingness depended on the true value of outcome. |
|  |  | All | Adherence | Low | Low | Some concerns | Low | Some concerns | Some concerns | D3: Some missing data but unlikely that missingness depended on the true value of outcome.  D5: No evidence of a pre-specified analysis plan. |
|  |  | All | Extent of use (frequency; EoT) | Low | Low | Some concerns | Low | Some concerns | Some concerns | D3: Some missing data but unlikely that missingness depended on the true value of outcome.  D5: No evidence of a pre-specified analysis plan. |
|  |  | All | Extent of use (frequency; follow-up) | Low | Low | Some concerns | Low | Some concerns | Some concerns | D3: Some missing data but unlikely that missingness depended on the true value of outcome.  D5: No evidence of a pre-specified analysis plan. |
|  |  | All | Extent of use (quantity; EoT) | Low | Low | Some concerns | Low | Some concerns | Some concerns | D3: Some missing data but unlikely that missingness depended on the true value of outcome.  D5: No evidence of a pre-specified analysis plan. |
|  |  | All | Extent of use (quantity; follow-up) | Low | Low | Some concerns | Low | Some concerns | Some concerns | D3: Some missing data but unlikely that missingness depended on the true value of outcome.  D5: No evidence of a pre-specified analysis plan. |
| LaRowe 2013 | 1. Placebo 2. N-acetylcysteine (2400mg) 3. N-acetylcysteine (1200mg) | All | Dropout (any reason) | Low | Low | Low | Low | Low | Low |  |
|  |  | All | Adherence | Low | High | Some concerns | Low | Some concerns | High | D2: Appears that a per protocol analysis was used (e.g., exclusions due to non-compliance). D3: Some missing data but unlikely that missingness depended on the true value of outcome.  D5: No evidence of a pre-specified analysis plan. |
|  |  | All | Extent of use (BE level; EoT) | Low | High | Some concerns | Low | Some concerns | High | D2: Appears that a per protocol analysis was used (e.g., exclusions due to non-compliance). D3: Some missing data but unlikely that missingness depended on the true value of outcome.  D5: No evidence of a pre-specified analysis plan. |
| Levin 1999 | 1. Placebo 2. Risperidone | All | Dropout (any reason) | Some concerns | Low | Low | Low | Low | Some concerns | D1: Limited information regarding randomization and allocation process. |
| Levin 2007 | 1. Placebo 2. Methylphenidate | All | Dropout (any reason) | Some concerns | Low | Low | Low | Low | Some concerns | D1: Limited information regarding allocation process. |
|  |  | All | Dropout (adverse events) | Some concerns | Low | High | Low | Low | High | D1: Limited information regarding allocation process. D3: Some missing data. Amount and difference between groups unclear. |
|  |  | All | Adherence | Some concerns | Low | Some concerns | Low | Some concerns | Some concerns | D1: Limited information regarding allocation process. D3: Some missing data but unlikely that missingness depended on the true value of outcome.  D5: No evidence of a pre-specified analysis plan. |
|  |  | All | Continuous abstinence (treatment) | Some concerns | Low | High | Low | Some concerns | High | D1: Limited information regarding allocation process. D3: Some missing data. Amount and difference between groups unclear.  D5: No evidence of a pre-specified analysis plan. |
|  |  | All | Extent of use (proportion tests/days; EoT) | Some concerns | Low | High | Low | Some concerns | High | D1: Limited information regarding allocation process. D3: Some missing data. Amount and difference between groups unclear.  D5: No evidence of a pre-specified analysis plan. |
| Levin 2015 | 1. Placebo 2. MAS-ER + topiramate (60mg) 3. MAS-ER + topiramate (80mg) | All | Dropout (any reason) | Low | Low | Low | Low | Low | Low |  |
|  |  | All | Serious adverse events | Low | Low | Low | Low | Low | Low |  |
|  |  | All | Continuous abstinence (treatment) | Low | Low | Some concerns | Low | Some concerns | Some concerns | D3: Some missing data but unlikely that missingness depended on the true value of outcome.  D5: No evidence of a pre-specified analysis plan. |
|  |  | All | Point abstinence (EoT) | Low | Low | Low | Low | Some concerns | Some concerns | D5: No evidence of a pre-specified analysis plan. |
| Levin 2020 | 1. Placebo 2. MAS-ER + topiramate | All | Dropout (any reason) | Low | Low | Low | Low | Low | Low |  |
|  |  | All | Adherence | Low | Low | Some concerns | Low | Some concerns | Some concerns | D3: Some missing data but unlikely that missingness depended on the true value of outcome.  D5: No evidence of a pre-specified analysis plan. |
|  |  | All | Serious adverse events | Low | Low | Low | Low | Low | Low |  |
|  |  | All | Continuous abstinence (treatment) | Low | Low | Some concerns | Low | Some concerns | Some concerns | D3: Some missing data but unlikely that missingness depended on the true value of outcome.  D5: No evidence of a pre-specified analysis plan. |
|  |  | All | Craving | Low | Low | Some concerns | Low | Some concerns | Some concerns | D3: Some missing data but unlikely that missingness depended on the true value of outcome.  D5: No evidence of a pre-specified analysis plan. |
| Licata 2011 | 1. Placebo 2. Citicoline | All | Dropout (any reason) | Some concerns | Low | Low | Low | Low | Some concerns | D1: Limited information regarding randomization and allocation process. |
|  |  | All | Extent of use (frequency; EoT) | Some concerns | Low | Some concerns | Low | Some concerns | Some concerns | D1: Limited information regarding randomization and allocation process. D3: Some missing data but unlikely that missingness depended on the true value of outcome.  D5: No evidence of a pre-specified analysis plan. |
|  |  | All | Extent of use (frequency; follow-up) | Some concerns | Low | Some concerns | Low | Some concerns | Some concerns | D1: Limited information regarding randomization and allocation process. D3: Some missing data but unlikely that missingness depended on the true value of outcome.  D5: No evidence of a pre-specified analysis plan. |
|  |  | All | Craving | Some concerns | Low | Some concerns | Low | Some concerns | Some concerns | D1: Limited information regarding randomization and allocation process. D3: Some missing data but unlikely that missingness depended on the true value of outcome.  D5: No evidence of a pre-specified analysis plan. |
| Ling 2016 | 1. Placebo 2. Buprenorphine + naloxone (16/4mg) 3. Buprenorphine + naloxone (4/1mg) | All | Dropout (any reason) | Low | Low | Low | Low | Low | Low |  |
|  |  | All | Adherence | Low | Low | Low | Low | Some concerns | Some concerns | D5: No evidence of a pre-specified analysis plan. |
|  |  | All | Serious adverse events | Low | Low | Low | Low | Low | Low |  |
|  |  | All | Mortality | Low | Low | Low | Low | Low | Low |  |
|  |  | All | Continuous abstinence (treatment) | Low | Low | Low | Low | High | High | D5: Discrepancy between registry/protocol and report over outcome measure used. |
|  |  | All | Extent of use (proportion tests/days; EoT) | Low | Low | Low | Low | Low | Low |  |
| Loebl 2008 | 1. Placebo 2. Risperidone | All | Dropout (any reason) | Some concerns | Low | Low | Low | Low | Some concerns | D1: Limited information regarding randomization and allocation process. |
|  |  | All | Dropout (adverse events) | Some concerns | Low | High | Low | Low | High | D1: Limited information regarding randomization and allocation process.  D3: Some missing data and likely that it could depend on the true value of the outcome. |
|  |  | All | Extent of use (proportion tests/days; EoT) | Some concerns | Low | High | Low | Some concerns | High | D1: Limited information regarding randomization and allocation process. D3: Some missing data and likely that it could depend on the true value of the outcome.  D5: No evidence of a pre-specified analysis plan. |
|  |  | All | Extent of use (frequency; EoT) | Some concerns | Low | High | Low | Some concerns | High | D1: Limited information regarding randomization and allocation process. D3: Some missing data and likely that it could depend on the true value of the outcome.  D5: No evidence of a pre-specified analysis plan. |
| Lynch 2022 | 1. Placebo 2. Varenicline | All | Dropout (any reason) | Low | Low | Low | Low | Low | Low |  |
|  |  | All | Dropout (adverse events) | Low | Low | Some concerns | Low | Low | Some concerns | D3: Some missing data but unlikely that missingness depended on the true value of outcome. |
|  |  | All | Adherence | Low | Low | Some concerns | Low | High | High | D3: Some missing data but unlikely that missingness depended on the true value of outcome. D5: Discrepancy between registry and report over outcome measure used. |
|  |  | All | Serious adverse events | Low | Low | Low | Low | Low | Low |  |
|  |  | All | Continuous abstinence (treatment) | Low | Low | Low | Low | High | High | D5: Discrepancy between registry and report over outcome measure used. |
|  |  | All | Point abstinence (EoT) | Low | Low | Some concerns | Low | Some concerns | Some concerns | D3: Some missing data but unlikely that missingness depended on the true value of outcome.  D5: No evidence of a pre-specified analysis plan. |
| Malcolm 2000 | 1. Placebo 2. Pergolide (0.25mg) 3. Pergolide (0.05mg) | All | Dropout (any reason) | Some concerns | Low | Low | Low | Low | Some concerns | D1: Limited information regarding allocation process. |
|  |  | All | Adherence | Some concerns | High | High | Low | Some concerns | High | D1: Limited information regarding allocation process. D2: Analysis was restricted to N treated (i.e., not ITT or mITT). D3: Some missing data and likely that it could depend on the true value of the outcome.  D5: No evidence of a pre-specified analysis plan. |
|  |  | All | Point abstinence (follow-up) | Some concerns | High | High | Low | Some concerns | High | D1: Limited information regarding allocation process. D2: Analysis was restricted to N treated (i.e., not ITT or mITT).  D3: Some missing data and likely that it could depend on the true value of the outcome.  D5: No evidence of a pre-specified analysis plan. |
| Malcolm 2005 | 1. Placebo 2. Amlodipine | All | Dropout (any reason) | Some concerns | High | Low | Low | Low | High | D1: Limited information regarding allocation process. Baseline demographics only reported for subset of randomised participants.  D2: Analysis was restricted to N treated (i.e., not ITT or mITT). |
|  |  | All | Adherence | Some concerns | High | High | Low | Some concerns | High | D1: Limited information regarding allocation process. Baseline demographics only reported for subset of randomised participants.  D2: Analysis was restricted to N treated (i.e., not ITT or mITT).  D3: Some missing data and likely that it could depend on the true value of the outcome.  D5: No evidence of a pre-specified analysis plan. |
|  |  | All | Point abstinence (EoT) | Some concerns | High | High | Low | Some concerns | High | D1: Limited information regarding allocation process. Baseline demographics only reported for subset of randomised participants.  D2: Analysis was restricted to N treated (i.e., not ITT or mITT).  D3: Some missing data and likely that it could depend on the true value of the outcome.  D5: No evidence of a pre-specified analysis plan. |
|  |  | All | Extent of use (BE level; EoT) | Some concerns | High | High | Low | Some concerns | High | D1: Limited information regarding allocation process. Baseline demographics only reported for subset of randomised participants.  D2: Analysis was restricted to N treated (i.e., not ITT or mITT).  D3: Some missing data and likely that it could depend on the true value of the outcome.  D5: No evidence of a pre-specified analysis plan. |
| Mancino 2014 | 1. Placebo 2. Sertraline 3. Gabapentin + sertraline | All | Dropout (any reason) | Low | Low | Low | Low | Low | Low |  |
|  |  | All | Adherence | Low | High | High | Low | Some concerns | High | D2: Analysis was restricted to N treated (i.e., not ITT or mITT).  D3: Some missing data and likely that it could depend on the true value of the outcome.  D5: No evidence of a pre-specified analysis plan. |
|  |  | All | Serious adverse events | Low | Low | Low | Low | Low | Low |  |
|  |  | All | Continuous abstinence (treatment) | Low | High | High | Low | Some concerns | High | D2: Analysis was restricted to N treated (i.e., not ITT or mITT).  D3: Some missing data and likely that it could depend on the true value of the outcome.  D5: No evidence of a pre-specified analysis plan. |
| Margolin 1995 | 1. Placebo 2. Bupropion | All | Dropout (any reason) | Some concerns | Some concerns | Low | Low | Low | Some concerns | D1: Limited information regarding randomization and allocation process. D2: Analysis restricted to those receiving first dose - but judged unlikely to have substantial impact on the results. |
|  |  | All | Extent of use (proportion tests/days; EoT) | Some concerns | Some concerns | Some concerns | Low | Some concerns | Some concerns | D1: Limited information regarding randomization and allocation process D2: Analysis restricted to those receiving first dose - but judged unlikely to have substantial impact on the results.  D3: Some missing data but unlikely that missingness depended on the true value of outcome.  D5: No evidence of a pre-specified analysis plan. |
|  |  | All | Extent of use (frequency; EoT) | Some concerns | Some concerns | Some concerns | Low | Some concerns | Some concerns | D1: Limited information regarding randomization and allocation process D2: Analysis restricted to those receiving first dose - but judged unlikely to have substantial impact on the results.  D3: Some missing data but unlikely that missingness depended on the true value of outcome.  D5: No evidence of a pre-specified analysis plan. |
|  |  | All | Extent of use (quantity; EoT) | Some concerns | Some concerns | Some concerns | Low | Some concerns | Some concerns | D1: Limited information regarding randomization and allocation process D2: Analysis restricted to those receiving first dose - but judged unlikely to have substantial impact on the results.  D3: Some missing data but unlikely that missingness depended on the true value of outcome.  D5: No evidence of a pre-specified analysis plan. |
|  |  | All | Craving | Some concerns | Some concerns | Some concerns | Low | Some concerns | Some concerns | D1: Limited information regarding randomization and allocation process D2: Analysis restricted to those receiving first dose - but judged unlikely to have substantial impact on the results.  D3: Some missing data but unlikely that missingness depended on the true value of outcome.  D5: No evidence of a pre-specified analysis plan. |
| Margolin 1997 | 1. Placebo 2. Mazindol (1mg) 3. Mazindol (8mg) | All | Dropout (any reason) | Some concerns | Low | Low | Low | Low | Some concerns | D1: Limited information regarding randomization and allocation process. Baseline demographics not reported by trial arm. |
|  |  | All | Point abstinence (EoT) | Some concerns | Low | High | Low | Some concerns | High | D1: Limited information regarding randomization and allocation process. Baseline demographics not reported by trial arm.  D3: Some missing data and likely that it could depend on the true value of the outcome.  D5: No evidence of a pre-specified analysis plan. |
|  |  | All | Craving | Some concerns | Low | High | Low | Some concerns | High | D1: Limited information regarding randomization and allocation process. Baseline demographics not reported by trial arm.  D3: Some missing data and likely that it could depend on the true value of the outcome.  D5: No evidence of a pre-specified analysis plan. |
| Mariani 2012 | 1. Placebo 2. MAS-ER + topiramate | All | Dropout (any reason) | Low | Low | Low | Low | Low | Low |  |
|  |  | All | Dropout (adverse events) | Low | Low | Some concerns | Low | Low | Some concerns | D3: Some missing data but unlikely that missingness depended on the true value of outcome. |
|  |  | All | Serious adverse events | Low | Low | Low | Low | Low | Low |  |
|  |  | All | Continuous abstinence (treatment) | Low | Low | Some concerns | Low | High | High | D3: Some missing data but unlikely that missingness depended on the true value of outcome.  D5: Discrepancy between registry and report over outcome measure used. |
| Martell 2009 | 1. Placebo 2. TA-CD vaccine | All | Dropout (any reason) | Low | Low | Low | Low | Low | Low |  |
|  |  | All | Dropout (adverse events) | Low | Low | Some concerns | Low | Low | Some concerns | D3: Some missing data but unlikely that missingness depended on the true value of outcome. |
|  |  | All | Serious adverse events | Low | Some concerns | Low | Low | Low | Some concerns | D2: Analysis restricted to those receiving first dose - but judged unlikely to have substantial impact on the results. |
| McCann 2024 | 1. Placebo 2. Lorcaserin | All | Dropout (any reason) | Low | Low | Low | Low | Low | Low |  |
|  |  | All | Serious adverse events | Low | Low | Low | Low | Low | Low |  |
|  |  | All | Continuous abstinence (treatment) | Low | Low | Some concerns | Low | Low | Some concerns | D3: Some missing data but unlikely that missingness depended on the true value of outcome. |
|  |  | All | Craving | Low | High | Some concerns | Low | Low | High | D2: Appears that a per protocol analysis was used (e.g., exclusions due to non-compliance)  D3: Some missing data but unlikely that missingness depended on the true value of outcome. |
| McDowell 2005 | 1. Placebo 2. Desipramine | All | Dropout (any reason) | Low | Low | Low | Low | Low | Low |  |
|  |  | All | Dropout (adverse events) | Low | Low | High | Low | Low | High | D3: Some missing data and likely that it could depend on the true value of the outcome. |
|  |  | All | Serious adverse events | Low | Low | Low | Low | Low | Low |  |
|  |  | All | Continuous abstinence (treatment) | Low | Low | High | Low | Some concerns | High | D3: Some missing data and likely that it could depend on the true value of the outcome.  D5: No evidence of a pre-specified analysis plan. |
|  |  | All | Extent of use (frequency; EoT) | Low | Low | High | Low | Some concerns | High | D3: Some missing data and likely that it could depend on the true value of the outcome.  D5: No evidence of a pre-specified analysis plan. |
| NCT03953612 | 1. Placebo 2. Pregnenolone (500mg) 3. Pregnenolone (300mg) | All | Dropout (any reason) | Low | Low | Low | Low | Low | Low |  |
|  |  | All | Serious adverse events | Low | Some concerns | Low | Low | Low | Some concerns | D2: Analysis restricted to those receiving first dose - but judged unlikely to have substantial impact on the results. |
|  |  | 2 vs 1 | Extent of use (frequency; EoT) | Low | Some concerns | Low | Low | Some concerns | Some concerns | D2: Analysis restricted to those receiving first dose - but judged unlikely to have substantial impact on the results.  D5: No evidence of a pre-specified analysis plan. |
|  |  | 3 vs 1;  3 vs 2 | Extent of use (frequency; EoT) | Low | Some concerns | Some concerns | Low | Some concerns | Some concerns | D2: Analysis restricted to those receiving first dose - but judged unlikely to have substantial impact on the results.  D3: Some missing data but unlikely that missingness depended on the true value of outcome.  D5: No evidence of a pre-specified analysis plan. |
|  |  | 2 vs 1 | Extent of use (quantity; EoT) | Low | Some concerns | Low | Low | Some concerns | Some concerns | D2: Analysis restricted to those receiving first dose - but judged unlikely to have substantial impact on the results.  D5: No evidence of a pre-specified analysis plan. |
|  |  | 3 vs 1;  3 vs 2 | Extent of use (quantity; EoT) | Low | Some concerns | Some concerns | Low | Some concerns | Some concerns | D2: Analysis restricted to those receiving first dose - but judged unlikely to have substantial impact on the results.  D3: Some missing data but unlikely that missingness depended on the true value of outcome.  D5: No evidence of a pre-specified analysis plan. |
| Moeller 2001 | 1. Placebo 2. Buspirone | All | Dropout (any reason) | Some concerns | High | Low | Low | Low | High | D1: Limited information regarding allocation process. Baseline demographics only reported for subset of randomised participants.  D2: Analysis was restricted to N treated (i.e., not ITT or mITT). |
|  |  | All | Craving | Some concerns | High | High | Low | Some concerns | High | D1: Limited information regarding allocation process. Baseline demographics only reported for subset of randomised participants.  D2: Analysis was restricted to N treated (i.e., not ITT or mITT).  D3: Some missing data and likely that it could depend on the true value of the outcome.  D5: No evidence of a pre-specified analysis plan. |
| Mongeau-Perusse 2021 | 1. Placebo 2. Cannabidiol | All | Dropout (any reason) | Low | Low | Low | Low | Low | Low |  |
|  |  | All | Serious adverse events | Low | High | Low | Low | Low | High | D2: Analysis was restricted to N treated (i.e., not ITT or mITT). |
|  |  | All | Extent of use (proportion tests/days; EoT) | Low | High | Some concerns | Low | Low | High | D2: Analysis was restricted to N treated (i.e., not ITT or mITT).  D3: Some missing data but unlikely that missingness depended on the true value of outcome. |
| Montoya 1995 | 1. Placebo 2. Carbamazepine | All | Dropout (any reason) | Some concerns | High | Low | Low | Low | High | D1: Limited information regarding randomization and allocation process. Baseline demographics only reported for subset of randomised participants.  D2: Analysis was restricted to N treated (i.e., not ITT or mITT). |
| Mooney 2007 (study 1) | 1. Placebo 2. Levodopa + carbidopa | All | Dropout (any reason) | Some concerns | Low | Low | Low | Low | Some concerns | D1: Limited information regarding randomization and allocation process. |
|  |  | All | Dropout (adverse events) | Some concerns | Low | Some concerns | Low | Low | Some concerns | D1: Limited information regarding allocation process. D3: Some missing data but unlikely that missingness depended on the true value of outcome. |
| Mooney 2007 (study 2) | 1. Placebo 2. Levodopa + Carbidopa (800/200 mg) 3. Levodopa + Carbidopa (400/100 mg) | All | Dropout due to any reason | Some concerns | Low | Low | Low | Low | Some concerns | D1: Limited information regarding allocation process. |
| Mooney 2009 | 1. Placebo 2. Methamphetamine (immediate release) 3. Methamphetamine (sustained release) | All | Dropout (any reason) | Some concerns | Low | Low | Low | Low | Some concerns | D1: Limited information regarding allocation process. |
|  |  | 2 vs 1;  3 vs 1 | Dropout (adverse events) | Some concerns | Low | Some concerns | Low | Low | Some concerns | D1: Limited information regarding allocation process. D3: Some missing data but unlikely that missingness depended on the true value of outcome. |
|  |  | 3 vs 2 | Dropout (adverse events) | Some concerns | Low | High | Low | Low | High | D1: Limited information regarding allocation process. D3: Some missing data and likely that it could depend on the true value of the outcome. |
|  |  | All | Adherence | Some concerns | Low | High | Low | Some concerns | High | D1: Limited information regarding allocation process. D3: Some missing data and likely that it could depend on the true value of the outcome.  D5: No evidence of a pre-specified analysis plan. |
|  |  | All | Craving | Some concerns | Low | High | Low | Some concerns | High | D1: Limited information regarding allocation process. D3: Some missing data and likely that it could depend on the true value of the outcome.  D5: No evidence of a pre-specified analysis plan. |
| Mooney 2015 | 1. Placebo 2. Lisdexamfetamine | All | Dropout (any reason) | Some concerns | Low | Low | Low | Low | Some concerns | D1: Limited information regarding randomization and allocation process. |
|  |  | All | Dropout (adverse events) | Some concerns | Low | High | High | Low | High | D1: Limited information regarding randomization and allocation process. D3: Some missing data and likely that it could depend on the true value of the outcome.  D4: The outcome was self-reported and participants were likely aware of their group allocation. |
|  |  | All | Serious adverse events | Some concerns | Low | Low | Low | Low | Some concerns | D1: Limited information regarding randomization and allocation process |
|  |  | All | Point abstinence (EoT) | Some concerns | Low | Low | Low | Some concerns | Some concerns | D1: Limited information regarding randomization and allocation process. D5: No evidence of a pre-specified analysis plan. |
|  |  | All | Craving | Some concerns | Low | High | High | Some concerns | High | D1: Limited information regarding randomization and allocation process. D3: Some missing data and likely that it could depend on the true value of the outcome.  D4: The outcome was self-reported and participants were likely aware of their group allocation.  D5: No evidence of a pre-specified analysis plan. |
| Morgan 2016 | 1. Placebo 2. Modafinil | All | Dropout (any reason) | Low | Low | Low | Low | Low | Low |  |
|  |  | All | Longest duration of continuous abstinence | Low | Low | High | Low | Some concerns | High | D3: Some missing data and likely that it could depend on the true value of the outcome. D5: No evidence of a pre-specified analysis plan. |
| Nanni-Alvarado 2022 | 1. Placebo 2. Mirtazapine | All | Dropout (any reason) | Low | Low | Low | Low | Low | Low |  |
|  |  | All | Craving | Low | High | High | Low | Some concerns | High | D2: Appears that a per protocol analysis was used (e.g., exclusions due to non-compliance). D3: Some missing data and likely that it could depend on the true value of the outcome.  D5: No evidence of a pre-specified analysis plan; registration appears to be retrospective. |
| Noel Raby 2022 | 1. Placebo 2. Oxytocin | All | Dropout (any reason) | Low | High | Low | Low | Low | High | D2: Analysis was restricted to N treated (i.e., not ITT or mITT). |
|  |  | All | Point abstinence (EoT) | Low | High | High | Low | Some concerns | High | D2: Analysis was restricted to N treated (i.e., not ITT or mITT). D3: Some missing data and likely that it could depend on the true value of the outcome.  D5: No evidence of a pre-specified analysis plan. |
|  |  | All | Longest duration of continuous abstinence | Low | High | High | Low | Some concerns | High | D2: Analysis was restricted to N treated (i.e., not ITT or mITT). D3: Some missing data and likely that it could depend on the true value of the outcome.  D5: No evidence of a pre-specified analysis plan. |
| Nuijten 2016 | 1. Placebo 2. Dexamfetamine | All | Dropout (any reason) | Low | Low | Low | Low | Low | Low |  |
|  |  | All | Dropout (adverse events) | Low | Low | Some concerns | Low | Low | Some concerns | D3: Some missing data but unlikely that missingness depended on the true value of outcome. |
|  |  | All | Serious adverse events | Low | Low | Low | Low | Low | Low |  |
|  |  | All | Continuous abstinence (treatment) | Low | Low | Some concerns | Low | Some concerns | Some concerns | D3: Some missing data but unlikely that missingness depended on the true value of outcome. D5: No evidence of a pre-specified analysis plan. |
|  |  | All | Longest duration of continuous abstinence | Low | Low | Some concerns | Low | Low | Some concerns | D3: Some missing data but unlikely that missingness depended on the true value of outcome. |
|  |  | All | Extent of use (frequency; EoT) | Low | Low | Some concerns | Low | Low | Some concerns | D3: Some missing data but unlikely that missingness depended on the true value of outcome. |
| Oliva 2022 | 1. Placebo 2. Progesterone | All | Dropout (any reason) | Some concerns | Low | Low | Low | Low | Some concerns | D1: Limited information regarding allocation process. |
|  |  | All | Serious adverse events | Some concerns | Low | Low | Low | Low | Some concerns | D1: Limited information regarding allocation process. |
|  |  | All | Continuous abstinence (treatment) | Some concerns | Low | Some concerns | Low | High | High | D1: Limited information regarding allocation process. D3: Some missing data but unlikely that missingness depended on the true value of outcome. D5: Discrepancy between registry and report over outcome measure used. |
| Oliveto 2011 | 1. Placebo 2. Disulfiram (62.5mg) 3. Disulfiram (125mg) 4. Disulfiram (250mg) | All | Dropout (any reason) | Low | Low | Low | Low | Low | Low |  |
|  |  | 2 vs 1;  4 vs 1;  3 vs 2;  4 vs 2;  4 vs 3 | Point abstinence (EoT) | Low | Some concerns | Some concerns | Low | Some concerns | Some concerns | D2: Analysis restricted to those receiving first dose - but judged unlikely to have substantial impact on the results. D3: Some missing data but unlikely that missingness depended on the true value of outcome. D5: No evidence of a pre-specified analysis plan. |
|  |  | 3 vs 1 | Point abstinence (EoT) | Low | some concerns | High | Low | Some concerns | High | D2: Analysis restricted to those receiving first dose - but judged unlikely to have substantial impact on the results. D3: Some missing data and likely that it could depend on the true value of the outcome. D5: No evidence of a pre-specified analysis plan. |
|  |  | 2 vs 1;  3 vs 2;  4 vs 2;  4 vs 3 | Extent of use (quantity; EoT) | Low | Some concerns | Some concerns | Low | Some concerns | Some concerns | D2: Analysis restricted to those receiving first dose - but judged unlikely to have substantial impact on the results. D3: Some missing data but unlikely that missingness depended on the true value of outcome. D5: No evidence of a pre-specified analysis plan. |
|  |  | 3 vs 1;  4 vs 1 | Extent of use (quantity; EoT) | Low | Some concerns | High | Low | Some concerns | High | D2: Analysis restricted to those receiving first dose - but judged unlikely to have substantial impact on the results. D3: Some missing data and likely that it could depend on the true value of the outcome. D5: No evidence of a pre-specified analysis plan. |
| Oliveto 2012 | 1. Placebo 2. Sertraline | All | Dropout (any reason) | Low | Low | Low | Low | Low | Low |  |
|  |  | All | Dropout (adverse events) | Low | Low | High | Low | Low | High | D3: Some missing data and likely that it could depend on the true value of the outcome. |
|  |  | All | Continuous abstinence (treatment) | Low | High | High | Low | Some concerns | High | D2: Appears that a per protocol analysis was used (e.g., exclusions due to non-compliance) D3: Some missing data and likely that it could depend on the true value of the outcome. D5: No evidence of a pre-specified analysis plan. |
| Passos 2005 | 1. Placebo 2. Nefazodone | All | Dropout (any reason) | Low | Low | Low | Low | Low | Low |  |
|  |  | All | Dropout (adverse events) | Low | Low | High | Low | Low | High | D3: Some missing data and likely that it could depend on the true value of the outcome. |
|  |  | All | Adherence | Low | Low | High | Low | Some concerns | High | D3: Some missing data and likely that it could depend on the true value of the outcome. D5: No evidence of a pre-specified analysis plan. |
|  |  | All | Craving | Low | Low | High | Low | Some concerns | High | D3: Some missing data and likely that it could depend on the true value of the outcome. D5: No evidence of a pre-specified analysis plan. |
| Petrakis 2000 | 1. Placebo 2. Disulfiram | All | Dropout (any reason) | Some concerns | Some concerns | Low | Low | Low | Some concerns | D1: Limited information regarding allocation process. Baseline demographics only reported for subset of randomised participants. D2: Analysis restricted to those receiving first dose - but judged unlikely to have substantial impact on the results |
|  |  | All | Dropout (adverse events) | Some concerns | Some concerns | High | Low | Low | High | D1: Limited information regarding allocation process. Baseline demographics only reported for subset of randomised participants. D2: Analysis restricted to those receiving first dose - but judged unlikely to have substantial impact on the results. D3: Some missing data and likely that it could depend on the true value of the outcome. |
|  |  | All | Extent of use (frequency; EoT) | Some concerns | Some concerns | High | Low | Some concerns | High | D1: Limited information regarding allocation process. Baseline demographics only reported for subset of randomised participants. D2: Analysis restricted to those receiving first dose - but judged unlikely to have substantial impact on the results. D3: Some missing data and likely that it could depend on the true value of the outcome. D5: No evidence of a pre-specified analysis plan. |
|  |  | All | Extent of use (quantity; EoT) | Some concerns | Some concerns | Some concerns | Low | Some concerns | Some concerns | D1: Limited information regarding allocation process. Baseline demographics only reported for subset of randomised participants. D2: Analysis restricted to those receiving first dose - but judged unlikely to have substantial impact on the results. D3: Some missing data but unlikely that missingness depended on the true value of outcome. D5: No evidence of a pre-specified analysis plan. |
| Pettinati 2008 | 1. Placebo 2. Naltrexone 3. Disulfiram 4. Disulfiram + Naltrexone | All | Dropout (any reason) | Some concerns | Low | Low | Low | Low | Some concerns | D1: Limited information regarding randomization and allocation process. |
|  |  | 2 vs 1;  4 vs 1;  3 vs 2;  4 vs 2 | Continuous abstinence (treatment) | Some concerns | Low | Some concerns | Low | Some concerns | Some concerns | D1: Limited information regarding randomization and allocation process. D3: Some missing data but unlikely that missingness depended on the true value of outcome. D5: No evidence of a pre-specified analysis plan. |
|  |  | 3 vs 1;  4 vs 3 | Continuous abstinence (treatment) | Some concerns | Low | High | Low | Some concerns | High | D1: Limited information regarding randomization and allocation process. D3: Some missing data and likely that it could depend on the true value of the outcome. D5: No evidence of a pre-specified analysis plan. |
| Pettinati 2014 | 1. Placebo 2. Naltrexone | All | Dropout (any reason) | Low | Low | Low | Low | Low | Low |  |
|  |  | All | Serious adverse events | Low | Low | Low | Low | Low | Low |  |
|  |  | All | Continuous abstinence (treatment) | Low | Low | Some concerns | Low | Some concerns | Some concerns | D3: Some missing data but unlikely that missingness depended on the true value of outcome. D5: No evidence of a pre-specified analysis plan. |
| Plebani 2012 | 1. Placebo 2. Varenicline | All | Point abstinence (EoT) | Some concerns | Low | Low | Low | Some concerns | Some concerns | D1: Limited information regarding allocation process. D5: No evidence of a pre-specified analysis plan. |
| Raby 2014 | 1. Placebo 2. Venlafaxine | All | Dropout (any reason) | Low | Low | Low | Low | Low | Low |  |
|  |  | All | Dropout (adverse events) | Low | Low | High | Low | Low | High | D3: Some missing data and likely that it could depend on the true value of the outcome. |
|  |  | All | Serious adverse events | Low | Low | Low | Low | Low | Low |  |
|  |  | All | Continuous abstinence (treatment) | Low | Low | High | Low | Some concerns | High | D3: Some missing data and likely that it could depend on the true value of the outcome. D5: No evidence of a pre-specified analysis plan. |
|  |  | All | Extent of use (proportion tests/days; EoT) | Low | Low | High | Low | Some concerns | High | D3: Some missing data and likely that it could depend on the true value of the outcome. D5: No evidence of a pre-specified analysis plan. |
|  |  | All | Extent of use (frequency; EoT) | Low | Low | High | Low | Some concerns | High | D3: Some missing data and likely that it could depend on the true value of the outcome. D5: No evidence of a pre-specified analysis plan. |
| Reid 2005a | 1. Placebo 2. Celecoxib | All | Dropout (any reason) | Some concerns | High | Low | Low | Low | High | D1: Limited information regarding allocation process. D2: Part of CREST protocol is modified blind, meaning participants and people responsible for intervention delivery likely unblinded. |
|  |  | All | Dropout (adverse events) | Some concerns | High | High | Some concerns | Low | High | D1: Limited information regarding allocation process. D2: Part of CREST protocol is modified blind, meaning participants and people responsible for intervention delivery likely unblinded. D3: Some missing data and likely that it could depend on the true value of the outcome. D4: The outcome was self-reported and participants may be aware of their group allocation, but unlikely to be a substantial portion of participants. |
|  |  | All | Point abstinence (EoT) | Some concerns | High | High | Low | Some concerns | High | D1: Limited information regarding allocation process. D2: Part of CREST protocol is modified blind, meaning participants and people responsible for intervention delivery likely unblinded. D3: Some missing data and likely that it could depend on the true value of the outcome. D5: No evidence of a pre-specified analysis plan. |
|  |  | All | Extent of use (BE level; EoT) | Some concerns | High | High | Low | Some concerns | High | D1: Limited information regarding allocation process. D2: Part of CREST protocol is modified blind, meaning participants and people responsible for intervention delivery likely unblinded. D3: Some missing data and likely that it could depend on the true value of the outcome. D5: No evidence of a pre-specified analysis plan. |
|  |  | All | Extent of use (frequency; EoT) | Some concerns | High | High | Some concerns | Some concerns | High | D1: Limited information regarding allocation process. D2: Part of CREST protocol is modified blind, meaning participants and people responsible for intervention delivery likely unblinded. D3: Some missing data and likely that it could depend on the true value of the outcome. D4: The outcome was self-reported and participants may be aware of their group allocation, but unlikely to be a substantial portion of participants. D5: No evidence of a pre-specified analysis plan. |
| Reid 2005b | 1. Placebo 2. Olanzapine 3. Valproate 4. Coenzyme Q10 + L-carnitine | All | Dropout (any reason) | Some concerns | High | Low | Low | Low | High | D1: Limited information regarding allocation process. Baseline demographics only reported for subset of randomised participants. D2: Part of CREST protocol is modified blind, meaning participants and people responsible for intervention delivery likely unblinded. |
|  |  | All | Serious adverse events | Some concerns | High | Low | Low | Low | High | D1: Limited information regarding allocation process. Baseline demographics only reported for subset of randomised participants. D2: Part of CREST protocol is modified blind, meaning participants and people responsible for intervention delivery likely unblinded. |
|  |  | All | Mortality | Some concerns | High | Low | Low | Low | High | D1: Limited information regarding allocation process. Baseline demographics only reported for subset of randomised participants. D2: Part of CREST protocol is modified blind, meaning participants and people responsible for intervention delivery likely unblinded. |
|  |  | 2 vs 1;  3 vs 1;  4 vs 1;  4 vs 2;  3 vs 2 | Point abstinence (EoT) | Some concerns | High | Some concerns | Low | Some concerns | High | D1: Limited information regarding allocation process. Baseline demographics only reported for subset of randomised participants. D2: Part of CREST protocol is modified blind, meaning participants and people responsible for intervention delivery likely unblinded; also analysis limited to specific subgroup. D3: Some missing data but unlikely that missingness depended on the true value of outcome. D5: No evidence of a pre-specified analysis plan. |
|  |  | 4 vs 3 | Point abstinence (EoT) | Some concerns | High | High | Low | Some concerns | High | D1: Limited information regarding allocation process. Baseline demographics only reported for subset of randomised participants. D2: Part of CREST protocol is modified blind, meaning participants and people responsible for intervention delivery likely unblinded; also analysis limited to specific subgroup. D3: Some missing data and likely that it could depend on the true value of the outcome. D5: No evidence of a pre-specified analysis plan. |
|  |  | 2 vs 1;  3 vs 1;  4 vs 1;  4 vs 2;  3 vs 2 | Extent of use (BE level; EoT) | Some concerns | High | Some concerns | Low | Some concerns | High | D1: Limited information regarding allocation process. Baseline demographics only reported for subset of randomised participants. D2: Part of CREST protocol is modified blind, meaning participants and people responsible for intervention delivery likely unblinded; also analysis limited to specific subgroup. D3: Some missing data but unlikely that missingness depended on the true value of outcome. D5: No evidence of a pre-specified analysis plan. |
|  |  | 4 vs 3 | Extent of use (BE level; EoT) | Some concerns | High | High | Low | Some concerns | High | D1: Limited information regarding allocation process. Baseline demographics only reported for subset of randomised participants. D2: Part of CREST protocol is modified blind, meaning participants and people responsible for intervention delivery likely unblinded; also analysis limited to specific subgroup. D3: Some missing data and likely that it could depend on the true value of the outcome. D5: No evidence of a pre-specified analysis plan. |
|  |  | 2 vs 1;  3 vs 1;  4 vs 1;  4 vs 2;  3 vs 2 | Extent of use (frequency; EoT) | Some concerns | High | Some concerns | Some concerns | Some concerns | High | D1: Limited information regarding allocation process. Baseline demographics only reported for subset of randomised participants. D2: Part of CREST protocol is modified blind, meaning participants and people responsible for intervention delivery likely unblinded; also analysis limited to specific subgroup. D3: Some missing data but unlikely that missingness depended on the true value of outcome. D4: The outcome was self-reported and participants may be aware of their group allocation, but unlikely to be a substantial portion of participants. D5: No evidence of a pre-specified analysis plan. |
|  |  | 4 vs 3 | Extent of use (frequency; EoT) | Some concerns | High | High | Some concerns | Some concerns | High | D1: Limited information regarding allocation process. Baseline demographics only reported for subset of randomised participants. D2: Part of CREST protocol is modified blind, meaning participants and people responsible for intervention delivery likely unblinded; also analysis limited to specific subgroup. D3: Some missing data and likely that it could depend on the true value of the outcome. D4: The outcome was self-reported and participants may be aware of their group allocation, but unlikely to be a substantial portion of participants. D5: No evidence of a pre-specified analysis plan. |
|  |  | 2 vs 1;  3 vs 1;  4 vs 1;  4 vs 2;  3 vs 2 | Craving | Some concerns | High | Some concerns | Some concerns | Some concerns | High | D1: Limited information regarding allocation process. Baseline demographics only reported for subset of randomised participants. D2: Part of CREST protocol is modified blind, meaning participants and people responsible for intervention delivery likely unblinded; also analysis limited to specific subgroup. D3: Some missing data but unlikely that missingness depended on the true value of outcome. D4: The outcome was self-reported and participants may be aware of their group allocation, but unlikely to be a substantial portion of participants. D5: No evidence of a pre-specified analysis plan. |
|  |  | 4 vs 3 | Craving | Some concerns | High | High | Some concerns | Some concerns | High | D1: Limited information regarding allocation process. Baseline demographics only reported for subset of randomised participants. D2: Part of CREST protocol is modified blind, meaning participants and people responsible for intervention delivery likely unblinded; also analysis limited to specific subgroup. D3: Some missing data and likely that it could depend on the true value of the outcome. D4: The outcome was self-reported and participants may be aware of their group allocation, but unlikely to be a substantial portion of participants. D5: No evidence of a pre-specified analysis plan. |
| Reid 2006 | 1. Placebo 2. Mecamylamine | All | Dropout (any reason) | Some concerns | Low | Low | Low | Low | Some concerns | D1: Limited information regarding allocation process. |
|  |  | All | Serious adverse events | Some concerns | Low | Low | Low | Low | Some concerns | D1: Limited information regarding allocation process. |
|  |  | All | Extent of use (frequency; EoT) | Some concerns | Low | High | Low | Some concerns | High | D1: Limited information regarding allocation process. D3: Some missing data and likely that it could depend on the true value of the outcome. D5: No evidence of a pre-specified analysis plan. |
|  |  | All | Craving | Some concerns | Low | High | Low | Some concerns | High | D1: Limited information regarding allocation process. D3: Some missing data and likely that it could depend on the true value of the outcome. D5: No evidence of a pre-specified analysis plan. |
| Santos 2021 | 1. Placebo 2. Lorcaserin | All | Dropout (any reason) | Low | Low | Low | Low | Low | Low |  |
|  |  | All | Adherence | Low | Low | High | Low | Low | High | D3: Some missing data and likely that it could depend on the true value of the outcome. |
|  |  | All | Serious adverse events | Low | Low | Low | Low | Low | Low |  |
|  |  | All | Point abstinence (EoT) | Low | Low | High | Low | Some concerns | High | D3: Some missing data and likely that it could depend on the true value of the outcome. D5: Outcome not listed in registration history prior to study completion. |
| Schmitz 2001 | 1. Placebo 2. Fluoxetine | All | Dropout (any reason) | Some concerns | Low | Low | Low | Low | Some concerns | D1: Limited information regarding allocation process. |
|  |  | All | Adherence | Some concerns | Low | High | Low | Some concerns | High | D1: Limited information regarding allocation process. D3: Some missing data and likely that it could depend on the true value of the outcome. D5: No evidence of a pre-specified analysis plan. |
| Schmitz 2008 | 1. Placebo 2. Levodopa + Carbidopa | All | Dropout (any reason) | Low | Low | Low | Low | Low | Low |  |
|  |  | All | Continuous abstinence (treatment) | Low | High | High | Low | Some concerns | High | D2: Analysis was restricted to N treated (i.e., not ITT or mITT).  D3: Some missing data and likely that it could depend on the true value of the outcome. D5: No evidence of a pre-specified analysis plan. |
| Schmitz 2009 | 1. Placebo 2. Naltrexone | All | Extent of use (proportion tests/days; EoT) | Low | Some concerns | High | Low | Some concerns | High | D2: Appears that a per protocol analysis was used - but judged unlikely to have substantial impact on the results. D3: Some missing data. Amount and difference between groups unclear. D5: No evidence of a pre-specified analysis plan. |
| Schmitz 2012 | 1. Placebo 2. Dexamfetamine + Modafinil 3. Modafinil 4. Dexamfetamine | All | Dropout (any reason) | Low | Low | Low | Low | Low | Low |  |
|  |  | All | Extent of use (proportion tests/days; EoT) | Low | High | High | Low | High | High | D2: Analysis was restricted to N treated (i.e., not ITT or mITT).  D3: Some missing data and likely that it could depend on the true value of the outcome. D5: No evidence of a pre-specified analysis plan. The result taken from the registration – outcome is not mentioned in the paper. |
| Schmitz 2014 | 1. Placebo 2. Modafinil 3. Carbidopa + levodopa 4. Naltrexone | All | Dropout (any reason) | Some concerns | Low | Low | Low | Low | Some concerns | D1: Limited information regarding allocation process. |
|  |  | 2 vs 1;  4 vs 1;  3 vs 2;  4 vs 2;  4 vs 3 | Adherence | Some concerns | High | High | Low | Some concerns | High | D1: Limited information regarding allocation process. D2: Analysis was restricted to N treated (i.e., not ITT or mITT).  D3: Some missing data and likely that it could depend on the true value of the outcome. D5: No evidence of a pre-specified analysis plan. |
|  |  | 3 vs 1 | Adherence | Some concerns | Some concerns | High | Low | Some concerns | High | D1: Limited information regarding allocation process. D2: Analysis restricted to those receiving first dose - but judged unlikely to have substantial impact on the results. D3: Some missing data and likely that it could depend on the true value of the outcome. D5: No evidence of a pre-specified analysis plan. |
|  |  | 2 vs 1;  4 vs 1;  3 vs 2;  4 vs 2;  4 vs 3 | Serious adverse events | Some concerns | High | Low | Low | Low | High | D1: Limited information regarding allocation process. D2: Analysis was restricted to N treated (i.e., not ITT or mITT). |
|  |  | 3 vs 1 | Serious adverse events | Some concerns | Some concerns | Low | Low | Low | Some concerns | D1: Limited information regarding allocation process. D2: Analysis restricted to those receiving first dose - but judged unlikely to have substantial impact on the results. |
|  |  | 2 vs 1;  4 vs 1;  3 vs 2;  4 vs 2;  4 vs 3 | Extent of use (proportion tests/days; EoT) | Some concerns | High | High | Low | Some concerns | High | D1: Limited information regarding allocation process. D2: Analysis was restricted to N treated (i.e., not ITT or mITT).  D3: Some missing data and likely that it could depend on the true value of the outcome. D5: No evidence of a pre-specified analysis plan. |
|  |  | 3 vs 1 | Extent of use (proportion tests/days; EoT) | Some concerns | Some concerns | High | Low | Some concerns | High | D1: Limited information regarding allocation process. D2: Analysis restricted to those receiving first dose - but judged unlikely to have substantial impact on the results. D3: Some missing data and likely that it could depend on the true value of the outcome. D5: No evidence of a pre-specified analysis plan. |
| Schmitz 2017 | 1. Placebo 2. Pioglitazone | All | Dropout (any reason) | Low | Low | Low | Low | Low | Low |  |
|  |  | All | Adherence | Low | High | Some concerns | Low | Some concerns | High | D2: Analysis was restricted to N treated (i.e., not ITT or mITT).  D3: Some missing data but unlikely that missingness depended on the true value of outcome. D5: No evidence of a pre-specified analysis plan. |
|  |  | All | Extent of use (proportion tests/days; EoT) | Low | High | Some concerns | Low | Some concerns | High | D2: Analysis was restricted to N treated (i.e., not ITT or mITT).  D3: Some missing data but unlikely that missingness depended on the true value of outcome. D5: No evidence of a pre-specified analysis plan. |
|  |  | All | Craving | Low | High | Some concerns | Low | Some concerns | High | D2: Analysis was restricted to N treated (i.e., not ITT or mITT).  D3: Some missing data but unlikely that missingness depended on the true value of outcome. D5: No evidence of a pre-specified analysis plan. |
| Schmitz 2021 | 1. Placebo 2. Levodopa + carbidopa 3. Levodopa + carbidopa + ropinirole (2mg) 4. Levodopa + carbidopa + ropinirole (4mg) | All | Dropout (any reason) | Low | Low | Low | Low | Low | Low |  |
|  |  | All | Adherence | Low | Some concerns | Some concerns | Low | Some concerns | Some concerns | D2: Analysis restricted to those receiving first dose - but judged unlikely to have substantial impact on the results. D3: Some missing data but unlikely that missingness depended on the true value of outcome. D5: No evidence of a pre-specified analysis plan. |
|  |  | All | Serious adverse events | Low | Low | Low | Low | Low | Low |  |
|  |  | 2 vs 1;  3 vs 2;  4 vs 2 | Longest duration of continuous abstinence | Low | Some concerns | Some concerns | Low | Some concerns | Some concerns | D2: Analysis restricted to those receiving first dose - but judged unlikely to have substantial impact on the results. D3: Some missing data but unlikely that missingness depended on the true value of outcome. D5: No evidence of a pre-specified analysis plan. |
|  |  | 3 vs 1;  4 vs 1;  4 vs 2 | Longest duration of continuous abstinence | Low | Low | Some concerns | Low | Some concerns | Some concerns | D3: Some missing data but unlikely that missingness depended on the true value of outcome. D5: No evidence of a pre-specified analysis plan. |
|  |  | 2 vs 1;  3 vs 2;  4 vs 2 | Extent of use (proportion tests/days; EoT) | Low | Some concerns | Some concerns | Low | Some concerns | Some concerns | D2: Analysis restricted to those receiving first dose - but judged unlikely to have substantial impact on the results. D3: Some missing data but unlikely that missingness depended on the true value of outcome. D5: No evidence of a pre-specified analysis plan. |
|  |  | 3 vs 1;  4 vs 1;  4 vs 2 | Extent of use (proportion tests/days; EoT) | Low | Low | Some concerns | Low | Some concerns | Some concerns | D3: Some missing data but unlikely that missingness depended on the true value of outcome. D5: No evidence of a pre-specified analysis plan. |
| Schmitz 2024 | 1. Placebo 2. Modafinil | All | Adherence | Low | High | High | Low | Some concerns | High | D2: Analysis was restricted to N treated (i.e., not ITT or mITT).  D3: Some missing data. Amount and difference between groups unclear. D5: No evidence of a pre-specified analysis plan. |
|  |  | All | Serious adverse events | Low | Low | Low | Low | Low | Low |  |
|  |  | All | Extent of use (proportion tests/days; EoT) | Low | High | High | Low | Some concerns | High | D2: Analysis was restricted to N treated (i.e., not ITT or mITT).  D3: Some missing data. Amount and difference between groups unclear. D5: No evidence of a pre-specified analysis plan. |
|  |  | All | Extent of use (frequency; EoT) | Low | High | High | Low | Some concerns | High | D2: Analysis was restricted to N treated (i.e., not ITT or mITT).  D3: Some missing data. Amount and difference between groups unclear. D5: No evidence of a pre-specified analysis plan. |
| Schubiner 2002 | 1. Placebo 2. Methylphenidate | All | Dropout (any reason) | Low | Low | Low | Low | Low | Low |  |
|  |  | All | Dropout (adverse events) | Low | Low | High | Low | Low | High | D3: Some missing data and likely that it could depend on the true value of the outcome. |
|  |  | All | Longest duration of continuous abstinence | Low | Low | High | Low | Some concerns | High | D3: Some missing data and likely that it could depend on the true value of the outcome. D5: No evidence of a pre-specified analysis plan. |
|  |  | All | Extent of use (frequency; EoT) | Low | Low | High | Low | Some concerns | High | D3: Some missing data and likely that it could depend on the true value of the outcome. D5: No evidence of a pre-specified analysis plan. |
| Shearer 2003 | 1. Placebo 2. Dexamphetamine | All | Dropout (any reason) | Low | Low | Low | Low | Low | Low |  |
|  |  | All | Craving | Low | High | High | Low | Some concerns | High | D2: Analysis was restricted to N treated (i.e., not ITT or mITT).  D3: Some missing data. Amount and difference between groups unclear. D5: No evidence of a pre-specified analysis plan. |
| Shoptaw 2002 | 1. Placebo 2. Amantadine | All | Dropout (any reason) | Some concerns | Low | Low | Low | Low | Some concerns | D1: Limited information regarding allocation process. |
|  |  | All | Serious adverse events | Some concerns | Some concerns | Low | Low | Low | Some concerns | D1: Limited information regarding allocation process. D2: Analysis restricted to those receiving first dose - but judged unlikely to have substantial impact on the results. |
|  |  | All | Continuous abstinence (treatment) | Some concerns | Some concerns | High | Low | Some concerns | High | D1: Limited information regarding allocation process. D2: Analysis restricted to those receiving first dose - but judged unlikely to have substantial impact on the results. D3: Some missing data and likely that it could depend on the true value of the outcome. D5: No evidence of a pre-specified analysis plan. |
|  |  |  | Point abstinence (EoT) | Some concerns | Some concerns | High | Low | Some concerns | High | D1: Limited information regarding allocation process. D2: Analysis restricted to those receiving first dose - but judged unlikely to have substantial impact on the results. D3: Some missing data and likely that it could depend on the true value of the outcome. D5: No evidence of a pre-specified analysis plan. |
|  |  | All | Point abstinence (follow-up) | Some concerns | Some concerns | High | Low | Some concerns | High | D1: Limited information regarding allocation process. D2: Analysis restricted to those receiving first dose - but judged unlikely to have substantial impact on the results. D3: Some missing data and likely that it could depend on the true value of the outcome. D5: No evidence of a pre-specified analysis plan. |
|  |  | All | Longest duration of continuous abstinence | Some concerns | Some concerns | High | Low | Some concerns | High | D1: Limited information regarding allocation process. D2: Analysis restricted to those receiving first dose - but judged unlikely to have substantial impact on the results. D3: Some missing data and likely that it could depend on the true value of the outcome. D5: No evidence of a pre-specified analysis plan. |
|  |  | All | Extent of use (proportion tests/days; EoT) | Some concerns | Some concerns | High | Low | Some concerns | High | D1: Limited information regarding allocation process. D2: Analysis restricted to those receiving first dose - but judged unlikely to have substantial impact on the results. D3: Some missing data and likely that it could depend on the true value of the outcome. D5: No evidence of a pre-specified analysis plan. |
|  |  | All | Craving | Some concerns | Some concerns | High | Low | Some concerns | High | D1: Limited information regarding allocation process. D2: Analysis restricted to those receiving first dose - but judged unlikely to have substantial impact on the results. D3: Some missing data and likely that it could depend on the true value of the outcome. D5: No evidence of a pre-specified analysis plan. |
| Shoptaw 2003 | 1. Placebo 2. Baclofen | All | Dropout (any reason) | Some concerns | Low | Low | Low | Low | Some concerns | D1: Limited information regarding allocation process. |
|  |  | All | Dropout (adverse events) | Some concerns | Low | High | Low | Low | High | D1: Limited information regarding allocation process. D3: Some missing data and likely that it could depend on the true value of the outcome. |
|  |  | All | Adherence | Some concerns | Low | High | Low | Some concerns | High | D1: Limited information regarding allocation process. D3: Some missing data and likely that it could depend on the true value of the outcome. D5: No evidence of a pre-specified analysis plan. |
|  |  | All | Continuous abstinence (treatment) | Some concerns | Low | High | Low | Some concerns | High | D1: Limited information regarding allocation process. D3: Some missing data and likely that it could depend on the true value of the outcome. D5: No evidence of a pre-specified analysis plan. |
|  |  | All | Point abstinence (EoT) | Some concerns | Low | High | Low | Some concerns | High | D1: Limited information regarding allocation process. D3: Some missing data and likely that it could depend on the true value of the outcome. D5: No evidence of a pre-specified analysis plan. |
|  |  | All | Longest duration of continuous abstinence | Some concerns | Low | High | Low | Some concerns | High | D1: Limited information regarding allocation process. D3: Some missing data and likely that it could depend on the true value of the outcome. D5: No evidence of a pre-specified analysis plan. |
| Shoptaw 2004 | 1. Placebo 2. Dehydroepiandrosterone | All | Dropout (any reason) | Some concerns | Low | Low | Low | Low | Low | D1: Limited information regarding allocation process. |
|  |  | All | Continuous abstinence (treatment) | Some concerns | Low | High | Low | Some concerns | High | D1: Limited information regarding allocation process. D3: Some missing data and likely that it could depend on the true value of the outcome. D5: No evidence of a pre-specified analysis plan. |
|  |  | All | Extent of use (proportion tests/days; EoT) | Some concerns | Low | High | Low | Some concerns | High | D1: Limited information regarding allocation process. D3: Some missing data and likely that it could depend on the true value of the outcome. D5: No evidence of a pre-specified analysis plan. |
|  |  | All | Craving | Some concerns | Low | High | Low | Some concerns | High | D1: Limited information regarding allocation process. D3: Some missing data and likely that it could depend on the true value of the outcome. D5: No evidence of a pre-specified analysis plan. |
| Shoptaw 2005 | 1. Placebo 2. Cabergoline 3. Carbidopa + levodopa 4. Ergoloid mesylates | All | Dropout (any reason) | Some concerns | High | Low | Low | Low | High | D1: Limited information regarding allocation process. D2: Part of CREST protocol is modified blind, meaning participants and people responsible for intervention delivery likely unblinded. |
|  |  | All | Point abstinence (EoT) | Some concerns | High | High | Low | Some concerns | High | D1: Limited information regarding allocation process. D2: Part of CREST protocol is modified blind, meaning participants and people responsible for intervention delivery likely unblinded. D3: Some missing data and likely that it could depend on the true value of the outcome. D5: No evidence of a pre-specified analysis plan. |
|  |  | All | Extent of use (proportion tests/days; EoT) | Some concerns | High | High | Low | Some concerns | High | D1: Limited information regarding allocation process. D2: Part of CREST protocol is modified blind, meaning participants and people responsible for intervention delivery likely unblinded. D3: Some missing data and likely that it could depend on the true value of the outcome. D5: No evidence of a pre-specified analysis plan. |
|  |  | All | Craving | Some concerns | High | High | Some concerns | Some concerns | High | D1: Limited information regarding allocation process. D2: Part of CREST protocol is modified blind, meaning participants and people responsible for intervention delivery likely unblinded. D3: Some missing data and likely that it could depend on the true value of the outcome. D4: The outcome was self-reported and participants may be aware of their group allocation, but unlikely to be a substantial portion of participants. D5: No evidence of a pre-specified analysis plan. |
| Shorter 2013 | 1. Placebo 2. Doxazosin (fast) 3. Doxazosin (slow) | All | Dropout (any reason) | Some concerns | High | Low | Low | Low | High | D1: Limited information regarding allocation process. D2: Analysis was restricted to N treated (i.e., not ITT or mITT). |
|  |  | 2 vs 1;  3 vs 1 | Continuous abstinence (treatment) | Some concerns | High | High | Low | Some concerns | High | D1: Limited information regarding allocation process. D2: Analysis was restricted to N treated (i.e., not ITT or mITT).  D3: Some missing data and likely that it could depend on the true value of the outcome. D5: No evidence of a pre-specified analysis plan. |
|  |  | 3 vs 2 | Continuous abstinence (treatment) | Some concerns | High | Some concerns | Low | Some concerns | High | D1: Limited information regarding allocation process. D2: Analysis was restricted to N treated (i.e., not ITT or mITT).  D3: Some missing data but unlikely that missingness depended on the true value of outcome. D5: No evidence of a pre-specified analysis plan. |
| Sofuoglu 2007 | 1. Placebo 2. Progesterone | All | Dropout (any reason) | Some concerns | Low | Low | Low | Low | Some concerns | D1: Limited information regarding allocation process. |
| Sofuoglu 2011 | 1. Placebo 2. Galantamine | All | Dropout (any reason) | Some concerns | Low | Low | Low | Low | Some concerns | D1: Limited information regarding allocation process. |
|  |  | All | Dropout (adverse events) | Some concerns | Low | High | Low | Low | High | D1: Limited information regarding allocation process. D3: Some missing data and likely that it could depend on the true value of the outcome. |
|  |  | All | Extent of use (frequency; EoT) | Some concerns | Low | High | Low | Some concerns | High | D1: Limited information regarding allocation process. D3: Some missing data and likely that it could depend on the true value of the outcome. D5: No evidence of a pre-specified analysis plan. |
| Sofuoglu 2017 | 1. Placebo 2. Carvedilol (25mg) 3. Carvedilol (50mg) | All | Dropout (any reason) | Low | Low | Low | Low | Low | Low |  |
|  |  | All | Adherence | Low | Low | Low | Low | Some concerns | Some concerns | D5: No evidence of a pre-specified analysis plan. |
|  |  | 2 vs 1 | Extent of use (proportion tests/days; EoT) | Low | Low | High | Low | Some concerns | High | D3: Some missing data and likely that it could depend on the true value of the outcome. D5: No evidence of a pre-specified analysis plan. |
|  |  | 3 vs 1;  3 vs 2 | Extent of use (proportion tests/days; EoT) | Low | Low | Some concerns | Low | Some concerns | Some concerns | D3: Some missing data but unlikely that missingness depended on the true value of outcome. D5: No evidence of a pre-specified analysis plan. |
| Somoza 2013 | 1. Placebo 2. Vigabatrin | All | Dropout (any reason) | Low | Low | Low | Low | Low | Low |  |
|  |  | All | Dropout (adverse events) | Low | Low | Some concerns | Low | Low | Some concerns | D3: Some missing data but unlikely that missingness depended on the true value of outcome. |
|  |  | All | Serious adverse events | Low | Low | Low | Low | Low | Low |  |
|  |  | All | Mortality | Low | Low | Low | Low | Low | Low |  |
|  |  | All | Continuous abstinence (treatment) | Low | Low | Some concerns | Low | Low | Some concerns | D3: Some missing data but unlikely that missingness depended on the true value of outcome. |
|  |  | All | Extent of use (proportion tests/days; EoT) | Low | Low | Some concerns | Low | Low | Some concerns | D3: Some missing data but unlikely that missingness depended on the true value of outcome. |
|  |  | All | Craving | Low | Low | Some concerns | Low | Some concerns | Some concerns | D3: Some missing data but unlikely that missingness depended on the true value of outcome. D5: No evidence of a pre-specified analysis plan. |
| Stine 1995 | 1. Placebo 2. Mazindol | All | Dropout (any reason) | Some concerns | Low | Low | Low | Low | Some concerns | D1: Limited information regarding randomization and allocation process. |
|  |  | All | Continuous abstinence (treatment) | Some concerns | Low | High | Low | Some concerns | High | D1: Limited information regarding randomization and allocation process. D3: Some missing data and likely that it could depend on the true value of the outcome. D5: No evidence of a pre-specified analysis plan. |
|  |  | All | Extent of use (frequency; EoT) | Some concerns | Low | High | Low | Some concerns | High | D1: Limited information regarding randomization and allocation process. D3: Some missing data and likely that it could depend on the true value of the outcome. D5: No evidence of a pre-specified analysis plan. |
|  |  | All | Extent of use (quantity; EoT) | Some concerns | Low | High | Low | Some concerns | High | D1: Limited information regarding randomization and allocation process. D3: Some missing data and likely that it could depend on the true value of the outcome. D5: No evidence of a pre-specified analysis plan. |
| Suchting 2021 | 1. Placebo 2. Citalopram (40mg) 3. Citalopram (20 mg) | All | Dropout (any reason) | Low | Low | Low | Low | Low | Low |  |
|  |  | 2 vs 1;  3 vs 2 | Adherence | Low | Some concerns | Some concerns | Low | Some concerns | Some concerns | D2: Analysis restricted to those receiving first dose - but judged unlikely to have substantial impact on the results. D3: Some missing data but unlikely that missingness depended on the true value of outcome. D5: Outcome not listed in registration history prior to study completion. |
|  |  | 3 vs 1 | Adherence | Low | Low | Some concerns | Low | Some concerns | Some concerns | D3: Some missing data but unlikely that missingness depended on the true value of outcome. D5: Outcome not listed in registration history prior to study completion. |
|  |  | 2 vs 1;  3 vs 2 | Serious adverse events | Low | Some concerns | Low | Low | Low | Some concerns | D2: Analysis restricted to those receiving first dose - but judged unlikely to have substantial impact on the results. |
|  |  | 3 vs 1 | Serious adverse events | Low | Low | Low | Low | Low | Low |  |
|  |  | 2 vs 1;  3 vs 2 | Continuous abstinence (treatment) | Low | Some concerns | Some concerns | Low | Low | Some concerns | D2: Analysis restricted to those receiving first dose - but judged unlikely to have substantial impact on the results. D3: Some missing data but unlikely that missingness depended on the true value of outcome. |
|  |  | 3 vs 1 | Continuous abstinence (treatment) | Low | Low | Some concerns | Low | Low | Some concerns | D3: Some missing data but unlikely that missingness depended on the true value of outcome. |
|  |  | 2 vs 1;  3 vs 2 | Longest duration of continuous abstinence | Low | Some concerns | Some concerns | Low | Low | Some concerns | D2: Analysis restricted to those receiving first dose - but judged unlikely to have substantial impact on the results. D3: Some missing data but unlikely that missingness depended on the true value of outcome. |
|  |  | 3 vs 1 | Longest duration of continuous abstinence | Low | Low | Some concerns | Low | Low | Some concerns | D3: Some missing data but unlikely that missingness depended on the true value of outcome. |
|  |  | 2 vs 1;  3 vs 2 | Extent of use (proportion tests/days; EoT) | Low | Some concerns | Low | Low | Low | Some concerns | D2: Analysis restricted to those receiving first dose - but judged unlikely to have substantial impact on the results. |
|  |  | 3 vs 1 | Extent of use (proportion tests/days; EoT) | Low | Low | Low | Low | Low | Low |  |
| Tapp 2015 | 1. Placebo 2. Quetiapine | All | Dropout (any reason) | Low | Low | Low | Low | Low | Low |  |
|  |  | All | Dropout (adverse events) | Low | High | High | Low | Low | High | D2: Appears that a per protocol analysis was used D3: Some missing data. Amount and difference between groups unclear. |
|  |  | All | Adherence | Low | Low | High | Low | Some concerns | High | D3: Some missing data and likely that it could depend on the true value of the outcome. D5: No evidence of a pre-specified analysis plan. |
|  |  | All | Continuous abstinence (treatment) | Low | Low | High | Low | Some concerns | High | D3: Some missing data and likely that it could depend on the true value of the outcome. D5: No evidence of a pre-specified analysis plan. |
|  |  | All | Extent of use (proportion tests/days; EoT) | Low | Low | High | Low | Some concerns | High | D3: Some missing data and likely that it could depend on the true value of the outcome. D5: No evidence of a pre-specified analysis plan. |
|  |  | All | Extent of use (frequency; EoT) | Low | Low | High | Low | Some concerns | High | D3: Some missing data and likely that it could depend on the true value of the outcome. D5: No evidence of a pre-specified analysis plan. |
|  |  | All | Extent of use (quantity; EoT) | Low | Low | High | Low | Some concerns | High | D3: Some missing data and likely that it could depend on the true value of the outcome. D5: No evidence of a pre-specified analysis plan. |
|  |  | All | Craving | Low | Low | High | Low | Some concerns | High | D3: Some missing data and likely that it could depend on the true value of the outcome. D5: No evidence of a pre-specified analysis plan. |
| Umbricht 2014 | 1. Placebo 2. Topiramate | All | Dropout (any reason) | Low | Low | Low | Low | Low | Low |  |
|  |  | All | Dropout (adverse events) | Low | Low | Some concerns | Low | Low | Some concerns | D3: Some missing data but unlikely that missingness depended on the true value of outcome. |
|  |  | All | Longest duration of continuous abstinence | Low | Low | Some concerns | Low | Some concerns | Some concerns | D3: Some missing data but unlikely that missingness depended on the true value of outcome. D5: No evidence of a pre-specified analysis plan. |
|  |  | All | Extent of use (proportion tests/days; EoT) | Low | Low | Low | Low | Some concerns | Some concerns | D5: No evidence of a pre-specified analysis plan. |
| Walsh 2013 | 1. Placebo 2. Atomoxetine | All | Dropout (any reason) | Low | High | Low | Low | Low | High | D2: Analysis was restricted to N treated (i.e., not ITT or mITT). |
|  |  | All | Extent of use (proportion tests/days; EoT) | Low | High | High | Low | Low | High | D2: Analysis was restricted to N treated (i.e., not ITT or mITT).  D3: Some missing data and likely that it could depend on the true value of the outcome. |
| Wardle 2017 | 1. Placebo 2. Carbidopa + levodopa | All | Dropout (any reason) | Low | Low | Low | Low | Low | Low |  |
|  |  | All | Dropout (adverse events) | Low | Low | High | Low | Low | High | D3: Some missing data and likely that it could depend on the true value of the outcome. |
|  |  | All | Adherence | Low | Low | High | Low | Some concerns | High | D3: Some missing data and likely that it could depend on the true value of the outcome. D5: No evidence of a pre-specified analysis plan. |
|  |  | All | Serious adverse events | Low | Low | Low | Low | Low | Low |  |
|  |  | All | Extent of use (proportion tests/days; EoT) | Low | Low | High | Low | Some concerns | High | D3: Some missing data and likely that it could depend on the true value of the outcome. D5: No evidence of a pre-specified analysis plan. |
| Ware 2023 | 1. Placebo 2. Bupropion | All | Dropout (any reason) | Low | Low | Low | Low | Low | Low |  |
|  |  | All | Point abstinence (follow-up) | Low | Some concerns | High | Low | Some concerns | High | D2: Analysis restricted to those receiving first dose - but judged unlikely to have substantial impact on the results. D3: Some missing data and likely that it could depend on the true value of the outcome. D5: Outcome not listed in protocol. |
|  |  | All | Longest duration of continuous abstinence | Low | Some concerns | High | Low | Low | High | D2: Analysis restricted to those receiving first dose - but judged unlikely to have substantial impact on the results. D3: Some missing data and likely that it could depend on the true value of the outcome. |
| Winhusen 2005 | 1. Placebo 2. Tiagabine 3. Sertraline 4. Donepezil | All | Dropout (any reason) | Some concerns | High | Low | Low | Low | High | D1: Limited information regarding allocation process. D2: Part of CREST protocol is modified blind, meaning participants and people responsible for intervention delivery likely unblinded. |
|  |  | 2 vs 1;  3 vs 1;  4 vs 2;  4 vs 3 | Dropout (adverse events) | Some concerns | High | High | Some concerns | Low | High | D1: Limited information regarding allocation process. D2: Part of CREST protocol is modified blind, meaning participants and people responsible for intervention delivery likely unblinded. D3: Some missing data and likely that it could depend on the true value of the outcome. D4: The outcome was self-reported and participants may be aware of their group allocation, but unlikely to be a substantial portion of participants. |
|  |  | 4 vs 1;  3 vs 2 | Dropout (adverse events) | Some concerns | High | Some concerns | Some concerns | Low | High | D1: Limited information regarding allocation process. D2: Part of CREST protocol is modified blind, meaning participants and people responsible for intervention delivery likely unblinded. D3: Some missing data but unlikely that missingness depended on the true value of outcome. D4: The outcome was self-reported and participants may be aware of their group allocation, but unlikely to be a substantial portion of participants. |
|  |  | 2 vs 1;  3 vs 1;  4 vs 2;  4 vs 3 | Adherence | Some concerns | High | High | Low | Low | High | D1: Limited information regarding allocation process. D2: Part of CREST protocol is modified blind, meaning participants and people responsible for intervention delivery likely unblinded. D3: Some missing data and likely that it could depend on the true value of the outcome. |
|  |  | 4 vs 1;  3 vs 2 | Adherence | Some concerns | High | Some concerns | Low | Low | High | D1: Limited information regarding allocation process. D2: Part of CREST protocol is modified blind, meaning participants and people responsible for intervention delivery likely unblinded. D3: Some missing data but unlikely that missingness depended on the true value of outcome. |
|  |  | All | Serious adverse events | Some concerns | High | Low | Low | Low | High | D1: Limited information regarding allocation process. D2: Part of CREST protocol is modified blind, meaning participants and people responsible for intervention delivery likely unblinded. |
|  |  | 2 vs 1;  3 vs 1;  4 vs 2;  4 vs 3 | Extent of use (BE level; EoT) | Some concerns | High | High | Low | Low | High | D1: Limited information regarding allocation process. D2: Part of CREST protocol is modified blind, meaning participants and people responsible for intervention delivery likely unblinded. D3: Some missing data and likely that it could depend on the true value of the outcome. |
|  |  | 4 vs 1;  3 vs 2 | Extent of use (BE level; EoT) | Some concerns | High | Some concerns | Low | Low | High | D1: Limited information regarding allocation process. D2: Part of CREST protocol is modified blind, meaning participants and people responsible for intervention delivery likely unblinded. D3: Some missing data but unlikely that missingness depended on the true value of outcome. |
|  |  | 2 vs 1;  3 vs 1;  4 vs 2;  4 vs 3 | Extent of use (frequency; EoT) | Some concerns | High | High | Some concerns | Low | High | D1: Limited information regarding allocation process. D2: Part of CREST protocol is modified blind, meaning participants and people responsible for intervention delivery likely unblinded. D3: Some missing data and likely that it could depend on the true value of the outcome. D4: The outcome was self-reported and participants may be aware of their group allocation, but unlikely to be a substantial portion of participants. |
|  |  | 4 vs 1;  3 vs 2 | Extent of use (frequency; EoT) | Some concerns | High | Some concerns | Some concerns | Low | High | D1: Limited information regarding allocation process. D2: Part of CREST protocol is modified blind, meaning participants and people responsible for intervention delivery likely unblinded. D3: Some missing data but unlikely that missingness depended on the true value of outcome. D4: The outcome was self-reported and participants may be aware of their group allocation, but unlikely to be a substantial portion of participants. |
|  |  | 2 vs 1;  3 vs 1;  4 vs 2;  4 vs 3 | Craving | Some concerns | High | High | Some concerns | Low | High | D1: Limited information regarding allocation process. D2: Part of CREST protocol is modified blind, meaning participants and people responsible for intervention delivery likely unblinded. D3: Some missing data and likely that it could depend on the true value of the outcome. D4: The outcome was self-reported and participants may be aware of their group allocation, but unlikely to be a substantial portion of participants. |
|  |  | 4 vs 1;  3 vs 2 | Craving | Some concerns | High | Some concerns | Some concerns | Low | High | D1: Limited information regarding allocation process. D2: Part of CREST protocol is modified blind, meaning participants and people responsible for intervention delivery likely unblinded. D3: Some missing data but unlikely that missingness depended on the true value of outcome. D4: The outcome was self-reported and participants may be aware of their group allocation, but unlikely to be a substantial portion of participants. |
| Winhusen 2007a | 1. Placebo 2. Reserpine | All | Dropout (any reason) | Low | Low | Low | Low | Low | Low |  |
|  |  | All | Dropout (adverse events) | Low | Low | Some concerns | Low | Low | Some concerns | D3: Some missing data but unlikely that missingness depended on the true value of outcome. |
|  |  | All | Adherence | Low | Low | Some concerns | Low | Some concerns | Some concerns | D3: Some missing data but unlikely that missingness depended on the true value of outcome. D5: No evidence of a pre-specified analysis plan. |
|  |  | All | Extent of use (BE level; EoT) | Low | Low | Some concerns | Low | Some concerns | Some concerns | D3: Some missing data but unlikely that missingness depended on the true value of outcome. D5: No evidence of a pre-specified analysis plan. |
|  |  | All | Extent of use (proportion tests/days; EoT) | Low | Low | Some concerns | Low | Some concerns | Some concerns | D3: Some missing data but unlikely that missingness depended on the true value of outcome. D5: No evidence of a pre-specified analysis plan. |
|  |  | All | Craving | Low | Low | Some concerns | Low | Some concerns | Some concerns | D3: Some missing data but unlikely that missingness depended on the true value of outcome. D5: No evidence of a pre-specified analysis plan. |
| Winhusen 2007b | 1. Placebo 2. Tiagabine | All | Dropout (any reason) | Low | Low | Low | Low | Low | Low |  |
|  |  | All | Dropout (adverse events) | Low | Low | Some concerns | Low | Low | Some concerns | D3: Some missing data but unlikely that missingness depended on the true value of outcome. |
|  |  | All | Adherence | Low | Low | Some concerns | Low | Some concerns | Some concerns | D3: Some missing data but unlikely that missingness depended on the true value of outcome. D5: No evidence of a pre-specified analysis plan. |
|  |  | All | Serious adverse events | Low | Low | Low | Low | Low | Low |  |
|  |  | All | Extent of use (proportion tests/days; EoT) | Low | Low | Some concerns | Low | Some concerns | Some concerns | D3: Some missing data but unlikely that missingness depended on the true value of outcome. D5: No evidence of a pre-specified analysis plan. |
|  |  | All | Craving | Low | Low | High | Low | Some concerns | High | D3: Some missing data. Amount and difference between groups unclear. D5: No evidence of a pre-specified analysis plan. |
| Winhusen 2014 | 1. Placebo 2. Buspirone | All | Dropout (any reason) | Low | Low | Low | Low | Low | Low |  |
|  |  | All | Adherence | Low | Low | Some concerns | Low | Some concerns | Some concerns | D3: Some missing data but unlikely that missingness depended on the true value of outcome. D5: No evidence of a pre-specified analysis plan. |
|  |  | All | Serious adverse events | Low | Low | Low | Low | Low | Low |  |
|  |  | All | Longest duration of continuous abstinence | Low | Low | Some concerns | Low | Low | Some concerns | D3: Some missing data but unlikely that missingness depended on the true value of outcome. |
|  |  | All | Extent of use (proportion tests/days; EoT) | Low | Low | Some concerns | Low | Low | Some concerns | D3: Some missing data but unlikely that missingness depended on the true value of outcome. |
| Winstanley 2011 | 1. Placebo 2. Fluoxetine | All | Dropout (any reason) | Low | Low | Low | Low | Low | Low |  |
|  |  | All | Dropout (adverse events) | Low | Low | High | Low | Low | High | D3: Some missing data and likely that it could depend on the true value of the outcome. |
| Zhang 2019 | 1. Placebo 2. Doxazosin | All | Dropout (adverse events)* | Some concerns | High | High | Low | Low | High | D1: Limited information regarding allocation process. Baseline demographics only reported for subset of randomised participants. D2: Analysis was restricted to N treated (i.e., not ITT or mITT).  D3: Some missing data. Amount and difference between groups unclear. |
|  |  | All | Serious adverse events* | Some concerns | High | High | Low | Low | High | D1: Limited information regarding allocation process. Baseline demographics only reported for subset of randomised participants. D2: Analysis was restricted to N treated (i.e., not ITT or mITT).  D3: Some missing data. Amount and difference between groups unclear. |
| Note. *result reported in secondary study. BE = benzoylecgonine; EoT = end of treatment | | | | | | | | | | |
